# Effects of a standardised *Amaranthus tricolor*–*Spinacia oleracea* extract on exercise performance, muscle strength, endurance and recovery in physically active adults: a randomised, double-blind, placebo-controlled trial

**DOI:** 10.64898/2026.09.30.26363751

**Authors:** Siddharth J. Modi, Surajkumar Patil, Dhananjay Zutshi, Vaishnavi Patil, Gayatri Ganu, Anand Bhaskar, Gunjan Gujrati, Prem Muthuraj, Anirudh Mehta, Ashit Vora, Dilip Mehta, Anselm Desouza, Juhi Srivastava

## Abstract

Dietary supplements are increasingly being adopted by physically active individuals to support exercise performance and recovery. This randomised, double-blind, placebo-controlled trial evaluated the ergogenic potential of SpinoleRG™, an HPLC-PDA-standardised combination of *Amaranthus tricolor* L. and *Spinacia oleracea* L. in healthy, recreationally active adults over 12 weeks. Participants received a total of 1,600 mg of the extract or a placebo daily before exercise. Primary endpoints included cardiopulmonary exercise testing, muscle strength, and endurance. Secondary endpoints included treadmill-based time to exhaustion (TTE), body composition, repetitions to failure, biochemical markers, delayed onset muscle soreness (DOMS), and vitals. Routine investigations and safety monitoring were also conducted. Four participants per group were withdrawn due to non-compliance and 64 participants completed the study. Compared to the placebo, the test group showed a significant increase in the ventilatory threshold, but not in VO₂ max or maximal ventilation. They also showed significantly improved one-repetition maximum strength, fatigue indices, TTE, repetitions to failure, plasma nitrite, blood lactate, creatine kinase, lactate dehydrogenase, and DOMS outcomes. Vital signs or routine parameters were not impacted meaningfully. Three minor adverse events across both groups were observed, considered unrelated to the interventions and fully resolved. Overall, the findings support the tolerability and ergogenic potential of the extract.

**Trial registration:** Clinical Trials Registry – India (CTRI) on 18 November 2025 (<u>CTRI/2025/11/097623</u>).

## 1 Introduction

Interest in evidence-based nutritional strategies to support athletic performance, recovery, fatigue management and metabolic health has increased. Multi-ingredient dietary supplements and functional foods have an advantage, in that they simultaneously target distinct biological pathways for multifaceted beneficial effects.

SpinoleRG™ is a combination extract aimed at physically active individuals, made from standardised extracts of commonly consumed leafy vegetables - *Amaranthus tricolor* (red amaranth) and *Spinacia oleracea* (green spinach) of the family Amaranthaceae. *A. tricolor* and *S. oleracea* are rich sources of diverse phytochemicals, dietary nitrate, phytoecdysteroids, nutrient-rich essential amino acids, minerals, vitamins, essential fatty acids, flavonoids and pigments [1,2]. *A. tricolor* is characterized by dietary nitrate, a precursor to nitric oxide (NO) [3–5]. NO modulates diverse cellular processes, including vasodilation, musculoskeletal efficiency and cardiovascular homeostasis [3]. *A. tricolor* has therefore shown therapeutic potential in enhancing athletic performance. *S. oleracea*, which is rich in iron, calcium, folic acid and phytoecdysteroids, notably 3′,4′,5-trihydroxy-3-methoxy-6,7-methylenedioxyflavone 4′-glucuronide (TMMG), has been clinically demonstrated to augment muscle mass and physical strength during resistance training [6,7]. It also enhances cartilage integrity while delaying extracellular matrix degradation [8].

Prior to exploring its ergogenic potential, the extract has been comprehensively characterised and its safety established with toxicological evaluations in *in vitro* and *in vivo* models. Acute toxicity studies on male and female Sprague-Dawley rats, conducted independently at a single oral dose of 2000 mg/kg body weight for 14 days did not reveal any mortality, and the extract was found to be safe and well tolerated at a single dose of 2000 mg/kg body weight. Furthermore, a single repeat oral dose was administered daily for 90 consecutive days at 0, 250, 500 and 1000 mg/kg body weight. The results indicated no toxicity; the treatment was well tolerated. The no-observed-adverse-effect-level (NOAEL) was found to be 1000 mg/kg body weight (manuscript under peer review).

The present study was a randomised, double-blind, placebo-controlled, multi-centre clinical study conducted for a period of 12 weeks on physically active healthy individuals. The trial aimed to evaluate the extract’s clinical safety, tolerability, and efficacy on cardiorespiratory health, endurance, changes in muscle strength, and recovery from post-exercise activities in the cohort. To the best of our knowledge, this study represents the first investigation into the effects of such a combination on exercise performance. This research aims to fill a gap in the existing literature by exploring the potential synergistic benefits of combining bioactives from these widespread foods in promoting overall physical fitness and well-being in physically active populations.

## 2 Methods

The study was conducted by MPREX Healthcare Pvt. Ltd. Pune, India, an external contract research organisation (CRO), on behalf of Phytoveda Pvt. Ltd, Mumbai, India.

### 2.1 Study design

The study was a randomized, double-blind, placebo-controlled, multi-centre clinical trial.

#### 2.1.1 Sample size calculation

Mean time to exhaustion (TTE) was the main efficacy endpoint used for sample size calculation. The sample size was calculated for a two-sample, two-tailed t-test with the level of significance (α) = 5 % (*z*_α/2_ = 1.96) and power (1-β) = 90% (*z*_β_ = 1.2816), assuming mean TTE (test) = 14.95 min and mean TTE (placebo) = 12.56 min; mean difference (*d*) = 2.39 min; equal allocation (κ = 1); and standard deviation (σ) = 2.00 [9].

Using the following formula for sample size estimation [10]:

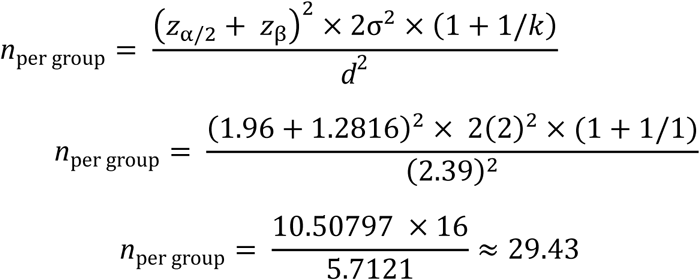

A sample size of 29.43 per group was obtained and rounded up to 30 evaluable subjects per group (a total of 60 evaluable subjects). To allow for an anticipated 15% dropout, the enrolment target was inflated:

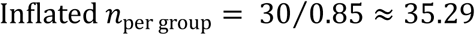

Rounded up to 36 subjects per group, a total of 72 subjects were targeted for enrolment to achieve a minimum of 60 completers.

### 2.2 Ethics and registration

#### 2.2.1 Ethical committee approval

The study was conducted at Sangvi Multispecialty Hospital and Gunjkar Multispeciality Hospital, Pune, Maharashtra, India, geographically proximal sites that operate under a shared organisational and operational framework. The study was thus approved and oversighted by a joint institutional ethical committee (IEC). There were no changes made to the protocol after approval, and participants were only recruited after approval and registration of the study.

#### 2.2.2 Study registration

After IEC approval, the study was prospectively registered online with the Clinical Trials Registry – India (CTRI) on 18 November 2025 (CTRI/2025/11/097623).

#### 2.2.3 Study conduct

The study was conducted, recorded and reported strictly in accordance with the guidelines of the Ministry of Ayurveda, Yoga and naturopathy, Unani, Siddha and Homoeopathy and Good Clinical Practice of the International Council of Harmonisation. The investigational products were prepared in compliance with Indian Good Manufacturing Practices. During conduct, the rights, safety, and well-being of the study participants were given foremost importance. There were no changes to the established trial protocol after commencement.

#### 2.2.4 Informed consent

On the initial screening visit, voluntary written informed consent from participants for their participation in the study was obtained using an informed consent form (ICF) printed in the language best understood by the participants. If the participant was illiterate, the ICF was read to them by an impartial witness in the language they best understood. The participants were informed about the objectives of the study, the design of the study, and the expected risks and benefits of participation. They were given enough time and opportunities to obtain clarifications and fully understand the study. If the participant then agreed to participate in the study, their thumb impression, along with the signature of the impartial witness (if present) was obtained on the ICF. Each participant received a copy of their ICF. The identity of the study participants was disclosed only to the primary study personnel and ethics committees. The data generated from the study were kept confidential, and participant identities were coded.

### 2.3 Study objectives

The study aimed to:

a. evaluate the impact of the test product compared to the placebo on physical performance measures, cardiorespiratory health and endurance, anthropometric measurements, hypertrophy, and body composition over 12 weeks in physically active healthy individuals.
b. assess the efficacy of the test product compared to the placebo on changes in muscle strength, biochemical markers, recovery from exercise, and laboratory investigations in physically active healthy individuals.
c. assess safety via monitoring of adverse events, tolerability, and compliance.

### 2.4 Study endpoints

The study’s primary and secondary efficacy and safety endpoints are detailed in **Table 1**.

**Table 1.** Efficacy and safety endpoints.

| Endpoint | Test and method | Outcome measure | Time frame |
| --- | --- | --- | --- |
| **Primary endpoint 1** | Cardiopulmonary function: Cardiopulmonary exercise test (CPET) | VO<sub>2</sub> max (oxygen consumption), maximum ventilation, ventilatory threshold (VT) | Baseline, week 6 and week 12 |
| **Primary endpoint 2** | Muscular strength: Leg extension and bench press through one-repetition maximum (1RM) | Muscle strength (kg) | Baseline, week 6 and week 12 |
| **Primary endpoint 3** | 4 sets at 80% 1RM load until voluntary exhaustion (muscular endurance) | Fatigue index (%) | Baseline, week 6 and week 12 |
| **Primary endpoint 4** | Muscular endurance markers | Creatine kinase, Lactate dehydrogenase | Baseline and week 12 |
| **Secondary endpoint 1** | Treadmill-based time to exhaustion test at a velocity that corresponded to 85% of the peak oxygen uptake | Time to exhaustion (TTE) | Baseline, week 6 and week 12 |
| **Secondary endpoint 2** | Physical measures (Resting vital signs assessment) | Heart rate (BPM), systolic blood pressure (mmHg), and diastolic blood pressure (mmHg), respiratory exchange ratio, and oxygen saturation | Baseline, week 6 and week 12 |
| **Secondary endpoint 3** | Anthropometric parameters and hypertrophy | Body Weight (kg), BMI (kg/m<sup>2</sup>) and hip, waist, chest circumferences (cm) | Baseline, week 6 and week 12 |
| **Secondary endpoint 4** | Body composition using bioelectric impedance analysis (BIA) | % fat and % skeletal muscle | Baseline, week 6 and week 12 |
| **Secondary endpoint 5** | 48 hours post-1RM test using leg press & bench press at 70% 1RM. | Muscle endurance (repetitions to failure) | Baseline, week 6 and week 12 |
| **Secondary endpoint 6** | Biochemical marker levels | Plasma nitrite ([NO<sub>2</sub><sup>-</sup>]), serum creatine, cortisol, serum ferritin, serum iron, blood lactate (immediately after acute exercise) | Baseline and week 12 |
| **Secondary endpoint 7** | Changes in recovery from exercise measures - post-delayed onset muscle soreness (DOMS) symptoms | Post-delayed onset muscle soreness (DOMS) is defined as an aching pain and soreness in a given muscle following a novel exercise bout. | Baseline, week 6 and week 12 |
| **Secondary endpoint 8** | Complete blood count (CBC), liver function test (LFT, AST, ALT), renal function test (RFT), lipid profile, serum electrolytes, urine analysis, etc. | Haematological and biochemical parameter levels | Baseline and week 12 |
| **Secondary endpoint 9** | Adverse events profile | - | From baseline to week 12 |
| **Secondary endpoint 10** | Compliance and tolerability | - | From baseline to week 12 |

### 2.5 Participant recruitment

**Figure 1** presents participant enrolment and progression through the study in accordance with the 2025 CONSORT guidelines. Participants who provided written informed consent, were ready to provide regular follow-ups till the completion of the study, and met the inclusion and exclusion criteria were recruited. Of 76 individuals screened in the study, four were ineligible and 72 were randomised. During the study, eight participants (four in each group) discontinued, and 64 completed the study. Data were collected from December 2025 through April 2026.

**Figure 1.**
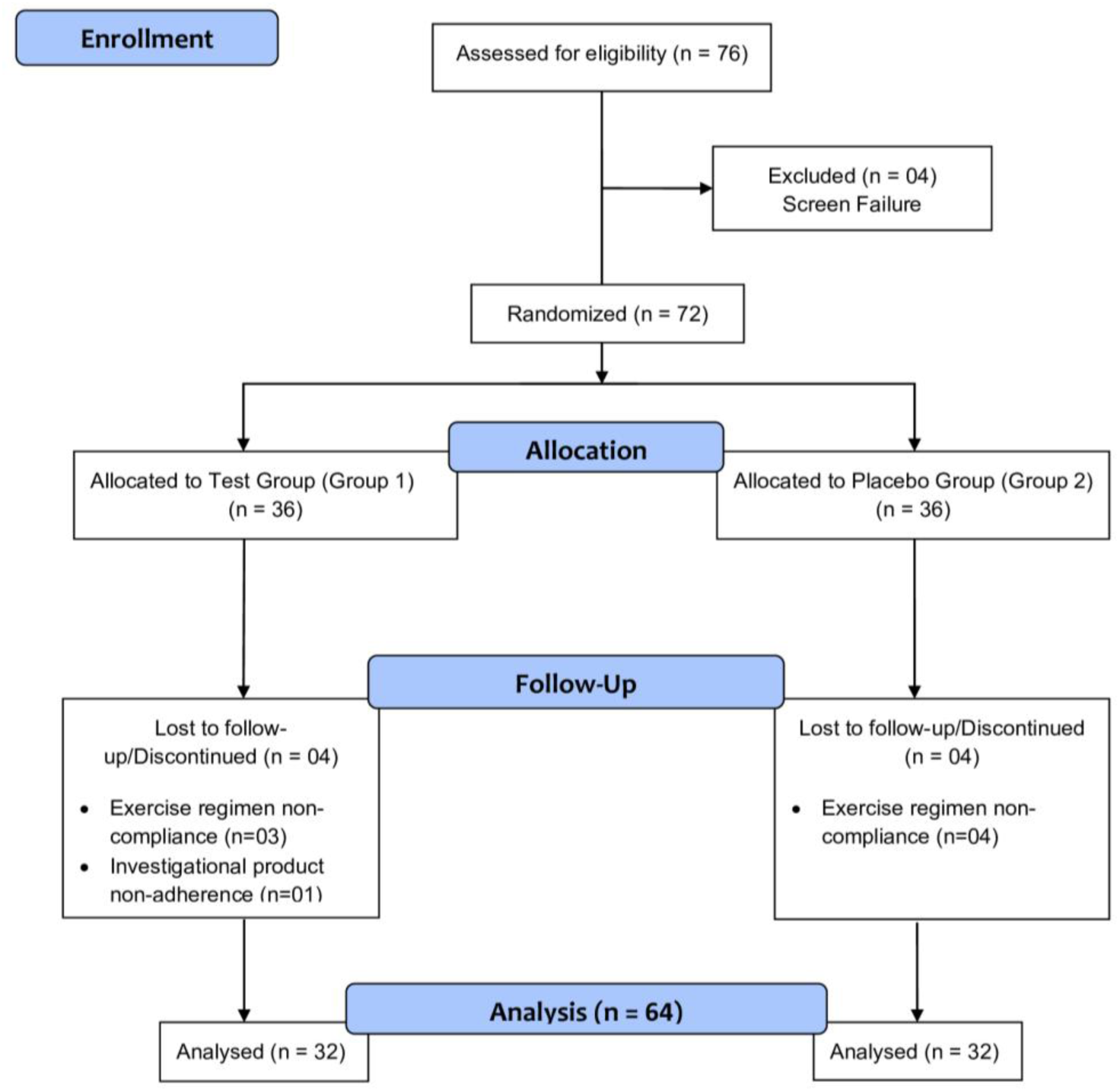
Consolidated standards of reporting trials (CONSORT) diagram for the study.

#### 2.5.1 Inclusion criteria

Participants were eligible for enrolment in the study if they were male or female adults aged 18–40 years (inclusive) with a body mass index between 25–30 kg/m² (inclusive) and a resting blood pressure, based on the average of screening assessments, of <140/90 mmHg. They were also not receiving any medication for cardiovascular or metabolic disorders and had engaged in regular resistance training, comprising both cardiovascular and strength-training exercises, for a period of six to twelve months prior to screening. Participants were required to be willing to continue their existing exercise regimen throughout the study period. In addition, they were required to be physically active and be willing to maintain a stable, healthy, and balanced diet with adequate intake of carbohydrates, proteins, and fats during the study. They were also required not to initiate any new dietary interventions, weight-management programs, restrictive diets, or fad dietary regimens during study participation. Further, they were expected to possess an adequate understanding of the importance of dietary and lifestyle factors relevant to the study objectives, as determined by the investigator. Finally, they had to be willing to discontinue the use of mouthwash products for the duration of the study, provide written informed consent prior to the conduct of any study-related procedures, and comply with all protocol requirements, scheduled follow-up visits, and assessments.

#### 2.5.2 Exclusion criteria

Participants were excluded from the study if they met any of the following criteria: presence of an acute illness requiring immediate medical attention; evidence of clinically significant hepatic dysfunction, defined as alanine aminotransferase or aspartate aminotransferase levels >3 times the upper limit of the normal; uncontrolled diabetes mellitus (HbA1c >6.5%); thyroid disorders including hypothyroidism or hyperthyroidism (thyroid-stimulating hormone [TSH] <0.5 or >4.5 mIU/L), adrenal insufficiency, or uncontrolled asthma despite medication. Participants with a history or diagnosis of major depressive disorder, bipolar disorder, schizophrenia, dementia, delusional disorders, or insomnia were also excluded. Additional exclusion criteria included anaemia (haemoglobin <13 g/dL and <12 g/dL in male and female individuals, respectively), any chronic medical condition, or a history of malignant tumours within the previous 5 years. Participants were also excluded if they consumed alcohol at levels of ≥40 g/day (male participants) or ≥20 g/day (female participants), in accordance with the WHO’s alcohol consumption criteria; smoked more than 20 cigarettes per day; used any nicotine-containing products, including tobacco, electronic cigarettes, or nicotine replacement therapies; or consumed more than 400 mg of caffeine per day from coffee, tea, energy drinks, soft drinks, supplements, or other sources. Individuals with a sedentary lifestyle who were unable or medically advised against participating in the resistance-training program specified in the protocol were not eligible. Participants presenting with any clinically relevant macro- or micronutrient deficiency, as determined by the investigator, or with any known cardiovascular or metabolic disease were also excluded. Participants who were pregnant, lactating, or those with reproductive potential not using an adequate method of contraception, or who had a positive urine pregnancy test at screening, were excluded from the study. Participants currently using herbal products or nutraceutical supplements, or those unwilling or unable to refrain from consuming nitrate-rich foods (e.g., beetroot, spinach, or similar dietary sources) during the study period, were excluded. Additional exclusion criteria included inability to perform study-required exercise procedures (e.g., treadmill testing), engagement in excessive occupational physical labor or participation as a professional athlete, known hypersensitivity or adverse reactions to any ingredient of the IP, participation in another clinical trial within three months prior to screening, or the presence of any other medical, psychological, or social condition that, in the opinion of the investigator, could compromise participant safety, study compliance, or the validity of the study results.

### 2.6 Study groups and participant allocation

The study involved two treatment groups:

a. Test group: received SpinoleRG™ capsules
b. Placebo group: received placebo capsules

36 participants were randomly allocated to the test group and placebo group each (a total of 72). 32 participants finished the study in each group (64 in total).

### 2.7 Randomisation and blinding

The random allocation of participants to either study arm was performed by a computer-generated randomisation list.

Investigators, designated study staff, participants, sponsors, and study monitors were blinded to the assigned treatments. Each study site had a designated dispenser, who was to dispense the capsules, and maintain dispensing records and logs completely and accurately. The participants were requested not to discuss the appearance of the capsules with other study participants. To minimise potential bias, the randomisation scheme was securely held by the packaging company until after the database was locked. The IPs were packaged and labelled with the participant IDs in a manner that prevented the participants from identifying the treatment.

### 2.8 Investigational products (IPs)

#### 2.8.1 Extract preparation

Fresh leaves of both plants were identified, collected, cleaned, shade-dried, and coarsely milled. The dried leaf material was extracted with ethanol-water (90:10, v/v), then filtered, concentrated, and dried.

#### 2.8.2 Extract standardisation

The composition of the resulting extract was quantitatively analysed and standardised to contain not less than (NLT) 0.50% TMMG and 5.00% nitrates, using high-performance liquid chromatography with photodiode array detection and validated reference standards. For TMMG, a 15 cm x 4.6 mm, 5 µm C18 analytical column (Phenomenex, USA) with a mobile phase of acidified water and acetonitrile was used. The flow rate was set at 0.5 mL/min and mobile phase B at 15%. The concentration of mobile phase B was raised to 70% at 18 min. TMMG was detected at 350 nm. For nitrate characterisation, a combination of buffer solution and acetonitrile was used in a Shim-pack GIST C18 column (Shimadzu Corporation, Japan). The flow rate was set at 0.8 mL/min. Ionic nitrates were detected at 222 nm (**Figure 2**).

**Figure 2.**
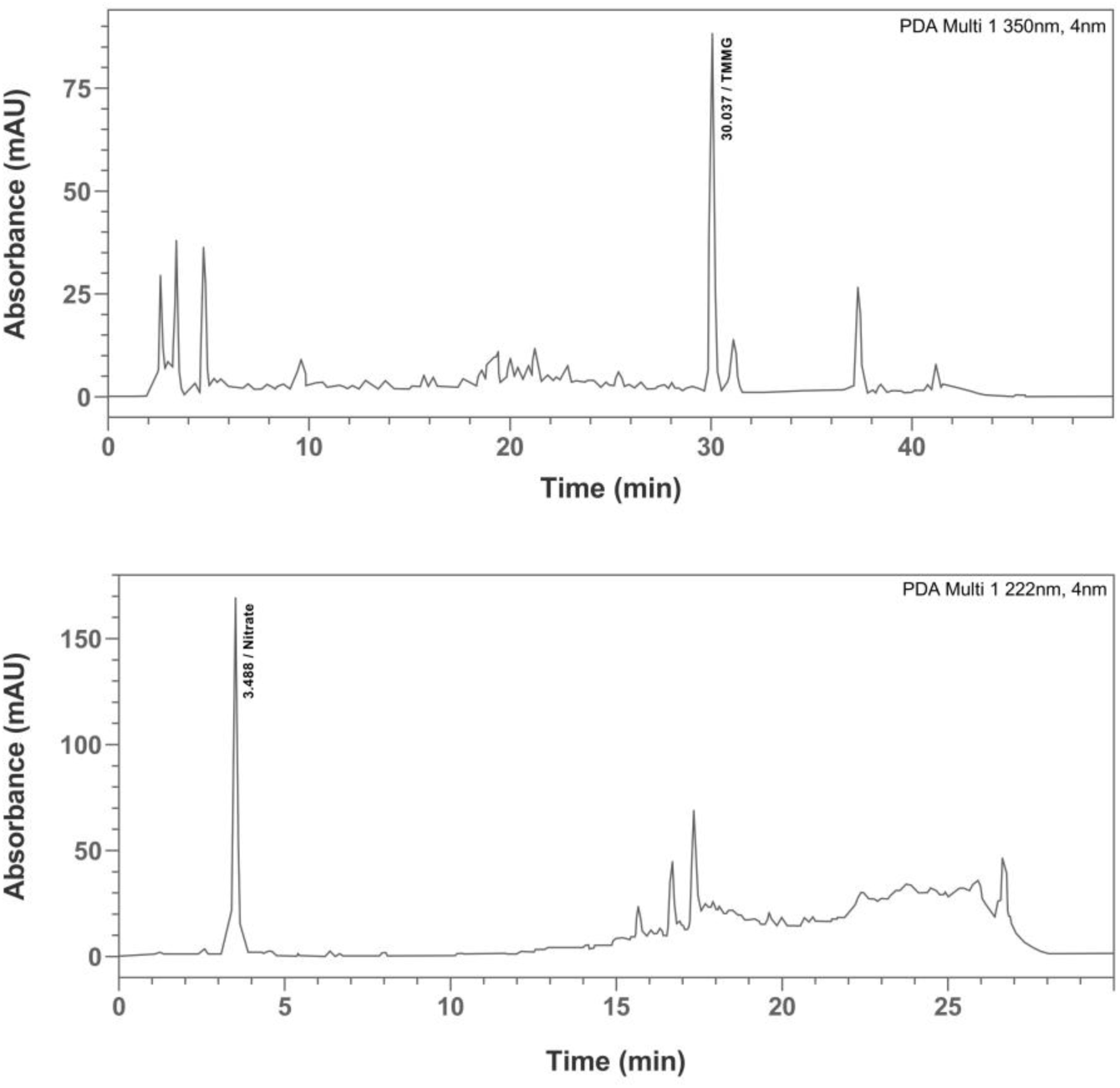
Typical chromatograms for characterisation and standardisation of TMMG and nitrate in the test extract, SpinoleRG™.

In addition, the extract was also standardised to meet heavy metal limits and to ensure passage of at least 90% of the particles through a 60-mesh sieve. It was then formulated as granules and filled into capsules, which were each standardised to contain NLT 2 mg TMMG and NLT 20 mg nitrates.

#### 2.8.3 Composition of the IPs

The test product was prepared in 800 mg capsules, consisting of 200 mg *A. tricolor* extract, 200 mg *S. oleracea* extract, and excipients q.s. (microcrystalline cellulose, maltodextrin, calcium carbonate, hydroxypropyl methylcellulose, water). The placebo was also prepared in 800 mg capsules of hydroxypropyl methylcellulose, microcrystalline cellulose, calcium carbonate, sodium starch glycolate, and croscarmellose sodium.

#### 2.8.4 Packaging and storage

The capsules were packaged in high-density polyethylene bottles, each containing 65 capsules. The bottles were labelled with information such as product code, participant IDs, manufacturing date, expiry date, storage conditions, dosage and usage instructions and the statement: ‘For Clinical Study Use Only’, then stored in closed containers below 30°C, protected from light and moisture. The storage area was secure and only accessible to authorised personnel.

### 2.9 Permitted and prohibited concomitant therapy

Treatment for concomitant medical conditions, where applicable, was permitted at the discretion of the investigator. Any concomitant medication considered necessary during the study was allowed and was documented in the case report form (CRF). Since participants with significant medical conditions were excluded from the study, the use of concomitant medications was expected to be minimal. The use of Ayurveda, homoeopathy, Siddha, Unani medicines, herbal products, nutraceutical supplements, creatine supplements, and any other supplements considered by the investigator to have the potential to influence study outcomes was prohibited during the study period.

### 2.10 Interventions

Participants received a total daily dose of 1,600 mg of SpinoleRG™ (two 800 mg capsules, each containing 200 mg of *A. tricolor* and *S.* oleracea and excipients q.s.) or matching placebo, administered in sequence 30–60 minutes before exercise. All participants received standardised counselling at baseline and during each follow-up visit, regarding adherence to a balanced diet and regular physical activity throughout the study period. Each participant received a daily diary to record smoking habits, caffeine consumption, alcohol consumption, daily protein intake, physical activity (gym-based & non-gym-based activity), and dietary indulgence (junk/processed food, sweets, fried food, or outside food).

Resting heart rate, systolic and diastolic blood pressure, respiratory exchange ratio, and oxygen saturation were assessed at baseline, week 6, and week 12. Cardiorespiratory health and endurance were evaluated through a cardiopulmonary exercise test (CPET), including maximal oxygen uptake (VO₂ max), maximum ventilation, and ventilatory threshold (VT), at baseline, week 6, and week 12. Anthropometric parameters such as body weight, BMI, and hip, waist, and chest circumferences, along with body composition (% fat and % skeletal muscle) measured by bioelectrical impedance analysis (BIA), were assessed at baseline, week 6, and week 12. Muscle strength was evaluated using one-repetition maximum (1RM) tests for leg extension and bench press. At baseline, week 6, and week 12. Muscle endurance was assessed by repetitions to failure at 70% of 1RM at baseline, week 6, and week 12, and creatine kinase (CK) and lactate dehydrogenase (LDH) levels at baseline and week 12. Fatigue index (FI) was determined through repetitions to voluntary exhaustion at 80% of 1RM. Treadmill-based time to exhaustion (TTE) at 85% of peak VO₂ was also assessed. Recovery from exercise was measured using delayed onset muscle soreness (DOMS) scores. FI, TTE and DOMS scores were assessed at baseline, week 6, and week 12. Treatment compliance and tolerability were evaluated at week 6 and week 12 to ensure adherence and monitor responses to the IPs.

### 2.11 Adverse events

An adverse event (AE) was defined as any untoward medical occurrence in a participant administered the IP, irrespective of causality. This included any unfavourable and unintended sign, symptom, disease, or abnormal laboratory finding temporally associated with the use of the IP.

#### 2.11.1 Reporting and recording adverse events

AEs were identified through participant self-reports, clinical evaluations, and laboratory assessments at the baseline, week 6, and week 12. All AEs were recorded in the AE log of the case report form.

#### 2.11.2 Follow-up

All reported AEs occurred within the treatment period and were resolved by the end of the study, thus requiring no additional follow-up beyond the scheduled end-of-study follow-up visit.

### 2.12 Tolerability and compliance

Treatment compliance and tolerability were evaluated at weeks 6 and 12 to ensure adherence and monitor responses to the intervention.

Tolerability was evaluated based on the incidence and severity of AEs related to the investigational products using the following scoring criteria: 0 (poor tolerability): severe AEs necessitating discontinuation; 1 (fair tolerability): moderate to severe AEs, which resolved with or without medication and did not require discontinuation; 2 (good tolerability): mild AEs related to the IP, which resolved with or without medication and 3 (excellent tolerability): no related AE.

Compliance with IP intake, lifestyle restrictions and physical activity requirements was recorded in the CRF for each participant. Participants missing more than four consecutive or cumulative days of IP consumption within a six-week period were considered dropouts. Additionally, participants exceeding the protocol-defined limits on alcohol (≥40 g/day for males, ≥20 g/day for females), caffeine (>400 mg/day), or smoking/nicotine use (>20 cigarettes/day or use of any nicotine-containing products for more than four cumulative or consecutive days within a six-week period) were considered dropouts. Participants were required to perform gym-based exercise at least four times per week and would be withdrawn upon failure to meet this requirement for two consecutive weeks or cumulatively within six weeks.

### 2.13 Statistical analysis

All statistical analyses were performed using IBM SPSS Statistics version 30.0. All statistical tests were two-tailed with α set at 0.05. A *p*-value of <0.05 was considered significant. Analyses were based on available observations at each assessment time point, with no imputation for missing data. Continuous variables were summarized using descriptive statistics: number of observations (n), mean, standard deviation (SD), median, minimum, and maximum. Categorical variables were summarized using frequency counts and percentages.

Efficacy endpoints were analysed using the per-protocol (PP) population, and safety variables using the modified intention-to-treat analysis. Efficacy endpoints showing substantial deviation from normality were analysed using appropriate non-parametric methods. The eight participants excluded from the PP population were therefore not included in the primary PP efficacy analyses. No formal adjustment for multiplicity across the multiple efficacy endpoints was applied; therefore, reported *p*-values were interpreted using a two-sided α of 0.05. Primary efficacy endpoints were analysed using a linear mixed-effects model (LMM) with ‘treatment group’, ‘visit’, and ‘treatment-by-visit’ interactions as fixed effects and ‘participant’ as a random effect, while secondary efficacy endpoints were analysed using an LMM with ‘treatment group’ and ‘visit’ as fixed effects and ‘participant’ as a random effect, to account for repeated measurements. For non-normally distributed variables in either primary or secondary analyses, non-parametric methods, including the Mann–Whitney *U* test for between-group comparisons, were applied. Changes from baseline in muscular endurance biomarkers were analysed with analysis of covariance (ANCOVA), with treatment group as a fixed factor and baseline value as a covariate. Between-group comparisons of biochemical markers and haematological and biochemical safety parameters were also analysed using ANCOVA with the inclusion of the week 12 value as the dependent variable. Recovery from exercise was analysed using an LMM with ‘treatment group’, ‘visit’, and ‘treatment-by-visit’ interaction as fixed effects and ‘participant’ as a random effect. The Shapiro–Wilk test was used to assess data normality.

## 3 Results

### 3.1 Demographic characteristics

The test and placebo groups were similar in age and sex distribution at baseline. Mean age was 31.50 ± 5.28 years in the test group and 31.69 ± 5.18 years in the placebo group (*p* = 0.884); sex distributions also did not differ significantly (**Table 2**).

**Table 2.** Assessment of demographic characteristics.

| Parameter | Test | Placebo | *p*-value<br>(between groups) |
| --- | --- | --- | --- |
| Age (years) | $31.50 \pm 5.28$ | $31.69 \pm 5.18$ | 0.884 |
| Sex (male/female) | 24 / 08 | 25 / 07 | 0.767 |

### 3.2 Primary efficacy endpoints

#### 3.2.1 CPET

The effects of SpinoleRG™ supplementation on CPET parameters, including VO₂ max, maximal ventilation, and VT, were evaluated over the 12-week intervention period. At baseline, no statistically significant differences were observed between the test and placebo groups (VO₂ max: *p* = 0.936; maximal ventilation: *p* = 0.777; VT: *p* = 0.767). VO₂ max and maximum ventilation data met the assumptions for LMM analysis. VT data were not normally distributed and were analysed with the Mann–Whitney *U* test. *p*-values for week 6 and week 12 reflect comparisons of change from baseline to the respective visit.

In the test group, mean VO₂ max increased from 2.10 ± 0.41 L/min at baseline to 2.23 ± 0.43 L/min at week 12, compared with an increase from 2.09 ± 0.45 L/min to 2.12 ± 0.45 L/min in the placebo group. Maximal ventilation increased from 79.26 ± 14.91 L/min at baseline to 80.73 ± 15.63 L/min at week 12 in the test group, whereas values decreased from 80.61 ± 21.41 L/min to 74.59 ± 20.45 L/min in the placebo group (**Table 3**). However, neither of these changes were significant. By contrast, the change in VT from baseline to week 12 differed significantly between groups (*p* = 0.016) (**Table 4**).

**Table 3.** Assessment of VO_2_ max and maximum ventilation.

| Parameter | Timepoint | Test<br>[95%CI] | Placebo<br>[95%CI] | *p*-value<br>(between groups) |
| --- | --- | --- | --- | --- |
| VO<sub>2</sub> max (L/min) | Baseline | 2.10 ± 0.41<br>[1.94, 2.27] | 2.09 ± 0.45<br>[1.95, 2.29] | 0.936 |
|  | Week 6 | 2.12 ± 0.40<br>[1.97, 2.28] | 2.05 ± 0.43<br>[1.86, 2.20] | — |
|  | Week 12 | 2.23 ± 0.43<br>[2.09, 2.42] | 2.12 ± 0.45<br>[1.99, 2.35] | — |
|  | Mean difference<br>(Baseline – week 6) | 0.02 ± 0.27 | 0.09 ± 0.28 | — |
|  | Mean difference<br>(Baseline – week 12) | 0.13 ± 0.36 | 0.03 ± 0.45 | — |
| Fixed effect | Treatment |  |  | 0.868 |
|  | Visit |  |  | 0.966 |
|  | Treatment*Visit |  |  | 0.973 |
| Max ventilation (L/min) | Baseline | 79.26 ± 14.91<br>[73.14, 85.08] | 80.61 ± 21.41<br>[72.94, 89.07] | 0.777 |
|  | Week 6 | 75.50 ± 13.93<br>[70.10, 80.90] | 72.39 ± 18.90<br>[64.83, 80.38] | — |
|  | Week 12 | 80.73 ± 15.63<br>[74.98, 87.42] | 74.59 ± 20.45<br>[68.60, 85.12] | — |
|  | Mean difference<br>(Baseline – week 6) | 3.61 ± 13.60 | 8.22 ± 12.70 | — |
|  | Mean difference<br>(Baseline – week 12) | 1.47 ± 17.62 | 6.03 ± 18.80 | — |
| Fixed effect | Treatment |  |  | 0.872 |
|  | Visit |  |  | 0.930 |
|  | Treatment*Visit |  |  | 0.921 |

<sup>1</sup> Statistically significant at *p* < 0.05. LMM; linear mixed effects model. 95% CI values are provided in square brackets.

**Table 4.** Ventilatory threshold (VT) assessment.

| Parameter | Timepoint | Test<br>[95%CI] | Placebo<br>[95%CI] | *p*-value<br>(between groups) |
| --- | --- | --- | --- | --- |
| Ventilatory threshold (VT) | Baseline | 1.81 ± 0.38<br>[1.67, 1.97] | 1.81 ± 0.49<br>[1.64, 2.04] | 0.701 |
|  | week 6 | 1.78 ± 0.35<br>[1.64, 1.91] | 1.73 ± 0.35<br>[1.58, 1.86] | — |
|  | week 12 | 1.92 ± 0.39<br>[1.79, 2.08] | 1.74 ± 0.38<br>[1.60, 1.89] | — |
|  | Mean difference<br>(Baseline – week 6) | 0.04 ± 0.21 | 0.11 ± 0.34 | 0.625 |
|  | Mean difference<br>(Baseline – week 12) | 0.11 ± 0.28 | 0.07 ± 0.42 | 0.016* |

<sup>1</sup> Statistically significant at *p* < 0.05. The non-parametric Mann-Whitney *U* test was used for assessment. *p*-values for week 6 and week 12 reflect comparisons of change from the baseline to the respective visit. 95% CI values are provided in square brackets.

#### 3.2.2 Muscular strength – 1 RM

1RM data for bench and leg presses were not normally distributed and did not meet the assumptions required for LMM analysis. Therefore, a Mann–Whitney *U* test was used. *p*-values for week 6 and week 12 reflect comparisons of change from baseline to the respective visit.

At 12 weeks, mean 1RM bench press increased from 71.12 ± 8.26 kg at baseline to 77.67 ± 6.68 kg at week 12 in the test group, compared with an increase from 70.60 ± 7.91 kg to 72.90 ± 6.65 kg in the placebo group, with a statistically significant between-group difference (*p* = 0.007). Mean 1RM leg press increased from 90.00 ± 7.30 kg at baseline to 97.16 ± 6.23 kg in the test group, compared with an increase from 88.79 ± 8.84 kg to 92.59 ± 7.56 kg in the placebo group, with a statistically significant between-group difference (*p* = 0.0001) (**Figure 3**).

**Figure 3.**
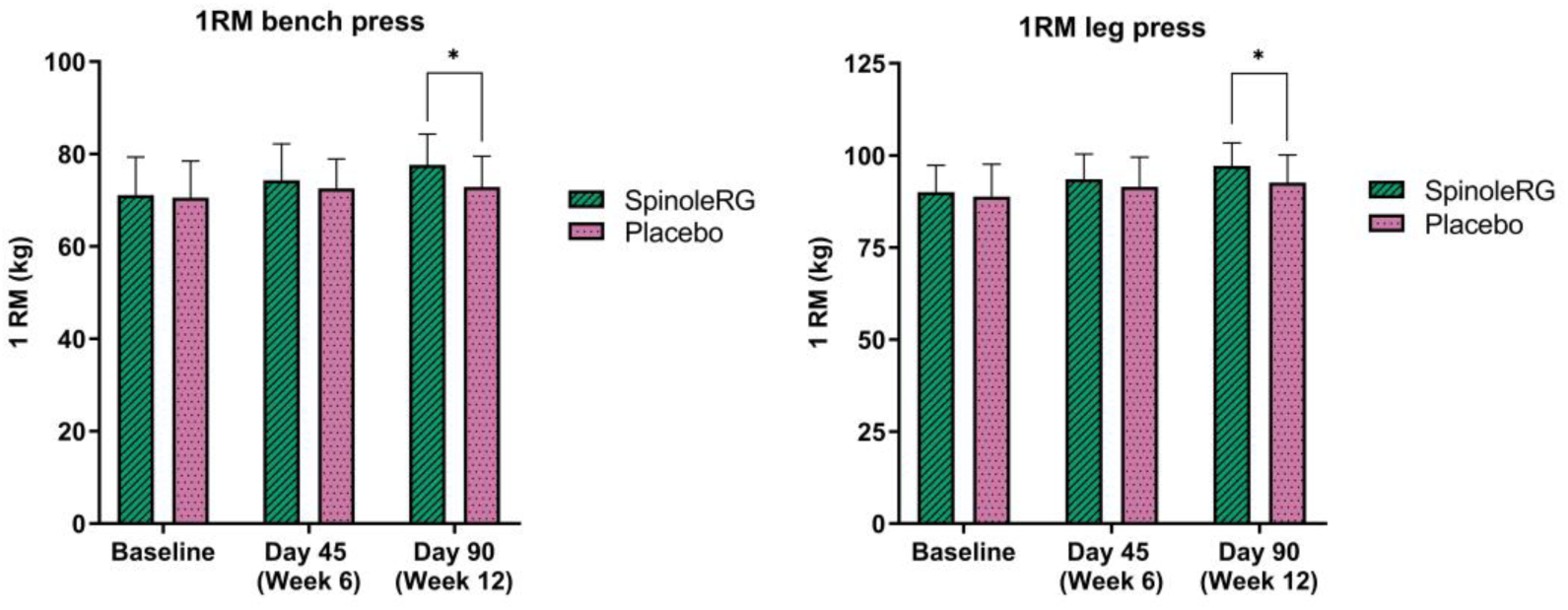
One-repetition maximum (1RM) bench and leg press results (in kg) in the test and placebo groups over 12 weeks. \**p* <0.05

#### 3.2.3 Muscle endurance - FI

FI data were analysed with a Mann-Whitney *U* test due to a non-normal distribution. *p*-values for week 6 and week 12 reflect comparisons of change from baseline to the respective visit.

At baseline, FI values for both leg press and bench press were comparable between the test and placebo groups (*p* > 0.05). For bench press FI, mean values decreased from 31.84 ± 5.53% at baseline to 22.68 ± 6.72% at week 12 in the test group, compared with a decrease from 31.39 ± 6.29% to 29.27 ± 7.92% in the placebo group. Statistically significant between-group differences were observed at week 6 (*p* = 0.035) and week 12 (*p* = 0.001). For the leg press fatigue index, mean values decreased from 23.42 ± 5.76% at baseline to 16.93 ± 4.13% at week 12 in the test group, compared with a decrease from 23.38 ± 4.49% to 22.08 ± 6.77% in the placebo group, with a statistically significant between-group difference (*p* = 0.012) (**Figure 4**).

**Figure 4.**
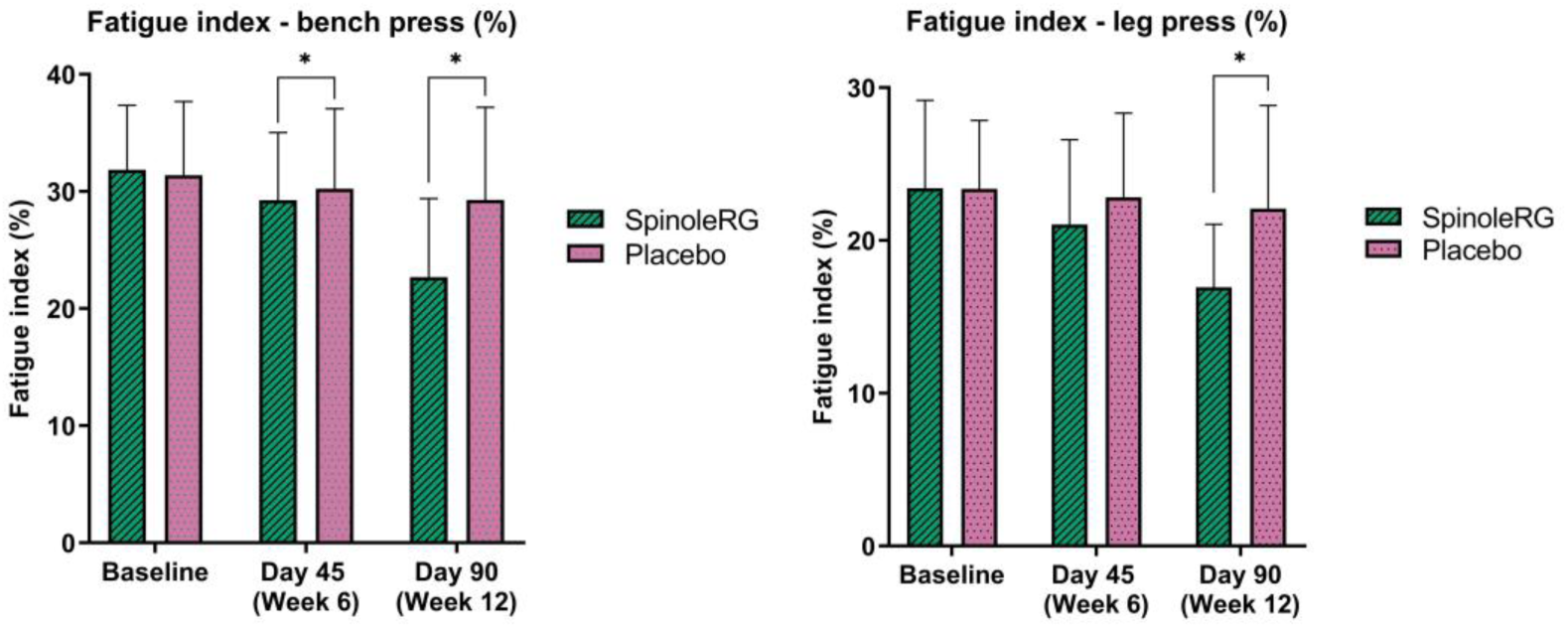
Fatigue indices for bench and leg press in the test and placebo groups over 12 weeks. \**p* <0.05

#### 3.2.4 Muscle endurance markers

Data were analysed by the Mann-Whitney *U* test due to being non-normally distributed. *p*-values for week 12 reflect comparisons of change from baseline to the respective visit.

CK and LDH were measured as indices of exercise-induced muscle damage. At baseline, both groups were comparable for CK (test: 122.52 ± 44.58 U/L; placebo: 118.05 ± 32.58 U/L; *p* = 0.914) and LDH (test: 234.88 ± 22.70 U/L; placebo: 237.93 ± 50.93 U/L; *p* = 0.327). At week 12, the test group demonstrated a 14.4% reduction in CK levels (from 122.52 to 104.87 U/L), compared to only a 5.9% reduction in the placebo group (from 118.05 to 111.03 U/L), with a statistically significant between-group difference (*p* = 0.001). Similarly, LDH declined by 13.3% in the test group (from 234.88 to 203.69 U/L), markedly greater than the 8.2% reduction observed in the placebo group (from 237.93 to 218.34 U/L; *p* = 0.001) (**Table 5**).

**Table 5.**
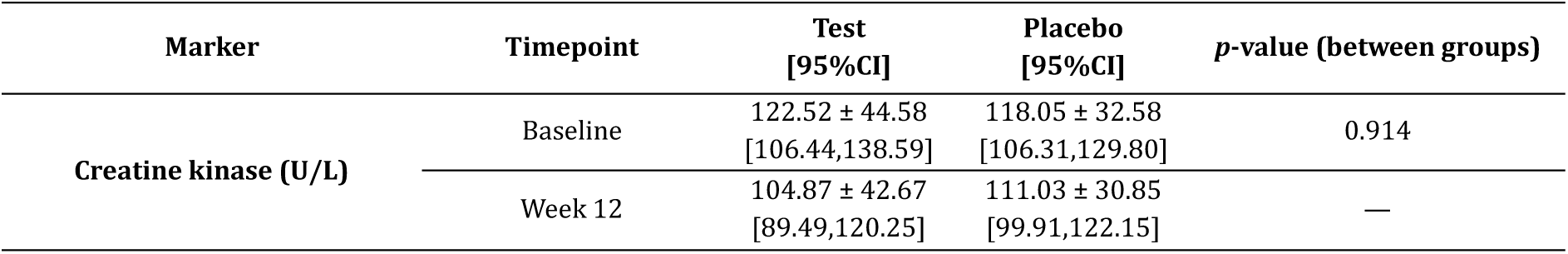
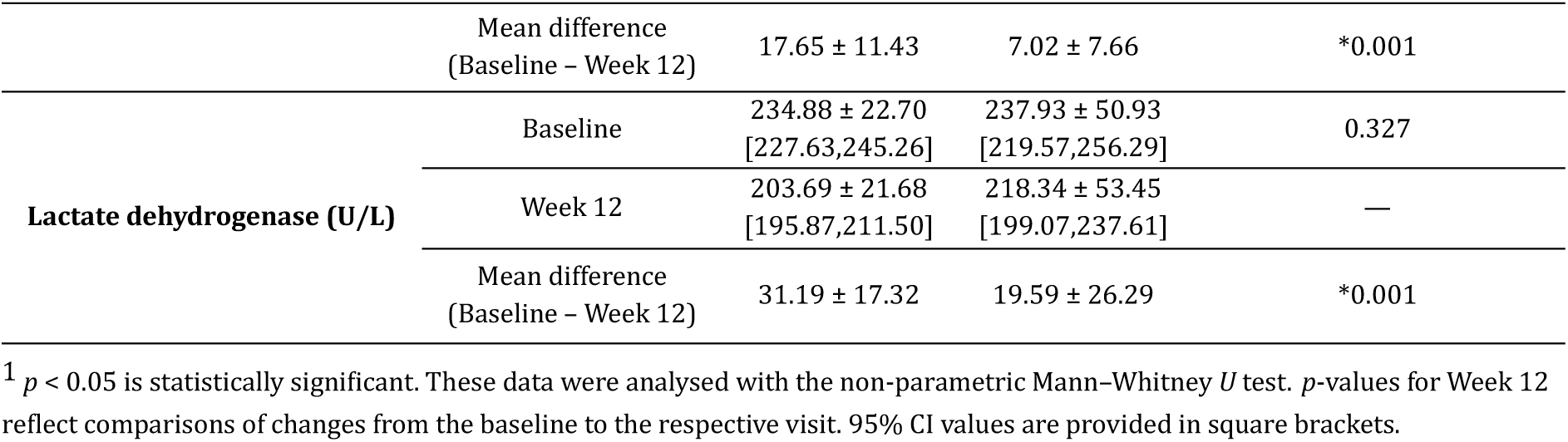
Assessment of muscle endurance markers.

| Marker | Timepoint | Test<br>[95%CI] | Placebo<br>[95%CI] | $p$-value (between groups) |
| --- | --- | --- | --- | --- |
| Creatine kinase (U/L) | Baseline | $122.52 \pm 44.58$<br>[106.44,138.59] | $118.05 \pm 32.58$<br>[106.31,129.80] | 0.914 |
| | Week 12 | $104.87 \pm 42.67$<br>[89.49,120.25] | $111.03 \pm 30.85$<br>[99.91,122.15] | — |
|  | Mean difference<br>(Baseline – Week 12) | 17.65 ± 11.43 | 7.02 ± 7.66 | *0.001 |
| Lactate dehydrogenase (U/L) | Baseline | 234.88 ± 22.70<br>[227.63,245.26] | 237.93 ± 50.93<br>[219.57,256.29] | 0.327 |
|  | Week 12 | 203.69 ± 21.68<br>[195.87,211.50] | 218.34 ± 53.45<br>[199.07,237.61] | — |
|  | Mean difference<br>(Baseline – Week 12) | 31.19 ± 17.32 | 19.59 ± 26.29 | *0.001 |

<sup>1</sup> $p < 0.05$ is statistically significant. These data were analysed with the non-parametric Mann-Whitney $U$ test. $p$ -values for Week 12 reflect comparisons of changes from the baseline to the respective visit. 95% CI values are provided in square brackets.

### 3.3 Secondary efficacy endpoints

#### 3.3.1 TTE

The Mann-Whitney *U* test was used for TTE data analysis due to a non-normal distribution. *p*-values for week 6 and week 12 reflect comparisons of change from baseline to the respective visit.

At week 12, the extract effected a statistically significant increase in the mean TTE over the placebo, from 8.07 ± 1.26 min at baseline to 10.45 ± 1.52 min in the test group, and from 7.65 ± 1.08 minutes to 8.61 ± 1.18 minutes in the placebo group. Statistically significant between-group differences were observed at week 6 (*p* = 0.004) and week 12 (*p* = 0.001) (**Figure 5**).

**Figure 5.**
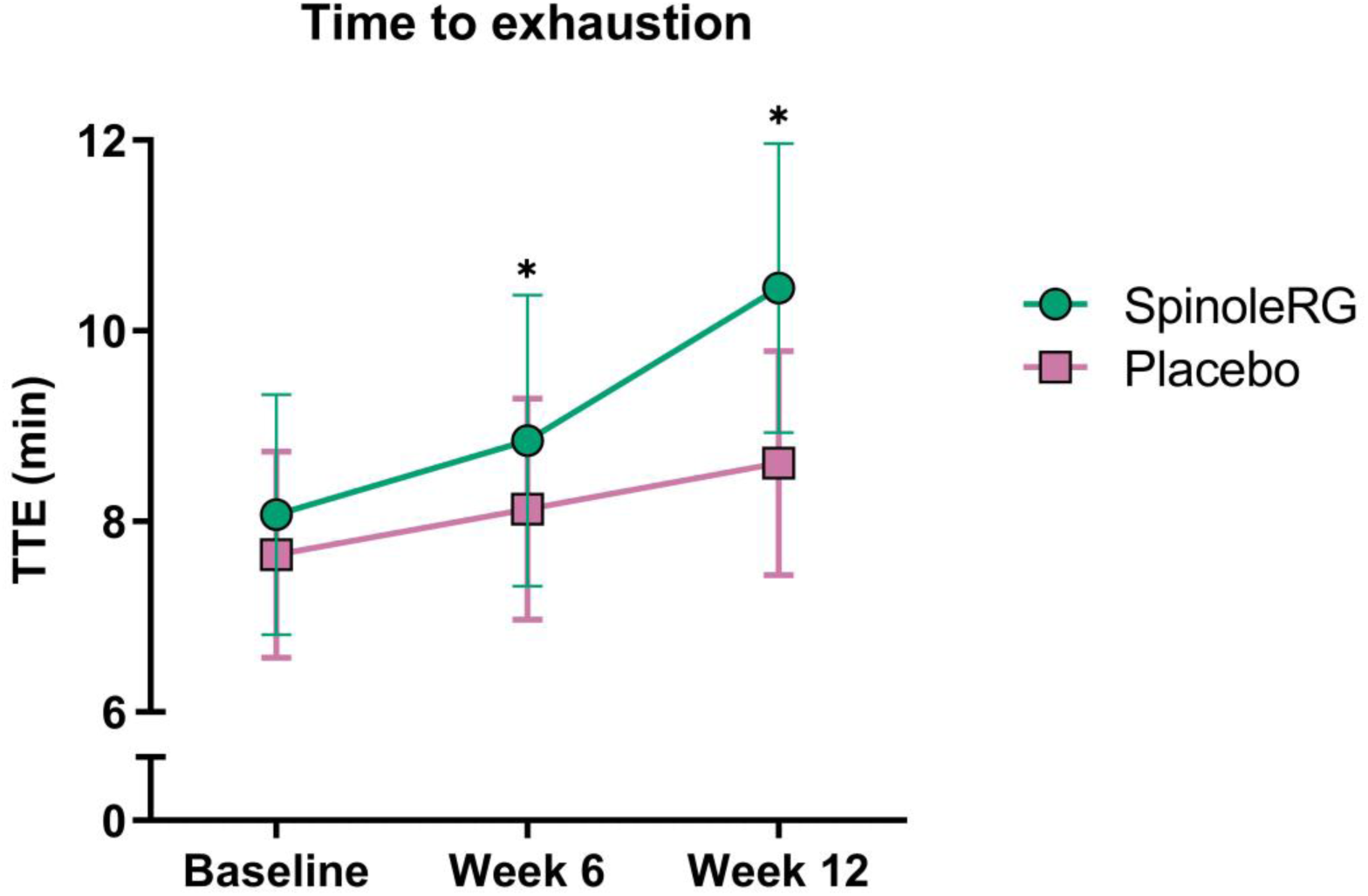
Mean time to exhaustion (TTE) in the test and placebo groups over 12 weeks. \**p* <0.05

#### 3.3.2 Anthropometric and body composition outcomes

Chest circumference and skeletal muscle percentage data met the assumptions for parametric analysis and were analysed using the LMM, whereas body weight, BMI, hip circumference, waist circumference, and body fat percentage data were not normally distributed and were analysed by a Mann–Whitney *U* test. *p*-values for week 6 and week 12 reflect comparisons of change from baseline to the respective visit. At baseline, chest circumference was marginally higher in the test group (*p* = 0.050). At week 12, mean body weight increased from 74.65 ± 6.02 kg at baseline to 75.80 ± 5.73 kg, while body fat percentage decreased from 18.54 ± 5.85% to 16.88 ± 4.90% and skeletal muscle percentage increased from 38.41 ± 4.94% to 39.70 ± 5.24%. In addition, reductions in waist circumference (85.31 ± 6.59 cm to 83.38 ± 6.35 cm) and hip circumference (93.53 ± 6.46 cm to 91.88 ± 5.74 cm), together with an increase in chest circumference (91.09 ± 4.89 cm to 93.44 ± 3.50 cm), were observed in the test group. Corresponding changes in the placebo group were generally less pronounced.

In the test group, statistically significant between-group differences were observed for body weight and BMI at week 6, and for body fat percentage at both week 6 and week 12. However, LMM analyses did not demonstrate significant treatment, time, or treatment-by-time interaction effects for chest circumference or skeletal muscle percentage. The test group demonstrated a progressive reduction in body fat percentage of 8.9% from baseline to week 12, compared to 3.9% in the placebo group, with significant between-group differences at both week 6 (*p* = 0.001) and week 12 (*p* = 0.019). Skeletal muscle percentage increased by 3.36% from baseline to week 12 in the test group, compared with a 1.66% increase in the placebo group, though this was not statistically significant (**Table 6** and **Table S1**).

**Table 6.** Assessment of chest circumference (cm) and skeletal muscle (%).

| Parameter | Timepoint | Test<br>[95%CI] | Placebo<br>[95%CI] | *p*-value (between<br>groups) |
| --- | --- | --- | --- | --- |
| Chest circumference (cm) | Baseline | $91.09 \pm 4.89$<br>[89.33, 92.86] | $89.06 \pm 3.00$<br>[87.98, 90.15] | *0.050 |
| | Week 6 | $93.09 \pm 3.71$<br>[91.76, 94.43] | $89.91 \pm 2.64$<br>[88.95, 90.86] | — |
| | Week 12 | $93.44 \pm 3.50$<br>[92.18, 94.70] | $90.31 \pm 2.63$<br>[89.36, 91.26] | — |
| | Mean difference<br>(Baseline – Week 6) | $2.00 \pm 4.03$ | $0.84 \pm 1.02$ | — |
| | Mean difference<br>(Baseline – Week 12) | $2.34 \pm 4.07$ | $1.25 \pm 1.08$ | — |

| Fixed effect | Treatment |  |  | 0.414 |
| --- | --- | --- | --- | --- |
|  | Visit |  |  | 0.834 |
|  | Treatment*Visit |  |  | 0.938 |
| Skeletal muscle (%) | Baseline | 38.41 ± 4.94<br>[36.63, 40.19] | 37.42 ± 4.62<br>[35.75, 39.08] | 0.408 |
|  | Week 6 | 39.35 ± 5.20<br>[37.47, 41.22] | 37.74 ± 4.66<br>[36.06, 39.42] | — |
|  | Week 12 | 39.70 ± 5.24<br>[37.81, 41.59] | 38.04 ± 4.56<br>[36.39, 39.68] | — |
|  | Mean difference<br>(Baseline – Week 6) | 0.93 ± 1.05 | 0.33 ± 0.76 | — |
|  | Mean difference<br>(Baseline – Week 12) | 1.29 ± 1.14 | 0.62 ± 1.00 | — |
| Fixed effect | Treatment |  |  | 0.751 |
|  | Visit |  |  | 0.975 |
|  | Treatment*Visit |  |  | 0.989 |

<sup>1</sup> Statistically significant at $p < 0.05$ . 95% CI values are provided in square brackets.

#### 3.3.3 Muscle endurance – repetitions to failure

These data, not being normally distributed, were analysed with the Mann– Whitney *U* test. *p*-values for week 6 and week 12 reflect comparisons of change from baseline to the respective visit.

Muscle endurance improved significantly in both groups, with the test group showing substantially greater gains. In bench press, a mean increase of +3.22 ± 1.24 repetitions versus +1.00 ± 0.72 in the placebo was observed (between-group *p* = 0.001). For leg press, the test product improved repetitions to +3.78 ± 1.81 versus +1.28 ± 0.68 in the placebo (between-group *p* = 0.001) (**Table 7**). These results are consistent with the concurrent fatigue index and biomarker data.

**Table 7.**
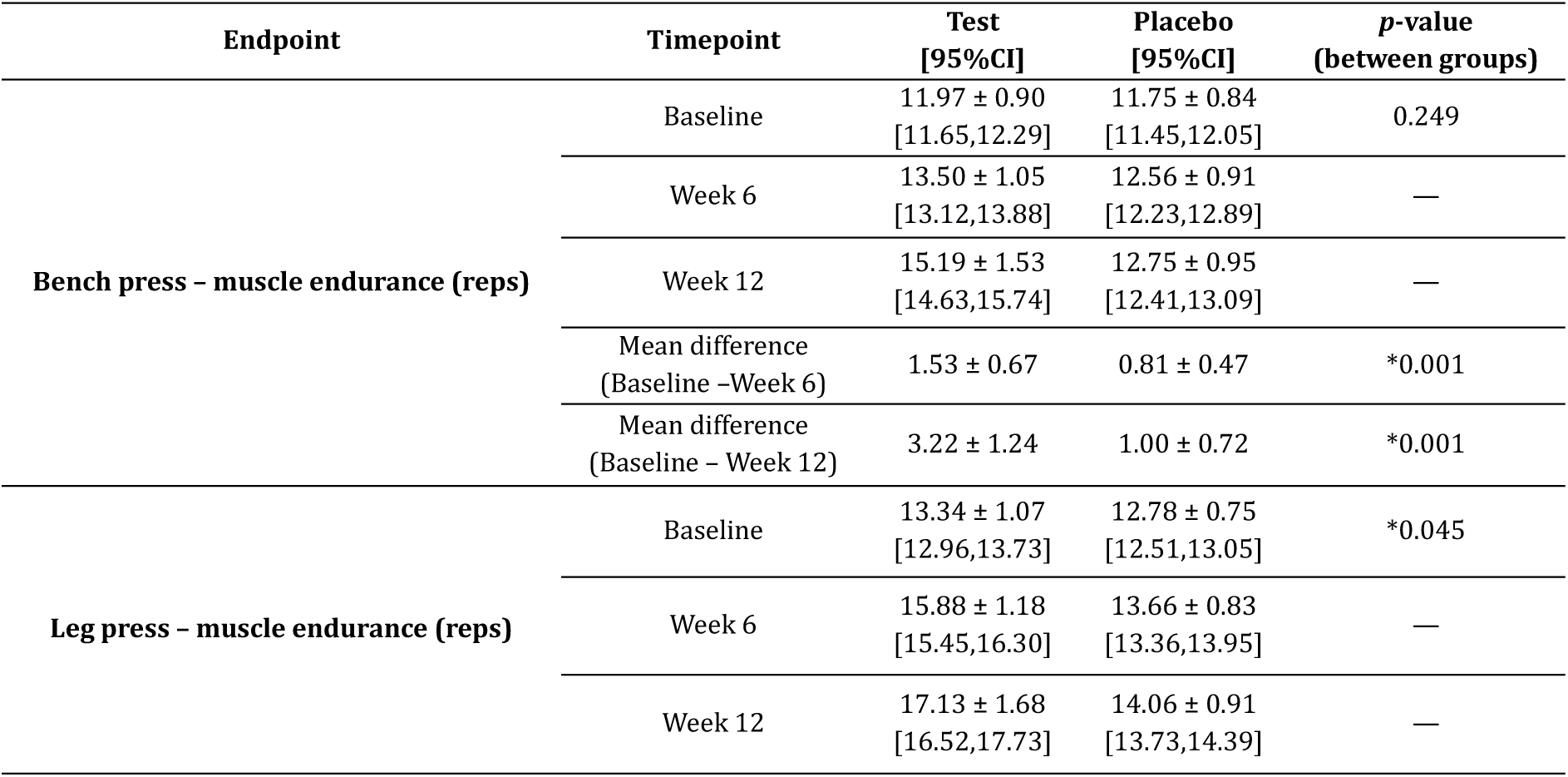
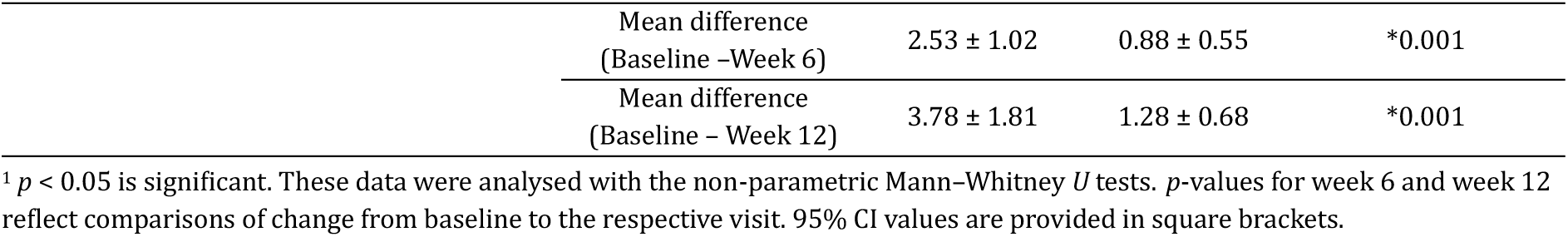
Assessment of repetitions to failure at bench and leg press.

#### 3.3.4 Biochemical markers

Blood lactate, cortisol, serum ferritin, and serum iron were analysed using the Mann-Whitney *U* test, while plasma nitrite and serum creatine were analysed using ANCOVA. *p*-values for week 12 reflect comparisons of change from baseline to the respective visit.

Blood lactate declined markedly in the test group by week 12 (68.76 to 40.06 mg/dL; ∼41.7% reduction) while remaining virtually unchanged in the placebo (72.87 to 71.32 mg/dL; ∼2.1% reduction) (between-group *p* = 0.001). Plasma nitrite increased by 42.6% in the test group (15.68 to 22.36 µmol/L) vs. a modest 6.1% rise in the placebo (13.85 to 14.69 µmol/L; *p* = 0.001). Serum creatine rose substantially in the test group (4.12 to 5.84 nmol/µL; ∼41.7% increase) compared to a negligible change in the placebo (3.88 to 4.16 nmol/µL; ∼7.2% increase; *p* = 0.001) (**Figure 6**). Cortisol levels declined in both groups, with a marginally greater reduction in the test group (145.97 to 127.49 nmol/L; ∼12.7%) vs. the placebo (144.01 to 132.27 nmol/L; ∼8.2%), with a significant between-group difference (*p* = 0.011). Serum ferritin increased in both groups at week 12, but the between-group difference was not significant (*p* = 0.405) (**Table 8**).

**Figure 6.**
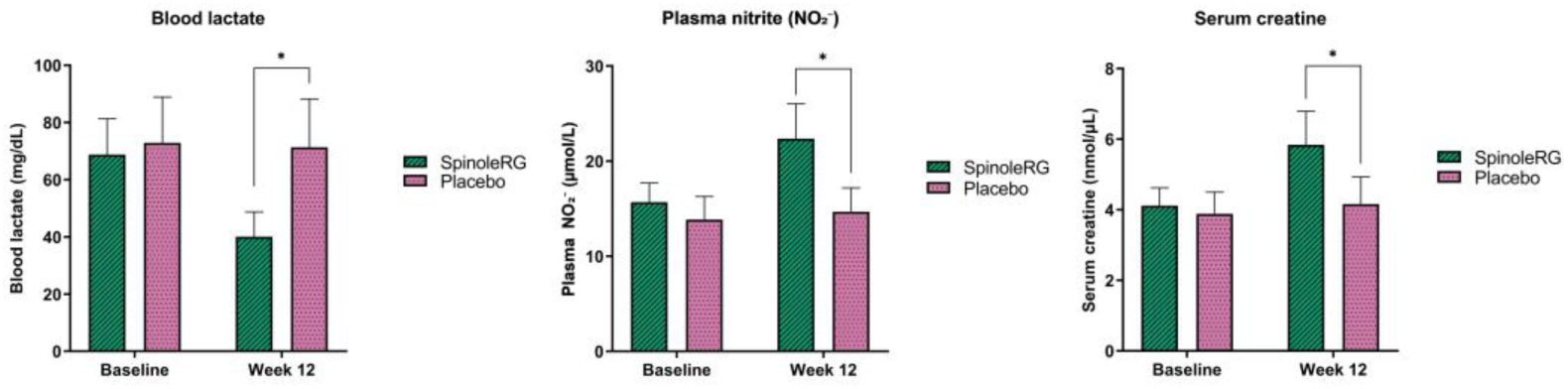
Levels of metabolically relevant biochemical markers in the test and placebo groups, measured at baseline and week 12. \**p* <0.05

**Table 8.** Assessment of biochemical markers.

| Biomarker | Timepoint | Test<br>[95%CI] | Placebo<br>[95%CI] | $p$-value<br>(between groups) |
| --- | --- | --- | --- | --- |
| **Blood lactate (mg/dL)** | Baseline | 68.76 ± 12.54<br>[64.24, 73.29] | 72.87 ± 16.02<br>[67.09, 78.64] | — |
|  | Week 12 | 40.06 ± 8.69<br>[36.93, 43.19] | 71.32 ± 16.94<br>[65.21, 77.43] | — |
|  | Mean difference<br>(Baseline – week 12) | 28.71 ± 15.25 | 1.55 ± 9.14 | *0.001 |
| **Plasma nitrite [NO<sub>2</sub><sup>-</sup>] (μmol/L)** | Baseline | 15.68 ± 2.03<br>[14.95, 16.41] | 13.85 ± 2.43<br>[12.97, 14.72] | — |
|  | Week 12 | 22.36 ± 3.68<br>[21.03, 23.69] | 14.69 ± 2.51<br>[13.78, 15.59] | — |
|  | Mean difference<br>(Baseline – week 12) | 6.69 ± 3.47 | 0.84 ± 1.04 | *0.001 |
| **Serum creatine (nmol/μL)** | Baseline | 4.12 ± 0.50<br>[3.93, 4.30] | 3.88 ± 0.62<br>[3.66, 4.10] | — |
|  | Week 12 | 5.84 ± 0.95<br>[5.49, 6.18] | 4.16 ± 0.77<br>[3.88, 4.43] | — |
|  | Mean difference<br>(Baseline – week 12) | 1.72 ± 0.90 | 0.28 ± 0.44 | *0.001 |
| **Cortisol (nmol/L)** | Baseline | 145.97 ± 53.30<br>[126.75, 165.18] | 144.01 ± 48.70<br>[126.46, 161.57] | 0.995 |
|  | Week 12 | 127.49 ± 51.63<br>[108.88, 146.11] | 132.27 ± 50.21<br>[114.17, 150.38] | — |
|  | Mean difference<br>(Baseline – week 12) | 18.48 ± 15.75 | 11.74 ± 29.51 | *0.011 |
| **Serum ferritin (ng/mL)** | Baseline | 41.24 ± 46.59<br>[24.44, 58.04] | 41.64 ± 57.83<br>[20.79, 62.49] | 0.596 |
|  | Week 12 | 51.50 ± 63.07<br>[28.76, 74.24] | 60.21 ± 62.13<br>[37.81, 82.60] | — |
|  | Mean difference<br>(Baseline – Week 12) | 10.25 ± 79.69 | 18.57 ± 90.74 | 0.405 |
| **Serum iron (μg/dl)** | Baseline | 80.03 ± 36.75<br>[66.64, 92.97] | 70.55 ± 36.75<br>[57.34, 83.81] | 0.274 |
|  | Week 12 | 97.91 ± 32.51<br>[86.19, 109.63] | 105.88 ± 37.69<br>[92.29, 119.47] | — |
|  | Mean difference<br>(Baseline – Week 12) | 17.88 ± 50.86 | 35.34 ± 45.05 | 0.100 |

<sup>1</sup> $p < 0.05$ is statistically significant. These data were analysed with the non-parametric ANCOVA and Mann-Whitney $U$ tests. 95% CI values are provided in square brackets.

#### 3.3.5 Recovery from exercise – DOMS

DOMS data were not normally distributed, requiring the Mann–Whitney *U* test for analysis. *p*-values for week 6 and week 12 reflect comparisons of change from baseline to the respective visit.

The extract effected a statistically significant reduction in exercise-induced muscle soreness and pain across both upper and lower body muscle groups. At week 12, upper body soreness scores (0–10 VAS) declined by a mean of 2.47 ± 1.05 points in the test vs. 1.28 ± 0.99 points in the placebo (between-group *p* = 0.001). Upper body pain scores declined by 1.28 ± 0.73 points in the test vs. 0.75 ± 0.57 points in the placebo (between-group *p* = 0.003). Lower body soreness decreased by 2.38 ± 1.13 points in the test versus 0.81 ± 0.82 points in the placebo (between-group *p* = 0.001), and pain scores by 1.53 ± 0.72 vs. 0.63 ± 0.61 points, respectively (between-group *p* = 0.001) (**Table 9**).

**Table 9.** Assessment of delayed onset muscle soreness (DOMS) across body regions.

| DOMS Parameter | Timepoint | Test<br>[95%CI] | Placebo<br>[95%CI] | *p*-value (between groups) |
| --- | --- | --- | --- | --- |
| Upper body soreness<br>(0–10) | Baseline | 5.72 ± 0.85<br>[5.41, 6.03] | 5.66 ± 0.75<br>[5.39, 5.93] | 0.724 |
|  | Week 6 | 4.69 ± 0.69<br>[4.44, 4.94] | 5.00 ± 0.72<br>[4.74, 5.26] | — |
|  | Week 12 | 3.25 ± 0.88<br>[2.93, 3.57] | 4.38 ± 0.61<br>[4.16, 4.59] | — |
|  | Mean difference<br>(Baseline – week 6) | 1.03 ± 0.74 | 0.66 ± 0.87 | 0.057 |
|  | Mean difference<br>(Baseline – week 12) | 2.47 ± 1.05 | 1.28 ± 0.99 | *0.001 |
| Upper body pain<br>(0–10) | Baseline | 2.69 ± 0.59<br>[2.47, 2.90] | 2.63 ± 0.61<br>[2.41, 2.84] | 0.649 |
|  | Week 6 | 1.97 ± 0.40<br>[1.82, 2.11] | 2.22 ± 0.61<br>[2.00, 2.44] | — |
|  | Week 12 | 1.41 ± 0.50<br>[1.23, 1.59] | 1.88 ± 0.61<br>[1.66, 2.09] | — |
|  | Mean difference<br>(Baseline – week 6) | 0.72 ± 0.58 | 0.41 ± 0.61 | *0.036 |
|  | Mean difference<br>(Baseline – week 12) | 1.28 ± 0.73 | 0.75 ± 0.57 | *0.003 |
| Lower body soreness<br>(0–10) | Baseline | 5.81 ± 0.86<br>[5.50, 6.12] | 5.63 ± 0.71<br>[5.37, 5.88] | 0.395 |
|  | Week 6 | 4.69 ± 0.82<br>[4.39, 4.98] | 5.06 ± 0.88<br>[4.75, 5.38] | — |
|  | Week 12 | 3.44 ± 0.88<br>[3.12, 3.75] | 4.81 ± 0.74<br>[4.55, 5.08] | — |
|  | Mean difference<br>(Baseline – week 6) | 1.13 ± 0.91 | 0.56 ± 0.84 | *0.010 |
|  | Mean difference<br>(Baseline – week 12) | 2.38 ± 1.13 | 0.81 ± 0.82 | *0.001 |
| Lower body pain<br>(0–10) | Baseline | 2.88 ± 0.66<br>[2.64, 3.11] | 2.59 ± 0.61<br>[2.37, 2.82] | 0.085 |
|  | Week 6 | 2.13 ± 0.49<br>[1.95, 2.30] | 2.19 ± 0.59<br>[1.97, 2.40] | — |
|  | Week 12 | 1.34 ± 0.48<br>[1.17, 1.52] | 1.97 ± 0.54<br>[1.77, 2.16] | — |
|  | Mean difference<br>(Baseline – week 6) | 0.75 ± 0.51 | 0.41 ± 0.50 | *0.010 |
|  | Mean difference<br>(Baseline – week 12) | 1.53 ± 0.72 | 0.63 ± 0.61 | *0.001 |

<sup>1</sup> $p < 0.05$ is significant. These data were analysed with the non-parametric Mann–Whitney $U$ tests. $p$ -values for week 6 and week 12 reflect comparisons of change from baseline to the respective visit. 95% CI values are provided in square brackets.

#### 3.3.6 Physical measures – resting vital signs

**Table 10** summarises this data. Vital signs remained generally stable and within physiological ranges in both treatment groups throughout the study period. No clinically meaningful changes were observed in blood pressure, heart rate, oxygen saturation, oral temperature, or respiratory rate from baseline to week 12. These findings did not identify a clinically apparent adverse effect of the test product on resting vital signs.

**Table 10.**
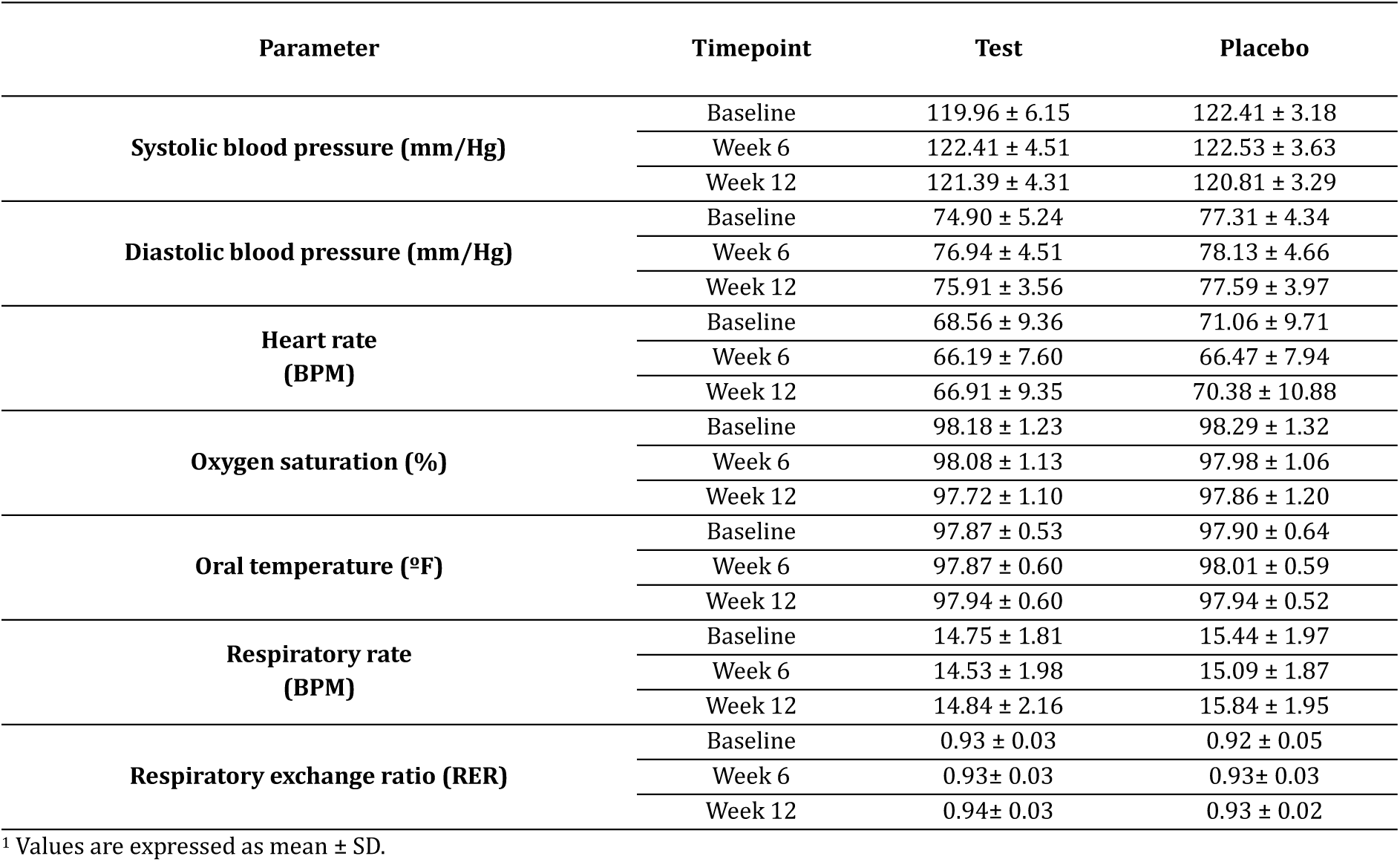
Assessment of resting vital signs.

| Parameter | Timepoint | Test | Placebo |
| --- | --- | --- | --- |
| Systolic blood pressure (mm/Hg) | Baseline | 119.96 ± 6.15 | 122.41 ± 3.18 |
|  | Week 6 | 122.41 ± 4.51 | 122.53 ± 3.63 |
|  | Week 12 | 121.39 ± 4.31 | 120.81 ± 3.29 |
| Diastolic blood pressure (mm/Hg) | Baseline | 74.90 ± 5.24 | 77.31 ± 4.34 |
|  | Week 6 | 76.94 ± 4.51 | 78.13 ± 4.66 |
|  | Week 12 | 75.91 ± 3.56 | 77.59 ± 3.97 |
| Heart rate (BPM) | Baseline | 68.56 ± 9.36 | 71.06 ± 9.71 |
|  | Week 6 | 66.19 ± 7.60 | 66.47 ± 7.94 |
|  | Week 12 | 66.91 ± 9.35 | 70.38 ± 10.88 |
| Oxygen saturation (%) | Baseline | 98.18 ± 1.23 | 98.29 ± 1.32 |
|  | Week 6 | 98.08 ± 1.13 | 97.98 ± 1.06 |
|  | Week 12 | 97.72 ± 1.10 | 97.86 ± 1.20 |
| Oral temperature (°F) | Baseline | 97.87 ± 0.53 | 97.90 ± 0.64 |
|  | Week 6 | 97.87 ± 0.60 | 98.01 ± 0.59 |
|  | Week 12 | 97.94 ± 0.60 | 97.94 ± 0.52 |
| Respiratory rate (BPM) | Baseline | 14.75 ± 1.81 | 15.44 ± 1.97 |
|  | Week 6 | 14.53 ± 1.98 | 15.09 ± 1.87 |
|  | Week 12 | 14.84 ± 2.16 | 15.84 ± 1.95 |
| Respiratory exchange ratio (RER) | Baseline | 0.93 ± 0.03 | 0.92 ± 0.05 |
|  | Week 6 | 0.93 ± 0.03 | 0.93 ± 0.03 |
|  | Week 12 | 0.94 ± 0.03 | 0.93 ± 0.02 |

<sup>1</sup> Values are expressed as mean ± SD.

### 3.4 Routine investigations

Haematological and biochemical investigations were performed at screening and study conclusion for all participants. Although some parameters exhibited statistically significant changes, all values remained within clinically acceptable ranges. Urinalysis parameters remained comparable between the test and placebo groups throughout the study, with no clinically meaningful changes observed from baseline to week 12. Overall, the findings were within normal limits and did not indicate any treatment-related urinary safety concerns (**Table S2**).

### 3.5 Adverse events

A total of three AEs were reported during the study. In the test group, one participant reported nausea, while in the placebo group, two participants reported AEs, comprising one case each of fever and diarrhoea (**Table S3**).

### 3.6 Tolerability and compliance

In this study, all participants demonstrated excellent tolerability (grade 3) to the IPs (**Table S3**). The compliance rates for IP consumption were consistently high in both test and placebo groups throughout the study duration (**Table S4**). In the test group, one participant was non-compliant with the IP intake requirements. Three and four participants in the test and placebo groups, respectively, were non-compliant with the protocol-defined exercise regimen.

### 3.7 Participant withdrawal

A total of eight participants were discontinued from the study. In the test group, four participants discontinued the study, of whom three participants were withdrawn due to non-compliance with the protocol-defined exercise regimen and one participant was withdrawn due to non-adherence to IP intake. In the placebo group, four participants were withdrawn from the study due to non-compliance with the protocol-defined exercise regimen. These participants were not replaced or followed up on, since they were not withdrawn due to AEs.

## 4 Discussion

This randomised, double-blind, placebo-controlled trial evaluated 12 weeks of SpinoleRG™ extract supplementation in recreationally active adults. Compared with the placebo, the test group was associated with greater improvements in CPET, muscular strength, muscular endurance, TTE, selected biochemical markers, and DOMS. The increase in plasma nitrite is consistent with engagement of the nitrate– nitrite–nitric oxide (NO) pathway, with possible downstream effects on vascular function, skeletal muscle contractility, metabolic efficiency, and oxidative stress attenuation (**Figure 7**).

**Figure 7.**
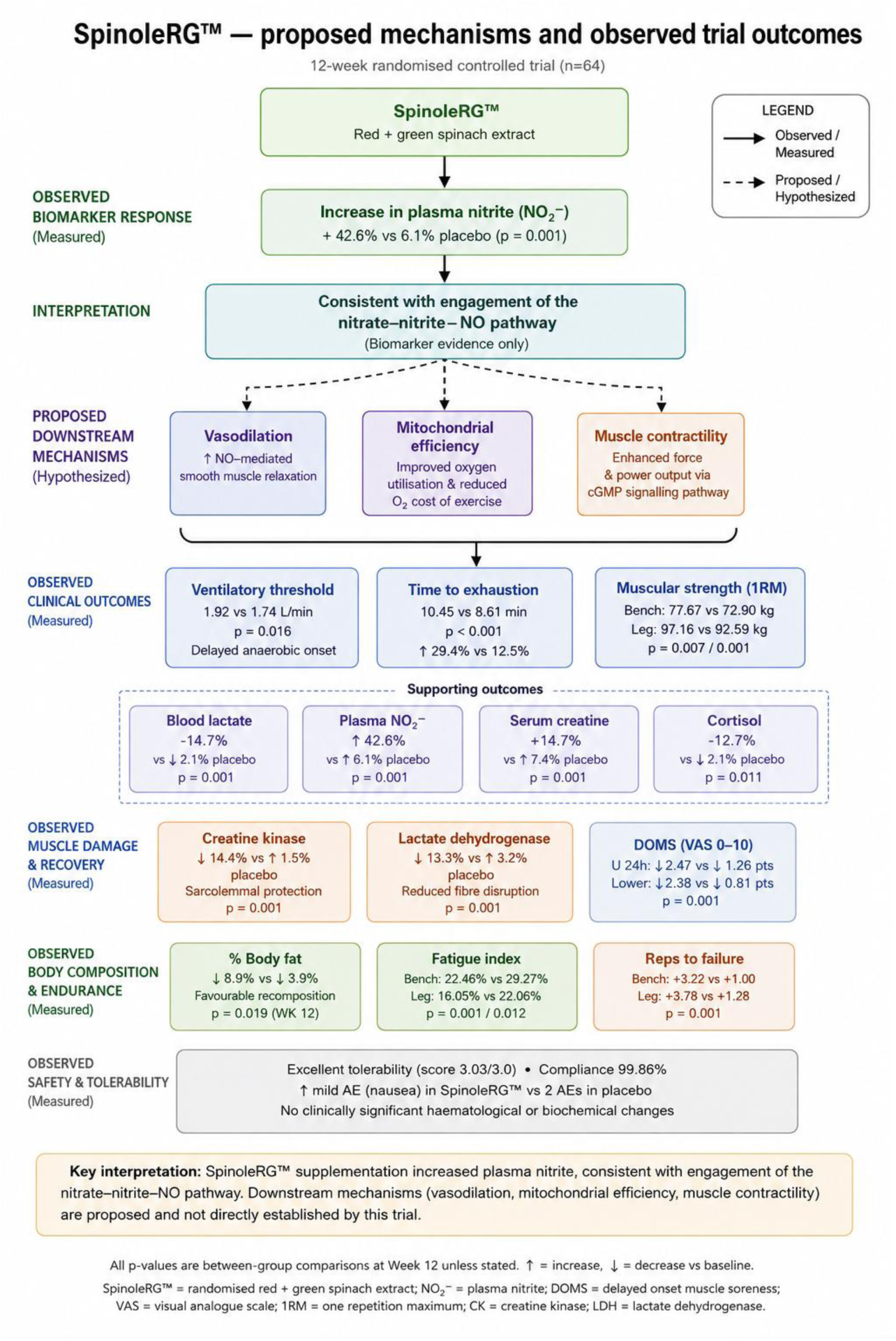
Summary of observed outcomes, interpretations and proposed mechanisms.

The mechanistic explanation for the observed ergogenic effects of the extract is consistent with the enterosalivary conversion of inorganic nitrate (NO₃⁻) to nitrite (NO₂⁻) and subsequently to NO. NO plays a central role in cardiovascular regulation, vasodilation, and glycaemic control [11]. It is also a ubiquitous signalling molecule with specific implications in skeletal muscle, modulating maximal shortening velocity and peak power output of muscle fibers, mitochondrial adaptation, and redox state [12]. In the present trial, the 42.6% increase in plasma nitrite in the test group, compared to a 6.1% in the placebo at 12 weeks, supports increased systemic plasma nitrite availability; however, it cannot be considered conclusive proof of either enhanced intramuscular NO production or mediation of the observed performance outcomes by NO. Nevertheless, the comprehensive improvements observed in this trial spanning strength, endurance, metabolic efficiency, and muscle recovery are consistent with NO’s multifaceted mechanisms of action.

VT was the only CPET parameter with a significant between-group difference in change from baseline at week 12. Conversely, an absence of significant effects in VO₂ max or maximal ventilation was observed. These findings accord with prior research, where using 1,000 mg of *Amaranthus* extract reported improvements in plasma nitrate and VT without significant differences in VO₂ peak or TTE under graded exercise testing, suggesting that VT is a more sensitive marker of nitrate-mediated benefit at submaximal exercise intensities than peak aerobic capacity over shorter study durations [11,13]. Further, nitrate loading from *Amaranthus* may delay the onset of anaerobic ventilatory compensation. As exercise intensity rises, lactic acid-derived hydrogen ions overwhelm bicarbonate buffering, triggering metabolic acidosis that stimulates chemoreceptors to drive the non-linear ventilatory increase defining VT. Enhanced NO bioavailability delays the transition by improving muscle oxygen delivery, reducing anaerobic glycolytic reliance, and shifting VT to higher oxygen uptake values [14]. An umbrella review of 20 systematic reviews encompassing 180 primary studies and 2,672 participants reported a beneficial effect of dietary nitrate with a standardised mean difference (SMD) of 0.33; 95% CI: 0.19–0.47 [15], which establishes a broader ergogenic profile for the molecule. This finding is directionally consistent with our trial’s observed within-group improvement of 29.4%; however, it is limited to this qualitative comparison and our limited cohort of recreationally active adults. It also does not account for placebo effects.

Maximum isometric force development, unloaded shortening velocity, and peak power are beneficially modulated by NO via the cGMP signalling pathway in skeletal muscle fibres [16]. NO also promotes vasodilation for improved blood flow and skeletal muscle performance, hypertrophy, and strength adaptations through improved nutrient and oxygen delivery to contracting muscle during resistance exercise [17]. The significant improvements in 1RM bench press and leg press, and increases in repetitions to failure in the test group, are consistent with this established NO activity. Nonetheless, detailed mechanistic investigations are required for definitive correlation of our findings with NO activity.

Similarly, the improvements in bench and leg press FI at week 12, indicate enhanced muscular endurance and fatigue resistance. Haynes et al. reported that daily supplementation of 2,000 mg *Amaranthus* extract in ten resistance-trained males did not significantly improve bench press performance, muscle oxygenation or cognitive function over seven days [18]. Our trial’s longer duration, relatively heterogeneous cohort and smaller IP dose may suggest that our observed bench and leg press improvements are generally time-based, sex-independent and dose-independent. However, we did not test for muscle oxygenation or cognitive function, and the contribution of *S. oleracea* towards muscular mass and function must also be considered.

Blood lactate decreased by 41.7% in the test group compared to 2.1% in the placebo group at week 12, which may reflect reduced lactate production or increased clearance due to exercise demand. The reduced lactate accumulation indicates decreased anaerobic glycolytic flux, which is consistent with the observed VT shift. In consonance with NO’s effects on VT, it likely indirectly affects blood lactate levels by promoting mitochondrial biogenesis and efficiency while maintaining ATP production [12]. Interestingly, prior studies on NO-enhancing supplements found that repetitions to failure significantly improved while blood lactate levels did not change significantly [19,20]. However, these trials did not measure changes in VT, and their shorter durations may not have allowed for the associated metabolic adaptations with NO.

The significantly greater reductions in serum CK and LDH at week 12 suggest reduced exercise-induced muscle stress, since CK and LDH elevations conventionally indicate muscle fibre damage and membrane integrity disruption, usually from reactive oxygen species (ROS) modifying sarcolemmal proteins. The antioxidant polyphenols in SpinoleRG™ and NO-related changes in perfusion are plausible contributors [11], however, oxidative stress markers were not measured in this study.

Serum creatine increased 41.7% in the test group compared to 7.2% in the placebo at week 12. Elevated serum creatine may result in an increase in the available amount of phosphocreatine substrate, the primary system for rapid ATP resynthesis during high-intensity efforts, in skeletal muscle. NO-mediated vasodilation likely facilitates greater substrate delivery and creatine shuttling to contracting muscle, while improving phosphocreatine regeneration between bouts [21]. This could explain the concurrent improvements in both 1RM strength and muscular endurance, as the phosphocreatine system contributes to energy supply across both performance domains.

The significantly greater cortisol reduction in the test group at week 12 suggests attenuation of exercise-induced physiological stress. Chronic exercise training is inherently a catabolic stimulus that activates the hypothalamic–pituitary–adrenal (HPA) axis, elevating cortisol as part of the adaptive stress response. Sustained elevation of cortisol, however, is associated with muscle protein catabolism, impaired recovery, and reduced training adaptation [22]. Overall reduced metabolic stress per training session, as can be inferred from the reduced CK, LDH, and blood lactate levels, translates to attenuated HPA axis activation. This finding is significant in that lower post-exercise cortisol in the test group supports an anabolic-to-catabolic hormonal environment more conducive to muscle protein synthesis and training adaptation.

Nutrition-based interventional approaches against DOMS targeting post-exercise inflammation and ROS have received considerable attention, given the evidence that both inflammatory responses and free radicals produced during exercise contribute to the soreness experience [23,24]. The polyphenolic and nitrate-containing components of SpinoleRG™ may address these mechanisms through antioxidant and anti-inflammatory actions. As we did not assess inflammatory and oxidative stress biomarkers, however, we interpret our findings as supporting a reduction in perceived soreness and pain, that could have contributed to the sustained performance improvements observed over the 12-week intervention period.

A progressive and significant reduction in body fat percentage and a non-significant increase in skeletal muscle percentage presents a body composition profile consistent with the discussed NO-mediated metabolic enhancement. A 12-week trial of spinach extract in adults over 50 years associated supplementation with fat reduction and increase in lean and muscle mass, which is directionally in line with the findings in our trial. Effects on body composition should be interpreted cautiously, however, due to the differing age and fitness of the cohort and our reliance on the participants to maintain consistency in their diet and training [25].

Three mild AEs considered unrelated to the interventions and resolved, besides normal vital signs and routine parameters, support the tolerability of the extract over 12 weeks, and are consistent with safety evaluations in the existing literature. For instance, Wen et al. evaluated spinach extract supplementation in 94 participants across four trials that reported significant improvements in most physical performance parameters with no complications [26].

This study has several limitations. The restriction of the sample population to physically active adults with a mean age of around 31 years limits generalisability to elite athletic populations, older or sedentary adults, and clinical populations. Our neglect towards multiplicity adjustment skews the probabilities of false positive results. Diet and training were monitored by participant diaries rather than minutely managed. BIA, despite not being the gold standard, was the only test used for analysing body composition. Finally, the absence of direct measures of intramuscular NO production, muscle biopsy data, or continuous lactate profiling constrain mechanistic inference beyond theory. Future studies should incorporate a longer follow-up period, include redundant testing, perform multiplicity control, and focus on the mechanistic interactions between the constituent bioactives and activated pathways behind the beneficial activities of SpinoleRG™.

## Supporting information

CONSORT checklist

Raw statistical outputs

## Data Availability

All data produced and reported in the present work are contained in the manuscript and its supplementary material.

## Abbreviations

1RM: one-repetition maximum
AE: adverse event
ALT: alanine aminotransferase
ANCOVA: analysis of covariance
AST: aspartate aminotransferase
BIA: bioelectrical impedance analysis
BMI: body mass index
BPM: beats per minute
CBC: complete blood count
CI: confidence interval
CK: creatine kinase
CONSORT: Consolidated Standards of Reporting Trials
CPET: cardiopulmonary exercise test
CRF: case report form
CRO: contract research organisation
CTRI: Clinical Trials Registry – India
DOMS: delayed-onset muscle soreness
FI: fatigue index
HPA: hypothalamic-pituitary-adrenal axis
HPLC: high-performance liquid chromatography
ICF: informed consent form
IEC: institutional ethics committee
IP: investigational product
LDH: lactate dehydrogenase
LFT: liver function test
LMM: linear mixed-effects model
NLT: not less than
NO: nitric oxide
NOAEL: no-observed-adverse-effect level
PDA: photodiode array detection
PP: per-protocol
RER: respiratory exchange ratio
RFT: renal function test
ROS: reactive oxygen species
SD: standard deviation
TMMG: 3′,4′,5-trihydroxy-3-methoxy-6,7-methylenedioxyflavone 4′-glucuronide TTE time to exhaustion
VT: ventilatory threshold

## Supplementary material

**Table S1:** Assessment of demographic characteristics.

**Table S2:** Assessment of haematological and biochemical parameters.

**Table S3:** Summary of adverse events.

**Table S4:** Assessment of tolerability and compliance.

## Ethical statement

The study was conducted in accordance with the Declaration of Helsinki and approved by the joint institutional ethical committee of Sangvi Multispeciality Hospital, and Gunjkar Multispeciality Hospital, Pune, Maharashtra, India (**MHC/CT/25-26/016, 25 August 2025**).

## Informed consent statement

Informed consent was obtained from all subjects involved in the study.

## Acknowledgements

The authors sincerely thank the participants for their time and compliance afforded to the study.

## Author contributions

**Conceptualisation,** SJM, VP, GGa, AM, AD; **Methodology,** SP, DZ, VP, GGa; **Software,** SP, DZ, VP, GGa; **Validation,** VP, GGa, PM; **Formal Analysis,** VP, GGa; **Investigation,** SP, DZ; **Resources,** SP, DZ, AM, AV, DM; **Data Curation,** VP, GGa, AB, GGu, PM; **Writing – Original Draft Preparation,** VP, GGa, AB; **Writing – Review & Editing,** SJM, JS, VP, GGa, AB; **Visualization,** AB, GGu; **Supervision,** SJM, AM, AV, DM, JS; **Project Administration,** SJM, VP, GGa, AM, AV, DM, AD, JS; **Funding Acquisition,** AM, AV, DM.

