## Supplementary tables for "Effects of a standardised *Amaranthus tricolor*–*Spinacia oleracea* extract on exercise performance, muscle strength, endurance and recovery in physically active adults: a randomised, double-blind, placebo-controlled trial"

**Table S1: Miscellaneous anthropometric and body composition parameters**

| **Parameter** | **Timepoint** | **Test**  **[95%CI]** | **Placebo**  **[95%CI]** | ***p*-value (Between Groups)** |
| --- | --- | --- | --- | --- |
| **Body Weight (kg)** | Baseline | 74.65 ± 6.02  [72.48,76.82] | 75.08 ± 5.57  [73.07,77.08] | 0.722 |
|  | Week 6 | 75.42 ± 5.84  [73.34,77.56] | 75.10 ± 5.28  [73.20,77.00] | — |
|  | Week 12 | 75.80 ± 5.73  [73.74,77.87] | 75.40 ± 5.20  [73.52,77.27] | — |
|  | Mean diff  (Baseline –Week 6) | 0.80 ± 1.69 | 0.02 ± 1.07 | *0.026 |
|  | Mean diff  (Baseline – Week 12) | 1.15 ± 1.96 | 0.32 ±1.70 | 0.092 |
| **BMI**  **(kg/m2)** | Baseline | 25.88 ± 0.82  [25.58,26.18] | 26.23 ± 1.03  [25.86,26.60] | 0.254 |
|  | Week 6 | 26.16 ± 0.79  [25.88,26.45] | 26.25 ± 1.09  [25.86,26.64] | — |
|  | Week 12 | 26.29 ± 0.89  [25.97,26.61] | 26.36 ± 1.18  [25.93,26.79] | — |
|  | Mean diff  (Baseline –Week 6) | 0.28 ± 0.59 | 0.02 ± 0.38 | *0.025 |
|  | Mean diff  (Baseline – Week 12) | 0.41 ± 0.71 | 0.13 ± 0.59 | 0.099 |
| **Hip Circumference (cm)** | Baseline | 93.53 ± 6.46  [91.20,95.86] | 91.63 ± 4.43  [90.03,93.22] | 0.175 |
|  | Week 6 | 92.38 ± 6.11  [90.17,94.58] | 90.79 ± 4.27  [89.25,92.33] | — |
|  | Week 12 | 91.88 ± 5.74  [89.81,93.94] | 90.54 ± 4.30  [88.99,92.09] | — |
|  | Mean diff  (Baseline –Week 6) | 1.16 ± 1.14 | 0.84 ± 0.51 | 0.375 |
|  | Mean diff  (Baseline – Week 12) | 1.66 ± 1.36 | 1.09 ± 0.74 | 0.064 |
| **Waist Circumference (cm)** | Baseline | 85.31 ± 6.59  [82.94,87.69] | 87.78 ± 5.15  [85.92,89.64] | 0.080 |
|  | Week 6 | 83.75 ± 6.48  [81.42,86.08] | 86.84 ± 5.11  [85.00,88.68] | — |
|  | Week 12 | 83.38 ± 6.35  [81.08,85.67] | 86.69 ± 4.90  [84.92,88.45] | — |
|  | Mean diff  (Baseline –Week 6) | 1.56 ± 1.29 | 0.94 ± 2.09 | 0.397 |
|  | Mean diff  (Baseline – Week 12) | 1.94 ± 1.44 | 1.09 ± 2.12 | 0.082 |
| **% Body Fat** | Baseline | 18.54 ± 5.85  [16.43,20.65] | 18.25 ± 5.44  [16.29,20.21] | 0.968 |
|  | Week 6 | 17.12 ± 5.31  [15.21,19.04] | 17.85 ± 5.31  [15.94,19.77] | — |
|  | Week 12 | 16.88 ± 4.90  [15.12,18.65] | 17.52 ± 5.03  [15.70,19.33] | — |
|  | Mean diff  (Baseline –Week 6) | 1.42 ± 1.84 | 0.40 ± 1.66 | *0.001 |
|  | Mean diff  (Baseline – Week 12) | 1.66 ± 1.62 | 0.73 ± 1.84 | *0.019 |

**Table S2: Assessment of haematological and biochemical parameters**

| **Timepoint** | **Test** | **Placebo** | **p-value**  **(Between Groups)** | |
| --- | --- | --- | --- | --- |
| **Total Leukocyte Count** | | | | |
| **Baseline** | 8419.34 ± 1524.44 | 7564.97 ± 1605.97 | | 0.065 |
| **Week 12** | 7607.81 ± 1985.71 | 7605.63 ± 1994.58 | | — |
| **Mean diff**  **(Baseline – Week 12)** | 811.53 ± 1729.20 | 40.66 ± 2194.66 | | 0.099 |
| **Neutrophils** | | | | |
| **Baseline** | 68.44 ± 7.78 | 64.59 ± 10.23 | | — |
| **Week 12** | 69.59 ± 6.37 | 66.78 ± 8.17 | | — |
| **Mean diff**  **(Baseline – Week 12)** | 1.16 ± 7.89 | 2.19 ± 12.20 | | 0.258 |
| **Lymphocytes** | | | | |
| **Baseline** | 29.09 ± 6.10 | 31.63 ± 9.50 | | 0.164 |
| **Week 12** | 26.66 ± 6.29 | 28.50 ± 6.13 | | — |
| **Mean diff**  **(Baseline – Week 12)** | 2.44 ± 6.31 | 3.13 ± 8.55 | | 0.995 |
| **Monocytes** | | | | |
| **Baseline** | 2.25 ± 0.44 | 2.41 ± 0.56 | | 0.257 |
| **Week 12** | 2.56 ± 0.67 | 2.84 ± 1.63 | | — |
| **Mean diff**  **(Baseline – Week 12)** | 0.31 ± 0.90 | 0.44 ± 1.79 | | 0.413 |
| **Eosinophils** | | | | |
| **Baseline** | 1.22 ± 0.42 | 1.38 ± 0.55 | | 0.246 |
| **Week 12** | 1.56 ± 0.62 | 1.72 ± 0.68 | | — |
| **Mean diff**  **(Baseline – Week 12)** | 0.34 ± 0.70 | 0.34 ± 0.94 | | 0.953 |
| **Basophils** | | | | |
| **Baseline** | 0.00 ± 0.00 | 0.00 ± 0.00 | | 1.000 |
| **Week 12** | 0.00 ± 0.00 | 0.00 ± 0.00 | | — |
| **Mean diff**  **(Baseline – Week 12)** | 0.00 ± 0.00 | 0.00 ± 0.00 | | 1.000 |
| **Total RBC Count** | | | | |
| **Baseline** | 5.21 ± 0.46 | 5.26 ± 0.76 | | — |
| **Week 12** | 5.30 ± 0.55 | 5.21 ± 0.58 | | — |
| **Mean diff**  **(Baseline – Week 12)** | 0.09 ± 0.61 | 0.05 ± 0.85 | | 0.460 |
| **Hemoglobin** | | | | |
| **Baseline** | 14.46 ± 0.92 | 14.32 ± 0.96 | | 0.638 |
| **Week 12** | 14.36 ± 0.85 | 14.23 ± 0.90 | | — |
| **Mean diff**  **(Baseline – Week 12)** | 0.12 ± 0.74 | 0.09 ± 0.35 | | 0.989 |
| **Hematocrit** | | | | |
| **Baseline** | 42.95 ± 4.56 | 42.83 ± 5.31 | | — |
| **Week 12** | 42.17 ± 3.70 | 41.87 ± 3.84 | | — |
| **Mean diff**  **(Baseline – Week 12)** | 0.78 ± 4.28 | 0.96 ± 6.20 | | 0.764 |
| **Platelets** | | | | |
| **Baseline** | 294.72 ± 81.01 | 279.75 ± 64.03 | | — |
| **Week 12** | 289.38 ± 82.84 | 302.53 ± 74.65 | | — |
| **Mean diff**  **(Baseline – Week 12)** | 5.34 ± 60.40 | 22.78 ± 77.28 | | 0.168 |
| **Bilirubin Total** | | | | |
| **Baseline** | 0.61 ± 0.21 | 0.65 ± 0.22 | | 0.586 |
| **Week 12** | 0.79 ± 0.25 | 0.76 ± 0.27 | | — |
| **Mean diff**  **(Baseline – Week 12)** | 0.18 ± 0.28 | 0.12 ± 0.33 | | 0.375 |
| **Bilirubin Direct** | | | | |
| **Baseline** | 0.21 ± 0.09 | 0.24 ± 0.08 | | — |
| **Week 12** | 0.26 ± 0.09 | 0.24 ± 0.08 | | — |
| **Mean diff**  **(Baseline – Week 12)** | 0.05 ± 0.10 | 0.00 ± 0.10 | | 0.216 |
| **Bilirubin Indirect** | | | | |
| **Baseline** | 0.41 ± 0.22 | 0.41 ± 0.22 | | 0.968 |
| **Week 12** | 0.54 ± 0.21 | 0.52 ± 0.24 | | — |
| **Mean diff**  **(Baseline – Week 12)** | 0.12 ± 0.27 | 0.11 ± 0.31 | | 0.846 |
| **SGOT** | | | | |
| **Baseline** | 28.63 ± 11.25 | 28.75 ± 8.78 | | — |
| **Week 12** | 28.29 ± 9.44 | 26.08 ± 10.41 | | — |
| **Mean diff**  **(Baseline – Week 12)** | 0.34 ± 12.30 | 2.66 ± 13.48 | | 0.370 |
| **SGPT** | | | | |
| **Baseline** | 29.54 ± 12.42 | 31.19 ± 9.30 | | 0.327 |
| **Week 12** | 30.22 ± 14.61 | 24.63 ± 10.14 | | — |
| **Mean diff**  **(Baseline – Week 12)** | 0.68 ± 14.05 | 6.55 ± 13.03 | | *0.035 |
| **Alkaline Phosphatase** | | | | |
| **Baseline** | 107.42 ± 22.29 | 107.27 ± 20.97 | | 0.978 |
| **Week 12** | 105.76 ± 23.95 | 102.02 ± 21.54 | | — |
| **Mean diff**  **(Baseline – Week 12)** | 1.66 ± 30.91 | 5.26 ± 27.91 | | 0.516 |
| **Total Proteins** | | | | |
| **Baseline** | 6.90 ± 0.35 | 6.96 ± 0.40 | | 0.424 |
| **Week 12** | 6.98 ± 0.77 | 6.83 ± 0.45 | | — |
| **Mean diff**  **(Baseline – Week 12)** | 0.09 ± 0.77 | 0.13 ± 0.48 | | 0.099 |
| **Serum Albumin** | | | | |
| **Baseline** | 4.00 ± 0.39 | 4.07 ± 0.47 | | 0.619 |
| **Week 12** | 3.92 ± 0.43 | 3.75 ± 0.29 | | — |
| **Mean diff**  **(Baseline – Week 12)** | 0.04 ± 0.98 | 0.32 ± 0.61 | | 0.161 |
| **Serum Globulin** | | | | |
| **Baseline** | 2.88 ± 0.31 | 2.89 ± 0.42 | | — |
| **Week 12** | 2.96 ± 0.55 | 3.08 ± 0.52 | | — |
| **Mean diff**  **(Baseline – Week 12)** | 0.08 ± 0.62 | 0.19 ± 0.71 | | 0.408 |
| **A/G Ratio** | | | | |
| **Baseline** | 1.42 ± 0.25 | 1.45 ± 0.32 | | 0.652 |
| **Week 12** | 1.37 ± 0.36 | 1.33 ± 0.51 | | — |
| **Mean diff**  **(Baseline – Week 12)** | 0.05 ± 0.11 | 0.12 ± 0.19 | | 0.331 |
| **Serum Uric Acid** | | | | |
| **Baseline** | 4.53 ± 1.28 | 4.81 ± 1.22 | | 0.197 |
| **Week 12** | 4.23 ± 1.04 | 4.48 ± 1.11 | | — |
| **Mean diff**  **(Baseline – Week 12)** | 0.30 ± 1.41 | 0.33 ± 1.29 | | 0.672 |
| **Serum Blood Urea** | | | | |
| **Baseline** | 20.95 ± 4.51 | 19.49 ± 4.22 | | 0.157 |
| **Week 12** | 18.73 ± 4.61 | 17.92 ± 3.80 | | — |
| **Mean diff**  **(Baseline – Week 12)** | 2.22 ± 4.91 | 1.57 ± 4.81 | | 0.489 |
| **Serum creatinine** | | | | |
| **Baseline** | 0.82 ± 0.11 | 0.85 ± 0.17 | | — |
| **Week 12** | 0.81 ± 0.14 | 0.81 ± 0.14 | | — |
| **Mean diff**  **(Baseline – Week 12)** | 0.01 ± 0.12 | 0.04 ± 0.15 | | 0.616 |
| **Total Cholesterol** | | | | |
| **Baseline** | 170.73 ± 32.36 | 169.13 ± 29.86 | | — |
| **Week 12** | 169.14 ± 28.69 | 159.98 ± 37.25 | | — |
| **Mean diff**  **(Baseline – Week 12)** | 1.59 ± 35.13 | 9.15 ± 40.38 | | 0.283 |
| **HDL Cholesterol** | | | | |
| **Baseline** | 54.82 ± 11.48 | 56.31 ± 11.94 | | 0.502 |
| **Week 12** | 51.56 ± 11.97 | 51.19 ± 11.93 | | — |
| **Mean diff**  **(Baseline – Week 12)** | 3.26 ± 14.70 | 5.12 ± 13.47 | | 0.702 |
| **Triglycerides** | | | | |
| **Baseline** | 129.63 ± 33.82 | 130.90 ± 34.97 | | — |
| **Week 12** | 125.68 ± 33.48 | 127.24 ± 31.34 | | — |
| **Mean diff**  **(Baseline – Week 12)** | 3.95 ± 38.46 | 3.66 ± 46.09 | | 0.869 |
| **LDL Cholesterol** | | | | |
| **Baseline** | 89.98 ± 33.43 | 86.64 ± 26.47 | | — |
| **Week 12** | 92.44 ± 25.78 | 83.34 ± 34.30 | | — |
| **Mean diff**  **(Baseline – Week 12)** | 2.46 ± 38.33 | 3.30 ± 38.75 | | 0.265 |
| **VLDL Cholesterol** | | | | |
| **Baseline** | 25.93 ± 6.76 | 26.18 ± 6.99 | | — |
| **Week 12** | 25.14 ± 6.70 | 25.45 ± 6.27 | | — |
| **Mean diff**  **(Baseline – Week 12)** | 0.79 ± 7.69 | 0.73 ± 9.22 | | 0.869 |
| **TC/HDL Ratio** | | | | |
| **Baseline** | 3.23 ± 0.83 | 3.10 ± 0.68 | | — |
| **Week 12** | 3.38 ± 0.68 | 3.22 ± 0.80 | | — |
| **Mean diff**  **(Baseline – Week 12)** | 0.15 ± 1.10 | 0.12 ± 0.94 | | 0.430 |
| **LDL/HDL Ratio** | | | | |
| **Baseline** | 1.73 ± 0.75 | 1.60 ± 0.56 | | — |
| **Week 12** | 1.85 ± 0.60 | 1.69 ± 0.71 | | — |
| **Mean diff**  **(Baseline – Week 12)** | 0.12 ± 0.99 | 0.09 ± 0.83 | | 0.359 |
| **Serum sodium** | | | | |
| **Baseline** | 138.11 ± 1.99 | 138.56 ± 2.29 | | — |
| **Week 12** | 138.87 ± 2.32 | 139.70 ± 2.30 | | — |
| **Mean diff**  **(Baseline – Week 12)** | 0.76 ± 2.82 | 1.14 ± 2.57 | | 0.219 |
| **Serum Potassium** | | | | |
| **Baseline** | 4.41 ± 0.78 | 4.45 ± 0.60 | | 0.493 |
| **Week 12** | 4.33 ± 0.60 | 4.32 ± 0.55 | | — |
| **Mean diff**  **(Baseline – Week 12)** | 0.07 ± 0.87 | 0.14 ± 0.82 | | 0.448 |
| **Serum Chloride** | | | | |
| **Baseline** | 98.77 ± 1.57 | 100.11 ± 1.82 | | — |
| **Week 12** | 99.53 ± 1.90 | 99.28 ± 2.07 | | — |
| **Mean diff**  **(Baseline – Week 12)** | 0.76 ± 0.87 | 0.83 ± 2.26 | | 0.128 |
| **Bicarbonate** | | | | |
| **Baseline** | 21.77 ± 2.12 | 22.02 ± 2.27 | | 0.712 |
| **Week 12** | 27.56 ± 2.64 | 26.37 ± 2.12 | | — |
| **Mean diff**  **(Baseline – Week 12)** | 5.79 ± 2.08 | 4.34 ± 1.68 | | *0.001 |
| **Inorganic Phosphorus** | | | | |
| **Baseline** | 4.27 ± 4.19 | 3.50 ± 0.51 | | 0.742 |
| **Week 12** | 3.75 ± 0.57 | 3.65 ± 0.91 | | — |
| **Mean diff**  **(Baseline – Week 12)** | 0.52 ± 4.31 | 0.15 ± 0.90 | | 0.596 |

** p < 0.05 significant. Data found to be not normal hence non-parametric ANCOVA and Mann–Whitney U tests applied.*

**Table S3: Summary of adverse events**

| **Adverse Events** | **Test** | | **Placebo** | |
| --- | --- | --- | --- | --- |
|  | **Number of participants** | **%** | **Number of participants** | **%** |
| Nausea | 01 | 3.12 | - | - |
| Fever | - | - | 01 | 3.12 |
| Diarrhea | - | - | 01 | 3.12 |
| **Total No. of Events** | **01** | **3.12** | **02** | **6.25** |
| **Total No. of Participants** | **01** | **3.12** | **02** | **6.25** |

**Table S4: Tolerability and compliance**

| **Visit** | **Test** | **Placebo** |
| --- | --- | --- |
| **Tolerability** | | |
| **Week 6** | 3.00 ± 0.00 | 3.00 ± 0.00 |
| **Week 12** | 3.00 ± 0.00 | 3.00 ± 0.00 |
| **Compliance** | | |
| **Week 6** | 99.86 ± 0.55 | 99.86 ± 0.55 |
| **Week 12** | 99.86 ± 0.55 | 99.93 ± 0.39 |

*Values are expressed as mean ± SD*
