## Supplementary material for "Effects of a standardised *Amaranthus tricolor*–*Spinacia oleracea* extract on exercise performance, muscle strength, endurance and recovery in physically active adults: a randomised, double-blind, placebo-controlled trial": Raw statistical outputs: Antho_Output_17.07.2026.doc

Explore


Notes	
Output Created	17-JUL-2026 11:47:10	
Comments		
Input	Active Dataset	DataSet0	
	Filter	<none>	
	Weight	<none>	
	Split File	<none>	
	N of Rows in Working Data File	64	
Missing Value Handling	Definition of Missing	User-defined missing values for dependent variables are treated as missing.	
	Cases Used	Statistics are based on cases with no missing values for any dependent variable or factor used.	
Syntax	EXAMINE VARIABLES=Body_Weight_B Body_Weight_W6 Body_Weight_W12 Body_Weight_Diff_W6
    Body_Weight_Diff_W12 Height_B Height_W6 Height_W12 Height_Diff_W6 Height_Diff_W12 BMI_B BMI_W6
    BMI_W12 BMI_Diff_W6 BMI_Diff_W12 Hip_Circumf_B Hip_Circumf_W6 Hip_Circumf_W12 Hip_Circumf_Diff_W6
    Hip_Circumf_Diff_W12 Waist_Circumf_B Waist_Circumf_W6 Waist_Circumf_W12 Waist_Circumf_Diff_W6
    Waist_Circumf_Diff_W12 Chest_Circumf_B Chest_Circumf_W6 Chest_Circumf_W12 Chest_Circumf_Diff_W6
    Chest_Circumf_Diff_W12 Body_Fat_B Body_Fat_W6 Body_Fat_W12 Body_Fat_Diff_W6 Body_Fat_Diff_W12
    Skeletal_Muscle_B Skeletal_Muscle_W6 Skeletal_Muscle_W12 Skeletal_Muscle_Diff_W6
    Skeletal_Muscle_Diff_W12 BY Group
  /PLOT BOXPLOT STEMLEAF
  /COMPARE GROUPS
  /STATISTICS DESCRIPTIVES
  /CINTERVAL 95
  /MISSING LISTWISE
  /NOTOTAL.	
Resources	Processor Time	00:00:03.25	
	Elapsed Time	00:00:03.16	


Group


Case Processing Summary	
	Group	Cases	
		Valid	Missing	Total	
		N	Percent	N	Percent	N	Percent	
Body_Weight_B	1.00	32	100.0%	0	0.0%	32	100.0%	
	2.00	32	100.0%	0	0.0%	32	100.0%	
Body_Weight_W6	1.00	32	100.0%	0	0.0%	32	100.0%	
	2.00	32	100.0%	0	0.0%	32	100.0%	
Body_Weight_W12	1.00	32	100.0%	0	0.0%	32	100.0%	
	2.00	32	100.0%	0	0.0%	32	100.0%	
Body_Weight_Diff_W6	1.00	32	100.0%	0	0.0%	32	100.0%	
	2.00	32	100.0%	0	0.0%	32	100.0%	
Body_Weight_Diff_W12	1.00	32	100.0%	0	0.0%	32	100.0%	
	2.00	32	100.0%	0	0.0%	32	100.0%	
Height_B	1.00	32	100.0%	0	0.0%	32	100.0%	
	2.00	32	100.0%	0	0.0%	32	100.0%	
Height_W6	1.00	32	100.0%	0	0.0%	32	100.0%	
	2.00	32	100.0%	0	0.0%	32	100.0%	
Height_W12	1.00	32	100.0%	0	0.0%	32	100.0%	
	2.00	32	100.0%	0	0.0%	32	100.0%	
Height_Diff_W6	1.00	32	100.0%	0	0.0%	32	100.0%	
	2.00	32	100.0%	0	0.0%	32	100.0%	
Height_Diff_W12	1.00	32	100.0%	0	0.0%	32	100.0%	
	2.00	32	100.0%	0	0.0%	32	100.0%	
BMI_B	1.00	32	100.0%	0	0.0%	32	100.0%	
	2.00	32	100.0%	0	0.0%	32	100.0%	
BMI_W6	1.00	32	100.0%	0	0.0%	32	100.0%	
	2.00	32	100.0%	0	0.0%	32	100.0%	
BMI_W12	1.00	32	100.0%	0	0.0%	32	100.0%	
	2.00	32	100.0%	0	0.0%	32	100.0%	
BMI_Diff_W6	1.00	32	100.0%	0	0.0%	32	100.0%	
	2.00	32	100.0%	0	0.0%	32	100.0%	
BMI_Diff_W12	1.00	32	100.0%	0	0.0%	32	100.0%	
	2.00	32	100.0%	0	0.0%	32	100.0%	
Hip_Circumf_B	1.00	32	100.0%	0	0.0%	32	100.0%	
	2.00	32	100.0%	0	0.0%	32	100.0%	
Hip_Circumf_W6	1.00	32	100.0%	0	0.0%	32	100.0%	
	2.00	32	100.0%	0	0.0%	32	100.0%	
Hip_Circumf_W12	1.00	32	100.0%	0	0.0%	32	100.0%	
	2.00	32	100.0%	0	0.0%	32	100.0%	
Hip_Circumf_Diff_W6	1.00	32	100.0%	0	0.0%	32	100.0%	
	2.00	32	100.0%	0	0.0%	32	100.0%	
Hip_Circumf_Diff_W12	1.00	32	100.0%	0	0.0%	32	100.0%	
	2.00	32	100.0%	0	0.0%	32	100.0%	
Waist_Circumf_B	1.00	32	100.0%	0	0.0%	32	100.0%	
	2.00	32	100.0%	0	0.0%	32	100.0%	
Waist_Circumf_W6	1.00	32	100.0%	0	0.0%	32	100.0%	
	2.00	32	100.0%	0	0.0%	32	100.0%	
Waist_Circumf_W12	1.00	32	100.0%	0	0.0%	32	100.0%	
	2.00	32	100.0%	0	0.0%	32	100.0%	
Waist_Circumf_Diff_W6	1.00	32	100.0%	0	0.0%	32	100.0%	
	2.00	32	100.0%	0	0.0%	32	100.0%	
Waist_Circumf_Diff_W12	1.00	32	100.0%	0	0.0%	32	100.0%	
	2.00	32	100.0%	0	0.0%	32	100.0%	
Chest_Circumf_B	1.00	32	100.0%	0	0.0%	32	100.0%	
	2.00	32	100.0%	0	0.0%	32	100.0%	
Chest_Circumf_W6	1.00	32	100.0%	0	0.0%	32	100.0%	
	2.00	32	100.0%	0	0.0%	32	100.0%	
Chest_Circumf_W12	1.00	32	100.0%	0	0.0%	32	100.0%	
	2.00	32	100.0%	0	0.0%	32	100.0%	
Chest_Circumf_Diff_W6	1.00	32	100.0%	0	0.0%	32	100.0%	
	2.00	32	100.0%	0	0.0%	32	100.0%	
Chest_Circumf_Diff_W12	1.00	32	100.0%	0	0.0%	32	100.0%	
	2.00	32	100.0%	0	0.0%	32	100.0%	
Body_Fat_B	1.00	32	100.0%	0	0.0%	32	100.0%	
	2.00	32	100.0%	0	0.0%	32	100.0%	
Body_Fat_W6	1.00	32	100.0%	0	0.0%	32	100.0%	
	2.00	32	100.0%	0	0.0%	32	100.0%	
Body_Fat_W12	1.00	32	100.0%	0	0.0%	32	100.0%	
	2.00	32	100.0%	0	0.0%	32	100.0%	
Body_Fat_Diff_W6	1.00	32	100.0%	0	0.0%	32	100.0%	
	2.00	32	100.0%	0	0.0%	32	100.0%	
Body_Fat_Diff_W12	1.00	32	100.0%	0	0.0%	32	100.0%	
	2.00	32	100.0%	0	0.0%	32	100.0%	
Skeletal_Muscle_B	1.00	32	100.0%	0	0.0%	32	100.0%	
	2.00	32	100.0%	0	0.0%	32	100.0%	
Skeletal_Muscle_W6	1.00	32	100.0%	0	0.0%	32	100.0%	
	2.00	32	100.0%	0	0.0%	32	100.0%	
Skeletal_Muscle_W12	1.00	32	100.0%	0	0.0%	32	100.0%	
	2.00	32	100.0%	0	0.0%	32	100.0%	
Skeletal_Muscle_Diff_W6	1.00	32	100.0%	0	0.0%	32	100.0%	
	2.00	32	100.0%	0	0.0%	32	100.0%	
Skeletal_Muscle_Diff_W12	1.00	32	100.0%	0	0.0%	32	100.0%	
	2.00	32	100.0%	0	0.0%	32	100.0%	


Descriptives	
	Group	Statistic	Std. Error	
Body_Weight_B	1.00	Mean	74.6531	1.06389	
		95% Confidence Interval for Mean	Lower Bound	72.4833		
			Upper Bound	76.8229		
		5% Trimmed Mean	74.2958		
		Median	72.6000		
		Variance	36.219		
		Std. Deviation	6.01825		
		Minimum	65.20		
		Maximum	90.20		
		Range	25.00		
		Interquartile Range	8.28		
		Skewness	.908	.414	
		Kurtosis	.561	.809	
	2.00	Mean	75.0781	.98395	
		95% Confidence Interval for Mean	Lower Bound	73.0713		
			Upper Bound	77.0849		
		5% Trimmed Mean	74.9569		
		Median	75.6500		
		Variance	30.981		
		Std. Deviation	5.56607		
		Minimum	63.60		
		Maximum	89.00		
		Range	25.40		
		Interquartile Range	7.92		
		Skewness	.233	.414	
		Kurtosis	.226	.809	
Body_Weight_W6	1.00	Mean	75.4500	1.03311	
		95% Confidence Interval for Mean	Lower Bound	73.3430		
			Upper Bound	77.5570		
		5% Trimmed Mean	75.1035		
		Median	74.4500		
		Variance	34.154		
		Std. Deviation	5.84416		
		Minimum	66.10		
		Maximum	90.70		
		Range	24.60		
		Interquartile Range	8.25		
		Skewness	.895	.414	
		Kurtosis	.736	.809	
	2.00	Mean	75.1000	.93326	
		95% Confidence Interval for Mean	Lower Bound	73.1966		
			Upper Bound	77.0034		
		5% Trimmed Mean	74.9979		
		Median	75.7500		
		Variance	27.871		
		Std. Deviation	5.27930		
		Minimum	64.10		
		Maximum	88.20		
		Range	24.10		
		Interquartile Range	7.60		
		Skewness	.249	.414	
		Kurtosis	.291	.809	
Body_Weight_W12	1.00	Mean	75.8031	1.01275	
		95% Confidence Interval for Mean	Lower Bound	73.7376		
			Upper Bound	77.8686		
		5% Trimmed Mean	75.5250		
		Median	74.8500		
		Variance	32.821		
		Std. Deviation	5.72896		
		Minimum	67.00		
		Maximum	90.10		
		Range	23.10		
		Interquartile Range	8.50		
		Skewness	.759	.414	
		Kurtosis	.221	.809	
	2.00	Mean	75.3969	.91884	
		95% Confidence Interval for Mean	Lower Bound	73.5229		
			Upper Bound	77.2709		
		5% Trimmed Mean	75.2493		
		Median	75.5000		
		Variance	27.016		
		Std. Deviation	5.19773		
		Minimum	65.40		
		Maximum	89.10		
		Range	23.70		
		Interquartile Range	6.55		
		Skewness	.390	.414	
		Kurtosis	.534	.809	
Body_Weight_Diff_W6	1.00	Mean	-.7969	.29843	
		95% Confidence Interval for Mean	Lower Bound	-1.4055		
			Upper Bound	-.1882		
		5% Trimmed Mean	-.7535		
		Median	-.8000		
		Variance	2.850		
		Std. Deviation	1.68819		
		Minimum	-5.90		
		Maximum	3.00		
		Range	8.90		
		Interquartile Range	2.28		
		Skewness	-.470	.414	
		Kurtosis	1.740	.809	
	2.00	Mean	-.0219	.18916	
		95% Confidence Interval for Mean	Lower Bound	-.4077		
			Upper Bound	.3639		
		5% Trimmed Mean	-.0347		
		Median	-.3000		
		Variance	1.145		
		Std. Deviation	1.07004		
		Minimum	-2.60		
		Maximum	2.90		
		Range	5.50		
		Interquartile Range	1.45		
		Skewness	.267	.414	
		Kurtosis	1.024	.809	
Body_Weight_Diff_W12	1.00	Mean	-1.1500	.34635	
		95% Confidence Interval for Mean	Lower Bound	-1.8564		
			Upper Bound	-.4436		
		5% Trimmed Mean	-1.2486		
		Median	-.9500		
		Variance	3.839		
		Std. Deviation	1.95926		
		Minimum	-4.40		
		Maximum	5.60		
		Range	10.00		
		Interquartile Range	2.40		
		Skewness	.947	.414	
		Kurtosis	3.358	.809	
	2.00	Mean	-.3188	.29988	
		95% Confidence Interval for Mean	Lower Bound	-.9304		
			Upper Bound	.2929		
		5% Trimmed Mean	-.3903		
		Median	-.6500		
		Variance	2.878		
		Std. Deviation	1.69638		
		Minimum	-3.60		
		Maximum	4.70		
		Range	8.30		
		Interquartile Range	2.08		
		Skewness	.877	.414	
		Kurtosis	1.414	.809	
Height_B	1.00	Mean	169.7500	1.20567	
		95% Confidence Interval for Mean	Lower Bound	167.2910		
			Upper Bound	172.2090		
		5% Trimmed Mean	169.6181		
		Median	168.0000		
		Variance	46.516		
		Std. Deviation	6.82027		
		Minimum	158.00		
		Maximum	185.00		
		Range	27.00		
		Interquartile Range	10.75		
		Skewness	.358	.414	
		Kurtosis	-.588	.809	
	2.00	Mean	169.1250	1.11871	
		95% Confidence Interval for Mean	Lower Bound	166.8434		
			Upper Bound	171.4066		
		5% Trimmed Mean	168.9375		
		Median	169.0000		
		Variance	40.048		
		Std. Deviation	6.32838		
		Minimum	159.00		
		Maximum	183.00		
		Range	24.00		
		Interquartile Range	11.00		
		Skewness	.266	.414	
		Kurtosis	-.585	.809	
Height_W6	1.00	Mean	169.7500	1.20567	
		95% Confidence Interval for Mean	Lower Bound	167.2910		
			Upper Bound	172.2090		
		5% Trimmed Mean	169.6181		
		Median	168.0000		
		Variance	46.516		
		Std. Deviation	6.82027		
		Minimum	158.00		
		Maximum	185.00		
		Range	27.00		
		Interquartile Range	10.75		
		Skewness	.358	.414	
		Kurtosis	-.588	.809	
	2.00	Mean	169.1250	1.11871	
		95% Confidence Interval for Mean	Lower Bound	166.8434		
			Upper Bound	171.4066		
		5% Trimmed Mean	168.9375		
		Median	169.0000		
		Variance	40.048		
		Std. Deviation	6.32838		
		Minimum	159.00		
		Maximum	183.00		
		Range	24.00		
		Interquartile Range	11.00		
		Skewness	.266	.414	
		Kurtosis	-.585	.809	
Height_W12	1.00	Mean	169.7500	1.20567	
		95% Confidence Interval for Mean	Lower Bound	167.2910		
			Upper Bound	172.2090		
		5% Trimmed Mean	169.6181		
		Median	168.0000		
		Variance	46.516		
		Std. Deviation	6.82027		
		Minimum	158.00		
		Maximum	185.00		
		Range	27.00		
		Interquartile Range	10.75		
		Skewness	.358	.414	
		Kurtosis	-.588	.809	
	2.00	Mean	169.1250	1.11871	
		95% Confidence Interval for Mean	Lower Bound	166.8434		
			Upper Bound	171.4066		
		5% Trimmed Mean	168.9375		
		Median	169.0000		
		Variance	40.048		
		Std. Deviation	6.32838		
		Minimum	159.00		
		Maximum	183.00		
		Range	24.00		
		Interquartile Range	11.00		
		Skewness	.266	.414	
		Kurtosis	-.585	.809	
Height_Diff_W6	1.00	Mean	.0000	.00000	
		95% Confidence Interval for Mean	Lower Bound	.0000		
			Upper Bound	.0000		
		5% Trimmed Mean	.0000		
		Median	.0000		
		Variance	.000		
		Std. Deviation	.00000		
		Minimum	.00		
		Maximum	.00		
		Range	.00		
		Interquartile Range	.00		
		Skewness	.	.	
		Kurtosis	.	.	
	2.00	Mean	.0000	.00000	
		95% Confidence Interval for Mean	Lower Bound	.0000		
			Upper Bound	.0000		
		5% Trimmed Mean	.0000		
		Median	.0000		
		Variance	.000		
		Std. Deviation	.00000		
		Minimum	.00		
		Maximum	.00		
		Range	.00		
		Interquartile Range	.00		
		Skewness	.	.	
		Kurtosis	.	.	
Height_Diff_W12	1.00	Mean	.0000	.00000	
		95% Confidence Interval for Mean	Lower Bound	.0000		
			Upper Bound	.0000		
		5% Trimmed Mean	.0000		
		Median	.0000		
		Variance	.000		
		Std. Deviation	.00000		
		Minimum	.00		
		Maximum	.00		
		Range	.00		
		Interquartile Range	.00		
		Skewness	.	.	
		Kurtosis	.	.	
	2.00	Mean	.0000	.00000	
		95% Confidence Interval for Mean	Lower Bound	.0000		
			Upper Bound	.0000		
		5% Trimmed Mean	.0000		
		Median	.0000		
		Variance	.000		
		Std. Deviation	.00000		
		Minimum	.00		
		Maximum	.00		
		Range	.00		
		Interquartile Range	.00		
		Skewness	.	.	
		Kurtosis	.	.	
BMI_B	1.00	Mean	25.8794	.14521	
		95% Confidence Interval for Mean	Lower Bound	25.5832		
			Upper Bound	26.1755		
		5% Trimmed Mean	25.8923		
		Median	25.8700		
		Variance	.675		
		Std. Deviation	.82144		
		Minimum	23.31		
		Maximum	27.83		
		Range	4.52		
		Interquartile Range	.97		
		Skewness	-.457	.414	
		Kurtosis	2.389	.809	
	2.00	Mean	26.2325	.18187	
		95% Confidence Interval for Mean	Lower Bound	25.8616		
			Upper Bound	26.6034		
		5% Trimmed Mean	26.1370		
		Median	26.1950		
		Variance	1.059		
		Std. Deviation	1.02884		
		Minimum	25.02		
		Maximum	29.76		
		Range	4.74		
		Interquartile Range	1.18		
		Skewness	1.460	.414	
		Kurtosis	3.265	.809	
BMI_W6	1.00	Mean	26.1628	.13963	
		95% Confidence Interval for Mean	Lower Bound	25.8780		
			Upper Bound	26.4476		
		5% Trimmed Mean	26.1588		
		Median	26.1300		
		Variance	.624		
		Std. Deviation	.78985		
		Minimum	24.76		
		Maximum	27.69		
		Range	2.93		
		Interquartile Range	1.31		
		Skewness	.342	.414	
		Kurtosis	-.698	.809	
	2.00	Mean	26.2503	.19234	
		95% Confidence Interval for Mean	Lower Bound	25.8580		
			Upper Bound	26.6426		
		5% Trimmed Mean	26.1665		
		Median	26.3200		
		Variance	1.184		
		Std. Deviation	1.08806		
		Minimum	24.81		
		Maximum	29.69		
		Range	4.88		
		Interquartile Range	1.40		
		Skewness	1.082	.414	
		Kurtosis	1.864	.809	
BMI_W12	1.00	Mean	26.2912	.15803	
		95% Confidence Interval for Mean	Lower Bound	25.9690		
			Upper Bound	26.6135		
		5% Trimmed Mean	26.3257		
		Median	26.2800		
		Variance	.799		
		Std. Deviation	.89394		
		Minimum	23.51		
		Maximum	28.16		
		Range	4.65		
		Interquartile Range	.92		
		Skewness	-.739	.414	
		Kurtosis	2.345	.809	
	2.00	Mean	26.3606	.20925	
		95% Confidence Interval for Mean	Lower Bound	25.9339		
			Upper Bound	26.7874		
		5% Trimmed Mean	26.3272		
		Median	26.3150		
		Variance	1.401		
		Std. Deviation	1.18367		
		Minimum	24.07		
		Maximum	29.55		
		Range	5.48		
		Interquartile Range	1.56		
		Skewness	.483	.414	
		Kurtosis	.541	.809	
BMI_Diff_W6	1.00	Mean	-.2834	.10379	
		95% Confidence Interval for Mean	Lower Bound	-.4951		
			Upper Bound	-.0718		
		5% Trimmed Mean	-.2733		
		Median	-.3150		
		Variance	.345		
		Std. Deviation	.58711		
		Minimum	-1.93		
		Maximum	1.11		
		Range	3.04		
		Interquartile Range	.79		
		Skewness	-.282	.414	
		Kurtosis	1.232	.809	
	2.00	Mean	-.0178	.06733	
		95% Confidence Interval for Mean	Lower Bound	-.1551		
			Upper Bound	.1195		
		5% Trimmed Mean	-.0189		
		Median	-.1000		
		Variance	.145		
		Std. Deviation	.38090		
		Minimum	-1.02		
		Maximum	1.00		
		Range	2.02		
		Interquartile Range	.52		
		Skewness	.080	.414	
		Kurtosis	1.327	.809	
BMI_Diff_W12	1.00	Mean	-.4119	.12493	
		95% Confidence Interval for Mean	Lower Bound	-.6667		
			Upper Bound	-.1571		
		5% Trimmed Mean	-.4455		
		Median	-.3200		
		Variance	.499		
		Std. Deviation	.70671		
		Minimum	-1.69		
		Maximum	2.06		
		Range	3.75		
		Interquartile Range	.89		
		Skewness	1.009	.414	
		Kurtosis	3.678	.809	
	2.00	Mean	-.1281	.10359	
		95% Confidence Interval for Mean	Lower Bound	-.3394		
			Upper Bound	.0831		
		5% Trimmed Mean	-.1508		
		Median	-.2200		
		Variance	.343		
		Std. Deviation	.58599		
		Minimum	-1.25		
		Maximum	1.53		
		Range	2.78		
		Interquartile Range	.72		
		Skewness	.760	.414	
		Kurtosis	.965	.809	
Hip_Circumf_B
	1.00	Mean	93.5313	1.14122	
		95% Confidence Interval for Mean	Lower Bound	91.2037		
			Upper Bound	95.8588		
		5% Trimmed Mean	93.3750		
		Median	92.5000		
		Variance	41.676		
		Std. Deviation	6.45573		
		Minimum	83.00		
		Maximum	108.00		
		Range	25.00		
		Interquartile Range	7.75		
		Skewness	.554	.414	
		Kurtosis	-.168	.809	
	2.00	Mean	91.6250	.78256	
		95% Confidence Interval for Mean	Lower Bound	90.0290		
			Upper Bound	93.2210		
		5% Trimmed Mean	91.5694		
		Median	91.0000		
		Variance	19.597		
		Std. Deviation	4.42682		
		Minimum	83.00		
		Maximum	102.00		
		Range	19.00		
		Interquartile Range	4.75		
		Skewness	.569	.414	
		Kurtosis	.671	.809	
Hip_Circumf_W6	1.00	Mean	92.3750	1.07927	
		95% Confidence Interval for Mean	Lower Bound	90.1738		
			Upper Bound	94.5762		
		5% Trimmed Mean	92.1667		
		Median	91.5000		
		Variance	37.274		
		Std. Deviation	6.10526		
		Minimum	83.00		
		Maximum	107.00		
		Range	24.00		
		Interquartile Range	7.75		
		Skewness	.528	.414	
		Kurtosis	-.199	.809	
	2.00	Mean	90.7875	.75402	
		95% Confidence Interval for Mean	Lower Bound	89.2497		
			Upper Bound	92.3253		
		5% Trimmed Mean	90.7500		
		Median	90.0000		
		Variance	18.193		
		Std. Deviation	4.26537		
		Minimum	82.00		
		Maximum	101.00		
		Range	19.00		
		Interquartile Range	4.55		
		Skewness	.487	.414	
		Kurtosis	.919	.809	
Hip_Circumf_W12	1.00	Mean	91.8750	1.01476	
		95% Confidence Interval for Mean	Lower Bound	89.8054		
			Upper Bound	93.9446		
		5% Trimmed Mean	91.7361		
		Median	91.0000		
		Variance	32.952		
		Std. Deviation	5.74035		
		Minimum	82.00		
		Maximum	105.00		
		Range	23.00		
		Interquartile Range	6.00		
		Skewness	.455	.414	
		Kurtosis	-.245	.809	
	2.00	Mean	90.5375	.75975	
		95% Confidence Interval for Mean	Lower Bound	88.9880		
			Upper Bound	92.0870		
		5% Trimmed Mean	90.5069		
		Median	90.0000		
		Variance	18.471		
		Std. Deviation	4.29777		
		Minimum	81.00		
		Maximum	101.00		
		Range	20.00		
		Interquartile Range	4.80		
		Skewness	.470	.414	
		Kurtosis	1.121	.809	
Hip_Circumf_Diff_W6	1.00	Mean	1.1563	.20135	
		95% Confidence Interval for Mean	Lower Bound	.7456		
			Upper Bound	1.5669		
		5% Trimmed Mean	1.0278		
		Median	1.0000		
		Variance	1.297		
		Std. Deviation	1.13903		
		Minimum	.00		
		Maximum	5.00		
		Range	5.00		
		Interquartile Range	.00		
		Skewness	2.189	.414	
		Kurtosis	5.068	.809	
	2.00	Mean	.8375	.09089	
		95% Confidence Interval for Mean	Lower Bound	.6521		
			Upper Bound	1.0229		
		5% Trimmed Mean	.8194		
		Median	1.0000		
		Variance	.264		
		Std. Deviation	.51415		
		Minimum	.00		
		Maximum	2.00		
		Range	2.00		
		Interquartile Range	.15		
		Skewness	-.222	.414	
		Kurtosis	.671	.809	
Hip_Circumf_Diff_W12	1.00	Mean	1.6563	.24017	
		95% Confidence Interval for Mean	Lower Bound	1.1664		
			Upper Bound	2.1461		
		5% Trimmed Mean	1.5625		
		Median	2.0000		
		Variance	1.846		
		Std. Deviation	1.35859		
		Minimum	.00		
		Maximum	5.00		
		Range	5.00		
		Interquartile Range	1.00		
		Skewness	1.171	.414	
		Kurtosis	1.574	.809	
	2.00	Mean	1.0875	.13012	
		95% Confidence Interval for Mean	Lower Bound	.8221		
			Upper Bound	1.3529		
		5% Trimmed Mean	1.0625		
		Median	1.0000		
		Variance	.542		
		Std. Deviation	.73605		
		Minimum	.00		
		Maximum	3.00		
		Range	3.00		
		Interquartile Range	.75		
		Skewness	.394	.414	
		Kurtosis	.239	.809	
Waist_Circumf_B	1.00	Mean	85.3125	1.16435	
		95% Confidence Interval for Mean	Lower Bound	82.9378		
			Upper Bound	87.6872		
		5% Trimmed Mean	85.0417		
		Median	85.5000		
		Variance	43.383		
		Std. Deviation	6.58658		
		Minimum	75.00		
		Maximum	102.00		
		Range	27.00		
		Interquartile Range	9.50		
		Skewness	.577	.414	
		Kurtosis	.058	.809	
	2.00	Mean	87.7813	.91110	
		95% Confidence Interval for Mean	Lower Bound	85.9230		
			Upper Bound	89.6395		
		5% Trimmed Mean	87.5764		
		Median	87.5000		
		Variance	26.564		
		Std. Deviation	5.15398		
		Minimum	80.00		
		Maximum	100.00		
		Range	20.00		
		Interquartile Range	6.00		
		Skewness	.745	.414	
		Kurtosis	.133	.809	
Waist_Circumf_W6	1.00	Mean	83.7500	1.14476	
		95% Confidence Interval for Mean	Lower Bound	81.4152		
			Upper Bound	86.0848		
		5% Trimmed Mean	83.3958		
		Median	83.5000		
		Variance	41.935		
		Std. Deviation	6.47576		
		Minimum	74.00		
		Maximum	101.00		
		Range	27.00		
		Interquartile Range	8.50		
		Skewness	.690	.414	
		Kurtosis	.413	.809	
	2.00	Mean	86.8438	.90263	
		95% Confidence Interval for Mean	Lower Bound	85.0028		
			Upper Bound	88.6847		
		5% Trimmed Mean	86.6250		
		Median	86.5000		
		Variance	26.072		
		Std. Deviation	5.10603		
		Minimum	80.00		
		Maximum	98.00		
		Range	18.00		
		Interquartile Range	5.75		
		Skewness	.742	.414	
		Kurtosis	-.167	.809	
Waist_Circumf_W12	1.00	Mean	83.3750	1.12321	
		95% Confidence Interval for Mean	Lower Bound	81.0842		
			Upper Bound	85.6658		
		5% Trimmed Mean	83.0556		
		Median	83.0000		
		Variance	40.371		
		Std. Deviation	6.35382		
		Minimum	74.00		
		Maximum	100.00		
		Range	26.00		
		Interquartile Range	8.25		
		Skewness	.648	.414	
		Kurtosis	.245	.809	
	2.00	Mean	86.6875	.86653	
		95% Confidence Interval for Mean	Lower Bound	84.9202		
			Upper Bound	88.4548		
		5% Trimmed Mean	86.4861		
		Median	86.5000		
		Variance	24.028		
		Std. Deviation	4.90186		
		Minimum	80.00		
		Maximum	97.00		
		Range	17.00		
		Interquartile Range	5.75		
		Skewness	.694	.414	
		Kurtosis	-.192	.809	
Waist_Circumf_Diff_W6	1.00	Mean	1.5625	.22868	
		95% Confidence Interval for Mean	Lower Bound	1.0961		
			Upper Bound	2.0289		
		5% Trimmed Mean	1.4444		
		Median	1.0000		
		Variance	1.673		
		Std. Deviation	1.29359		
		Minimum	.00		
		Maximum	6.00		
		Range	6.00		
		Interquartile Range	1.00		
		Skewness	1.379	.414	
		Kurtosis	3.301	.809	
	2.00	Mean	.9375	.37010	
		95% Confidence Interval for Mean	Lower Bound	.1827		
			Upper Bound	1.6923		
		5% Trimmed Mean	1.0903		
		Median	1.0000		
		Variance	4.383		
		Std. Deviation	2.09358		
		Minimum	-7.00		
		Maximum	5.00		
		Range	12.00		
		Interquartile Range	2.00		
		Skewness	-1.757	.414	
		Kurtosis	5.963	.809	
Waist_Circumf_Diff_W12	1.00	Mean	1.9375	.25375	
		95% Confidence Interval for Mean	Lower Bound	1.4200		
			Upper Bound	2.4550		
		5% Trimmed Mean	1.8056		
		Median	2.0000		
		Variance	2.060		
		Std. Deviation	1.43544		
		Minimum	.00		
		Maximum	7.00		
		Range	7.00		
		Interquartile Range	1.00		
		Skewness	1.582	.414	
		Kurtosis	4.327	.809	
	2.00	Mean	1.0938	.37395	
		95% Confidence Interval for Mean	Lower Bound	.3311		
			Upper Bound	1.8564		
		5% Trimmed Mean	1.2431		
		Median	1.0000		
		Variance	4.475		
		Std. Deviation	2.11537		
		Minimum	-7.00		
		Maximum	5.00		
		Range	12.00		
		Interquartile Range	2.00		
		Skewness	-1.636	.414	
		Kurtosis	6.167	.809	
Chest_Circumf_B	1.00	Mean	91.0938	.86411	
		95% Confidence Interval for Mean	Lower Bound	89.3314		
			Upper Bound	92.8561		
		5% Trimmed Mean	90.9444		
		Median	90.0000		
		Variance	23.894		
		Std. Deviation	4.88816		
		Minimum	82.00		
		Maximum	104.00		
		Range	22.00		
		Interquartile Range	4.00		
		Skewness	.548	.414	
		Kurtosis	.788	.809	
	2.00	Mean	89.0625	.53116	
		95% Confidence Interval for Mean	Lower Bound	87.9792		
			Upper Bound	90.1458		
		5% Trimmed Mean	88.9931		
		Median	89.0000		
		Variance	9.028		
		Std. Deviation	3.00470		
		Minimum	83.00		
		Maximum	96.00		
		Range	13.00		
		Interquartile Range	4.00		
		Skewness	.408	.414	
		Kurtosis	-.074	.809	
Chest_Circumf_W6	1.00	Mean	93.0938	.65587	
		95% Confidence Interval for Mean	Lower Bound	91.7561		
			Upper Bound	94.4314		
		5% Trimmed Mean	92.9792		
		Median	93.0000		
		Variance	13.765		
		Std. Deviation	3.71014		
		Minimum	86.00		
		Maximum	103.00		
		Range	17.00		
		Interquartile Range	5.00		
		Skewness	.443	.414	
		Kurtosis	.297	.809	
	2.00	Mean	89.9063	.46740	
		95% Confidence Interval for Mean	Lower Bound	88.9530		
			Upper Bound	90.8595		
		5% Trimmed Mean	89.8750		
		Median	89.0000		
		Variance	6.991		
		Std. Deviation	2.64404		
		Minimum	84.00		
		Maximum	96.00		
		Range	12.00		
		Interquartile Range	3.75		
		Skewness	.242	.414	
		Kurtosis	.139	.809	
Chest_Circumf_W12	1.00	Mean	93.4375	.61882	
		95% Confidence Interval for Mean	Lower Bound	92.1754		
			Upper Bound	94.6996		
		5% Trimmed Mean	93.3056		
		Median	93.5000		
		Variance	12.254		
		Std. Deviation	3.50058		
		Minimum	87.00		
		Maximum	103.00		
		Range	16.00		
		Interquartile Range	4.00		
		Skewness	.528	.414	
		Kurtosis	.468	.809	
	2.00	Mean	90.3125	.46541	
		95% Confidence Interval for Mean	Lower Bound	89.3633		
			Upper Bound	91.2617		
		5% Trimmed Mean	90.3264		
		Median	90.0000		
		Variance	6.931		
		Std. Deviation	2.63277		
		Minimum	84.00		
		Maximum	96.00		
		Range	12.00		
		Interquartile Range	3.00		
		Skewness	.078	.414	
		Kurtosis	.190	.809	
Chest_Circumf_Diff_W6	1.00	Mean	-2.0000	.71279	
		95% Confidence Interval for Mean	Lower Bound	-3.4537		
			Upper Bound	-.5463		
		5% Trimmed Mean	-1.8333		
		Median	-1.5000		
		Variance	16.258		
		Std. Deviation	4.03213		
		Minimum	-11.00		
		Maximum	4.00		
		Range	15.00		
		Interquartile Range	6.00		
		Skewness	-.551	.414	
		Kurtosis	-.471	.809	
	2.00	Mean	-.8438	.18022	
		95% Confidence Interval for Mean	Lower Bound	-1.2113		
			Upper Bound	-.4762		
		5% Trimmed Mean	-.8819		
		Median	-1.0000		
		Variance	1.039		
		Std. Deviation	1.01947		
		Minimum	-3.00		
		Maximum	2.00		
		Range	5.00		
		Interquartile Range	1.75		
		Skewness	.447	.414	
		Kurtosis	.878	.809	
Chest_Circumf_Diff_W12	1.00	Mean	-2.3438	.71928	
		95% Confidence Interval for Mean	Lower Bound	-3.8107		
			Upper Bound	-.8768		
		5% Trimmed Mean	-2.1250		
		Median	-2.0000		
		Variance	16.555		
		Std. Deviation	4.06884		
		Minimum	-12.00		
		Maximum	3.00		
		Range	15.00		
		Interquartile Range	6.00		
		Skewness	-.633	.414	
		Kurtosis	-.119	.809	
	2.00	Mean	-1.2500	.19050	
		95% Confidence Interval for Mean	Lower Bound	-1.6385		
			Upper Bound	-.8615		
		5% Trimmed Mean	-1.2778		
		Median	-1.0000		
		Variance	1.161		
		Std. Deviation	1.07763		
		Minimum	-3.00		
		Maximum	1.00		
		Range	4.00		
		Interquartile Range	1.00		
		Skewness	.206	.414	
		Kurtosis	-.298	.809	
Body_Fat_B	1.00	Mean	18.5438	1.03488	
		95% Confidence Interval for Mean	Lower Bound	16.4331		
			Upper Bound	20.6544		
		5% Trimmed Mean	18.3382		
		Median	16.3500		
		Variance	34.272		
		Std. Deviation	5.85419		
		Minimum	9.30		
		Maximum	31.30		
		Range	22.00		
		Interquartile Range	7.67		
		Skewness	.728	.414	
		Kurtosis	-.534	.809	
	2.00	Mean	18.2500	.96166	
		95% Confidence Interval for Mean	Lower Bound	16.2887		
			Upper Bound	20.2113		
		5% Trimmed Mean	18.0778		
		Median	17.5000		
		Variance	29.594		
		Std. Deviation	5.44000		
		Minimum	8.20		
		Maximum	30.20		
		Range	22.00		
		Interquartile Range	6.92		
		Skewness	.659	.414	
		Kurtosis	-.131	.809	
Body_Fat_W6	1.00	Mean	17.1219	.93900	
		95% Confidence Interval for Mean	Lower Bound	15.2068		
			Upper Bound	19.0370		
		5% Trimmed Mean	17.0174		
		Median	14.9000		
		Variance	28.215		
		Std. Deviation	5.31181		
		Minimum	8.20		
		Maximum	27.10		
		Range	18.90		
		Interquartile Range	6.90		
		Skewness	.639	.414	
		Kurtosis	-.811	.809	
	2.00	Mean	17.8531	.93792	
		95% Confidence Interval for Mean	Lower Bound	15.9402		
			Upper Bound	19.7660		
		5% Trimmed Mean	17.5132		
		Median	16.5500		
		Variance	28.150		
		Std. Deviation	5.30569		
		Minimum	11.20		
		Maximum	32.20		
		Range	21.00		
		Interquartile Range	7.35		
		Skewness	.961	.414	
		Kurtosis	.353	.809	
Body_Fat_W12	1.00	Mean	16.8844	.86678	
		95% Confidence Interval for Mean	Lower Bound	15.1166		
			Upper Bound	18.6522		
		5% Trimmed Mean	16.7201		
		Median	15.2000		
		Variance	24.042		
		Std. Deviation	4.90326		
		Minimum	8.00		
		Maximum	27.90		
		Range	19.90		
		Interquartile Range	7.52		
		Skewness	.687	.414	
		Kurtosis	-.396	.809	
	2.00	Mean	17.5156	.88833	
		95% Confidence Interval for Mean	Lower Bound	15.7039		
			Upper Bound	19.3274		
		5% Trimmed Mean	17.4028		
		Median	17.0000		
		Variance	25.252		
		Std. Deviation	5.02517		
		Minimum	9.60		
		Maximum	26.90		
		Range	17.30		
		Interquartile Range	7.45		
		Skewness	.456	.414	
		Kurtosis	-.867	.809	
Body_Fat_Diff_W6	1.00	Mean	1.4219	.32505	
		95% Confidence Interval for Mean	Lower Bound	.7589		
			Upper Bound	2.0848		
		5% Trimmed Mean	1.3958		
		Median	1.0000		
		Variance	3.381		
		Std. Deviation	1.83878		
		Minimum	-3.20		
		Maximum	7.00		
		Range	10.20		
		Interquartile Range	2.25		
		Skewness	.434	.414	
		Kurtosis	2.607	.809	
	2.00	Mean	.3969	.29308	
		95% Confidence Interval for Mean	Lower Bound	-.2009		
			Upper Bound	.9946		
		5% Trimmed Mean	.2743		
		Median	.0000		
		Variance	2.749		
		Std. Deviation	1.65792		
		Minimum	-3.00		
		Maximum	6.60		
		Range	9.60		
		Interquartile Range	1.30		
		Skewness	1.721	.414	
		Kurtosis	6.197	.809	
Body_Fat_Diff_W12	1.00	Mean	1.6594	.28631	
		95% Confidence Interval for Mean	Lower Bound	1.0754		
			Upper Bound	2.2433		
		5% Trimmed Mean	1.5917		
		Median	1.4000		
		Variance	2.623		
		Std. Deviation	1.61961		
		Minimum	-1.20		
		Maximum	6.00		
		Range	7.20		
		Interquartile Range	2.50		
		Skewness	.671	.414	
		Kurtosis	.338	.809	
	2.00	Mean	.7344	.32542	
		95% Confidence Interval for Mean	Lower Bound	.0707		
			Upper Bound	1.3981		
		5% Trimmed Mean	.6910		
		Median	.6500		
		Variance	3.389		
		Std. Deviation	1.84086		
		Minimum	-4.40		
		Maximum	6.50		
		Range	10.90		
		Interquartile Range	1.38		
		Skewness	.548	.414	
		Kurtosis	3.702	.809	
Skeletal_Muscle_B	1.00	Mean	38.4125	.87281	
		95% Confidence Interval for Mean	Lower Bound	36.6324		
			Upper Bound	40.1926		
		5% Trimmed Mean	38.3194		
		Median	37.9500		
		Variance	24.377		
		Std. Deviation	4.93733		
		Minimum	30.30		
		Maximum	48.80		
		Range	18.50		
		Interquartile Range	7.42		
		Skewness	.240	.414	
		Kurtosis	-.760	.809	
	2.00	Mean	37.4156	.81690	
		95% Confidence Interval for Mean	Lower Bound	35.7496		
			Upper Bound	39.0817		
		5% Trimmed Mean	37.3153		
		Median	37.6000		
		Variance	21.354		
		Std. Deviation	4.62107		
		Minimum	30.50		
		Maximum	46.10		
		Range	15.60		
		Interquartile Range	8.20		
		Skewness	.210	.414	
		Kurtosis	-.892	.809	
Skeletal_Muscle_W6	1.00	Mean	39.3469	.91956	
		95% Confidence Interval for Mean	Lower Bound	37.4714		
			Upper Bound	41.2223		
		5% Trimmed Mean	39.2743		
		Median	39.6000		
		Variance	27.059		
		Std. Deviation	5.20180		
		Minimum	31.60		
		Maximum	49.00		
		Range	17.40		
		Interquartile Range	9.38		
		Skewness	.013	.414	
		Kurtosis	-1.138	.809	
	2.00	Mean	37.7437	.82370	
		95% Confidence Interval for Mean	Lower Bound	36.0638		
			Upper Bound	39.4237		
		5% Trimmed Mean	37.7000		
		Median	38.1000		
		Variance	21.712		
		Std. Deviation	4.65957		
		Minimum	30.10		
		Maximum	46.10		
		Range	16.00		
		Interquartile Range	8.40		
		Skewness	.065	.414	
		Kurtosis	-.941	.809	
Skeletal_Muscle_W12	1.00	Mean	39.7000	.92672	
		95% Confidence Interval for Mean	Lower Bound	37.8099		
			Upper Bound	41.5901		
		5% Trimmed Mean	39.6868		
		Median	39.6500		
		Variance	27.482		
		Std. Deviation	5.24232		
		Minimum	30.90		
		Maximum	49.00		
		Range	18.10		
		Interquartile Range	8.85		
		Skewness	-.016	.414	
		Kurtosis	-1.049	.809	
	2.00	Mean	38.0375	.80650	
		95% Confidence Interval for Mean	Lower Bound	36.3926		
			Upper Bound	39.6824		
		5% Trimmed Mean	37.9812		
		Median	38.4500		
		Variance	20.814		
		Std. Deviation	4.56224		
		Minimum	30.90		
		Maximum	46.30		
		Range	15.40		
		Interquartile Range	7.37		
		Skewness	-.023	.414	
		Kurtosis	-.922	.809	
Skeletal_Muscle_Diff_W6	1.00	Mean	-.9344	.18609	
		95% Confidence Interval for Mean	Lower Bound	-1.3139		
			Upper Bound	-.5548		
		5% Trimmed Mean	-.9236		
		Median	-.9500		
		Variance	1.108		
		Std. Deviation	1.05268		
		Minimum	-3.20		
		Maximum	1.10		
		Range	4.30		
		Interquartile Range	1.60		
		Skewness	-.243	.414	
		Kurtosis	-.006	.809	
	2.00	Mean	-.3281	.13502	
		95% Confidence Interval for Mean	Lower Bound	-.6035		
			Upper Bound	-.0527		
		5% Trimmed Mean	-.3007		
		Median	.0000		
		Variance	.583		
		Std. Deviation	.76379		
		Minimum	-2.00		
		Maximum	1.00		
		Range	3.00		
		Interquartile Range	1.15		
		Skewness	-.642	.414	
		Kurtosis	-.373	.809	
Skeletal_Muscle_Diff_W12	1.00	Mean	-1.2875	.20224	
		95% Confidence Interval for Mean	Lower Bound	-1.7000		
			Upper Bound	-.8750		
		5% Trimmed Mean	-1.2243		
		Median	-1.0000		
		Variance	1.309		
		Std. Deviation	1.14406		
		Minimum	-4.20		
		Maximum	.60		
		Range	4.80		
		Interquartile Range	1.48		
		Skewness	-.928	.414	
		Kurtosis	.629	.809	
	2.00	Mean	-.6219	.17619	
		95% Confidence Interval for Mean	Lower Bound	-.9812		
			Upper Bound	-.2625		
		5% Trimmed Mean	-.5715		
		Median	-.3000		
		Variance	.993		
		Std. Deviation	.99668		
		Minimum	-4.00		
		Maximum	1.70		
		Range	5.70		
		Interquartile Range	.98		
		Skewness	-1.155	.414	
		Kurtosis	3.693	.809	

	
