## Supplementary material for "Effects of a standardised *Amaranthus tricolor*–*Spinacia oleracea* extract on exercise performance, muscle strength, endurance and recovery in physically active adults: a randomised, double-blind, placebo-controlled trial": Raw statistical outputs: Bench___Leg_press_Output_17.07.2026.doc

Explore


Notes	
Output Created	17-JUL-2026 11:26:14	
Comments		
Input	Active Dataset	DataSet0	
	Filter	<none>	
	Weight	<none>	
	Split File	<none>	
	N of Rows in Working Data File	64	
Missing Value Handling	Definition of Missing	User-defined missing values for dependent variables are treated as missing.	
	Cases Used	Statistics are based on cases with no missing values for any dependent variable or factor used.	
Syntax	EXAMINE VARIABLES=Bench_1RM_B Bench_1RM_W6 Bench_1RM_W12 Bench_1RM_Diff_W6 Bench_1RM_Diff_W12
    Leg_1RM_B Leg_1RM_W6 Leg_1RM_W12 Leg_1RM_Diff_W6 Leg_1RM_Diff_W12 Bench_KG_B Bench_KG_W6
    Bench_KG_W12 Bench_KG_Diff_W6 Bench_KG_Diff_W12 Bench_Reps_B Bench_Reps_W6 Bench_Reps_W12
    Bench_Reps_Diff_W6 Bench_Reps_Diff_W12 Leg_KG_B Leg_KG_W6 Leg_KG_W12 Leg_KG_Diff_W6 Leg_KG_Diff_W12
    Leg_Reps_B Leg_Reps_W6 Leg_Reps_W12 Leg_Reps_Diff_W6 Leg_Reps_Diff_W12 Fatigue_Index_Leg_B
    Fatigue_Index_Leg_W6 Fatigue_Index_Leg_W12 Fatigue_Index_Leg_Diff_W6 Fatigue_Index_Leg_Diff_W12
    Fatigue_Index_Bench_B Fatigue_Index_Bench_W6 Fatigue_Index_Bench_W12 Fatigue_Index_Bench_Diff_W6
    Fatigue_Index_Bench_Diff_W12 BY Group
  /PLOT BOXPLOT STEMLEAF
  /COMPARE GROUPS
  /STATISTICS DESCRIPTIVES
  /CINTERVAL 95
  /MISSING LISTWISE
  /NOTOTAL.	
Resources	Processor Time	00:00:03.78	
	Elapsed Time	00:00:03.25	


Group


Case Processing Summary	
	Group	Cases	
		Valid	Missing	Total	
		N	Percent	N	Percent	N	Percent	
Bench_1RM_B	1.00	32	100.0%	0	0.0%	32	100.0%	
	2.00	32	100.0%	0	0.0%	32	100.0%	
Bench_1RM_W6	1.00	32	100.0%	0	0.0%	32	100.0%	
	2.00	32	100.0%	0	0.0%	32	100.0%	
Bench_1RM_W12	1.00	32	100.0%	0	0.0%	32	100.0%	
	2.00	32	100.0%	0	0.0%	32	100.0%	
Bench_1RM_Diff_W6	1.00	32	100.0%	0	0.0%	32	100.0%	
	2.00	32	100.0%	0	0.0%	32	100.0%	
Bench_1RM_Diff_W12	1.00	32	100.0%	0	0.0%	32	100.0%	
	2.00	32	100.0%	0	0.0%	32	100.0%	
Leg_1RM_B	1.00	32	100.0%	0	0.0%	32	100.0%	
	2.00	32	100.0%	0	0.0%	32	100.0%	
Leg_1RM_W6	1.00	32	100.0%	0	0.0%	32	100.0%	
	2.00	32	100.0%	0	0.0%	32	100.0%	
Leg_1RM_W12	1.00	32	100.0%	0	0.0%	32	100.0%	
	2.00	32	100.0%	0	0.0%	32	100.0%	
Leg_1RM_Diff_W6	1.00	32	100.0%	0	0.0%	32	100.0%	
	2.00	32	100.0%	0	0.0%	32	100.0%	
Leg_1RM_Diff_W12	1.00	32	100.0%	0	0.0%	32	100.0%	
	2.00	32	100.0%	0	0.0%	32	100.0%	
Bench_KG_B	1.00	32	100.0%	0	0.0%	32	100.0%	
	2.00	32	100.0%	0	0.0%	32	100.0%	
Bench_KG_W6	1.00	32	100.0%	0	0.0%	32	100.0%	
	2.00	32	100.0%	0	0.0%	32	100.0%	
Bench_KG_W12	1.00	32	100.0%	0	0.0%	32	100.0%	
	2.00	32	100.0%	0	0.0%	32	100.0%	
Bench_KG_Diff_W6	1.00	32	100.0%	0	0.0%	32	100.0%	
	2.00	32	100.0%	0	0.0%	32	100.0%	
Bench_KG_Diff_W12	1.00	32	100.0%	0	0.0%	32	100.0%	
	2.00	32	100.0%	0	0.0%	32	100.0%	
Bench_Reps_B	1.00	32	100.0%	0	0.0%	32	100.0%	
	2.00	32	100.0%	0	0.0%	32	100.0%	
Bench_Reps_W6	1.00	32	100.0%	0	0.0%	32	100.0%	
	2.00	32	100.0%	0	0.0%	32	100.0%	
Bench_Reps_W12	1.00	32	100.0%	0	0.0%	32	100.0%	
	2.00	32	100.0%	0	0.0%	32	100.0%	
Bench_Reps_Diff_W6	1.00	32	100.0%	0	0.0%	32	100.0%	
	2.00	32	100.0%	0	0.0%	32	100.0%	
Bench_Reps_Diff_W12	1.00	32	100.0%	0	0.0%	32	100.0%	
	2.00	32	100.0%	0	0.0%	32	100.0%	
Leg_KG_B	1.00	32	100.0%	0	0.0%	32	100.0%	
	2.00	32	100.0%	0	0.0%	32	100.0%	
Leg_KG_W6	1.00	32	100.0%	0	0.0%	32	100.0%	
	2.00	32	100.0%	0	0.0%	32	100.0%	
Leg_KG_W12	1.00	32	100.0%	0	0.0%	32	100.0%	
	2.00	32	100.0%	0	0.0%	32	100.0%	
Leg_KG_Diff_W6	1.00	32	100.0%	0	0.0%	32	100.0%	
	2.00	32	100.0%	0	0.0%	32	100.0%	
Leg_KG_Diff_W12	1.00	32	100.0%	0	0.0%	32	100.0%	
	2.00	32	100.0%	0	0.0%	32	100.0%	
Leg_Reps_B	1.00	32	100.0%	0	0.0%	32	100.0%	
	2.00	32	100.0%	0	0.0%	32	100.0%	
Leg_Reps_W6	1.00	32	100.0%	0	0.0%	32	100.0%	
	2.00	32	100.0%	0	0.0%	32	100.0%	
Leg_Reps_W12	1.00	32	100.0%	0	0.0%	32	100.0%	
	2.00	32	100.0%	0	0.0%	32	100.0%	
Leg_Reps_Diff_W6	1.00	32	100.0%	0	0.0%	32	100.0%	
	2.00	32	100.0%	0	0.0%	32	100.0%	
Leg_Reps_Diff_W12	1.00	32	100.0%	0	0.0%	32	100.0%	
	2.00	32	100.0%	0	0.0%	32	100.0%	
Fatigue_Index_Leg_B	1.00	32	100.0%	0	0.0%	32	100.0%	
	2.00	32	100.0%	0	0.0%	32	100.0%	
Fatigue_Index_Leg_W6	1.00	32	100.0%	0	0.0%	32	100.0%	
	2.00	32	100.0%	0	0.0%	32	100.0%	
Fatigue_Index_Leg_W12	1.00	32	100.0%	0	0.0%	32	100.0%	
	2.00	32	100.0%	0	0.0%	32	100.0%	
Fatigue_Index_Leg_Diff_W6	1.00	32	100.0%	0	0.0%	32	100.0%	
	2.00	32	100.0%	0	0.0%	32	100.0%	
Fatigue_Index_Leg_Diff_W12	1.00	32	100.0%	0	0.0%	32	100.0%	
	2.00	32	100.0%	0	0.0%	32	100.0%	
Fatigue_Index_Bench_B	1.00	32	100.0%	0	0.0%	32	100.0%	
	2.00	32	100.0%	0	0.0%	32	100.0%	
Fatigue_Index_Bench_W6	1.00	32	100.0%	0	0.0%	32	100.0%	
	2.00	32	100.0%	0	0.0%	32	100.0%	
Fatigue_Index_Bench_W12	1.00	32	100.0%	0	0.0%	32	100.0%	
	2.00	32	100.0%	0	0.0%	32	100.0%	
Fatigue_Index_Bench_Diff_W6	1.00	32	100.0%	0	0.0%	32	100.0%	
	2.00	32	100.0%	0	0.0%	32	100.0%	
Fatigue_Index_Bench_Diff_W12	1.00	32	100.0%	0	0.0%	32	100.0%	
	2.00	32	100.0%	0	0.0%	32	100.0%	


Descriptives	
	Group	Statistic	Std. Error	
Bench_1RM_B	1.00	Mean	71.1174	1.46058	
		95% Confidence Interval for Mean	Lower Bound	68.1385		
			Upper Bound	74.0963		
		5% Trimmed Mean	71.0130		
		Median	70.4116		
		Variance	68.265		
		Std. Deviation	8.26227		
		Minimum	57.18		
		Maximum	87.00		
		Range	29.82		
		Interquartile Range	12.71		
		Skewness	.188	.414	
		Kurtosis	-1.114	.809	
	2.00	Mean	70.6029	1.39851	
		95% Confidence Interval for Mean	Lower Bound	67.7506		
			Upper Bound	73.4552		
		5% Trimmed Mean	71.0226		
		Median	70.4151		
		Variance	62.587		
		Std. Deviation	7.91117		
		Minimum	44.00		
		Maximum	84.71		
		Range	40.71		
		Interquartile Range	11.42		
		Skewness	-.994	.414	
		Kurtosis	2.713	.809	
Bench_1RM_W6	1.00	Mean	74.3603	1.39304	
		95% Confidence Interval for Mean	Lower Bound	71.5191		
			Upper Bound	77.2014		
		5% Trimmed Mean	74.1711		
		Median	73.0481		
		Variance	62.098		
		Std. Deviation	7.88023		
		Minimum	62.47		
		Maximum	90.01		
		Range	27.54		
		Interquartile Range	14.73		
		Skewness	.328	.414	
		Kurtosis	-1.084	.809	
	2.00	Mean	72.5546	1.12465	
		95% Confidence Interval for Mean	Lower Bound	70.2608		
			Upper Bound	74.8483		
		5% Trimmed Mean	72.3126		
		Median	70.3688		
		Variance	40.475		
		Std. Deviation	6.36198		
		Minimum	63.28		
		Maximum	86.19		
		Range	22.91		
		Interquartile Range	11.73		
		Skewness	.554	.414	
		Kurtosis	-.923	.809	
Bench_1RM_W12	1.00	Mean	77.6677	1.18112	
		95% Confidence Interval for Mean	Lower Bound	75.2588		
			Upper Bound	80.0766		
		5% Trimmed Mean	77.5747		
		Median	78.7579		
		Variance	44.642		
		Std. Deviation	6.68143		
		Minimum	66.38		
		Maximum	91.00		
		Range	24.62		
		Interquartile Range	10.69		
		Skewness	.068	.414	
		Kurtosis	-.872	.809	
	2.00	Mean	72.9010	1.17517	
		95% Confidence Interval for Mean	Lower Bound	70.5042		
			Upper Bound	75.2978		
		5% Trimmed Mean	72.7829		
		Median	70.3527		
		Variance	44.193		
		Std. Deviation	6.64779		
		Minimum	63.28		
		Maximum	84.38		
		Range	21.11		
		Interquartile Range	13.64		
		Skewness	.367	.414	
		Kurtosis	-1.391	.809	
Bench_1RM_Diff_W6	1.00	Mean	-3.2429	1.33877	
		95% Confidence Interval for Mean	Lower Bound	-5.9733		
			Upper Bound	-.5124		
		5% Trimmed Mean	-3.3738		
		Median	-2.1829		
		Variance	57.354		
		Std. Deviation	7.57323		
		Minimum	-16.14		
		Maximum	12.00		
		Range	28.14		
		Interquartile Range	8.38		
		Skewness	.270	.414	
		Kurtosis	-.379	.809	
	2.00	Mean	-1.9517	1.22383	
		95% Confidence Interval for Mean	Lower Bound	-4.4477		
			Upper Bound	.5444		
		5% Trimmed Mean	-1.6508		
		Median	-2.0554		
		Variance	47.928		
		Std. Deviation	6.92303		
		Minimum	-21.46		
		Maximum	9.78		
		Range	31.24		
		Interquartile Range	2.39		
		Skewness	-.529	.414	
		Kurtosis	1.142	.809	
Bench_1RM_Diff_W12	1.00	Mean	-6.5503	1.24704	
		95% Confidence Interval for Mean	Lower Bound	-9.0937		
			Upper Bound	-4.0070		
		5% Trimmed Mean	-6.6694		
		Median	-4.7372		
		Variance	49.764		
		Std. Deviation	7.05433		
		Minimum	-19.64		
		Maximum	8.69		
		Range	28.33		
		Interquartile Range	9.82		
		Skewness	.106	.414	
		Kurtosis	-.387	.809	
	2.00	Mean	-2.2981	1.65521	
		95% Confidence Interval for Mean	Lower Bound	-5.6739		
			Upper Bound	1.0777		
		5% Trimmed Mean	-1.9742		
		Median	-2.4247		
		Variance	87.671		
		Std. Deviation	9.36331		
		Minimum	-32.00		
		Maximum	19.45		
		Range	51.45		
		Interquartile Range	9.72		
		Skewness	-.693	.414	
		Kurtosis	2.586	.809	
Leg_1RM_B	1.00	Mean	90.0003	1.29026	
		95% Confidence Interval for Mean	Lower Bound	87.3688		
			Upper Bound	92.6318		
		5% Trimmed Mean	89.9951		
		Median	90.0042		
		Variance	53.273		
		Std. Deviation	7.29884		
		Minimum	75.18		
		Maximum	103.77		
		Range	28.59		
		Interquartile Range	10.06		
		Skewness	.003	.414	
		Kurtosis	-.271	.809	
	2.00	Mean	88.7903	1.56315	
		95% Confidence Interval for Mean	Lower Bound	85.6022		
			Upper Bound	91.9783		
		5% Trimmed Mean	89.4169		
		Median	90.0042		
		Variance	78.190		
		Std. Deviation	8.84251		
		Minimum	56.00		
		Maximum	102.71		
		Range	46.71		
		Interquartile Range	11.12		
		Skewness	-1.549	.414	
		Kurtosis	4.867	.809	
Leg_1RM_W6	1.00	Mean	93.5241	1.20886	
		95% Confidence Interval for Mean	Lower Bound	91.0586		
			Upper Bound	95.9896		
		5% Trimmed Mean	93.5515		
		Median	94.3705		
		Variance	46.763		
		Std. Deviation	6.83837		
		Minimum	80.73		
		Maximum	106.92		
		Range	26.18		
		Interquartile Range	9.68		
		Skewness	-.249	.414	
		Kurtosis	-.545	.809	
	2.00	Mean	91.4634	1.41963	
		95% Confidence Interval for Mean	Lower Bound	88.5680		
			Upper Bound	94.3587		
		5% Trimmed Mean	91.6008		
		Median	92.4280		
		Variance	64.491		
		Std. Deviation	8.03062		
		Minimum	74.12		
		Maximum	105.83		
		Range	31.70		
		Interquartile Range	10.93		
		Skewness	-.271	.414	
		Kurtosis	-.599	.809	
Leg_1RM_W12	1.00	Mean	97.1572	1.10177	
		95% Confidence Interval for Mean	Lower Bound	94.9102		
			Upper Bound	99.4043		
		5% Trimmed Mean	97.4282		
		Median	97.8848		
		Variance	38.845		
		Std. Deviation	6.23256		
		Minimum	81.01		
		Maximum	109.14		
		Range	28.13		
		Interquartile Range	6.47		
		Skewness	-.945	.414	
		Kurtosis	1.149	.809	
	2.00	Mean	92.5878	1.33561	
		95% Confidence Interval for Mean	Lower Bound	89.8639		
			Upper Bound	95.3118		
		5% Trimmed Mean	92.6075		
		Median	94.9160		
		Variance	57.083		
		Std. Deviation	7.55536		
		Minimum	77.46		
		Maximum	109.14		
		Range	31.68		
		Interquartile Range	10.56		
		Skewness	-.283	.414	
		Kurtosis	-.201	.809	
Leg_1RM_Diff_W6	1.00	Mean	-3.5238	.85851	
		95% Confidence Interval for Mean	Lower Bound	-5.2748		
			Upper Bound	-1.7729		
		5% Trimmed Mean	-3.5970		
		Median	-3.6602		
		Variance	23.585		
		Std. Deviation	4.85648		
		Minimum	-15.05		
		Maximum	9.14		
		Range	24.20		
		Interquartile Range	3.69		
		Skewness	.336	.414	
		Kurtosis	1.913	.809	
	2.00	Mean	-2.6731	.80882	
		95% Confidence Interval for Mean	Lower Bound	-4.3227		
			Upper Bound	-1.0235		
		5% Trimmed Mean	-2.4739		
		Median	-2.5531		
		Variance	20.934		
		Std. Deviation	4.57538		
		Minimum	-18.12		
		Maximum	8.42		
		Range	26.54		
		Interquartile Range	3.86		
		Skewness	-.971	.414	
		Kurtosis	3.935	.809	
Leg_1RM_Diff_W12	1.00	Mean	-7.1569	.98594	
		95% Confidence Interval for Mean	Lower Bound	-9.1678		
			Upper Bound	-5.1461		
		5% Trimmed Mean	-7.3206		
		Median	-7.0866		
		Variance	31.107		
		Std. Deviation	5.57735		
		Minimum	-18.47		
		Maximum	6.48		
		Range	24.95		
		Interquartile Range	4.83		
		Skewness	.852	.414	
		Kurtosis	1.245	.809	
	2.00	Mean	-3.7976	.83469	
		95% Confidence Interval for Mean	Lower Bound	-5.4999		
			Upper Bound	-2.0952		
		5% Trimmed Mean	-3.5910		
		Median	-3.6923		
		Variance	22.294		
		Std. Deviation	4.72169		
		Minimum	-21.46		
		Maximum	6.86		
		Range	28.32		
		Interquartile Range	5.11		
		Skewness	-1.258	.414	
		Kurtosis	5.847	.809	
Bench_KG_B	1.00	Mean	49.3875	.95604	
		95% Confidence Interval for Mean	Lower Bound	47.4376		
			Upper Bound	51.3373		
		5% Trimmed Mean	49.3683		
		Median	48.5275		
		Variance	29.248		
		Std. Deviation	5.40817		
		Minimum	40.03		
		Maximum	59.30		
		Range	19.27		
		Interquartile Range	8.73		
		Skewness	.177	.414	
		Kurtosis	-1.140	.809	
	2.00	Mean	49.7103	.83373	
		95% Confidence Interval for Mean	Lower Bound	48.0099		
			Upper Bound	51.4107		
		5% Trimmed Mean	49.7158		
		Median	49.2906		
		Variance	22.243		
		Std. Deviation	4.71630		
		Minimum	40.03		
		Maximum	59.30		
		Range	19.27		
		Interquartile Range	7.99		
		Skewness	.041	.414	
		Kurtosis	-.796	.809	
Bench_KG_W6	1.00	Mean	52.0624	.97765	
		95% Confidence Interval for Mean	Lower Bound	50.0685		
			Upper Bound	54.0563		
		5% Trimmed Mean	51.9257		
		Median	51.1674		
		Variance	30.586		
		Std. Deviation	5.53043		
		Minimum	43.73		
		Maximum	63.01		
		Range	19.27		
		Interquartile Range	10.31		
		Skewness	.334	.414	
		Kurtosis	-1.070	.809	
	2.00	Mean	50.7882	.78726	
		95% Confidence Interval for Mean	Lower Bound	49.1826		
			Upper Bound	52.3938		
		5% Trimmed Mean	50.6188		
		Median	49.2581		
		Variance	19.833		
		Std. Deviation	4.45339		
		Minimum	44.29		
		Maximum	60.33		
		Range	16.04		
		Interquartile Range	8.21		
		Skewness	.554	.414	
		Kurtosis	-.923	.809	
Bench_KG_W12	1.00	Mean	54.3950	.83721	
		95% Confidence Interval for Mean	Lower Bound	52.6875		
			Upper Bound	56.1024		
		5% Trimmed Mean	54.3023		
		Median	55.1305		
		Variance	22.429		
		Std. Deviation	4.73596		
		Minimum	46.47		
		Maximum	64.58		
		Range	18.11		
		Interquartile Range	7.48		
		Skewness	.129	.414	
		Kurtosis	-.738	.809	
	2.00	Mean	51.0626	.82595	
		95% Confidence Interval for Mean	Lower Bound	49.3781		
			Upper Bound	52.7472		
		5% Trimmed Mean	50.9835		
		Median	49.2469		
		Variance	21.830		
		Std. Deviation	4.67229		
		Minimum	44.29		
		Maximum	59.07		
		Range	14.77		
		Interquartile Range	9.55		
		Skewness	.348	.414	
		Kurtosis	-1.427	.809	
Bench_KG_Diff_W6	1.00	Mean	-2.6749	.99951	
		95% Confidence Interval for Mean	Lower Bound	-4.7134		
			Upper Bound	-.6364		
		5% Trimmed Mean	-2.7215		
		Median	-1.6074		
		Variance	31.969		
		Std. Deviation	5.65407		
		Minimum	-13.42		
		Maximum	8.40		
		Range	21.82		
		Interquartile Range	6.39		
		Skewness	.177	.414	
		Kurtosis	-.413	.809	
	2.00	Mean	-1.0779	.75035	
		95% Confidence Interval for Mean	Lower Bound	-2.6082		
			Upper Bound	.4525		
		5% Trimmed Mean	-1.0326		
		Median	-1.4388		
		Variance	18.017		
		Std. Deviation	4.24464		
		Minimum	-9.79		
		Maximum	6.85		
		Range	16.64		
		Interquartile Range	1.67		
		Skewness	.101	.414	
		Kurtosis	.045	.809	
Bench_KG_Diff_W12	1.00	Mean	-5.0075	.93122	
		95% Confidence Interval for Mean	Lower Bound	-6.9067		
			Upper Bound	-3.1082		
		5% Trimmed Mean	-5.1043		
		Median	-3.4312		
		Variance	27.750		
		Std. Deviation	5.26778		
		Minimum	-13.50		
		Maximum	6.08		
		Range	19.58		
		Interquartile Range	8.34		
		Skewness	.104	.414	
		Kurtosis	-.613	.809	
	2.00	Mean	-1.3523	1.03166	
		95% Confidence Interval for Mean	Lower Bound	-3.4564		
			Upper Bound	.7518		
		5% Trimmed Mean	-1.3820		
		Median	-1.6973		
		Variance	34.058		
		Std. Deviation	5.83595		
		Minimum	-14.20		
		Maximum	13.62		
		Range	27.81		
		Interquartile Range	6.80		
		Skewness	.088	.414	
		Kurtosis	.581	.809	
Bench_Reps_B	1.00	Mean	11.9688	.15865	
		95% Confidence Interval for Mean	Lower Bound	11.6452		
			Upper Bound	12.2923		
		5% Trimmed Mean	11.9861		
		Median	12.0000		
		Variance	.805		
		Std. Deviation	.89747		
		Minimum	10.00		
		Maximum	14.00		
		Range	4.00		
		Interquartile Range	.00		
		Skewness	-.507	.414	
		Kurtosis	1.003	.809	
	2.00	Mean	11.7500	.14892	
		95% Confidence Interval for Mean	Lower Bound	11.4463		
			Upper Bound	12.0537		
		5% Trimmed Mean	11.7778		
		Median	12.0000		
		Variance	.710		
		Std. Deviation	.84242		
		Minimum	10.00		
		Maximum	13.00		
		Range	3.00		
		Interquartile Range	1.00		
		Skewness	-.173	.414	
		Kurtosis	-.466	.809	
Bench_Reps_W6	1.00	Mean	13.5000	.18513	
		95% Confidence Interval for Mean	Lower Bound	13.1224		
			Upper Bound	13.8776		
		5% Trimmed Mean	13.5000		
		Median	13.0000		
		Variance	1.097		
		Std. Deviation	1.04727		
		Minimum	11.00		
		Maximum	16.00		
		Range	5.00		
		Interquartile Range	1.00		
		Skewness	.090	.414	
		Kurtosis	.429	.809	
	2.00	Mean	12.5625	.16150	
		95% Confidence Interval for Mean	Lower Bound	12.2331		
			Upper Bound	12.8919		
		5% Trimmed Mean	12.6042		
		Median	13.0000		
		Variance	.835		
		Std. Deviation	.91361		
		Minimum	10.00		
		Maximum	14.00		
		Range	4.00		
		Interquartile Range	1.00		
		Skewness	-.603	.414	
		Kurtosis	.830	.809	
Bench_Reps_W12	1.00	Mean	15.1875	.27104	
		95% Confidence Interval for Mean	Lower Bound	14.6347		
			Upper Bound	15.7403		
		5% Trimmed Mean	15.1528		
		Median	15.0000		
		Variance	2.351		
		Std. Deviation	1.53323		
		Minimum	13.00		
		Maximum	18.00		
		Range	5.00		
		Interquartile Range	2.00		
		Skewness	.294	.414	
		Kurtosis	-.767	.809	
	2.00	Mean	12.7500	.16801	
		95% Confidence Interval for Mean	Lower Bound	12.4074		
			Upper Bound	13.0926		
		5% Trimmed Mean	12.7431		
		Median	13.0000		
		Variance	.903		
		Std. Deviation	.95038		
		Minimum	11.00		
		Maximum	15.00		
		Range	4.00		
		Interquartile Range	1.00		
		Skewness	.060	.414	
		Kurtosis	-.008	.809	
Bench_Reps_Diff_W6	1.00	Mean	-1.5313	.11867	
		95% Confidence Interval for Mean	Lower Bound	-1.7733		
			Upper Bound	-1.2892		
		5% Trimmed Mean	-1.4792		
		Median	-1.0000		
		Variance	.451		
		Std. Deviation	.67127		
		Minimum	-3.00		
		Maximum	-1.00		
		Range	2.00		
		Interquartile Range	1.00		
		Skewness	-.903	.414	
		Kurtosis	-.243	.809	
	2.00	Mean	-.8125	.08325	
		95% Confidence Interval for Mean	Lower Bound	-.9823		
			Upper Bound	-.6427		
		5% Trimmed Mean	-.8819		
		Median	-1.0000		
		Variance	.222		
		Std. Deviation	.47093		
		Minimum	-1.00		
		Maximum	1.00		
		Range	2.00		
		Interquartile Range	.00		
		Skewness	2.610	.414	
		Kurtosis	6.692	.809	
Bench_Reps_Diff_W12	1.00	Mean	-3.2188	.21875	
		95% Confidence Interval for Mean	Lower Bound	-3.6649		
			Upper Bound	-2.7726		
		5% Trimmed Mean	-3.1319		
		Median	-3.0000		
		Variance	1.531		
		Std. Deviation	1.23744		
		Minimum	-6.00		
		Maximum	-2.00		
		Range	4.00		
		Interquartile Range	2.00		
		Skewness	-1.081	.414	
		Kurtosis	.414	.809	
	2.00	Mean	-1.0000	.12700	
		95% Confidence Interval for Mean	Lower Bound	-1.2590		
			Upper Bound	-.7410		
		5% Trimmed Mean	-1.0347		
		Median	-1.0000		
		Variance	.516		
		Std. Deviation	.71842		
		Minimum	-2.00		
		Maximum	1.00		
		Range	3.00		
		Interquartile Range	.00		
		Skewness	.557	.414	
		Kurtosis	.802	.809	
Leg_KG_B	1.00	Mean	63.0002	.90319	
		95% Confidence Interval for Mean	Lower Bound	61.1582		
			Upper Bound	64.8423		
		5% Trimmed Mean	62.9966		
		Median	63.0030		
		Variance	26.104		
		Std. Deviation	5.10919		
		Minimum	52.63		
		Maximum	72.64		
		Range	20.01		
		Interquartile Range	7.04		
		Skewness	.003	.414	
		Kurtosis	-.271	.809	
	2.00	Mean	62.5496	.87602	
		95% Confidence Interval for Mean	Lower Bound	60.7629		
			Upper Bound	64.3362		
		5% Trimmed Mean	62.5919		
		Median	63.0030		
		Variance	24.557		
		Std. Deviation	4.95551		
		Minimum	51.88		
		Maximum	71.90		
		Range	20.01		
		Interquartile Range	7.78		
		Skewness	-.161	.414	
		Kurtosis	-.689	.809	
Leg_KG_W6	1.00	Mean	65.4591	.84406	
		95% Confidence Interval for Mean	Lower Bound	63.7376		
			Upper Bound	67.1806		
		5% Trimmed Mean	65.4826		
		Median	66.0593		
		Variance	22.798		
		Std. Deviation	4.77473		
		Minimum	56.51		
		Maximum	74.84		
		Range	18.33		
		Interquartile Range	6.77		
		Skewness	-.258	.414	
		Kurtosis	-.546	.809	
	2.00	Mean	64.0278	.99099	
		95% Confidence Interval for Mean	Lower Bound	62.0066		
			Upper Bound	66.0489		
		5% Trimmed Mean	64.1295		
		Median	64.6996		
		Variance	31.426		
		Std. Deviation	5.60586		
		Minimum	51.88		
		Maximum	74.08		
		Range	22.19		
		Interquartile Range	7.65		
		Skewness	-.285	.414	
		Kurtosis	-.593	.809	
Leg_KG_W12	1.00	Mean	67.9900	.77041	
		95% Confidence Interval for Mean	Lower Bound	66.4188		
			Upper Bound	69.5613		
		5% Trimmed Mean	68.1828		
		Median	68.5194		
		Variance	18.993		
		Std. Deviation	4.35810		
		Minimum	56.71		
		Maximum	76.40		
		Range	19.69		
		Interquartile Range	4.53		
		Skewness	-.944	.414	
		Kurtosis	1.134	.809	
	2.00	Mean	64.5845	.92288	
		95% Confidence Interval for Mean	Lower Bound	62.7022		
			Upper Bound	66.4667		
		5% Trimmed Mean	64.5730		
		Median	65.2266		
		Variance	27.255		
		Std. Deviation	5.22060		
		Minimum	54.22		
		Maximum	76.40		
		Range	22.17		
		Interquartile Range	6.87		
		Skewness	-.169	.414	
		Kurtosis	-.091	.809	
Leg_KG_Diff_W6	1.00	Mean	-2.4589	.59763	
		95% Confidence Interval for Mean	Lower Bound	-3.6778		
			Upper Bound	-1.2400		
		5% Trimmed Mean	-2.5179		
		Median	-2.5621		
		Variance	11.429		
		Std. Deviation	3.38068		
		Minimum	-10.29		
		Maximum	6.40		
		Range	16.69		
		Interquartile Range	2.58		
		Skewness	.377	.414	
		Kurtosis	1.876	.809	
	2.00	Mean	-1.4782	.43925	
		95% Confidence Interval for Mean	Lower Bound	-2.3740		
			Upper Bound	-.5823		
		5% Trimmed Mean	-1.5058		
		Median	-1.7760		
		Variance	6.174		
		Std. Deviation	2.48480		
		Minimum	-7.32		
		Maximum	5.53		
		Range	12.86		
		Interquartile Range	2.53		
		Skewness	.175	.414	
		Kurtosis	1.535	.809	
Leg_KG_Diff_W12	1.00	Mean	-4.9898	.69016	
		95% Confidence Interval for Mean	Lower Bound	-6.3974		
			Upper Bound	-3.5822		
		5% Trimmed Mean	-5.1021		
		Median	-4.9606		
		Variance	15.242		
		Std. Deviation	3.90416		
		Minimum	-12.93		
		Maximum	4.54		
		Range	17.47		
		Interquartile Range	3.47		
		Skewness	.836	.414	
		Kurtosis	1.225	.809	
	2.00	Mean	-2.0349	.47634	
		95% Confidence Interval for Mean	Lower Bound	-3.0064		
			Upper Bound	-1.0634		
		5% Trimmed Mean	-2.1823		
		Median	-2.4722		
		Variance	7.261		
		Std. Deviation	2.69458		
		Minimum	-6.31		
		Maximum	4.81		
		Range	11.12		
		Interquartile Range	4.05		
		Skewness	.863	.414	
		Kurtosis	.555	.809	
Leg_Reps_B	1.00	Mean	13.3438	.18842	
		95% Confidence Interval for Mean	Lower Bound	12.9595		
			Upper Bound	13.7280		
		5% Trimmed Mean	13.2708		
		Median	13.0000		
		Variance	1.136		
		Std. Deviation	1.06587		
		Minimum	12.00		
		Maximum	16.00		
		Range	4.00		
		Interquartile Range	1.00		
		Skewness	.949	.414	
		Kurtosis	.802	.809	
	2.00	Mean	12.7813	.13270	
		95% Confidence Interval for Mean	Lower Bound	12.5106		
			Upper Bound	13.0519		
		5% Trimmed Mean	12.8125		
		Median	13.0000		
		Variance	.564		
		Std. Deviation	.75067		
		Minimum	11.00		
		Maximum	14.00		
		Range	3.00		
		Interquartile Range	1.00		
		Skewness	-.586	.414	
		Kurtosis	.584	.809	
Leg_Reps_W6	1.00	Mean	15.8750	.20941	
		95% Confidence Interval for Mean	Lower Bound	15.4479		
			Upper Bound	16.3021		
		5% Trimmed Mean	15.8611		
		Median	16.0000		
		Variance	1.403		
		Std. Deviation	1.18458		
		Minimum	14.00		
		Maximum	18.00		
		Range	4.00		
		Interquartile Range	2.00		
		Skewness	.008	.414	
		Kurtosis	-.607	.809	
	2.00	Mean	13.6563	.14625	
		95% Confidence Interval for Mean	Lower Bound	13.3580		
			Upper Bound	13.9545		
		5% Trimmed Mean	13.6736		
		Median	14.0000		
		Variance	.684		
		Std. Deviation	.82733		
		Minimum	12.00		
		Maximum	15.00		
		Range	3.00		
		Interquartile Range	1.00		
		Skewness	-.718	.414	
		Kurtosis	.134	.809	
Leg_Reps_W12	1.00	Mean	17.1250	.29699	
		95% Confidence Interval for Mean	Lower Bound	16.5193		
			Upper Bound	17.7307		
		5% Trimmed Mean	17.1389		
		Median	18.0000		
		Variance	2.823		
		Std. Deviation	1.68005		
		Minimum	14.00		
		Maximum	20.00		
		Range	6.00		
		Interquartile Range	2.00		
		Skewness	-.253	.414	
		Kurtosis	-.890	.809	
	2.00	Mean	14.0625	.16150	
		95% Confidence Interval for Mean	Lower Bound	13.7331		
			Upper Bound	14.3919		
		5% Trimmed Mean	14.0903		
		Median	14.0000		
		Variance	.835		
		Std. Deviation	.91361		
		Minimum	12.00		
		Maximum	16.00		
		Range	4.00		
		Interquartile Range	1.00		
		Skewness	-.400	.414	
		Kurtosis	.261	.809	
Leg_Reps_Diff_W6	1.00	Mean	-2.5313	.17952	
		95% Confidence Interval for Mean	Lower Bound	-2.8974		
			Upper Bound	-2.1651		
		5% Trimmed Mean	-2.4444		
		Median	-2.0000		
		Variance	1.031		
		Std. Deviation	1.01550		
		Minimum	-6.00		
		Maximum	-1.00		
		Range	5.00		
		Interquartile Range	1.00		
		Skewness	-1.684	.414	
		Kurtosis	4.028	.809	
	2.00	Mean	-.8750	.09786	
		95% Confidence Interval for Mean	Lower Bound	-1.0746		
			Upper Bound	-.6754		
		5% Trimmed Mean	-.8958		
		Median	-1.0000		
		Variance	.306		
		Std. Deviation	.55358		
		Minimum	-2.00		
		Maximum	1.00		
		Range	3.00		
		Interquartile Range	.00		
		Skewness	1.293	.414	
		Kurtosis	4.155	.809	
Leg_Reps_Diff_W12	1.00	Mean	-3.7813	.31982	
		95% Confidence Interval for Mean	Lower Bound	-4.4335		
			Upper Bound	-3.1290		
		5% Trimmed Mean	-3.7917		
		Median	-4.0000		
		Variance	3.273		
		Std. Deviation	1.80919		
		Minimum	-7.00		
		Maximum	.00		
		Range	7.00		
		Interquartile Range	2.75		
		Skewness	.000	.414	
		Kurtosis	-.676	.809	
	2.00	Mean	-1.2813	.12077	
		95% Confidence Interval for Mean	Lower Bound	-1.5276		
			Upper Bound	-1.0349		
		5% Trimmed Mean	-1.3125		
		Median	-1.0000		
		Variance	.467		
		Std. Deviation	.68318		
		Minimum	-2.00		
		Maximum	.00		
		Range	2.00		
		Interquartile Range	1.00		
		Skewness	.423	.414	
		Kurtosis	-.736	.809	
Fatigue_Index_Leg_B	1.00	Mean	23.4159	1.01818	
		95% Confidence Interval for Mean	Lower Bound	21.3393		
			Upper Bound	25.4925		
		5% Trimmed Mean	23.3974		
		Median	24.0385		
		Variance	33.174		
		Std. Deviation	5.75970		
		Minimum	9.09		
		Maximum	35.71		
		Range	26.62		
		Interquartile Range	6.82		
		Skewness	.089	.414	
		Kurtosis	.716	.809	
	2.00	Mean	23.3836	.79423	
		95% Confidence Interval for Mean	Lower Bound	21.7637		
			Upper Bound	25.0034		
		5% Trimmed Mean	23.2040		
		Median	25.0000		
		Variance	20.186		
		Std. Deviation	4.49287		
		Minimum	16.67		
		Maximum	33.33		
		Range	16.67		
		Interquartile Range	6.82		
		Skewness	.183	.414	
		Kurtosis	-.278	.809	
Fatigue_Index_Leg_W6	1.00	Mean	21.0409	.98531	
		95% Confidence Interval for Mean	Lower Bound	19.0314		
			Upper Bound	23.0505		
		5% Trimmed Mean	20.9927		
		Median	23.0769		
		Variance	31.067		
		Std. Deviation	5.57378		
		Minimum	9.09		
		Maximum	35.71		
		Range	26.62		
		Interquartile Range	6.41		
		Skewness	.194	.414	
		Kurtosis	1.125	.809	
	2.00	Mean	22.8212	.97753	
		95% Confidence Interval for Mean	Lower Bound	20.8275		
			Upper Bound	24.8149		
		5% Trimmed Mean	22.3731		
		Median	23.0769		
		Variance	30.578		
		Std. Deviation	5.52973		
		Minimum	15.38		
		Maximum	42.86		
		Range	27.47		
		Interquartile Range	6.82		
		Skewness	1.534	.414	
		Kurtosis	4.496	.809	
Fatigue_Index_Leg_W12	1.00	Mean	16.9345	.72936	
		95% Confidence Interval for Mean	Lower Bound	15.4470		
			Upper Bound	18.4221		
		5% Trimmed Mean	17.0164		
		Median	15.3846		
		Variance	17.023		
		Std. Deviation	4.12589		
		Minimum	7.69		
		Maximum	26.67		
		Range	18.97		
		Interquartile Range	6.87		
		Skewness	.016	.414	
		Kurtosis	.427	.809	
	2.00	Mean	22.0770	1.19673	
		95% Confidence Interval for Mean	Lower Bound	19.6363		
			Upper Bound	24.5177		
		5% Trimmed Mean	21.7540		
		Median	21.4286		
		Variance	45.829		
		Std. Deviation	6.76971		
		Minimum	9.09		
		Maximum	42.86		
		Range	33.77		
		Interquartile Range	9.66		
		Skewness	.952	.414	
		Kurtosis	1.737	.809	
Fatigue_Index_Leg_Diff_W6	1.00	Mean	2.3750	1.37653	
		95% Confidence Interval for Mean	Lower Bound	-.4325		
			Upper Bound	5.1824		
		5% Trimmed Mean	2.2041		
		Median	1.9231		
		Variance	60.635		
		Std. Deviation	7.78684		
		Minimum	-14.10		
		Maximum	26.62		
		Range	40.73		
		Interquartile Range	4.48		
		Skewness	.481	.414	
		Kurtosis	2.399	.809	
	2.00	Mean	.5624	1.13066	
		95% Confidence Interval for Mean	Lower Bound	-1.7436		
			Upper Bound	2.8684		
		5% Trimmed Mean	.9263		
		Median	1.7333		
		Variance	40.908		
		Std. Deviation	6.39596		
		Minimum	-22.86		
		Maximum	11.90		
		Range	34.76		
		Interquartile Range	2.19		
		Skewness	-1.373	.414	
		Kurtosis	5.009	.809	
Fatigue_Index_Leg_Diff_W12	1.00	Mean	6.4814	1.15005	
		95% Confidence Interval for Mean	Lower Bound	4.1358		
			Upper Bound	8.8269		
		5% Trimmed Mean	6.0617		
		Median	4.4322		
		Variance	42.324		
		Std. Deviation	6.50566		
		Minimum	-5.19		
		Maximum	28.02		
		Range	33.22		
		Interquartile Range	6.82		
		Skewness	1.277	.414	
		Kurtosis	3.415	.809	
	2.00	Mean	1.3066	1.35788	
		95% Confidence Interval for Mean	Lower Bound	-1.4628		
			Upper Bound	4.0760		
		5% Trimmed Mean	1.3286		
		Median	3.1843		
		Variance	59.003		
		Std. Deviation	7.68135		
		Minimum	-15.58		
		Maximum	20.00		
		Range	35.58		
		Interquartile Range	8.51		
		Skewness	-.256	.414	
		Kurtosis	.554	.809	
Fatigue_Index_Bench_B	1.00	Mean	31.8355	.97776	
		95% Confidence Interval for Mean	Lower Bound	29.8414		
			Upper Bound	33.8297		
		5% Trimmed Mean	31.6340		
		Median	30.0000		
		Variance	30.593		
		Std. Deviation	5.53107		
		Minimum	22.22		
		Maximum	45.45		
		Range	23.23		
		Interquartile Range	8.33		
		Skewness	.650	.414	
		Kurtosis	-.095	.809	
	2.00	Mean	31.3905	1.11254	
		95% Confidence Interval for Mean	Lower Bound	29.1214		
			Upper Bound	33.6595		
		5% Trimmed Mean	31.3208		
		Median	30.0000		
		Variance	39.608		
		Std. Deviation	6.29346		
		Minimum	20.00		
		Maximum	45.45		
		Range	25.45		
		Interquartile Range	9.19		
		Skewness	.298	.414	
		Kurtosis	-.275	.809	
Fatigue_Index_Bench_W6	1.00	Mean	29.2561	1.02203	
		95% Confidence Interval for Mean	Lower Bound	27.1717		
			Upper Bound	31.3406		
		5% Trimmed Mean	29.2545		
		Median	30.0000		
		Variance	33.425		
		Std. Deviation	5.78146		
		Minimum	18.18		
		Maximum	41.67		
		Range	23.48		
		Interquartile Range	7.77		
		Skewness	-.232	.414	
		Kurtosis	-.398	.809	
	2.00	Mean	30.2375	1.20945	
		95% Confidence Interval for Mean	Lower Bound	27.7708		
			Upper Bound	32.7042		
		5% Trimmed Mean	30.2723		
		Median	30.0000		
		Variance	46.809		
		Std. Deviation	6.84169		
		Minimum	18.18		
		Maximum	41.67		
		Range	23.48		
		Interquartile Range	11.36		
		Skewness	.010	.414	
		Kurtosis	-.806	.809	
Fatigue_Index_Bench_W12	1.00	Mean	22.6764	1.18797	
		95% Confidence Interval for Mean	Lower Bound	20.2536		
			Upper Bound	25.0993		
		5% Trimmed Mean	22.8654		
		Median	24.0385		
		Variance	45.161		
		Std. Deviation	6.72017		
		Minimum	8.33		
		Maximum	33.33		
		Range	25.00		
		Interquartile Range	9.09		
		Skewness	-.435	.414	
		Kurtosis	-.356	.809	
	2.00	Mean	29.2680	1.40069	
		95% Confidence Interval for Mean	Lower Bound	26.4113		
			Upper Bound	32.1248		
		5% Trimmed Mean	29.2477		
		Median	30.0000		
		Variance	62.782		
		Std. Deviation	7.92350		
		Minimum	16.67		
		Maximum	41.67		
		Range	25.00		
		Interquartile Range	14.14		
		Skewness	-.034	.414	
		Kurtosis	-1.258	.809	
Fatigue_Index_Bench_Diff_W6	1.00	Mean	2.5794	.89131	
		95% Confidence Interval for Mean	Lower Bound	.7616		
			Upper Bound	4.3972		
		5% Trimmed Mean	2.6505		
		Median	3.1818		
		Variance	25.422		
		Std. Deviation	5.04203		
		Minimum	-8.33		
		Maximum	11.82		
		Range	20.15		
		Interquartile Range	1.48		
		Skewness	-.404	.414	
		Kurtosis	.269	.809	
	2.00	Mean	1.1529	.96270	
		95% Confidence Interval for Mean	Lower Bound	-.8105		
			Upper Bound	3.1164		
		5% Trimmed Mean	1.2016		
		Median	.0000		
		Variance	29.658		
		Std. Deviation	5.44587		
		Minimum	-16.36		
		Maximum	15.15		
		Range	31.52		
		Interquartile Range	3.41		
		Skewness	-.227	.414	
		Kurtosis	3.947	.809	
Fatigue_Index_Bench_Diff_W12	1.00	Mean	9.1591	1.33749	
		95% Confidence Interval for Mean	Lower Bound	6.4313		
			Upper Bound	11.8869		
		5% Trimmed Mean	9.1095		
		Median	11.2121		
		Variance	57.244		
		Std. Deviation	7.56601		
		Minimum	-3.50		
		Maximum	23.33		
		Range	26.83		
		Interquartile Range	9.92		
		Skewness	-.205	.414	
		Kurtosis	-.631	.809	
	2.00	Mean	2.1224	1.39003	
		95% Confidence Interval for Mean	Lower Bound	-.7126		
			Upper Bound	4.9574		
		5% Trimmed Mean	2.3375		
		Median	.0000		
		Variance	61.830		
		Std. Deviation	7.86319		
		Minimum	-16.36		
		Maximum	15.15		
		Range	31.52		
		Interquartile Range	11.48		
		Skewness	-.181	.414	
		Kurtosis	-.331	.809	
