## Supplementary material for "Effects of a standardised *Amaranthus tricolor*–*Spinacia oleracea* extract on exercise performance, muscle strength, endurance and recovery in physically active adults: a randomised, double-blind, placebo-controlled trial": Raw statistical outputs: Biomarkers_Output_17.07.2026.doc

Explore


Notes	
Output Created	17-JUL-2026 11:34:27	
Comments		
Input	Active Dataset	DataSet0	
	Filter	<none>	
	Weight	<none>	
	Split File	<none>	
	N of Rows in Working Data File	64	
Missing Value Handling	Definition of Missing	User-defined missing values for dependent variables are treated as missing.	
	Cases Used	Statistics are based on cases with no missing values for any dependent variable or factor used.	
Syntax	EXAMINE VARIABLES=Creat_Kinase_B Creat_Kinase_W12 Creat_Kinase_Diff_W12 Lact_Dehydr_B
    Lact_Dehydr_W12 Lact_Dehydr_Diff_W12 Blood_Lac_B Blood_Lac_W12 Blood_Lac_Diff_W12 Plasma_Nitr_B
    Plasma_Nitr_W12 Plasma_Nitr_Diff_W12 Serum_Creat_B Serum_Creat_W12 Serum_Creat_Diff_W12 Cortisol_B
    Cortisol_W12 Cortisol_Diff_W12 Serum_Ferritin_B Serum_Ferritin_W12 Serum_Ferritin_Diff_W12
    Serum_iron_B Serum_iron_W12 Serum_iron_Diff_W12 BY Group
  /PLOT BOXPLOT STEMLEAF
  /COMPARE GROUPS
  /STATISTICS DESCRIPTIVES
  /CINTERVAL 95
  /MISSING LISTWISE
  /NOTOTAL.	
Resources	Processor Time	00:00:02.14	
	Elapsed Time	00:00:01.92	


Group


Case Processing Summary	
	Group	Cases	
		Valid	Missing	Total	
		N	Percent	N	Percent	N	Percent	
Creat_Kinase_B	1.00	32	100.0%	0	0.0%	32	100.0%	
	2.00	32	100.0%	0	0.0%	32	100.0%	
Creat_Kinase_W12	1.00	32	100.0%	0	0.0%	32	100.0%	
	2.00	32	100.0%	0	0.0%	32	100.0%	
Creat_Kinase_Diff_W12	1.00	32	100.0%	0	0.0%	32	100.0%	
	2.00	32	100.0%	0	0.0%	32	100.0%	
Lact_Dehydr_B	1.00	32	100.0%	0	0.0%	32	100.0%	
	2.00	32	100.0%	0	0.0%	32	100.0%	
Lact_Dehydr_W12	1.00	32	100.0%	0	0.0%	32	100.0%	
	2.00	32	100.0%	0	0.0%	32	100.0%	
Lact_Dehydr_Diff_W12	1.00	32	100.0%	0	0.0%	32	100.0%	
	2.00	32	100.0%	0	0.0%	32	100.0%	
Blood_Lac_B	1.00	32	100.0%	0	0.0%	32	100.0%	
	2.00	32	100.0%	0	0.0%	32	100.0%	
Blood_Lac_W12	1.00	32	100.0%	0	0.0%	32	100.0%	
	2.00	32	100.0%	0	0.0%	32	100.0%	
Blood_Lac_Diff_W12	1.00	32	100.0%	0	0.0%	32	100.0%	
	2.00	32	100.0%	0	0.0%	32	100.0%	
Plasma_Nitr_B	1.00	32	100.0%	0	0.0%	32	100.0%	
	2.00	32	100.0%	0	0.0%	32	100.0%	
Plasma_Nitr_W12	1.00	32	100.0%	0	0.0%	32	100.0%	
	2.00	32	100.0%	0	0.0%	32	100.0%	
Plasma_Nitr_Diff_W12	1.00	32	100.0%	0	0.0%	32	100.0%	
	2.00	32	100.0%	0	0.0%	32	100.0%	
Serum_Creat_B	1.00	32	100.0%	0	0.0%	32	100.0%	
	2.00	32	100.0%	0	0.0%	32	100.0%	
Serum_Creat_W12	1.00	32	100.0%	0	0.0%	32	100.0%	
	2.00	32	100.0%	0	0.0%	32	100.0%	
Serum_Creat_Diff_W12	1.00	32	100.0%	0	0.0%	32	100.0%	
	2.00	32	100.0%	0	0.0%	32	100.0%	
Cortisol_B	1.00	32	100.0%	0	0.0%	32	100.0%	
	2.00	32	100.0%	0	0.0%	32	100.0%	
Cortisol_W12	1.00	32	100.0%	0	0.0%	32	100.0%	
	2.00	32	100.0%	0	0.0%	32	100.0%	
Cortisol_Diff_W12	1.00	32	100.0%	0	0.0%	32	100.0%	
	2.00	32	100.0%	0	0.0%	32	100.0%	
Serum_Ferritin_B	1.00	32	100.0%	0	0.0%	32	100.0%	
	2.00	32	100.0%	0	0.0%	32	100.0%	
Serum_Ferritin_W12	1.00	32	100.0%	0	0.0%	32	100.0%	
	2.00	32	100.0%	0	0.0%	32	100.0%	
Serum_Ferritin_Diff_W12	1.00	32	100.0%	0	0.0%	32	100.0%	
	2.00	32	100.0%	0	0.0%	32	100.0%	
Serum_iron_B	1.00	32	100.0%	0	0.0%	32	100.0%	
	2.00	32	100.0%	0	0.0%	32	100.0%	
Serum_iron_W12	1.00	32	100.0%	0	0.0%	32	100.0%	
	2.00	32	100.0%	0	0.0%	32	100.0%	
Serum_iron_Diff_W12	1.00	32	100.0%	0	0.0%	32	100.0%	
	2.00	32	100.0%	0	0.0%	32	100.0%	


Descriptives	
	Group	Statistic	Std. Error	
Creat_Kinase_B	1.00	Mean	122.5181	7.88158	
		95% Confidence Interval for Mean	Lower Bound	106.4435		
			Upper Bound	138.5927		
		5% Trimmed Mean	121.8391		
		Median	105.3100		
		Variance	1987.816		
		Std. Deviation	44.58493		
		Minimum	49.20		
		Maximum	210.59		
		Range	161.39		
		Interquartile Range	68.10		
		Skewness	.470	.414	
		Kurtosis	-.733	.809	
	2.00	Mean	118.0516	5.75930	
		95% Confidence Interval for Mean	Lower Bound	106.3054		
			Upper Bound	129.7977		
		5% Trimmed Mean	116.5887		
		Median	110.9250		
		Variance	1061.424		
		Std. Deviation	32.57951		
		Minimum	56.32		
		Maximum	198.00		
		Range	141.68		
		Interquartile Range	34.97		
		Skewness	.926	.414	
		Kurtosis	.656	.809	
Creat_Kinase_W12	1.00	Mean	104.8706	7.54253	
		95% Confidence Interval for Mean	Lower Bound	89.4875		
			Upper Bound	120.2537		
		5% Trimmed Mean	103.4718		
		Median	91.0950		
		Variance	1820.474		
		Std. Deviation	42.66702		
		Minimum	39.23		
		Maximum	199.89		
		Range	160.66		
		Interquartile Range	65.97		
		Skewness	.640	.414	
		Kurtosis	-.553	.809	
	2.00	Mean	111.0291	5.45277	
		95% Confidence Interval for Mean	Lower Bound	99.9081		
			Upper Bound	122.1501		
		5% Trimmed Mean	109.7752		
		Median	101.7050		
		Variance	951.447		
		Std. Deviation	30.84554		
		Minimum	51.93		
		Maximum	183.09		
		Range	131.16		
		Interquartile Range	27.68		
		Skewness	.963	.414	
		Kurtosis	.546	.809	
Creat_Kinase_Diff_W12	1.00	Mean	17.6475	2.02120	
		95% Confidence Interval for Mean	Lower Bound	13.5252		
			Upper Bound	21.7698		
		5% Trimmed Mean	16.8122		
		Median	13.4300		
		Variance	130.728		
		Std. Deviation	11.43362		
		Minimum	3.73		
		Maximum	48.90		
		Range	45.17		
		Interquartile Range	15.84		
		Skewness	1.163	.414	
		Kurtosis	.625	.809	
	2.00	Mean	7.0225	1.35345	
		95% Confidence Interval for Mean	Lower Bound	4.2621		
			Upper Bound	9.7829		
		5% Trimmed Mean	7.4347		
		Median	8.3550		
		Variance	58.619		
		Std. Deviation	7.65628		
		Minimum	-15.39		
		Maximum	19.92		
		Range	35.31		
		Interquartile Range	8.49		
		Skewness	-.908	.414	
		Kurtosis	1.327	.809	
Lact_Dehydr_B	1.00	Mean	236.4444	4.32321	
		95% Confidence Interval for Mean	Lower Bound	227.6271		
			Upper Bound	245.2616		
		5% Trimmed Mean	235.8833		
		Median	235.2500		
		Variance	598.086		
		Std. Deviation	24.45580		
		Minimum	181.70		
		Maximum	298.70		
		Range	117.00		
		Interquartile Range	19.97		
		Skewness	.644	.414	
		Kurtosis	1.203	.809	
	2.00	Mean	237.9303	9.00351	
		95% Confidence Interval for Mean	Lower Bound	219.5675		
			Upper Bound	256.2931		
		5% Trimmed Mean	236.9719		
		Median	238.2600		
		Variance	2594.019		
		Std. Deviation	50.93152		
		Minimum	115.80		
		Maximum	409.00		
		Range	293.20		
		Interquartile Range	16.92		
		Skewness	.492	.414	
		Kurtosis	4.545	.809	
Lact_Dehydr_W12	1.00	Mean	203.6878	3.83188	
		95% Confidence Interval for Mean	Lower Bound	195.8727		
			Upper Bound	211.5030		
		5% Trimmed Mean	204.4191		
		Median	200.2700		
		Variance	469.865		
		Std. Deviation	21.67636		
		Minimum	135.10		
		Maximum	243.10		
		Range	108.00		
		Interquartile Range	23.95		
		Skewness	-.489	.414	
		Kurtosis	2.166	.809	
	2.00	Mean	218.3431	9.44838	
		95% Confidence Interval for Mean	Lower Bound	199.0730		
			Upper Bound	237.6132		
		5% Trimmed Mean	217.3549		
		Median	226.6000		
		Variance	2856.703		
		Std. Deviation	53.44814		
		Minimum	102.30		
		Maximum	394.10		
		Range	291.80		
		Interquartile Range	39.53		
		Skewness	.412	.414	
		Kurtosis	3.503	.809	
Lact_Dehydr_Diff_W12	1.00	Mean	32.7566	3.30210	
		95% Confidence Interval for Mean	Lower Bound	26.0219		
			Upper Bound	39.4912		
		5% Trimmed Mean	32.6762		
		Median	29.5000		
		Variance	348.924		
		Std. Deviation	18.67952		
		Minimum	-3.70		
		Maximum	72.20		
		Range	75.90		
		Interquartile Range	19.69		
		Skewness	.366	.414	
		Kurtosis	.129	.809	
	2.00	Mean	19.5872	4.64720	
		95% Confidence Interval for Mean	Lower Bound	10.1092		
			Upper Bound	29.0652		
		5% Trimmed Mean	17.4420		
		Median	11.8000		
		Variance	691.088		
		Std. Deviation	26.28855		
		Minimum	-19.60		
		Maximum	96.30		
		Range	115.90		
		Interquartile Range	18.03		
		Skewness	1.616	.414	
		Kurtosis	2.445	.809	
Blood_Lac_B	1.00	Mean	68.7647	2.21646	
		95% Confidence Interval for Mean	Lower Bound	64.2442		
			Upper Bound	73.2852		
		5% Trimmed Mean	68.6556		
		Median	65.8900		
		Variance	157.206		
		Std. Deviation	12.53819		
		Minimum	44.03		
		Maximum	96.97		
		Range	52.94		
		Interquartile Range	20.46		
		Skewness	.271	.414	
		Kurtosis	-.455	.809	
	2.00	Mean	72.8669	2.83179	
		95% Confidence Interval for Mean	Lower Bound	67.0914		
			Upper Bound	78.6423		
		5% Trimmed Mean	72.8874		
		Median	74.3900		
		Variance	256.608		
		Std. Deviation	16.01900		
		Minimum	41.56		
		Maximum	105.60		
		Range	64.04		
		Interquartile Range	19.67		
		Skewness	.017	.414	
		Kurtosis	-.314	.809	
Blood_Lac_W12	1.00	Mean	40.0591	1.53569	
		95% Confidence Interval for Mean	Lower Bound	36.9270		
			Upper Bound	43.1911		
		5% Trimmed Mean	39.9596		
		Median	40.2300		
		Variance	75.467		
		Std. Deviation	8.68717		
		Minimum	23.68		
		Maximum	60.76		
		Range	37.08		
		Interquartile Range	12.25		
		Skewness	.116	.414	
		Kurtosis	-.159	.809	
	2.00	Mean	71.3169	2.99515	
		95% Confidence Interval for Mean	Lower Bound	65.2082		
			Upper Bound	77.4255		
		5% Trimmed Mean	70.4681		
		Median	69.9750		
		Variance	287.069		
		Std. Deviation	16.94312		
		Minimum	40.69		
		Maximum	123.06		
		Range	82.37		
		Interquartile Range	22.32		
		Skewness	.809	.414	
		Kurtosis	1.448	.809	
Blood_Lac_Diff_W12	1.00	Mean	28.7056	2.69604	
		95% Confidence Interval for Mean	Lower Bound	23.2070		
			Upper Bound	34.2042		
		5% Trimmed Mean	28.8275		
		Median	27.0450		
		Variance	232.596		
		Std. Deviation	15.25109		
		Minimum	-.91		
		Maximum	53.03		
		Range	53.94		
		Interquartile Range	24.85		
		Skewness	.203	.414	
		Kurtosis	-1.037	.809	
	2.00	Mean	1.5500	1.61594	
		95% Confidence Interval for Mean	Lower Bound	-1.7457		
			Upper Bound	4.8457		
		5% Trimmed Mean	1.7884		
		Median	2.8500		
		Variance	83.560		
		Std. Deviation	9.14113		
		Minimum	-17.46		
		Maximum	16.01		
		Range	33.47		
		Interquartile Range	14.99		
		Skewness	-.408	.414	
		Kurtosis	-.874	.809	
Plasma_Nitr_B	1.00	Mean	15.6763	.35819	
		95% Confidence Interval for Mean	Lower Bound	14.9457		
			Upper Bound	16.4068		
		5% Trimmed Mean	15.6652		
		Median	15.5200		
		Variance	4.106		
		Std. Deviation	2.02622		
		Minimum	11.06		
		Maximum	20.09		
		Range	9.03		
		Interquartile Range	3.16		
		Skewness	.069	.414	
		Kurtosis	-.198	.809	
	2.00	Mean	13.8487	.42909	
		95% Confidence Interval for Mean	Lower Bound	12.9736		
			Upper Bound	14.7239		
		5% Trimmed Mean	13.8145		
		Median	14.0200		
		Variance	5.892		
		Std. Deviation	2.42730		
		Minimum	9.86		
		Maximum	18.45		
		Range	8.59		
		Interquartile Range	3.39		
		Skewness	-.019	.414	
		Kurtosis	-.637	.809	
Plasma_Nitr_W12	1.00	Mean	22.3619	.65129	
		95% Confidence Interval for Mean	Lower Bound	21.0336		
			Upper Bound	23.6902		
		5% Trimmed Mean	22.1901		
		Median	21.3300		
		Variance	13.574		
		Std. Deviation	3.68426		
		Minimum	17.14		
		Maximum	31.29		
		Range	14.15		
		Interquartile Range	4.80		
		Skewness	.889	.414	
		Kurtosis	.014	.809	
	2.00	Mean	14.6881	.44441	
		95% Confidence Interval for Mean	Lower Bound	13.7817		
			Upper Bound	15.5945		
		5% Trimmed Mean	14.6947		
		Median	15.0450		
		Variance	6.320		
		Std. Deviation	2.51397		
		Minimum	10.11		
		Maximum	19.26		
		Range	9.15		
		Interquartile Range	3.28		
		Skewness	-.207	.414	
		Kurtosis	-.527	.809	
Plasma_Nitr_Diff_W12	1.00	Mean	-6.6856	.61383	
		95% Confidence Interval for Mean	Lower Bound	-7.9375		
			Upper Bound	-5.4337		
		5% Trimmed Mean	-6.6934		
		Median	-6.6850		
		Variance	12.057		
		Std. Deviation	3.47233		
		Minimum	-13.45		
		Maximum	1.70		
		Range	15.15		
		Interquartile Range	4.33		
		Skewness	-.165	.414	
		Kurtosis	.281	.809	
	2.00	Mean	-.8394	.18300	
		95% Confidence Interval for Mean	Lower Bound	-1.2126		
			Upper Bound	-.4661		
		5% Trimmed Mean	-.8488		
		Median	-.8750		
		Variance	1.072		
		Std. Deviation	1.03521		
		Minimum	-4.19		
		Maximum	2.49		
		Range	6.68		
		Interquartile Range	1.02		
		Skewness	.038	.414	
		Kurtosis	5.552	.809	
Serum_Creat_B	1.00	Mean	4.1163	.08903	
		95% Confidence Interval for Mean	Lower Bound	3.9347		
			Upper Bound	4.2978		
		5% Trimmed Mean	4.1313		
		Median	4.1950		
		Variance	.254		
		Std. Deviation	.50363		
		Minimum	3.12		
		Maximum	4.83		
		Range	1.71		
		Interquartile Range	.85		
		Skewness	-.259	.414	
		Kurtosis	-.859	.809	
	2.00	Mean	3.8800	.10925	
		95% Confidence Interval for Mean	Lower Bound	3.6572		
			Upper Bound	4.1028		
		5% Trimmed Mean	3.9026		
		Median	3.8100		
		Variance	.382		
		Std. Deviation	.61801		
		Minimum	2.20		
		Maximum	4.82		
		Range	2.62		
		Interquartile Range	.87		
		Skewness	-.323	.414	
		Kurtosis	.083	.809	
Serum_Creat_W12	1.00	Mean	5.8353	.16769	
		95% Confidence Interval for Mean	Lower Bound	5.4933		
			Upper Bound	6.1773		
		5% Trimmed Mean	5.8094		
		Median	5.8000		
		Variance	.900		
		Std. Deviation	.94858		
		Minimum	4.10		
		Maximum	7.95		
		Range	3.85		
		Interquartile Range	1.26		
		Skewness	.554	.414	
		Kurtosis	.137	.809	
	2.00	Mean	4.1563	.13654	
		95% Confidence Interval for Mean	Lower Bound	3.8778		
			Upper Bound	4.4347		
		5% Trimmed Mean	4.1798		
		Median	4.2050		
		Variance	.597		
		Std. Deviation	.77239		
		Minimum	2.38		
		Maximum	5.43		
		Range	3.05		
		Interquartile Range	1.39		
		Skewness	-.431	.414	
		Kurtosis	-.336	.809	
Serum_Creat_Diff_W12	1.00	Mean	-1.7191	.15958	
		95% Confidence Interval for Mean	Lower Bound	-2.0445		
			Upper Bound	-1.3936		
		5% Trimmed Mean	-1.6408		
		Median	-1.4900		
		Variance	.815		
		Std. Deviation	.90270		
		Minimum	-4.55		
		Maximum	-.54		
		Range	4.01		
		Interquartile Range	.75		
		Skewness	-1.568	.414	
		Kurtosis	2.619	.809	
	2.00	Mean	-.2762	.07736	
		95% Confidence Interval for Mean	Lower Bound	-.4340		
			Upper Bound	-.1185		
		5% Trimmed Mean	-.3158		
		Median	-.3150		
		Variance	.191		
		Std. Deviation	.43759		
		Minimum	-.82		
		Maximum	1.07		
		Range	1.89		
		Interquartile Range	.39		
		Skewness	1.667	.414	
		Kurtosis	3.194	.809	
Cortisol_B	1.00	Mean	145.9663	9.42157	
		95% Confidence Interval for Mean	Lower Bound	126.7508		
			Upper Bound	165.1817		
		5% Trimmed Mean	141.7629		
		Median	120.9950		
		Variance	2840.514		
		Std. Deviation	53.29647		
		Minimum	87.00		
		Maximum	289.63		
		Range	202.63		
		Interquartile Range	78.20		
		Skewness	1.023	.414	
		Kurtosis	.367	.809	
	2.00	Mean	144.0147	8.60955	
		95% Confidence Interval for Mean	Lower Bound	126.4554		
			Upper Bound	161.5740		
		5% Trimmed Mean	142.0564		
		Median	120.2550		
		Variance	2371.980		
		Std. Deviation	48.70298		
		Minimum	88.02		
		Maximum	247.00		
		Range	158.98		
		Interquartile Range	84.20		
		Skewness	.522	.414	
		Kurtosis	-1.172	.809	
Cortisol_W12	1.00	Mean	127.4913	9.12701	
		95% Confidence Interval for Mean	Lower Bound	108.8766		
			Upper Bound	146.1059		
		5% Trimmed Mean	122.0604		
		Median	102.4100		
		Variance	2665.672		
		Std. Deviation	51.63015		
		Minimum	84.70		
		Maximum	274.21		
		Range	189.51		
		Interquartile Range	75.46		
		Skewness	1.407	.414	
		Kurtosis	1.261	.809	
	2.00	Mean	132.2741	8.87568	
		95% Confidence Interval for Mean	Lower Bound	114.1720		
			Upper Bound	150.3761		
		5% Trimmed Mean	128.3617		
		Median	109.5650		
		Variance	2520.884		
		Std. Deviation	50.20841		
		Minimum	86.70		
		Maximum	256.63		
		Range	169.93		
		Interquartile Range	96.07		
		Skewness	1.036	.414	
		Kurtosis	-.267	.809	
Cortisol_Diff_W12	1.00	Mean	18.4750	2.78398	
		95% Confidence Interval for Mean	Lower Bound	12.7970		
			Upper Bound	24.1530		
		5% Trimmed Mean	17.6485		
		Median	15.3450		
		Variance	248.017		
		Std. Deviation	15.74856		
		Minimum	-15.41		
		Maximum	76.12		
		Range	91.53		
		Interquartile Range	18.45		
		Skewness	1.321	.414	
		Kurtosis	4.996	.809	
	2.00	Mean	11.7406	5.21718	
		95% Confidence Interval for Mean	Lower Bound	1.1001		
			Upper Bound	22.3811		
		5% Trimmed Mean	8.8291		
		Median	3.7100		
		Variance	871.008		
		Std. Deviation	29.51284		
		Minimum	-30.51		
		Maximum	112.26		
		Range	142.77		
		Interquartile Range	21.82		
		Skewness	1.891	.414	
		Kurtosis	4.431	.809	
Serum_Ferritin_B	1.00	Mean	41.2441	8.23691	
		95% Confidence Interval for Mean	Lower Bound	24.4448		
			Upper Bound	58.0433		
		5% Trimmed Mean	35.2562		
		Median	20.6500		
		Variance	2171.093		
		Std. Deviation	46.59499		
		Minimum	9.32		
		Maximum	187.41		
		Range	178.09		
		Interquartile Range	25.66		
		Skewness	2.338	.414	
		Kurtosis	4.629	.809	
	2.00	Mean	41.6381	10.22330	
		95% Confidence Interval for Mean	Lower Bound	20.7876		
			Upper Bound	62.4887		
		5% Trimmed Mean	32.1691		
		Median	19.3050		
		Variance	3344.507		
		Std. Deviation	57.83172		
		Minimum	9.61		
		Maximum	244.00		
		Range	234.39		
		Interquartile Range	26.77		
		Skewness	2.984	.414	
		Kurtosis	8.608	.809	
Serum_Ferritin_W12	1.00	Mean	51.4975	11.14986	
		95% Confidence Interval for Mean	Lower Bound	28.7572		
			Upper Bound	74.2378		
		5% Trimmed Mean	43.6947		
		Median	20.4900		
		Variance	3978.223		
		Std. Deviation	63.07316		
		Minimum	12.33		
		Maximum	241.21		
		Range	228.88		
		Interquartile Range	30.94		
		Skewness	1.964	.414	
		Kurtosis	2.777	.809	
	2.00	Mean	60.2050	10.98242	
		95% Confidence Interval for Mean	Lower Bound	37.8062		
			Upper Bound	82.6038		
		5% Trimmed Mean	53.2356		
		Median	26.9250		
		Variance	3859.632		
		Std. Deviation	62.12594		
		Minimum	12.10		
		Maximum	254.10		
		Range	242.00		
		Interquartile Range	83.62		
		Skewness	1.601	.414	
		Kurtosis	2.055	.809	
Serum_Ferritin_Diff_W12	1.00	Mean	-10.2534	14.08781	
		95% Confidence Interval for Mean	Lower Bound	-38.9857		
			Upper Bound	18.4788		
		5% Trimmed Mean	-9.8251		
		Median	-.3450		
		Variance	6350.921		
		Std. Deviation	79.69267		
		Minimum	-198.78		
		Maximum	167.54		
		Range	366.32		
		Interquartile Range	23.18		
		Skewness	-.382	.414	
		Kurtosis	1.159	.809	
	2.00	Mean	-18.5669	16.04159	
		95% Confidence Interval for Mean	Lower Bound	-51.2839		
			Upper Bound	14.1502		
		5% Trimmed Mean	-21.2017		
		Median	-6.4300		
		Variance	8234.646		
		Std. Deviation	90.74495		
		Minimum	-235.09		
		Maximum	226.00		
		Range	461.09		
		Interquartile Range	81.53		
		Skewness	.446	.414	
		Kurtosis	2.621	.809	
Serum_iron_B	1.00	Mean	79.8084	6.45507	
		95% Confidence Interval for Mean	Lower Bound	66.6432		
			Upper Bound	92.9736		
		5% Trimmed Mean	77.2866		
		Median	78.7200		
		Variance	1333.373		
		Std. Deviation	36.51539		
		Minimum	30.41		
		Maximum	192.09		
		Range	161.68		
		Interquartile Range	46.73		
		Skewness	1.084	.414	
		Kurtosis	1.493	.809	
	2.00	Mean	70.5784	6.48859	
		95% Confidence Interval for Mean	Lower Bound	57.3449		
			Upper Bound	83.8120		
		5% Trimmed Mean	68.5269		
		Median	60.0200		
		Variance	1347.257		
		Std. Deviation	36.70500		
		Minimum	21.90		
		Maximum	157.80		
		Range	135.90		
		Interquartile Range	55.02		
		Skewness	.850	.414	
		Kurtosis	-.182	.809	
Serum_iron_W12	1.00	Mean	97.9108	5.74769	
		95% Confidence Interval for Mean	Lower Bound	86.1883		
			Upper Bound	109.6333		
		5% Trimmed Mean	96.7167		
		Median	89.3150		
		Variance	1057.151		
		Std. Deviation	32.51386		
		Minimum	58.00		
		Maximum	160.14		
		Range	102.14		
		Interquartile Range	56.19		
		Skewness	.372	.414	
		Kurtosis	-1.139	.809	
	2.00	Mean	105.8822	6.66353	
		95% Confidence Interval for Mean	Lower Bound	92.2918		
			Upper Bound	119.4725		
		5% Trimmed Mean	105.3730		
		Median	110.7750		
		Variance	1420.883		
		Std. Deviation	37.69460		
		Minimum	53.65		
		Maximum	170.25		
		Range	116.60		
		Interquartile Range	74.45		
		Skewness	.080	.414	
		Kurtosis	-1.402	.809	
Serum_iron_Diff_W12	1.00	Mean	-18.1023	8.94162	
		95% Confidence Interval for Mean	Lower Bound	-36.3389		
			Upper Bound	.1342		
		5% Trimmed Mean	-18.1133		
		Median	-14.9050		
		Variance	2558.485		
		Std. Deviation	50.58147		
		Minimum	-111.91		
		Maximum	82.36		
		Range	194.27		
		Interquartile Range	85.51		
		Skewness	-.101	.414	
		Kurtosis	-1.010	.809	
	2.00	Mean	-35.3038	7.95596	
		95% Confidence Interval for Mean	Lower Bound	-51.5300		
			Upper Bound	-19.0775		
		5% Trimmed Mean	-36.7822		
		Median	-29.3400		
		Variance	2025.515		
		Std. Deviation	45.00572		
		Minimum	-120.65		
		Maximum	95.21		
		Range	215.86		
		Interquartile Range	69.92		
		Skewness	.367	.414	
		Kurtosis	.911	.809	
