## Supplementary material for "Effects of a standardised *Amaranthus tricolor*–*Spinacia oleracea* extract on exercise performance, muscle strength, endurance and recovery in physically active adults: a randomised, double-blind, placebo-controlled trial": Raw statistical outputs: CPET_Output_17.07.2026.doc

Explore


Notes	
Output Created	17-JUL-2026 11:11:50	
Comments		
Input	Active Dataset	DataSet0	
	Filter	<none>	
	Weight	<none>	
	Split File	<none>	
	N of Rows in Working Data File	60	
Missing Value Handling	Definition of Missing	User-defined missing values for dependent variables are treated as missing.	
	Cases Used	Statistics are based on cases with no missing values for any dependent variable or factor used.	
Syntax	EXAMINE VARIABLES=VO2_max_B VO2_max_W6 VO2_max_W12 VO2_max_Diff_W6 VO2_max_Diff_W12 Max_Vent_B
    Max_Vent_W6 Max_Vent_W12 Max_Vent_Diff_W6 Max_Vent_Diff_W12 VO2_Peak_B VO2_Peak_W6 VO2_Peak_W12
    VO2_Peak_Diff_W6 VO2_Peak_Diff_W12 Vent_Thres_B Vent_Thres_W6 Vent_Thres_W12 Vent_Thres_Diff_W6
    Vent_Thres_Diff_W12 Resp_Thres_B Resp_Thres_W6 Resp_Thres_W12 Resp_Thres_Diff_W6
    Resp_Thres_Diff_W12 VO2_85_B VO2_85_W6 VO2_85_W12 VO2_85_Diff_W6 VO2_85_Diff_W12 Exhaust_B
    Exhaust_W6 Exhaust_W12 Exhaust_Diff_W6 Exhaust_Diff_W12 BY Group
  /PLOT BOXPLOT STEMLEAF
  /COMPARE GROUPS
  /STATISTICS DESCRIPTIVES
  /CINTERVAL 95
  /MISSING LISTWISE
  /NOTOTAL.	
Resources	Processor Time	00:00:08.27	
	Elapsed Time	00:00:03.82	


[DataSet0] 


Group


Case Processing Summary	
	Group	Cases	
		Valid	Missing	Total	
		N	Percent	N	Percent	N	Percent	
VO2_max_B	1.00	28	93.3%	2	6.7%	30	100.0%	
	2.00	26	86.7%	4	13.3%	30	100.0%	
VO2_max_W6	1.00	28	93.3%	2	6.7%	30	100.0%	
	2.00	26	86.7%	4	13.3%	30	100.0%	
VO2_max_W12	1.00	28	93.3%	2	6.7%	30	100.0%	
	2.00	26	86.7%	4	13.3%	30	100.0%	
VO2_max_Diff_W6	1.00	28	93.3%	2	6.7%	30	100.0%	
	2.00	26	86.7%	4	13.3%	30	100.0%	
VO2_max_Diff_W12	1.00	28	93.3%	2	6.7%	30	100.0%	
	2.00	26	86.7%	4	13.3%	30	100.0%	
Max_Vent_B	1.00	28	93.3%	2	6.7%	30	100.0%	
	2.00	26	86.7%	4	13.3%	30	100.0%	
Max_Vent_W6	1.00	28	93.3%	2	6.7%	30	100.0%	
	2.00	26	86.7%	4	13.3%	30	100.0%	
Max_Vent_W12	1.00	28	93.3%	2	6.7%	30	100.0%	
	2.00	26	86.7%	4	13.3%	30	100.0%	
Max_Vent_Diff_W6	1.00	28	93.3%	2	6.7%	30	100.0%	
	2.00	26	86.7%	4	13.3%	30	100.0%	
Max_Vent_Diff_W12	1.00	28	93.3%	2	6.7%	30	100.0%	
	2.00	26	86.7%	4	13.3%	30	100.0%	
VO2_Peak_B	1.00	28	93.3%	2	6.7%	30	100.0%	
	2.00	26	86.7%	4	13.3%	30	100.0%	
VO2_Peak_W6	1.00	28	93.3%	2	6.7%	30	100.0%	
	2.00	26	86.7%	4	13.3%	30	100.0%	
VO2_Peak_W12	1.00	28	93.3%	2	6.7%	30	100.0%	
	2.00	26	86.7%	4	13.3%	30	100.0%	
VO2_Peak_Diff_W6	1.00	28	93.3%	2	6.7%	30	100.0%	
	2.00	26	86.7%	4	13.3%	30	100.0%	
VO2_Peak_Diff_W12	1.00	28	93.3%	2	6.7%	30	100.0%	
	2.00	26	86.7%	4	13.3%	30	100.0%	
Vent_Thres_B	1.00	28	93.3%	2	6.7%	30	100.0%	
	2.00	26	86.7%	4	13.3%	30	100.0%	
Vent_Thres_W6	1.00	28	93.3%	2	6.7%	30	100.0%	
	2.00	26	86.7%	4	13.3%	30	100.0%	
Vent_Thres_W12	1.00	28	93.3%	2	6.7%	30	100.0%	
	2.00	26	86.7%	4	13.3%	30	100.0%	
Vent_Thres_Diff_W6	1.00	28	93.3%	2	6.7%	30	100.0%	
	2.00	26	86.7%	4	13.3%	30	100.0%	
Vent_Thres_Diff_W12	1.00	28	93.3%	2	6.7%	30	100.0%	
	2.00	26	86.7%	4	13.3%	30	100.0%	
Resp_Thres_B	1.00	28	93.3%	2	6.7%	30	100.0%	
	2.00	26	86.7%	4	13.3%	30	100.0%	
Resp_Thres_W6	1.00	28	93.3%	2	6.7%	30	100.0%	
	2.00	26	86.7%	4	13.3%	30	100.0%	
Resp_Thres_W12	1.00	28	93.3%	2	6.7%	30	100.0%	
	2.00	26	86.7%	4	13.3%	30	100.0%	
Resp_Thres_Diff_W6	1.00	28	93.3%	2	6.7%	30	100.0%	
	2.00	26	86.7%	4	13.3%	30	100.0%	
Resp_Thres_Diff_W12	1.00	28	93.3%	2	6.7%	30	100.0%	
	2.00	26	86.7%	4	13.3%	30	100.0%	
VO2_85_B	1.00	28	93.3%	2	6.7%	30	100.0%	
	2.00	26	86.7%	4	13.3%	30	100.0%	
VO2_85_W6	1.00	28	93.3%	2	6.7%	30	100.0%	
	2.00	26	86.7%	4	13.3%	30	100.0%	
VO2_85_W12	1.00	28	93.3%	2	6.7%	30	100.0%	
	2.00	26	86.7%	4	13.3%	30	100.0%	
VO2_85_Diff_W6	1.00	28	93.3%	2	6.7%	30	100.0%	
	2.00	26	86.7%	4	13.3%	30	100.0%	
VO2_85_Diff_W12	1.00	28	93.3%	2	6.7%	30	100.0%	
	2.00	26	86.7%	4	13.3%	30	100.0%	
Exhaust_B	1.00	28	93.3%	2	6.7%	30	100.0%	
	2.00	26	86.7%	4	13.3%	30	100.0%	
Exhaust_W6	1.00	28	93.3%	2	6.7%	30	100.0%	
	2.00	26	86.7%	4	13.3%	30	100.0%	
Exhaust_W12	1.00	28	93.3%	2	6.7%	30	100.0%	
	2.00	26	86.7%	4	13.3%	30	100.0%	
Exhaust_Diff_W6	1.00	28	93.3%	2	6.7%	30	100.0%	
	2.00	26	86.7%	4	13.3%	30	100.0%	
Exhaust_Diff_W12	1.00	28	93.3%	2	6.7%	30	100.0%	
	2.00	26	86.7%	4	13.3%	30	100.0%	


Descriptives	
	Group	Statistic	Std. Error	
VO2_max_B	1.00	Mean	2.1025	.08040	
		95% Confidence Interval for Mean	Lower Bound	1.9375		
			Upper Bound	2.2675		
		5% Trimmed Mean	2.1066		
		Median	2.1150		
		Variance	.181		
		Std. Deviation	.42545		
		Minimum	1.20		
		Maximum	2.94		
		Range	1.74		
		Interquartile Range	.52		
		Skewness	-.124	.441	
		Kurtosis	.189	.858	
	2.00	Mean	2.1200	.08103	
		95% Confidence Interval for Mean	Lower Bound	1.9531		
			Upper Bound	2.2869		
		5% Trimmed Mean	2.1143		
		Median	2.0650		
		Variance	.171		
		Std. Deviation	.41317		
		Minimum	1.36		
		Maximum	2.96		
		Range	1.60		
		Interquartile Range	.54		
		Skewness	.495	.456	
		Kurtosis	-.264	.887	
VO2_max_W6	1.00	Mean	2.1236	.07561	
		95% Confidence Interval for Mean	Lower Bound	1.9684		
			Upper Bound	2.2787		
		5% Trimmed Mean	2.1266		
		Median	2.0750		
		Variance	.160		
		Std. Deviation	.40009		
		Minimum	1.15		
		Maximum	3.10		
		Range	1.95		
		Interquartile Range	.43		
		Skewness	-.117	.441	
		Kurtosis	.955	.858	
	2.00	Mean	2.0308	.08306	
		95% Confidence Interval for Mean	Lower Bound	1.8597		
			Upper Bound	2.2018		
		5% Trimmed Mean	2.0247		
		Median	2.0200		
		Variance	.179		
		Std. Deviation	.42351		
		Minimum	1.34		
		Maximum	2.86		
		Range	1.52		
		Interquartile Range	.66		
		Skewness	.162	.456	
		Kurtosis	-.711	.887	
VO2_max_W12	1.00	Mean	2.2571	.08081	
		95% Confidence Interval for Mean	Lower Bound	2.0913		
			Upper Bound	2.4230		
		5% Trimmed Mean	2.2443		
		Median	2.2150		
		Variance	.183		
		Std. Deviation	.42761		
		Minimum	1.45		
		Maximum	3.25		
		Range	1.80		
		Interquartile Range	.58		
		Skewness	.550	.441	
		Kurtosis	.368	.858	
	2.00	Mean	2.1673	.08801	
		95% Confidence Interval for Mean	Lower Bound	1.9861		
			Upper Bound	2.3486		
		5% Trimmed Mean	2.1607		
		Median	2.1700		
		Variance	.201		
		Std. Deviation	.44875		
		Minimum	1.36		
		Maximum	3.08		
		Range	1.72		
		Interquartile Range	.62		
		Skewness	.284	.456	
		Kurtosis	-.353	.887	
VO2_max_Diff_W6	1.00	Mean	-.0211	.05175	
		95% Confidence Interval for Mean	Lower Bound	-.1273		
			Upper Bound	.0851		
		5% Trimmed Mean	-.0279		
		Median	-.0800		
		Variance	.075		
		Std. Deviation	.27384		
		Minimum	-.57		
		Maximum	.75		
		Range	1.32		
		Interquartile Range	.33		
		Skewness	.493	.441	
		Kurtosis	1.209	.858	
	2.00	Mean	.0892	.05527	
		95% Confidence Interval for Mean	Lower Bound	-.0246		
			Upper Bound	.2031		
		5% Trimmed Mean	.0823		
		Median	.0800		
		Variance	.079		
		Std. Deviation	.28181		
		Minimum	-.52		
		Maximum	.78		
		Range	1.30		
		Interquartile Range	.32		
		Skewness	.511	.456	
		Kurtosis	.906	.887	
VO2_max_Diff_W12	1.00	Mean	-.1546	.06892	
		95% Confidence Interval for Mean	Lower Bound	-.2960		
			Upper Bound	-.0132		
		5% Trimmed Mean	-.1455		
		Median	-.1100		
		Variance	.133		
		Std. Deviation	.36467		
		Minimum	-1.15		
		Maximum	.61		
		Range	1.76		
		Interquartile Range	.46		
		Skewness	-.368	.441	
		Kurtosis	1.006	.858	
	2.00	Mean	-.0473	.08804	
		95% Confidence Interval for Mean	Lower Bound	-.2286		
			Upper Bound	.1340		
		5% Trimmed Mean	-.0455		
		Median	-.0600		
		Variance	.202		
		Std. Deviation	.44891		
		Minimum	-1.02		
		Maximum	.92		
		Range	1.94		
		Interquartile Range	.42		
		Skewness	-.298	.456	
		Kurtosis	.869	.887	
Max_Vent_B	1.00	Mean	79.1089	2.90908	
		95% Confidence Interval for Mean	Lower Bound	73.1400		
			Upper Bound	85.0779		
		5% Trimmed Mean	78.8203		
		Median	79.7350		
		Variance	236.957		
		Std. Deviation	15.39340		
		Minimum	46.91		
		Maximum	118.08		
		Range	71.17		
		Interquartile Range	20.05		
		Skewness	.272	.441	
		Kurtosis	.748	.858	
	2.00	Mean	81.0077	3.91694	
		95% Confidence Interval for Mean	Lower Bound	72.9406		
			Upper Bound	89.0748		
		5% Trimmed Mean	80.9497		
		Median	79.7750		
		Variance	398.903		
		Std. Deviation	19.97255		
		Minimum	46.77		
		Maximum	117.49		
		Range	70.72		
		Interquartile Range	30.76		
		Skewness	.002	.456	
		Kurtosis	-.894	.887	
Max_Vent_W6	1.00	Mean	75.4979	2.63295	
		95% Confidence Interval for Mean	Lower Bound	70.0955		
			Upper Bound	80.9002		
		5% Trimmed Mean	76.0895		
		Median	75.8050		
		Variance	194.107		
		Std. Deviation	13.93224		
		Minimum	42.80		
		Maximum	98.96		
		Range	56.16		
		Interquartile Range	19.06		
		Skewness	-.689	.441	
		Kurtosis	.265	.858	
	2.00	Mean	72.6027	3.77386	
		95% Confidence Interval for Mean	Lower Bound	64.8303		
			Upper Bound	80.3751		
		5% Trimmed Mean	72.4093		
		Median	70.5550		
		Variance	370.292		
		Std. Deviation	19.24297		
		Minimum	38.74		
		Maximum	108.86		
		Range	70.12		
		Interquartile Range	31.24		
		Skewness	.205	.456	
		Kurtosis	-.668	.887	
Max_Vent_W12	1.00	Mean	81.2018	3.03241	
		95% Confidence Interval for Mean	Lower Bound	74.9798		
			Upper Bound	87.4238		
		5% Trimmed Mean	81.0144		
		Median	82.2100		
		Variance	257.474		
		Std. Deviation	16.04599		
		Minimum	53.60		
		Maximum	112.12		
		Range	58.52		
		Interquartile Range	26.90		
		Skewness	.057	.441	
		Kurtosis	-.807	.858	
	2.00	Mean	76.8592	4.01166	
		95% Confidence Interval for Mean	Lower Bound	68.5971		
			Upper Bound	85.1214		
		5% Trimmed Mean	75.7635		
		Median	72.4300		
		Variance	418.429		
		Std. Deviation	20.45553		
		Minimum	44.48		
		Maximum	131.53		
		Range	87.05		
		Interquartile Range	23.48		
		Skewness	1.034	.456	
		Kurtosis	.978	.887	
Max_Vent_Diff_W6	1.00	Mean	3.6111	2.56929	
		95% Confidence Interval for Mean	Lower Bound	-1.6607		
			Upper Bound	8.8828		
		5% Trimmed Mean	3.0541		
		Median	1.1450		
		Variance	184.835		
		Std. Deviation	13.59541		
		Minimum	-18.22		
		Maximum	38.07		
		Range	56.29		
		Interquartile Range	15.49		
		Skewness	.598	.441	
		Kurtosis	.236	.858	
	2.00	Mean	8.4050	2.53377	
		95% Confidence Interval for Mean	Lower Bound	3.1866		
			Upper Bound	13.6234		
		5% Trimmed Mean	9.1388		
		Median	10.5000		
		Variance	166.920		
		Std. Deviation	12.91976		
		Minimum	-31.27		
		Maximum	32.87		
		Range	64.14		
		Interquartile Range	12.41		
		Skewness	-1.083	.456	
		Kurtosis	2.813	.887	
Max_Vent_Diff_W12	1.00	Mean	-2.0929	3.41890	
		95% Confidence Interval for Mean	Lower Bound	-9.1079		
			Upper Bound	4.9221		
		5% Trimmed Mean	-1.6442		
		Median	-3.3000		
		Variance	327.288		
		Std. Deviation	18.09110		
		Minimum	-43.49		
		Maximum	31.92		
		Range	75.41		
		Interquartile Range	21.97		
		Skewness	-.301	.441	
		Kurtosis	.469	.858	
	2.00	Mean	4.1485	3.62180	
		95% Confidence Interval for Mean	Lower Bound	-3.3108		
			Upper Bound	11.6077		
		5% Trimmed Mean	4.2493		
		Median	4.3100		
		Variance	341.054		
		Std. Deviation	18.46765		
		Minimum	-35.30		
		Maximum	42.49		
		Range	77.79		
		Interquartile Range	25.31		
		Skewness	-.174	.456	
		Kurtosis	.193	.887	
VO2_Peak_B	1.00	Mean	27.9650	1.06245	
		95% Confidence Interval for Mean	Lower Bound	25.7850		
			Upper Bound	30.1450		
		5% Trimmed Mean	27.9220		
		Median	27.9150		
		Variance	31.607		
		Std. Deviation	5.62198		
		Minimum	17.37		
		Maximum	40.50		
		Range	23.13		
		Interquartile Range	8.56		
		Skewness	.169	.441	
		Kurtosis	-.292	.858	
	2.00	Mean	28.2537	.99553	
		95% Confidence Interval for Mean	Lower Bound	26.2034		
			Upper Bound	30.3041		
		5% Trimmed Mean	28.1213		
		Median	28.0150		
		Variance	25.768		
		Std. Deviation	5.07624		
		Minimum	19.51		
		Maximum	39.57		
		Range	20.06		
		Interquartile Range	6.34		
		Skewness	.458	.456	
		Kurtosis	.085	.887	
VO2_Peak_W6	1.00	Mean	28.0729	1.02105	
		95% Confidence Interval for Mean	Lower Bound	25.9778		
			Upper Bound	30.1679		
		5% Trimmed Mean	27.8977		
		Median	27.2600		
		Variance	29.191		
		Std. Deviation	5.40289		
		Minimum	16.91		
		Maximum	43.06		
		Range	26.15		
		Interquartile Range	6.05		
		Skewness	.676	.441	
		Kurtosis	1.443	.858	
	2.00	Mean	27.1300	1.07371	
		95% Confidence Interval for Mean	Lower Bound	24.9187		
			Upper Bound	29.3413		
		5% Trimmed Mean	26.9643		
		Median	26.7750		
		Variance	29.974		
		Std. Deviation	5.47486		
		Minimum	17.43		
		Maximum	40.57		
		Range	23.14		
		Interquartile Range	7.70		
		Skewness	.267	.456	
		Kurtosis	.196	.887	
VO2_Peak_W12	1.00	Mean	29.5050	.98778	
		95% Confidence Interval for Mean	Lower Bound	27.4782		
			Upper Bound	31.5318		
		5% Trimmed Mean	29.2414		
		Median	29.0800		
		Variance	27.320		
		Std. Deviation	5.22685		
		Minimum	20.86		
		Maximum	42.82		
		Range	21.96		
		Interquartile Range	4.62		
		Skewness	.992	.441	
		Kurtosis	.987	.858	
	2.00	Mean	28.7585	1.10991	
		95% Confidence Interval for Mean	Lower Bound	26.4726		
			Upper Bound	31.0444		
		5% Trimmed Mean	28.6809		
		Median	28.7400		
		Variance	32.029		
		Std. Deviation	5.65945		
		Minimum	17.55		
		Maximum	41.12		
		Range	23.57		
		Interquartile Range	7.06		
		Skewness	.229	.456	
		Kurtosis	.142	.887	
VO2_Peak_Diff_W6	1.00	Mean	-.1079	.76158	
		95% Confidence Interval for Mean	Lower Bound	-1.6705		
			Upper Bound	1.4548		
		5% Trimmed Mean	-.2464		
		Median	-.3000		
		Variance	16.240		
		Std. Deviation	4.02992		
		Minimum	-8.37		
		Maximum	11.98		
		Range	20.35		
		Interquartile Range	4.87		
		Skewness	.693	.441	
		Kurtosis	1.868	.858	
	2.00	Mean	1.1237	.69388	
		95% Confidence Interval for Mean	Lower Bound	-.3053		
			Upper Bound	2.5528		
		5% Trimmed Mean	1.0371		
		Median	1.2100		
		Variance	12.518		
		Std. Deviation	3.53810		
		Minimum	-6.65		
		Maximum	9.93		
		Range	16.58		
		Interquartile Range	4.51		
		Skewness	.445	.456	
		Kurtosis	1.004	.887	
VO2_Peak_Diff_W12	1.00	Mean	-1.5400	.90192	
		95% Confidence Interval for Mean	Lower Bound	-3.3906		
			Upper Bound	.3106		
		5% Trimmed Mean	-1.4883		
		Median	-1.2550		
		Variance	22.777		
		Std. Deviation	4.77251		
		Minimum	-13.57		
		Maximum	9.72		
		Range	23.29		
		Interquartile Range	5.62		
		Skewness	-.078	.441	
		Kurtosis	1.061	.858	
	2.00	Mean	-.5047	1.15289	
		95% Confidence Interval for Mean	Lower Bound	-2.8792		
			Upper Bound	1.8697		
		5% Trimmed Mean	-.4906		
		Median	-.6440		
		Variance	34.558		
		Std. Deviation	5.87863		
		Minimum	-12.93		
		Maximum	12.11		
		Range	25.04		
		Interquartile Range	5.34		
		Skewness	-.362	.456	
		Kurtosis	.836	.887	
Vent_Thres_B	1.00	Mean	1.8186	.07463	
		95% Confidence Interval for Mean	Lower Bound	1.6654		
			Upper Bound	1.9717		
		5% Trimmed Mean	1.8153		
		Median	1.7600		
		Variance	.156		
		Std. Deviation	.39492		
		Minimum	1.01		
		Maximum	2.76		
		Range	1.75		
		Interquartile Range	.47		
		Skewness	.116	.441	
		Kurtosis	.437	.858	
	2.00	Mean	1.8388	.09853	
		95% Confidence Interval for Mean	Lower Bound	1.6359		
			Upper Bound	2.0418		
		5% Trimmed Mean	1.7909		
		Median	1.8150		
		Variance	.252		
		Std. Deviation	.50240		
		Minimum	1.05		
		Maximum	3.74		
		Range	2.69		
		Interquartile Range	.47		
		Skewness	2.161	.456	
		Kurtosis	7.662	.887	
Vent_Thres_W6	1.00	Mean	1.7750	.06562	
		95% Confidence Interval for Mean	Lower Bound	1.6404		
			Upper Bound	1.9096		
		5% Trimmed Mean	1.7872		
		Median	1.8500		
		Variance	.121		
		Std. Deviation	.34723		
		Minimum	.91		
		Maximum	2.40		
		Range	1.49		
		Interquartile Range	.51		
		Skewness	-.636	.441	
		Kurtosis	.148	.858	
	2.00	Mean	1.7200	.07033	
		95% Confidence Interval for Mean	Lower Bound	1.5752		
			Upper Bound	1.8648		
		5% Trimmed Mean	1.7298		
		Median	1.7600		
		Variance	.129		
		Std. Deviation	.35861		
		Minimum	.92		
		Maximum	2.32		
		Range	1.40		
		Interquartile Range	.47		
		Skewness	-.353	.456	
		Kurtosis	-.235	.887	
Vent_Thres_W12	1.00	Mean	1.9346	.07285	
		95% Confidence Interval for Mean	Lower Bound	1.7852		
			Upper Bound	2.0841		
		5% Trimmed Mean	1.9293		
		Median	1.8850		
		Variance	.149		
		Std. Deviation	.38549		
		Minimum	1.09		
		Maximum	2.81		
		Range	1.72		
		Interquartile Range	.48		
		Skewness	.188	.441	
		Kurtosis	.482	.858	
	2.00	Mean	1.7427	.06931	
		95% Confidence Interval for Mean	Lower Bound	1.5999		
			Upper Bound	1.8854		
		5% Trimmed Mean	1.7376		
		Median	1.7150		
		Variance	.125		
		Std. Deviation	.35344		
		Minimum	.97		
		Maximum	2.68		
		Range	1.71		
		Interquartile Range	.51		
		Skewness	.273	.456	
		Kurtosis	1.115	.887	
Vent_Thres_Diff_W6	1.00	Mean	.0436	.03890	
		95% Confidence Interval for Mean	Lower Bound	-.0363		
			Upper Bound	.1234		
		5% Trimmed Mean	.0374		
		Median	.0450		
		Variance	.042		
		Std. Deviation	.20586		
		Minimum	-.26		
		Maximum	.49		
		Range	.75		
		Interquartile Range	.35		
		Skewness	.251	.441	
		Kurtosis	-.842	.858	
	2.00	Mean	.1188	.06767	
		95% Confidence Interval for Mean	Lower Bound	-.0205		
			Upper Bound	.2582		
		5% Trimmed Mean	.0712		
		Median	.0700		
		Variance	.119		
		Std. Deviation	.34507		
		Minimum	-.25		
		Maximum	1.62		
		Range	1.87		
		Interquartile Range	.17		
		Skewness	3.436	.456	
		Kurtosis	15.061	.887	
Vent_Thres_Diff_W12	1.00	Mean	-.1161	.05344	
		95% Confidence Interval for Mean	Lower Bound	-.2257		
			Upper Bound	-.0064		
		5% Trimmed Mean	-.1002		
		Median	-.1050		
		Variance	.080		
		Std. Deviation	.28278		
		Minimum	-1.10		
		Maximum	.44		
		Range	1.54		
		Interquartile Range	.25		
		Skewness	-1.247	.441	
		Kurtosis	4.715	.858	
	2.00	Mean	.0962	.08659	
		95% Confidence Interval for Mean	Lower Bound	-.0822		
			Upper Bound	.2745		
		5% Trimmed Mean	.0623		
		Median	.0600		
		Variance	.195		
		Std. Deviation	.44154		
		Minimum	-.79		
		Maximum	1.86		
		Range	2.65		
		Interquartile Range	.28		
		Skewness	2.316	.456	
		Kurtosis	10.575	.887	
Resp_Thres_B	1.00	Mean	.9336	.00649	
		95% Confidence Interval for Mean	Lower Bound	.9203		
			Upper Bound	.9469		
		5% Trimmed Mean	.9347		
		Median	.9400		
		Variance	.001		
		Std. Deviation	.03434		
		Minimum	.86		
		Maximum	.99		
		Range	.13		
		Interquartile Range	.04		
		Skewness	-.859	.441	
		Kurtosis	.381	.858	
	2.00	Mean	.9258	.00996	
		95% Confidence Interval for Mean	Lower Bound	.9053		
			Upper Bound	.9463		
		5% Trimmed Mean	.9315		
		Median	.9350		
		Variance	.003		
		Std. Deviation	.05077		
		Minimum	.73		
		Maximum	.99		
		Range	.26		
		Interquartile Range	.05		
		Skewness	-2.345	.456	
		Kurtosis	8.333	.887	
Resp_Thres_W6	1.00	Mean	.9254	.00648	
		95% Confidence Interval for Mean	Lower Bound	.9121		
			Upper Bound	.9386		
		5% Trimmed Mean	.9269		
		Median	.9300		
		Variance	.001		
		Std. Deviation	.03426		
		Minimum	.84		
		Maximum	.98		
		Range	.14		
		Interquartile Range	.04		
		Skewness	-.721	.441	
		Kurtosis	.475	.858	
	2.00	Mean	.9273	.00634	
		95% Confidence Interval for Mean	Lower Bound	.9143		
			Upper Bound	.9404		
		5% Trimmed Mean	.9287		
		Median	.9300		
		Variance	.001		
		Std. Deviation	.03232		
		Minimum	.85		
		Maximum	.98		
		Range	.13		
		Interquartile Range	.03		
		Skewness	-.816	.456	
		Kurtosis	.371	.887	
Resp_Thres_W12	1.00	Mean	.9364	.00609	
		95% Confidence Interval for Mean	Lower Bound	.9239		
			Upper Bound	.9489		
		5% Trimmed Mean	.9362		
		Median	.9400		
		Variance	.001		
		Std. Deviation	.03223		
		Minimum	.87		
		Maximum	1.01		
		Range	.14		
		Interquartile Range	.03		
		Skewness	.047	.441	
		Kurtosis	.230	.858	
	2.00	Mean	.9281	.00433	
		95% Confidence Interval for Mean	Lower Bound	.9192		
			Upper Bound	.9370		
		5% Trimmed Mean	.9284		
		Median	.9300		
		Variance	.000		
		Std. Deviation	.02209		
		Minimum	.88		
		Maximum	.97		
		Range	.09		
		Interquartile Range	.04		
		Skewness	-.267	.456	
		Kurtosis	-.330	.887	
Resp_Thres_Diff_W6	1.00	Mean	.0082	.00402	
		95% Confidence Interval for Mean	Lower Bound	.0000		
			Upper Bound	.0165		
		5% Trimmed Mean	.0088		
		Median	.0100		
		Variance	.000		
		Std. Deviation	.02127		
		Minimum	-.05		
		Maximum	.05		
		Range	.10		
		Interquartile Range	.03		
		Skewness	-.421	.441	
		Kurtosis	.904	.858	
	2.00	Mean	-.0015	.00916	
		95% Confidence Interval for Mean	Lower Bound	-.0204		
			Upper Bound	.0173		
		5% Trimmed Mean	.0017		
		Median	.0050		
		Variance	.002		
		Std. Deviation	.04671		
		Minimum	-.16		
		Maximum	.08		
		Range	.24		
		Interquartile Range	.04		
		Skewness	-1.359	.456	
		Kurtosis	4.583	.887	
Resp_Thres_Diff_W12	1.00	Mean	-.0029	.00468	
		95% Confidence Interval for Mean	Lower Bound	-.0125		
			Upper Bound	.0067		
		5% Trimmed Mean	-.0022		
		Median	.0000		
		Variance	.001		
		Std. Deviation	.02477		
		Minimum	-.06		
		Maximum	.04		
		Range	.10		
		Interquartile Range	.04		
		Skewness	-.339	.441	
		Kurtosis	-.403	.858	
	2.00	Mean	-.0023	.00955	
		95% Confidence Interval for Mean	Lower Bound	-.0220		
			Upper Bound	.0174		
		5% Trimmed Mean	.0021		
		Median	.0050		
		Variance	.002		
		Std. Deviation	.04869		
		Minimum	-.18		
		Maximum	.08		
		Range	.26		
		Interquartile Range	.05		
		Skewness	-1.948	.456	
		Kurtosis	6.433	.887	
VO2_85_B	1.00	Mean	1.7871	.06834	
		95% Confidence Interval for Mean	Lower Bound	1.6469		
			Upper Bound	1.9274		
		5% Trimmed Mean	1.7906		
		Median	1.7978		
		Variance	.131		
		Std. Deviation	.36163		
		Minimum	1.02		
		Maximum	2.50		
		Range	1.48		
		Interquartile Range	.44		
		Skewness	-.124	.441	
		Kurtosis	.189	.858	
	2.00	Mean	1.8020	.06888	
		95% Confidence Interval for Mean	Lower Bound	1.6601		
			Upper Bound	1.9439		
		5% Trimmed Mean	1.7971		
		Median	1.7553		
		Variance	.123		
		Std. Deviation	.35120		
		Minimum	1.16		
		Maximum	2.52		
		Range	1.36		
		Interquartile Range	.46		
		Skewness	.495	.456	
		Kurtosis	-.264	.887	
VO2_85_W6	1.00	Mean	1.8050	.06427	
		95% Confidence Interval for Mean	Lower Bound	1.6732		
			Upper Bound	1.9369		
		5% Trimmed Mean	1.8076		
		Median	1.7638		
		Variance	.116		
		Std. Deviation	.34008		
		Minimum	.98		
		Maximum	2.64		
		Range	1.66		
		Interquartile Range	.37		
		Skewness	-.117	.441	
		Kurtosis	.955	.858	
	2.00	Mean	1.7262	.07060	
		95% Confidence Interval for Mean	Lower Bound	1.5808		
			Upper Bound	1.8716		
		5% Trimmed Mean	1.7210		
		Median	1.7170		
		Variance	.130		
		Std. Deviation	.35998		
		Minimum	1.14		
		Maximum	2.43		
		Range	1.29		
		Interquartile Range	.56		
		Skewness	.162	.456	
		Kurtosis	-.711	.887	
VO2_85_W12	1.00	Mean	1.9186	.06869	
		95% Confidence Interval for Mean	Lower Bound	1.7776		
			Upper Bound	2.0595		
		5% Trimmed Mean	1.9076		
		Median	1.8828		
		Variance	.132		
		Std. Deviation	.36347		
		Minimum	1.23		
		Maximum	2.76		
		Range	1.53		
		Interquartile Range	.49		
		Skewness	.550	.441	
		Kurtosis	.368	.858	
	2.00	Mean	1.8422	.07481	
		95% Confidence Interval for Mean	Lower Bound	1.6881		
			Upper Bound	1.9963		
		5% Trimmed Mean	1.8366		
		Median	1.8445		
		Variance	.145		
		Std. Deviation	.38143		
		Minimum	1.16		
		Maximum	2.62		
		Range	1.46		
		Interquartile Range	.53		
		Skewness	.284	.456	
		Kurtosis	-.353	.887	
VO2_85_Diff_W6	1.00	Mean	-.0179	.04399	
		95% Confidence Interval for Mean	Lower Bound	-.1082		
			Upper Bound	.0723		
		5% Trimmed Mean	-.0237		
		Median	-.0680		
		Variance	.054		
		Std. Deviation	.23276		
		Minimum	-.48		
		Maximum	.64		
		Range	1.12		
		Interquartile Range	.28		
		Skewness	.493	.441	
		Kurtosis	1.209	.858	
	2.00	Mean	.0758	.04698	
		95% Confidence Interval for Mean	Lower Bound	-.0209		
			Upper Bound	.1726		
		5% Trimmed Mean	.0699		
		Median	.0680		
		Variance	.057		
		Std. Deviation	.23954		
		Minimum	-.44		
		Maximum	.66		
		Range	1.11		
		Interquartile Range	.27		
		Skewness	.511	.456	
		Kurtosis	.906	.887	
VO2_85_Diff_W12	1.00	Mean	-.1314	.05858	
		95% Confidence Interval for Mean	Lower Bound	-.2516		
			Upper Bound	-.0113		
		5% Trimmed Mean	-.1237		
		Median	-.0935		
		Variance	.096		
		Std. Deviation	.30997		
		Minimum	-.98		
		Maximum	.52		
		Range	1.50		
		Interquartile Range	.39		
		Skewness	-.368	.441	
		Kurtosis	1.006	.858	
	2.00	Mean	-.0402	.07483	
		95% Confidence Interval for Mean	Lower Bound	-.1943		
			Upper Bound	.1139		
		5% Trimmed Mean	-.0386		
		Median	-.0510		
		Variance	.146		
		Std. Deviation	.38158		
		Minimum	-.87		
		Maximum	.78		
		Range	1.65		
		Interquartile Range	.35		
		Skewness	-.298	.456	
		Kurtosis	.869	.887	
Exhaust_B	1.00	Mean	8.0679	.22510	
		95% Confidence Interval for Mean	Lower Bound	7.6060		
			Upper Bound	8.5297		
		5% Trimmed Mean	8.0592		
		Median	8.0000		
		Variance	1.419		
		Std. Deviation	1.19110		
		Minimum	6.00		
		Maximum	10.28		
		Range	4.28		
		Interquartile Range	1.96		
		Skewness	.225	.441	
		Kurtosis	-.873	.858	
	2.00	Mean	7.7608	.21505	
		95% Confidence Interval for Mean	Lower Bound	7.3179		
			Upper Bound	8.2037		
		5% Trimmed Mean	7.6982		
		Median	8.0000		
		Variance	1.202		
		Std. Deviation	1.09653		
		Minimum	6.02		
		Maximum	11.00		
		Range	4.98		
		Interquartile Range	1.15		
		Skewness	.849	.456	
		Kurtosis	1.815	.887	
Exhaust_W6	1.00	Mean	8.8511	.29001	
		95% Confidence Interval for Mean	Lower Bound	8.2560		
			Upper Bound	9.4461		
		5% Trimmed Mean	8.8660		
		Median	9.0200		
		Variance	2.355		
		Std. Deviation	1.53461		
		Minimum	5.50		
		Maximum	12.00		
		Range	6.50		
		Interquartile Range	2.04		
		Skewness	-.104	.441	
		Kurtosis	-.140	.858	
	2.00	Mean	8.1250	.22710	
		95% Confidence Interval for Mean	Lower Bound	7.6573		
			Upper Bound	8.5927		
		5% Trimmed Mean	8.0826		
		Median	8.0500		
		Variance	1.341		
		Std. Deviation	1.15800		
		Minimum	6.05		
		Maximum	11.20		
		Range	5.15		
		Interquartile Range	1.54		
		Skewness	.566	.456	
		Kurtosis	.978	.887	
Exhaust_W12	1.00	Mean	10.5514	.26754	
		95% Confidence Interval for Mean	Lower Bound	10.0025		
			Upper Bound	11.1004		
		5% Trimmed Mean	10.5929		
		Median	10.7000		
		Variance	2.004		
		Std. Deviation	1.41571		
		Minimum	7.07		
		Maximum	13.00		
		Range	5.93		
		Interquartile Range	1.24		
		Skewness	-.281	.441	
		Kurtosis	.261	.858	
	2.00	Mean	8.6792	.23910	
		95% Confidence Interval for Mean	Lower Bound	8.1868		
			Upper Bound	9.1717		
		5% Trimmed Mean	8.6523		
		Median	8.4350		
		Variance	1.486		
		Std. Deviation	1.21915		
		Minimum	6.12		
		Maximum	11.80		
		Range	5.68		
		Interquartile Range	1.18		
		Skewness	.444	.456	
		Kurtosis	.772	.887	
Exhaust_Diff_W6	1.00	Mean	-.7832	.26906	
		95% Confidence Interval for Mean	Lower Bound	-1.3353		
			Upper Bound	-.2311		
		5% Trimmed Mean	-.8540		
		Median	-1.0550		
		Variance	2.027		
		Std. Deviation	1.42374		
		Minimum	-3.13		
		Maximum	2.96		
		Range	6.09		
		Interquartile Range	1.53		
		Skewness	.993	.441	
		Kurtosis	1.243	.858	
	2.00	Mean	-.3642	.07186	
		95% Confidence Interval for Mean	Lower Bound	-.5122		
			Upper Bound	-.2162		
		5% Trimmed Mean	-.3411		
		Median	-.2150		
		Variance	.134		
		Std. Deviation	.36642		
		Minimum	-1.15		
		Maximum	-.02		
		Range	1.13		
		Interquartile Range	.71		
		Skewness	-.780	.456	
		Kurtosis	-.815	.887	
Exhaust_Diff_W12	1.00	Mean	-2.4836	.31355	
		95% Confidence Interval for Mean	Lower Bound	-3.1269		
			Upper Bound	-1.8402		
		5% Trimmed Mean	-2.6084		
		Median	-2.8850		
		Variance	2.753		
		Std. Deviation	1.65913		
		Minimum	-4.93		
		Maximum	2.95		
		Range	7.88		
		Interquartile Range	2.31		
		Skewness	1.270	.441	
		Kurtosis	2.831	.858	
	2.00	Mean	-.9185	.08629	
		95% Confidence Interval for Mean	Lower Bound	-1.0962		
			Upper Bound	-.7408		
		5% Trimmed Mean	-.9158		
		Median	-.9750		
		Variance	.194		
		Std. Deviation	.43997		
		Minimum	-1.88		
		Maximum	-.07		
		Range	1.81		
		Interquartile Range	.45		
		Skewness	.442	.456	
		Kurtosis	.474	.887	
