## Supplementary material for "Effects of a standardised *Amaranthus tricolor*–*Spinacia oleracea* extract on exercise performance, muscle strength, endurance and recovery in physically active adults: a randomised, double-blind, placebo-controlled trial": Raw statistical outputs: DOMS_Output_17.07.2026.doc

Explore


Notes	
Output Created	17-JUL-2026 11:54:43	
Comments		
Input	Active Dataset	DataSet0	
	Filter	<none>	
	Weight	<none>	
	Split File	<none>	
	N of Rows in Working Data File	64	
Missing Value Handling	Definition of Missing	User-defined missing values for dependent variables are treated as missing.	
	Cases Used	Statistics are based on cases with no missing values for any dependent variable or factor used.	
Syntax	EXAMINE VARIABLES=Soreness_Upper_Body_B Soreness_Upper_Body_W6 Soreness_Upper_Body_W12
    Soreness_Upper_Body_Diff_W6 Soreness_Upper_Body_Diff_W12 Pain_Upper_Body_B Pain_Upper_Body_W6
    Pain_Upper_Body_W12 Pain_Upper_Body_Diff_W6 Pain_Upper_Body_Diff_W12 Soreness_Lower_Body_B
    Soreness_Lower_Body_W6 Soreness_Lower_Body_W12 Soreness_Lower_Body_Diff_W6
    Soreness_Lower_Body_Diff_W12 Pain_Lower_Body_B Pain_Lower_Body_W6 Pain_Lower_Body_W12
    Pain_Lower_Body_Diff_W6 Pain_Lower_Body_Diff_W12 BY Group
  /PLOT BOXPLOT STEMLEAF
  /COMPARE GROUPS
  /STATISTICS DESCRIPTIVES
  /CINTERVAL 95
  /MISSING LISTWISE
  /NOTOTAL.	
Resources	Processor Time	00:00:01.62	
	Elapsed Time	00:00:01.57	


Group


Case Processing Summary	
	Group	Cases	
		Valid	Missing	Total	
		N	Percent	N	Percent	N	Percent	
Soreness_Upper_Body_B	1.00	32	100.0%	0	0.0%	32	100.0%	
	2.00	32	100.0%	0	0.0%	32	100.0%	
Soreness_Upper_Body_W6	1.00	32	100.0%	0	0.0%	32	100.0%	
	2.00	32	100.0%	0	0.0%	32	100.0%	
Soreness_Upper_Body_W12	1.00	32	100.0%	0	0.0%	32	100.0%	
	2.00	32	100.0%	0	0.0%	32	100.0%	
Soreness_Upper_Body_Diff_W6	1.00	32	100.0%	0	0.0%	32	100.0%	
	2.00	32	100.0%	0	0.0%	32	100.0%	
Soreness_Upper_Body_Diff_W12	1.00	32	100.0%	0	0.0%	32	100.0%	
	2.00	32	100.0%	0	0.0%	32	100.0%	
Pain_Upper_Body_B	1.00	32	100.0%	0	0.0%	32	100.0%	
	2.00	32	100.0%	0	0.0%	32	100.0%	
Pain_Upper_Body_W6	1.00	32	100.0%	0	0.0%	32	100.0%	
	2.00	32	100.0%	0	0.0%	32	100.0%	
Pain_Upper_Body_W12	1.00	32	100.0%	0	0.0%	32	100.0%	
	2.00	32	100.0%	0	0.0%	32	100.0%	
Pain_Upper_Body_Diff_W6	1.00	32	100.0%	0	0.0%	32	100.0%	
	2.00	32	100.0%	0	0.0%	32	100.0%	
Pain_Upper_Body_Diff_W12	1.00	32	100.0%	0	0.0%	32	100.0%	
	2.00	32	100.0%	0	0.0%	32	100.0%	
Soreness_Lower_Body_B	1.00	32	100.0%	0	0.0%	32	100.0%	
	2.00	32	100.0%	0	0.0%	32	100.0%	
Soreness_Lower_Body_W6	1.00	32	100.0%	0	0.0%	32	100.0%	
	2.00	32	100.0%	0	0.0%	32	100.0%	
Soreness_Lower_Body_W12	1.00	32	100.0%	0	0.0%	32	100.0%	
	2.00	32	100.0%	0	0.0%	32	100.0%	
Soreness_Lower_Body_Diff_W6	1.00	32	100.0%	0	0.0%	32	100.0%	
	2.00	32	100.0%	0	0.0%	32	100.0%	
Soreness_Lower_Body_Diff_W12	1.00	32	100.0%	0	0.0%	32	100.0%	
	2.00	32	100.0%	0	0.0%	32	100.0%	
Pain_Lower_Body_B	1.00	32	100.0%	0	0.0%	32	100.0%	
	2.00	32	100.0%	0	0.0%	32	100.0%	
Pain_Lower_Body_W6	1.00	32	100.0%	0	0.0%	32	100.0%	
	2.00	32	100.0%	0	0.0%	32	100.0%	
Pain_Lower_Body_W12	1.00	32	100.0%	0	0.0%	32	100.0%	
	2.00	32	100.0%	0	0.0%	32	100.0%	
Pain_Lower_Body_Diff_W6	1.00	32	100.0%	0	0.0%	32	100.0%	
	2.00	32	100.0%	0	0.0%	32	100.0%	
Pain_Lower_Body_Diff_W12	1.00	32	100.0%	0	0.0%	32	100.0%	
	2.00	32	100.0%	0	0.0%	32	100.0%	


Descriptives	
	Group	Statistic	Std. Error	
Soreness_Upper_Body_B	1.00	Mean	5.7188	.15050	
		95% Confidence Interval for Mean	Lower Bound	5.4118		
			Upper Bound	6.0257		
		5% Trimmed Mean	5.7431		
		Median	6.0000		
		Variance	.725		
		Std. Deviation	.85135		
		Minimum	4.00		
		Maximum	7.00		
		Range	3.00		
		Interquartile Range	1.00		
		Skewness	-.074	.414	
		Kurtosis	-.583	.809	
	2.00	Mean	5.6563	.13175	
		95% Confidence Interval for Mean	Lower Bound	5.3875		
			Upper Bound	5.9250		
		5% Trimmed Mean	5.6528		
		Median	6.0000		
		Variance	.555		
		Std. Deviation	.74528		
		Minimum	4.00		
		Maximum	7.00		
		Range	3.00		
		Interquartile Range	1.00		
		Skewness	.171	.414	
		Kurtosis	-.373	.809	
Soreness_Upper_Body_W6	1.00	Mean	4.6875	.12245	
		95% Confidence Interval for Mean	Lower Bound	4.4378		
			Upper Bound	4.9372		
		5% Trimmed Mean	4.6528		
		Median	5.0000		
		Variance	.480		
		Std. Deviation	.69270		
		Minimum	4.00		
		Maximum	6.00		
		Range	2.00		
		Interquartile Range	1.00		
		Skewness	.510	.414	
		Kurtosis	-.741	.809	
	2.00	Mean	5.0000	.12700	
		95% Confidence Interval for Mean	Lower Bound	4.7410		
			Upper Bound	5.2590		
		5% Trimmed Mean	5.0000		
		Median	5.0000		
		Variance	.516		
		Std. Deviation	.71842		
		Minimum	4.00		
		Maximum	6.00		
		Range	2.00		
		Interquartile Range	1.50		
		Skewness	.000	.414	
		Kurtosis	-.962	.809	
Soreness_Upper_Body_W12	1.00	Mean	3.2500	.15554	
		95% Confidence Interval for Mean	Lower Bound	2.9328		
			Upper Bound	3.5672		
		5% Trimmed Mean	3.2222		
		Median	3.0000		
		Variance	.774		
		Std. Deviation	.87988		
		Minimum	2.00		
		Maximum	5.00		
		Range	3.00		
		Interquartile Range	1.00		
		Skewness	.379	.414	
		Kurtosis	-.350	.809	
	2.00	Mean	4.3750	.10767	
		95% Confidence Interval for Mean	Lower Bound	4.1554		
			Upper Bound	4.5946		
		5% Trimmed Mean	4.4167		
		Median	4.0000		
		Variance	.371		
		Std. Deviation	.60907		
		Minimum	3.00		
		Maximum	5.00		
		Range	2.00		
		Interquartile Range	1.00		
		Skewness	-.400	.414	
		Kurtosis	-.582	.809	
Soreness_Upper_Body_Diff_W6	1.00	Mean	1.0313	.13079	
		95% Confidence Interval for Mean	Lower Bound	.7645		
			Upper Bound	1.2980		
		5% Trimmed Mean	1.0000		
		Median	1.0000		
		Variance	.547		
		Std. Deviation	.73985		
		Minimum	.00		
		Maximum	3.00		
		Range	3.00		
		Interquartile Range	.00		
		Skewness	.460	.414	
		Kurtosis	.375	.809	
	2.00	Mean	.6563	.15299	
		95% Confidence Interval for Mean	Lower Bound	.3442		
			Upper Bound	.9683		
		5% Trimmed Mean	.6389		
		Median	1.0000		
		Variance	.749		
		Std. Deviation	.86544		
		Minimum	-1.00		
		Maximum	3.00		
		Range	4.00		
		Interquartile Range	1.00		
		Skewness	.438	.414	
		Kurtosis	.694	.809	
Soreness_Upper_Body_Diff_W12	1.00	Mean	2.4688	.18505	
		95% Confidence Interval for Mean	Lower Bound	2.0913		
			Upper Bound	2.8462		
		5% Trimmed Mean	2.4306		
		Median	2.0000		
		Variance	1.096		
		Std. Deviation	1.04679		
		Minimum	1.00		
		Maximum	5.00		
		Range	4.00		
		Interquartile Range	1.00		
		Skewness	.358	.414	
		Kurtosis	-.304	.809	
	2.00	Mean	1.2813	.17526	
		95% Confidence Interval for Mean	Lower Bound	.9238		
			Upper Bound	1.6387		
		5% Trimmed Mean	1.2569		
		Median	1.0000		
		Variance	.983		
		Std. Deviation	.99139		
		Minimum	-1.00		
		Maximum	4.00		
		Range	5.00		
		Interquartile Range	1.00		
		Skewness	.232	.414	
		Kurtosis	1.021	.809	
Pain_Upper_Body_B	1.00	Mean	2.6875	.10470	
		95% Confidence Interval for Mean	Lower Bound	2.4740		
			Upper Bound	2.9010		
		5% Trimmed Mean	2.6528		
		Median	3.0000		
		Variance	.351		
		Std. Deviation	.59229		
		Minimum	2.00		
		Maximum	4.00		
		Range	2.00		
		Interquartile Range	1.00		
		Skewness	.194	.414	
		Kurtosis	-.518	.809	
	2.00	Mean	2.6250	.10767	
		95% Confidence Interval for Mean	Lower Bound	2.4054		
			Upper Bound	2.8446		
		5% Trimmed Mean	2.5833		
		Median	3.0000		
		Variance	.371		
		Std. Deviation	.60907		
		Minimum	2.00		
		Maximum	4.00		
		Range	2.00		
		Interquartile Range	1.00		
		Skewness	.400	.414	
		Kurtosis	-.582	.809	
Pain_Upper_Body_W6	1.00	Mean	1.9688	.07077	
		95% Confidence Interval for Mean	Lower Bound	1.8244		
			Upper Bound	2.1131		
		5% Trimmed Mean	1.9653		
		Median	2.0000		
		Variance	.160		
		Std. Deviation	.40035		
		Minimum	1.00		
		Maximum	3.00		
		Range	2.00		
		Interquartile Range	.00		
		Skewness	-.286	.414	
		Kurtosis	4.161	.809	
	2.00	Mean	2.2188	.10752	
		95% Confidence Interval for Mean	Lower Bound	1.9995		
			Upper Bound	2.4380		
		5% Trimmed Mean	2.2083		
		Median	2.0000		
		Variance	.370		
		Std. Deviation	.60824		
		Minimum	1.00		
		Maximum	4.00		
		Range	3.00		
		Interquartile Range	.75		
		Skewness	.786	.414	
		Kurtosis	1.589	.809	
Pain_Upper_Body_ _W12	1.00	Mean	1.4063	.08821	
		95% Confidence Interval for Mean	Lower Bound	1.2263		
			Upper Bound	1.5862		
		5% Trimmed Mean	1.3958		
		Median	1.0000		
		Variance	.249		
		Std. Deviation	.49899		
		Minimum	1.00		
		Maximum	2.00		
		Range	1.00		
		Interquartile Range	1.00		
		Skewness	.401	.414	
		Kurtosis	-1.967	.809	
	2.00	Mean	1.8750	.10767	
		95% Confidence Interval for Mean	Lower Bound	1.6554		
			Upper Bound	2.0946		
		5% Trimmed Mean	1.8611		
		Median	2.0000		
		Variance	.371		
		Std. Deviation	.60907		
		Minimum	1.00		
		Maximum	3.00		
		Range	2.00		
		Interquartile Range	.75		
		Skewness	.057	.414	
		Kurtosis	-.155	.809	
Pain_Upper_Body_Diff_W6	1.00	Mean	.7188	.10273	
		95% Confidence Interval for Mean	Lower Bound	.5092		
			Upper Bound	.9283		
		5% Trimmed Mean	.7431		
		Median	1.0000		
		Variance	.338		
		Std. Deviation	.58112		
		Minimum	-1.00		
		Maximum	2.00		
		Range	3.00		
		Interquartile Range	1.00		
		Skewness	-.956	.414	
		Kurtosis	1.388	.809	
	2.00	Mean	.4063	.10869	
		95% Confidence Interval for Mean	Lower Bound	.1846		
			Upper Bound	.6279		
		5% Trimmed Mean	.4514		
		Median	.0000		
		Variance	.378		
		Std. Deviation	.61484		
		Minimum	-1.00		
		Maximum	1.00		
		Range	2.00		
		Interquartile Range	1.00		
		Skewness	-.507	.414	
		Kurtosis	-.548	.809	
Pain_Upper_Body_Diff_W12	1.00	Mean	1.2813	.12885	
		95% Confidence Interval for Mean	Lower Bound	1.0185		
			Upper Bound	1.5440		
		5% Trimmed Mean	1.2778		
		Median	1.0000		
		Variance	.531		
		Std. Deviation	.72887		
		Minimum	.00		
		Maximum	3.00		
		Range	3.00		
		Interquartile Range	1.00		
		Skewness	.035	.414	
		Kurtosis	-.186	.809	
	2.00	Mean	.7500	.10040	
		95% Confidence Interval for Mean	Lower Bound	.5452		
			Upper Bound	.9548		
		5% Trimmed Mean	.7222		
		Median	1.0000		
		Variance	.323		
		Std. Deviation	.56796		
		Minimum	.00		
		Maximum	2.00		
		Range	2.00		
		Interquartile Range	1.00		
		Skewness	.000	.414	
		Kurtosis	-.257	.809	
Soreness_Lower_Body_B	1.00	Mean	5.8125	.15185	
		95% Confidence Interval for Mean	Lower Bound	5.5028		
			Upper Bound	6.1222		
		5% Trimmed Mean	5.8264		
		Median	6.0000		
		Variance	.738		
		Std. Deviation	.85901		
		Minimum	4.00		
		Maximum	7.00		
		Range	3.00		
		Interquartile Range	1.75		
		Skewness	.059	.414	
		Kurtosis	-1.017	.809	
	2.00	Mean	5.6250	.12500	
		95% Confidence Interval for Mean	Lower Bound	5.3701		
			Upper Bound	5.8799		
		5% Trimmed Mean	5.6181		
		Median	6.0000		
		Variance	.500		
		Std. Deviation	.70711		
		Minimum	4.00		
		Maximum	7.00		
		Range	3.00		
		Interquartile Range	1.00		
		Skewness	.109	.414	
		Kurtosis	-.185	.809	
Soreness_Lower_Body_W6	1.00	Mean	4.6875	.14506	
		95% Confidence Interval for Mean	Lower Bound	4.3916		
			Upper Bound	4.9834		
		5% Trimmed Mean	4.6181		
		Median	4.5000		
		Variance	.673		
		Std. Deviation	.82060		
		Minimum	4.00		
		Maximum	7.00		
		Range	3.00		
		Interquartile Range	1.00		
		Skewness	1.030	.414	
		Kurtosis	.498	.809	
	2.00	Mean	5.0625	.15514	
		95% Confidence Interval for Mean	Lower Bound	4.7461		
			Upper Bound	5.3789		
		5% Trimmed Mean	5.0139		
		Median	5.0000		
		Variance	.770		
		Std. Deviation	.87759		
		Minimum	4.00		
		Maximum	7.00		
		Range	3.00		
		Interquartile Range	.75		
		Skewness	.790	.414	
		Kurtosis	.354	.809	
Soreness_Lower_Body_W12	1.00	Mean	3.4375	.15514	
		95% Confidence Interval for Mean	Lower Bound	3.1211		
			Upper Bound	3.7539		
		5% Trimmed Mean	3.4306		
		Median	3.0000		
		Variance	.770		
		Std. Deviation	.87759		
		Minimum	2.00		
		Maximum	5.00		
		Range	3.00		
		Interquartile Range	1.00		
		Skewness	.203	.414	
		Kurtosis	-.513	.809	
	2.00	Mean	4.8125	.13043	
		95% Confidence Interval for Mean	Lower Bound	4.5465		
			Upper Bound	5.0785		
		5% Trimmed Mean	4.7917		
		Median	5.0000		
		Variance	.544		
		Std. Deviation	.73780		
		Minimum	4.00		
		Maximum	6.00		
		Range	2.00		
		Interquartile Range	1.00		
		Skewness	.317	.414	
		Kurtosis	-1.044	.809	
Soreness_Lower_Body_Diff_W6	1.00	Mean	1.1250	.16033	
		95% Confidence Interval for Mean	Lower Bound	.7980		
			Upper Bound	1.4520		
		5% Trimmed Mean	1.1181		
		Median	1.0000		
		Variance	.823		
		Std. Deviation	.90696		
		Minimum	-1.00		
		Maximum	3.00		
		Range	4.00		
		Interquartile Range	.75		
		Skewness	.294	.414	
		Kurtosis	.645	.809	
	2.00	Mean	.5625	.14850	
		95% Confidence Interval for Mean	Lower Bound	.2596		
			Upper Bound	.8654		
		5% Trimmed Mean	.5347		
		Median	1.0000		
		Variance	.706		
		Std. Deviation	.84003		
		Minimum	-1.00		
		Maximum	3.00		
		Range	4.00		
		Interquartile Range	1.00		
		Skewness	.313	.414	
		Kurtosis	1.337	.809	
Soreness_Lower_Body_Diff_W12	1.00	Mean	2.3750	.19955	
		95% Confidence Interval for Mean	Lower Bound	1.9680		
			Upper Bound	2.7820		
		5% Trimmed Mean	2.3958		
		Median	2.5000		
		Variance	1.274		
		Std. Deviation	1.12880		
		Minimum	-1.00		
		Maximum	5.00		
		Range	6.00		
		Interquartile Range	1.00		
		Skewness	-.529	.414	
		Kurtosis	1.780	.809	
	2.00	Mean	.8125	.14506	
		95% Confidence Interval for Mean	Lower Bound	.5166		
			Upper Bound	1.1084		
		5% Trimmed Mean	.7917		
		Median	1.0000		
		Variance	.673		
		Std. Deviation	.82060		
		Minimum	-1.00		
		Maximum	3.00		
		Range	4.00		
		Interquartile Range	1.00		
		Skewness	.371	.414	
		Kurtosis	.660	.809	
Pain_Lower_Body_B	1.00	Mean	2.8750	.11666	
		95% Confidence Interval for Mean	Lower Bound	2.6371		
			Upper Bound	3.1129		
		5% Trimmed Mean	2.8611		
		Median	3.0000		
		Variance	.435		
		Std. Deviation	.65991		
		Minimum	2.00		
		Maximum	4.00		
		Range	2.00		
		Interquartile Range	1.00		
		Skewness	.135	.414	
		Kurtosis	-.570	.809	
	2.00	Mean	2.5938	.10869	
		95% Confidence Interval for Mean	Lower Bound	2.3721		
			Upper Bound	2.8154		
		5% Trimmed Mean	2.5486		
		Median	3.0000		
		Variance	.378		
		Std. Deviation	.61484		
		Minimum	2.00		
		Maximum	4.00		
		Range	2.00		
		Interquartile Range	1.00		
		Skewness	.507	.414	
		Kurtosis	-.548	.809	
Pain_Lower_Body_W6	1.00	Mean	2.1250	.08695	
		95% Confidence Interval for Mean	Lower Bound	1.9477		
			Upper Bound	2.3023		
		5% Trimmed Mean	2.1389		
		Median	2.0000		
		Variance	.242		
		Std. Deviation	.49187		
		Minimum	1.00		
		Maximum	3.00		
		Range	2.00		
		Interquartile Range	.00		
		Skewness	.325	.414	
		Kurtosis	1.186	.809	
	2.00	Mean	2.1875	.10470	
		95% Confidence Interval for Mean	Lower Bound	1.9740		
			Upper Bound	2.4010		
		5% Trimmed Mean	2.2083		
		Median	2.0000		
		Variance	.351		
		Std. Deviation	.59229		
		Minimum	1.00		
		Maximum	3.00		
		Range	2.00		
		Interquartile Range	1.00		
		Skewness	-.054	.414	
		Kurtosis	-.160	.809	
Pain_Lower_Body_W12	1.00	Mean	1.3438	.08531	
		95% Confidence Interval for Mean	Lower Bound	1.1698		
			Upper Bound	1.5177		
		5% Trimmed Mean	1.3264		
		Median	1.0000		
		Variance	.233		
		Std. Deviation	.48256		
		Minimum	1.00		
		Maximum	2.00		
		Range	1.00		
		Interquartile Range	1.00		
		Skewness	.691	.414	
		Kurtosis	-1.629	.809	
	2.00	Mean	1.9688	.09508	
		95% Confidence Interval for Mean	Lower Bound	1.7748		
			Upper Bound	2.1627		
		5% Trimmed Mean	1.9653		
		Median	2.0000		
		Variance	.289		
		Std. Deviation	.53788		
		Minimum	1.00		
		Maximum	3.00		
		Range	2.00		
		Interquartile Range	.00		
		Skewness	-.035	.414	
		Kurtosis	.862	.809	
Pain_Lower_Body_Diff_W6	1.00	Mean	.7500	.08980	
		95% Confidence Interval for Mean	Lower Bound	.5668		
			Upper Bound	.9332		
		5% Trimmed Mean	.7431		
		Median	1.0000		
		Variance	.258		
		Std. Deviation	.50800		
		Minimum	.00		
		Maximum	2.00		
		Range	2.00		
		Interquartile Range	1.00		
		Skewness	-.394	.414	
		Kurtosis	-.154	.809	
	2.00	Mean	.4063	.08821	
		95% Confidence Interval for Mean	Lower Bound	.2263		
			Upper Bound	.5862		
		5% Trimmed Mean	.3958		
		Median	.0000		
		Variance	.249		
		Std. Deviation	.49899		
		Minimum	.00		
		Maximum	1.00		
		Range	1.00		
		Interquartile Range	1.00		
		Skewness	.401	.414	
		Kurtosis	-1.967	.809	
Pain_Lower_Body_Diff_W12	1.00	Mean	1.5313	.12688	
		95% Confidence Interval for Mean	Lower Bound	1.2725		
			Upper Bound	1.7900		
		5% Trimmed Mean	1.5347		
		Median	2.0000		
		Variance	.515		
		Std. Deviation	.71772		
		Minimum	.00		
		Maximum	3.00		
		Range	3.00		
		Interquartile Range	1.00		
		Skewness	-.116	.414	
		Kurtosis	-.058	.809	
	2.00	Mean	.6250	.10767	
		95% Confidence Interval for Mean	Lower Bound	.4054		
			Upper Bound	.8446		
		5% Trimmed Mean	.5833		
		Median	1.0000		
		Variance	.371		
		Std. Deviation	.60907		
		Minimum	.00		
		Maximum	2.00		
		Range	2.00		
		Interquartile Range	1.00		
		Skewness	.400	.414	
		Kurtosis	-.582	.809	
