## Supplementary material for "Effects of a standardised *Amaranthus tricolor*–*Spinacia oleracea* extract on exercise performance, muscle strength, endurance and recovery in physically active adults: a randomised, double-blind, placebo-controlled trial": Raw statistical outputs: Physical_Examination_Output_17.07.2026.doc

Explore


Notes	
Output Created	17-JUL-2026 11:40:50	
Comments		
Input	Active Dataset	DataSet0	
	Filter	<none>	
	Weight	<none>	
	Split File	<none>	
	N of Rows in Working Data File	64	
Missing Value Handling	Definition of Missing	User-defined missing values for dependent variables are treated as missing.	
	Cases Used	Statistics are based on cases with no missing values for any dependent variable or factor used.	
Syntax	EXAMINE VARIABLES=SBP_B SBP_W6 SBP_W12 SBP_Diff_W6 SBP_Diff_W12 DBP_B DBP_W6 DBP_W12 DBP_Diff_W6
    DBP_Diff_W12 Heart_Rate_B Heart_Rate_W6 Heart_Rate_W12 Heart_Rate_Diff_W6 Heart_Rate_Diff_W12
    Oxy_Saturatn_B Oxy_Saturatn_W6 Oxy_Saturatn_W12 Oxy_Saturatn_Diff_W6 Oxy_Saturatn_Diff_W12
    Oral_Temp_B Oral_Temp_W6 Oral_Temp_W12 Oral_Temp_Diff_W6 Oral_Temp_Diff_W12 Resp_Rate_B
    Resp_Rate_W6 Resp_Rate_W12 Resp_Rate_Diff_W6 Resp_Rate_Diff_W12 BY Group
  /PLOT BOXPLOT STEMLEAF
  /COMPARE GROUPS
  /STATISTICS DESCRIPTIVES
  /CINTERVAL 95
  /MISSING LISTWISE
  /NOTOTAL.	
Resources	Processor Time	00:00:02.52	
	Elapsed Time	00:00:02.38	


Group


Case Processing Summary	
	Group	Cases	
		Valid	Missing	Total	
		N	Percent	N	Percent	N	Percent	
SBP_B	1.00	32	100.0%	0	0.0%	32	100.0%	
	2.00	32	100.0%	0	0.0%	32	100.0%	
SBP_W6	1.00	32	100.0%	0	0.0%	32	100.0%	
	2.00	32	100.0%	0	0.0%	32	100.0%	
SBP_W12	1.00	32	100.0%	0	0.0%	32	100.0%	
	2.00	32	100.0%	0	0.0%	32	100.0%	
SBP_Diff_W6	1.00	32	100.0%	0	0.0%	32	100.0%	
	2.00	32	100.0%	0	0.0%	32	100.0%	
SBP_Diff_W12	1.00	32	100.0%	0	0.0%	32	100.0%	
	2.00	32	100.0%	0	0.0%	32	100.0%	
DBP_B	1.00	32	100.0%	0	0.0%	32	100.0%	
	2.00	32	100.0%	0	0.0%	32	100.0%	
DBP_W6	1.00	32	100.0%	0	0.0%	32	100.0%	
	2.00	32	100.0%	0	0.0%	32	100.0%	
DBP_W12	1.00	32	100.0%	0	0.0%	32	100.0%	
	2.00	32	100.0%	0	0.0%	32	100.0%	
DBP_Diff_W6	1.00	32	100.0%	0	0.0%	32	100.0%	
	2.00	32	100.0%	0	0.0%	32	100.0%	
DBP_Diff_W12	1.00	32	100.0%	0	0.0%	32	100.0%	
	2.00	32	100.0%	0	0.0%	32	100.0%	
Heart_Rate_B	1.00	32	100.0%	0	0.0%	32	100.0%	
	2.00	32	100.0%	0	0.0%	32	100.0%	
Heart_Rate_W6	1.00	32	100.0%	0	0.0%	32	100.0%	
	2.00	32	100.0%	0	0.0%	32	100.0%	
Heart_Rate_W12	1.00	32	100.0%	0	0.0%	32	100.0%	
	2.00	32	100.0%	0	0.0%	32	100.0%	
Heart_Rate_Diff_W6	1.00	32	100.0%	0	0.0%	32	100.0%	
	2.00	32	100.0%	0	0.0%	32	100.0%	
Heart_Rate_Diff_W12	1.00	32	100.0%	0	0.0%	32	100.0%	
	2.00	32	100.0%	0	0.0%	32	100.0%	
Oxy_Saturatn_B	1.00	32	100.0%	0	0.0%	32	100.0%	
	2.00	32	100.0%	0	0.0%	32	100.0%	
Oxy_Saturatn_W6	1.00	32	100.0%	0	0.0%	32	100.0%	
	2.00	32	100.0%	0	0.0%	32	100.0%	
Oxy_Saturatn_W12	1.00	32	100.0%	0	0.0%	32	100.0%	
	2.00	32	100.0%	0	0.0%	32	100.0%	
Oxy_Saturatn_Diff_W6	1.00	32	100.0%	0	0.0%	32	100.0%	
	2.00	32	100.0%	0	0.0%	32	100.0%	
Oxy_Saturatn_Diff_W12	1.00	32	100.0%	0	0.0%	32	100.0%	
	2.00	32	100.0%	0	0.0%	32	100.0%	
Oral_Temp_B	1.00	32	100.0%	0	0.0%	32	100.0%	
	2.00	32	100.0%	0	0.0%	32	100.0%	
Oral_Temp_W6	1.00	32	100.0%	0	0.0%	32	100.0%	
	2.00	32	100.0%	0	0.0%	32	100.0%	
Oral_Temp_W12	1.00	32	100.0%	0	0.0%	32	100.0%	
	2.00	32	100.0%	0	0.0%	32	100.0%	
Oral_Temp_Diff_W6	1.00	32	100.0%	0	0.0%	32	100.0%	
	2.00	32	100.0%	0	0.0%	32	100.0%	
Oral_Temp_Diff_W12	1.00	32	100.0%	0	0.0%	32	100.0%	
	2.00	32	100.0%	0	0.0%	32	100.0%	
Resp_Rate_B	1.00	32	100.0%	0	0.0%	32	100.0%	
	2.00	32	100.0%	0	0.0%	32	100.0%	
Resp_Rate_W6	1.00	32	100.0%	0	0.0%	32	100.0%	
	2.00	32	100.0%	0	0.0%	32	100.0%	
Resp_Rate_W12	1.00	32	100.0%	0	0.0%	32	100.0%	
	2.00	32	100.0%	0	0.0%	32	100.0%	
Resp_Rate_Diff_W6	1.00	32	100.0%	0	0.0%	32	100.0%	
	2.00	32	100.0%	0	0.0%	32	100.0%	
Resp_Rate_Diff_W12	1.00	32	100.0%	0	0.0%	32	100.0%	
	2.00	32	100.0%	0	0.0%	32	100.0%	


Descriptives	
	Group	Statistic	Std. Error	
SBP_B	1.00	Mean	119.9594	1.08631	
		95% Confidence Interval for Mean	Lower Bound	117.7438		
			Upper Bound	122.1749		
		5% Trimmed Mean	120.4201		
		Median	120.5000		
		Variance	37.762		
		Std. Deviation	6.14512		
		Minimum	100.00		
		Maximum	129.00		
		Range	29.00		
		Interquartile Range	7.50		
		Skewness	-1.241	.414	
		Kurtosis	2.388	.809	
	2.00	Mean	122.4063	.56236	
		95% Confidence Interval for Mean	Lower Bound	121.2593		
			Upper Bound	123.5532		
		5% Trimmed Mean	122.5069		
		Median	123.0000		
		Variance	10.120		
		Std. Deviation	3.18119		
		Minimum	115.00		
		Maximum	128.00		
		Range	13.00		
		Interquartile Range	5.00		
		Skewness	-.378	.414	
		Kurtosis	-.655	.809	
SBP_W6	1.00	Mean	122.4063	.79674	
		95% Confidence Interval for Mean	Lower Bound	120.7813		
			Upper Bound	124.0312		
		5% Trimmed Mean	122.5417		
		Median	123.0000		
		Variance	20.314		
		Std. Deviation	4.50705		
		Minimum	112.00		
		Maximum	130.00		
		Range	18.00		
		Interquartile Range	6.50		
		Skewness	-.302	.414	
		Kurtosis	-.442	.809	
	2.00	Mean	122.5313	.64129	
		95% Confidence Interval for Mean	Lower Bound	121.2233		
			Upper Bound	123.8392		
		5% Trimmed Mean	122.5556		
		Median	122.0000		
		Variance	13.160		
		Std. Deviation	3.62771		
		Minimum	116.00		
		Maximum	129.00		
		Range	13.00		
		Interquartile Range	6.00		
		Skewness	-.118	.414	
		Kurtosis	-.880	.809	
SBP_W12	1.00	Mean	121.3906	.76224	
		95% Confidence Interval for Mean	Lower Bound	119.8360		
			Upper Bound	122.9452		
		5% Trimmed Mean	121.4722		
		Median	122.0000		
		Variance	18.592		
		Std. Deviation	4.31190		
		Minimum	113.00		
		Maximum	128.50		
		Range	15.50		
		Interquartile Range	6.00		
		Skewness	-.102	.414	
		Kurtosis	-.714	.809	
	2.00	Mean	120.8125	.58101	
		95% Confidence Interval for Mean	Lower Bound	119.6275		
			Upper Bound	121.9975		
		5% Trimmed Mean	120.7778		
		Median	120.5000		
		Variance	10.802		
		Std. Deviation	3.28670		
		Minimum	114.00		
		Maximum	129.00		
		Range	15.00		
		Interquartile Range	5.75		
		Skewness	.142	.414	
		Kurtosis	.025	.809	
SBP_Diff_W6	1.00	Mean	-2.4469	1.25157	
		95% Confidence Interval for Mean	Lower Bound	-4.9995		
			Upper Bound	.1057		
		5% Trimmed Mean	-1.5590		
		Median	.0000		
		Variance	50.126		
		Std. Deviation	7.07996		
		Minimum	-30.00		
		Maximum	5.00		
		Range	35.00		
		Interquartile Range	5.00		
		Skewness	-2.489	.414	
		Kurtosis	7.259	.809	
	2.00	Mean	-.1250	.51929	
		95% Confidence Interval for Mean	Lower Bound	-1.1841		
			Upper Bound	.9341		
		5% Trimmed Mean	.0347		
		Median	.0000		
		Variance	8.629		
		Std. Deviation	2.93752		
		Minimum	-10.00		
		Maximum	5.00		
		Range	15.00		
		Interquartile Range	3.00		
		Skewness	-1.083	.414	
		Kurtosis	3.110	.809	
SBP_Diff_W12	1.00	Mean	-1.4313	1.21930	
		95% Confidence Interval for Mean	Lower Bound	-3.9180		
			Upper Bound	1.0555		
		5% Trimmed Mean	-.6319		
		Median	.0000		
		Variance	47.574		
		Std. Deviation	6.89738		
		Minimum	-28.00		
		Maximum	7.00		
		Range	35.00		
		Interquartile Range	5.00		
		Skewness	-2.435	.414	
		Kurtosis	7.061	.809	
	2.00	Mean	1.5938	.50772	
		95% Confidence Interval for Mean	Lower Bound	.5582		
			Upper Bound	2.6293		
		5% Trimmed Mean	1.9028		
		Median	2.0000		
		Variance	8.249		
		Std. Deviation	2.87211		
		Minimum	-10.00		
		Maximum	5.00		
		Range	15.00		
		Interquartile Range	2.00		
		Skewness	-2.160	.414	
		Kurtosis	7.839	.809	
DBP_B	1.00	Mean	74.9031	.92594	
		95% Confidence Interval for Mean	Lower Bound	73.0147		
			Upper Bound	76.7916		
		5% Trimmed Mean	75.0451		
		Median	76.0000		
		Variance	27.436		
		Std. Deviation	5.23792		
		Minimum	60.00		
		Maximum	85.00		
		Range	25.00		
		Interquartile Range	5.88		
		Skewness	-.411	.414	
		Kurtosis	1.275	.809	
	2.00	Mean	77.3125	.76785	
		95% Confidence Interval for Mean	Lower Bound	75.7465		
			Upper Bound	78.8785		
		5% Trimmed Mean	77.3958		
		Median	78.0000		
		Variance	18.867		
		Std. Deviation	4.34361		
		Minimum	67.00		
		Maximum	85.00		
		Range	18.00		
		Interquartile Range	6.00		
		Skewness	-.352	.414	
		Kurtosis	-.133	.809	
DBP_W6	1.00	Mean	76.9375	.79810	
		95% Confidence Interval for Mean	Lower Bound	75.3098		
			Upper Bound	78.5652		
		5% Trimmed Mean	76.8958		
		Median	76.5000		
		Variance	20.383		
		Std. Deviation	4.51476		
		Minimum	70.00		
		Maximum	85.00		
		Range	15.00		
		Interquartile Range	5.75		
		Skewness	.311	.414	
		Kurtosis	-.842	.809	
	2.00	Mean	78.1250	.82397	
		95% Confidence Interval for Mean	Lower Bound	76.4445		
			Upper Bound	79.8055		
		5% Trimmed Mean	78.1736		
		Median	78.0000		
		Variance	21.726		
		Std. Deviation	4.66109		
		Minimum	70.00		
		Maximum	85.00		
		Range	15.00		
		Interquartile Range	8.00		
		Skewness	.054	.414	
		Kurtosis	-1.223	.809	
DBP_W12	1.00	Mean	75.9063	.62919	
		95% Confidence Interval for Mean	Lower Bound	74.6230		
			Upper Bound	77.1895		
		5% Trimmed Mean	75.8056		
		Median	76.0000		
		Variance	12.668		
		Std. Deviation	3.55926		
		Minimum	70.00		
		Maximum	84.00		
		Range	14.00		
		Interquartile Range	5.75		
		Skewness	.293	.414	
		Kurtosis	-.518	.809	
	2.00	Mean	77.5938	.70262	
		95% Confidence Interval for Mean	Lower Bound	76.1608		
			Upper Bound	79.0267		
		5% Trimmed Mean	77.4931		
		Median	77.5000		
		Variance	15.797		
		Std. Deviation	3.97459		
		Minimum	72.00		
		Maximum	85.00		
		Range	13.00		
		Interquartile Range	7.75		
		Skewness	.292	.414	
		Kurtosis	-1.066	.809	
DBP_Diff_W6	1.00	Mean	-2.0344	1.22761	
		95% Confidence Interval for Mean	Lower Bound	-4.5381		
			Upper Bound	.4694		
		5% Trimmed Mean	-1.5660		
		Median	-.5000		
		Variance	48.225		
		Std. Deviation	6.94442		
		Minimum	-24.00		
		Maximum	10.00		
		Range	34.00		
		Interquartile Range	6.75		
		Skewness	-1.366	.414	
		Kurtosis	2.507	.809	
	2.00	Mean	-.8125	.90522	
		95% Confidence Interval for Mean	Lower Bound	-2.6587		
			Upper Bound	1.0337		
		5% Trimmed Mean	-.7361		
		Median	-1.0000		
		Variance	26.222		
		Std. Deviation	5.12072		
		Minimum	-13.00		
		Maximum	10.00		
		Range	23.00		
		Interquartile Range	6.50		
		Skewness	-.193	.414	
		Kurtosis	.161	.809	
DBP_Diff_W12	1.00	Mean	-1.0031	1.11831	
		95% Confidence Interval for Mean	Lower Bound	-3.2839		
			Upper Bound	1.2777		
		5% Trimmed Mean	-.6979		
		Median	.0000		
		Variance	40.020		
		Std. Deviation	6.32611		
		Minimum	-22.00		
		Maximum	13.00		
		Range	35.00		
		Interquartile Range	5.00		
		Skewness	-1.087	.414	
		Kurtosis	3.275	.809	
	2.00	Mean	-.2813	.92483	
		95% Confidence Interval for Mean	Lower Bound	-2.1675		
			Upper Bound	1.6050		
		5% Trimmed Mean	-.1458		
		Median	.0000		
		Variance	27.370		
		Std. Deviation	5.23163		
		Minimum	-13.00		
		Maximum	10.00		
		Range	23.00		
		Interquartile Range	5.50		
		Skewness	-.414	.414	
		Kurtosis	.432	.809	
Heart_Rate_B	1.00	Mean	68.5625	1.65401	
		95% Confidence Interval for Mean	Lower Bound	65.1891		
			Upper Bound	71.9359		
		5% Trimmed Mean	68.6597		
		Median	71.0000		
		Variance	87.544		
		Std. Deviation	9.35651		
		Minimum	50.00		
		Maximum	85.00		
		Range	35.00		
		Interquartile Range	16.25		
		Skewness	-.326	.414	
		Kurtosis	-.855	.809	
	2.00	Mean	71.0625	1.71682	
		95% Confidence Interval for Mean	Lower Bound	67.5610		
			Upper Bound	74.5640		
		5% Trimmed Mean	71.4167		
		Median	71.0000		
		Variance	94.319		
		Std. Deviation	9.71177		
		Minimum	50.00		
		Maximum	85.00		
		Range	35.00		
		Interquartile Range	16.25		
		Skewness	-.405	.414	
		Kurtosis	-.696	.809	
Heart_Rate_W6	1.00	Mean	66.1875	1.34287	
		95% Confidence Interval for Mean	Lower Bound	63.4487		
			Upper Bound	68.9263		
		5% Trimmed Mean	66.0556		
		Median	65.5000		
		Variance	57.706		
		Std. Deviation	7.59642		
		Minimum	51.00		
		Maximum	83.00		
		Range	32.00		
		Interquartile Range	9.75		
		Skewness	.325	.414	
		Kurtosis	.113	.809	
	2.00	Mean	66.4688	1.40419	
		95% Confidence Interval for Mean	Lower Bound	63.6049		
			Upper Bound	69.3326		
		5% Trimmed Mean	66.3403		
		Median	65.5000		
		Variance	63.096		
		Std. Deviation	7.94328		
		Minimum	53.00		
		Maximum	83.00		
		Range	30.00		
		Interquartile Range	12.75		
		Skewness	.253	.414	
		Kurtosis	-.875	.809	
Heart_Rate_W12	1.00	Mean	66.9063	1.65305	
		95% Confidence Interval for Mean	Lower Bound	63.5348		
			Upper Bound	70.2777		
		5% Trimmed Mean	66.8333		
		Median	65.5000		
		Variance	87.443		
		Std. Deviation	9.35107		
		Minimum	50.00		
		Maximum	84.00		
		Range	34.00		
		Interquartile Range	12.00		
		Skewness	.282	.414	
		Kurtosis	-.621	.809	
	2.00	Mean	70.3750	1.92383	
		95% Confidence Interval for Mean	Lower Bound	66.4513		
			Upper Bound	74.2987		
		5% Trimmed Mean	70.6944		
		Median	73.5000		
		Variance	118.435		
		Std. Deviation	10.88281		
		Minimum	50.00		
		Maximum	85.00		
		Range	35.00		
		Interquartile Range	18.25		
		Skewness	-.441	.414	
		Kurtosis	-1.041	.809	
Heart_Rate_Diff_W6	1.00	Mean	2.3750	1.88465	
		95% Confidence Interval for Mean	Lower Bound	-1.4688		
			Upper Bound	6.2188		
		5% Trimmed Mean	2.4167		
		Median	4.0000		
		Variance	113.661		
		Std. Deviation	10.66120		
		Minimum	-22.00		
		Maximum	26.00		
		Range	48.00		
		Interquartile Range	16.50		
		Skewness	-.098	.414	
		Kurtosis	-.284	.809	
	2.00	Mean	4.5938	1.73778	
		95% Confidence Interval for Mean	Lower Bound	1.0495		
			Upper Bound	8.1380		
		5% Trimmed Mean	4.7222		
		Median	6.0000		
		Variance	96.636		
		Std. Deviation	9.83037		
		Minimum	-15.00		
		Maximum	23.00		
		Range	38.00		
		Interquartile Range	12.50		
		Skewness	-.202	.414	
		Kurtosis	-.483	.809	
Heart_Rate_Diff_W12	1.00	Mean	1.6563	2.55646	
		95% Confidence Interval for Mean	Lower Bound	-3.5577		
			Upper Bound	6.8702		
		5% Trimmed Mean	1.9583		
		Median	6.0000		
		Variance	209.136		
		Std. Deviation	14.46154		
		Minimum	-29.00		
		Maximum	28.00		
		Range	57.00		
		Interquartile Range	24.00		
		Skewness	-.267	.414	
		Kurtosis	-.532	.809	
	2.00	Mean	.6875	2.66036	
		95% Confidence Interval for Mean	Lower Bound	-4.7383		
			Upper Bound	6.1133		
		5% Trimmed Mean	.4306		
		Median	.5000		
		Variance	226.480		
		Std. Deviation	15.04925		
		Minimum	-25.00		
		Maximum	31.00		
		Range	56.00		
		Interquartile Range	18.00		
		Skewness	.196	.414	
		Kurtosis	-.355	.809	
Oxy_Saturatn_B	1.00	Mean	98.1750	.21818	
		95% Confidence Interval for Mean	Lower Bound	97.7300		
			Upper Bound	98.6200		
		5% Trimmed Mean	98.1868		
		Median	98.2000		
		Variance	1.523		
		Std. Deviation	1.23419		
		Minimum	96.10		
		Maximum	100.00		
		Range	3.90		
		Interquartile Range	2.42		
		Skewness	-.235	.414	
		Kurtosis	-1.100	.809	
	2.00	Mean	98.2938	.23327	
		95% Confidence Interval for Mean	Lower Bound	97.8180		
			Upper Bound	98.7695		
		5% Trimmed Mean	98.3153		
		Median	98.5000		
		Variance	1.741		
		Std. Deviation	1.31956		
		Minimum	96.20		
		Maximum	100.00		
		Range	3.80		
		Interquartile Range	2.40		
		Skewness	-.417	.414	
		Kurtosis	-1.230	.809	
Oxy_Saturatn_W6	1.00	Mean	98.0750	.19899	
		95% Confidence Interval for Mean	Lower Bound	97.6692		
			Upper Bound	98.4808		
		5% Trimmed Mean	98.0889		
		Median	98.2000		
		Variance	1.267		
		Std. Deviation	1.12565		
		Minimum	96.10		
		Maximum	99.80		
		Range	3.70		
		Interquartile Range	1.90		
		Skewness	-.221	.414	
		Kurtosis	-1.234	.809	
	2.00	Mean	97.9750	.18762	
		95% Confidence Interval for Mean	Lower Bound	97.5923		
			Upper Bound	98.3577		
		5% Trimmed Mean	97.9632		
		Median	98.2000		
		Variance	1.126		
		Std. Deviation	1.06134		
		Minimum	96.30		
		Maximum	99.90		
		Range	3.60		
		Interquartile Range	1.78		
		Skewness	-.051	.414	
		Kurtosis	-1.012	.809	
Oxy_Saturatn_W12	1.00	Mean	97.7156	.19381	
		95% Confidence Interval for Mean	Lower Bound	97.3203		
			Upper Bound	98.1109		
		5% Trimmed Mean	97.6861		
		Median	97.4500		
		Variance	1.202		
		Std. Deviation	1.09636		
		Minimum	96.20		
		Maximum	99.80		
		Range	3.60		
		Interquartile Range	1.85		
		Skewness	.396	.414	
		Kurtosis	-1.037	.809	
	2.00	Mean	97.8625	.21269	
		95% Confidence Interval for Mean	Lower Bound	97.4287		
			Upper Bound	98.2963		
		5% Trimmed Mean	97.8472		
		Median	97.9500		
		Variance	1.448		
		Std. Deviation	1.20315		
		Minimum	96.10		
		Maximum	99.90		
		Range	3.80		
		Interquartile Range	2.30		
		Skewness	.104	.414	
		Kurtosis	-1.272	.809	
Oxy_Saturatn_Diff_W6	1.00	Mean	.1000	.30799	
		95% Confidence Interval for Mean	Lower Bound	-.5282		
			Upper Bound	.7282		
		5% Trimmed Mean	.0750		
		Median	.0000		
		Variance	3.035		
		Std. Deviation	1.74226		
		Minimum	-3.10		
		Maximum	3.90		
		Range	7.00		
		Interquartile Range	1.93		
		Skewness	.271	.414	
		Kurtosis	-.199	.809	
	2.00	Mean	.3188	.28854	
		95% Confidence Interval for Mean	Lower Bound	-.2697		
			Upper Bound	.9072		
		5% Trimmed Mean	.3125		
		Median	.5500		
		Variance	2.664		
		Std. Deviation	1.63222		
		Minimum	-2.80		
		Maximum	3.40		
		Range	6.20		
		Interquartile Range	2.30		
		Skewness	.071	.414	
		Kurtosis	-.850	.809	
Oxy_Saturatn_Diff_W12	1.00	Mean	.4594	.26239	
		95% Confidence Interval for Mean	Lower Bound	-.0758		
			Upper Bound	.9945		
		5% Trimmed Mean	.4604		
		Median	.3500		
		Variance	2.203		
		Std. Deviation	1.48430		
		Minimum	-2.60		
		Maximum	3.80		
		Range	6.40		
		Interquartile Range	2.10		
		Skewness	.097	.414	
		Kurtosis	-.034	.809	
	2.00	Mean	.4313	.33743	
		95% Confidence Interval for Mean	Lower Bound	-.2569		
			Upper Bound	1.1194		
		5% Trimmed Mean	.4764		
		Median	.7000		
		Variance	3.644		
		Std. Deviation	1.90880		
		Minimum	-3.50		
		Maximum	3.40		
		Range	6.90		
		Interquartile Range	2.68		
		Skewness	-.372	.414	
		Kurtosis	-.703	.809	
Oral_Temp_B	1.00	Mean	97.8656	.09361	
		95% Confidence Interval for Mean	Lower Bound	97.6747		
			Upper Bound	98.0565		
		5% Trimmed Mean	97.8528		
		Median	97.7500		
		Variance	.280		
		Std. Deviation	.52952		
		Minimum	97.00		
		Maximum	99.00		
		Range	2.00		
		Interquartile Range	.95		
		Skewness	.439	.414	
		Kurtosis	-.808	.809	
	2.00	Mean	97.9031	.11337	
		95% Confidence Interval for Mean	Lower Bound	97.6719		
			Upper Bound	98.1343		
		5% Trimmed Mean	97.8958		
		Median	97.8000		
		Variance	.411		
		Std. Deviation	.64131		
		Minimum	97.00		
		Maximum	98.90		
		Range	1.90		
		Interquartile Range	1.27		
		Skewness	.203	.414	
		Kurtosis	-1.401	.809	
Oral_Temp_W6	1.00	Mean	97.8656	.10565	
		95% Confidence Interval for Mean	Lower Bound	97.6502		
			Upper Bound	98.0811		
		5% Trimmed Mean	97.8486		
		Median	97.9000		
		Variance	.357		
		Std. Deviation	.59763		
		Minimum	97.00		
		Maximum	99.00		
		Range	2.00		
		Interquartile Range	1.18		
		Skewness	.271	.414	
		Kurtosis	-1.026	.809	
	2.00	Mean	98.0125	.10451	
		95% Confidence Interval for Mean	Lower Bound	97.7994		
			Upper Bound	98.2256		
		5% Trimmed Mean	98.0194		
		Median	98.0500		
		Variance	.350		
		Std. Deviation	.59120		
		Minimum	97.00		
		Maximum	98.90		
		Range	1.90		
		Interquartile Range	1.20		
		Skewness	-.212	.414	
		Kurtosis	-1.116	.809	
Oral_Temp_W12	1.00	Mean	97.9375	.10595	
		95% Confidence Interval for Mean	Lower Bound	97.7214		
			Upper Bound	98.1536		
		5% Trimmed Mean	97.9306		
		Median	97.8500		
		Variance	.359		
		Std. Deviation	.59933		
		Minimum	97.00		
		Maximum	99.00		
		Range	2.00		
		Interquartile Range	.90		
		Skewness	-.016	.414	
		Kurtosis	-1.213	.809	
	2.00	Mean	97.9406	.09211	
		95% Confidence Interval for Mean	Lower Bound	97.7528		
			Upper Bound	98.1285		
		5% Trimmed Mean	97.9375		
		Median	97.8500		
		Variance	.272		
		Std. Deviation	.52108		
		Minimum	97.10		
		Maximum	98.80		
		Range	1.70		
		Interquartile Range	.97		
		Skewness	.092	.414	
		Kurtosis	-1.380	.809	
Oral_Temp_Diff_W6	1.00	Mean	.0000	.13380	
		95% Confidence Interval for Mean	Lower Bound	-.2729		
			Upper Bound	.2729		
		5% Trimmed Mean	.0097		
		Median	.0000		
		Variance	.573		
		Std. Deviation	.75690		
		Minimum	-1.60		
		Maximum	1.50		
		Range	3.10		
		Interquartile Range	.88		
		Skewness	-.197	.414	
		Kurtosis	.034	.809	
	2.00	Mean	-.1094	.16384	
		95% Confidence Interval for Mean	Lower Bound	-.4435		
			Upper Bound	.2248		
		5% Trimmed Mean	-.1153		
		Median	-.1000		
		Variance	.859		
		Std. Deviation	.92679		
		Minimum	-1.60		
		Maximum	1.60		
		Range	3.20		
		Interquartile Range	1.78		
		Skewness	.048	.414	
		Kurtosis	-1.097	.809	
Oral_Temp_Diff_W12	1.00	Mean	-.0719	.13849	
		95% Confidence Interval for Mean	Lower Bound	-.3543		
			Upper Bound	.2106		
		5% Trimmed Mean	-.0819		
		Median	-.0500		
		Variance	.614		
		Std. Deviation	.78339		
		Minimum	-1.50		
		Maximum	1.50		
		Range	3.00		
		Interquartile Range	1.03		
		Skewness	.298	.414	
		Kurtosis	-.518	.809	
	2.00	Mean	-.0375	.15720	
		95% Confidence Interval for Mean	Lower Bound	-.3581		
			Upper Bound	.2831		
		5% Trimmed Mean	-.0417		
		Median	-.1000		
		Variance	.791		
		Std. Deviation	.88927		
		Minimum	-1.40		
		Maximum	1.40		
		Range	2.80		
		Interquartile Range	1.53		
		Skewness	.074	.414	
		Kurtosis	-1.134	.809	
Resp_Rate_B	1.00	Mean	14.7500	.32066	
		95% Confidence Interval for Mean	Lower Bound	14.0960		
			Upper Bound	15.4040		
		5% Trimmed Mean	14.7431		
		Median	15.0000		
		Variance	3.290		
		Std. Deviation	1.81392		
		Minimum	12.00		
		Maximum	18.00		
		Range	6.00		
		Interquartile Range	3.00		
		Skewness	-.017	.414	
		Kurtosis	-1.265	.809	
	2.00	Mean	15.4375	.34762	
		95% Confidence Interval for Mean	Lower Bound	14.7285		
			Upper Bound	16.1465		
		5% Trimmed Mean	15.4861		
		Median	15.5000		
		Variance	3.867		
		Std. Deviation	1.96645		
		Minimum	12.00		
		Maximum	18.00		
		Range	6.00		
		Interquartile Range	3.00		
		Skewness	-.172	.414	
		Kurtosis	-1.099	.809	
Resp_Rate_W6	1.00	Mean	14.5313	.35065	
		95% Confidence Interval for Mean	Lower Bound	13.8161		
			Upper Bound	15.2464		
		5% Trimmed Mean	14.5000		
		Median	14.5000		
		Variance	3.934		
		Std. Deviation	1.98355		
		Minimum	12.00		
		Maximum	18.00		
		Range	6.00		
		Interquartile Range	3.75		
		Skewness	.149	.414	
		Kurtosis	-1.508	.809	
	2.00	Mean	15.0938	.33105	
		95% Confidence Interval for Mean	Lower Bound	14.4186		
			Upper Bound	15.7689		
		5% Trimmed Mean	15.1042		
		Median	15.0000		
		Variance	3.507		
		Std. Deviation	1.87271		
		Minimum	12.00		
		Maximum	18.00		
		Range	6.00		
		Interquartile Range	3.75		
		Skewness	.075	.414	
		Kurtosis	-1.207	.809	
Resp_Rate_W12	1.00	Mean	14.8438	.38129	
		95% Confidence Interval for Mean	Lower Bound	14.0661		
			Upper Bound	15.6214		
		5% Trimmed Mean	14.8264		
		Median	15.0000		
		Variance	4.652		
		Std. Deviation	2.15690		
		Minimum	12.00		
		Maximum	18.00		
		Range	6.00		
		Interquartile Range	4.00		
		Skewness	.215	.414	
		Kurtosis	-1.369	.809	
	2.00	Mean	15.8438	.34521	
		95% Confidence Interval for Mean	Lower Bound	15.1397		
			Upper Bound	16.5478		
		5% Trimmed Mean	15.9375		
		Median	16.5000		
		Variance	3.814		
		Std. Deviation	1.95282		
		Minimum	12.00		
		Maximum	18.00		
		Range	6.00		
		Interquartile Range	4.00		
		Skewness	-.432	.414	
		Kurtosis	-1.065	.809	
Resp_Rate_Diff_W6	1.00	Mean	.2188	.40594	
		95% Confidence Interval for Mean	Lower Bound	-.6092		
			Upper Bound	1.0467		
		5% Trimmed Mean	.2222		
		Median	.0000		
		Variance	5.273		
		Std. Deviation	2.29634		
		Minimum	-5.00		
		Maximum	5.00		
		Range	10.00		
		Interquartile Range	2.75		
		Skewness	.072	.414	
		Kurtosis	.082	.809	
	2.00	Mean	.3438	.53808	
		95% Confidence Interval for Mean	Lower Bound	-.7537		
			Upper Bound	1.4412		
		5% Trimmed Mean	.4167		
		Median	.0000		
		Variance	9.265		
		Std. Deviation	3.04387		
		Minimum	-6.00		
		Maximum	5.00		
		Range	11.00		
		Interquartile Range	4.75		
		Skewness	-.027	.414	
		Kurtosis	-.655	.809	
Resp_Rate_Diff_W12	1.00	Mean	-.0938	.58778	
		95% Confidence Interval for Mean	Lower Bound	-1.2925		
			Upper Bound	1.1050		
		5% Trimmed Mean	-.0486		
		Median	.0000		
		Variance	11.055		
		Std. Deviation	3.32497		
		Minimum	-6.00		
		Maximum	5.00		
		Range	11.00		
		Interquartile Range	5.75		
		Skewness	-.104	.414	
		Kurtosis	-1.047	.809	
	2.00	Mean	-.4063	.50373	
		95% Confidence Interval for Mean	Lower Bound	-1.4336		
			Upper Bound	.6211		
		5% Trimmed Mean	-.5278		
		Median	-.5000		
		Variance	8.120		
		Std. Deviation	2.84955		
		Minimum	-5.00		
		Maximum	6.00		
		Range	11.00		
		Interquartile Range	4.00		
		Skewness	.672	.414	
		Kurtosis	.091	.809	
