## Supplementary material for "Effects of a standardised *Amaranthus tricolor*–*Spinacia oleracea* extract on exercise performance, muscle strength, endurance and recovery in physically active adults: a randomised, double-blind, placebo-controlled trial": Raw statistical outputs: Age___Gender_.rtf

T-Test


Notes	
Output Created	14-JUL-2026 09:56:26	
Comments		
Input	Active Dataset	DataSet0	
	Filter	<none>	
	Weight	<none>	
	Split File	<none>	
	N of Rows in Working Data File	64	
Missing Value Handling	Definition of Missing	User defined missing values are treated as missing.	
	Cases Used	Statistics for each analysis are based on the cases with no missing or out-of-range data for any variable in the analysis.	
Syntax	T-TEST GROUPS=Groups(1 2)
  /MISSING=ANALYSIS
  /VARIABLES=Age
  /ES DISPLAY(TRUE)
  /CRITERIA=CI(.95).	
Resources	Processor Time	00:00:00.00	
	Elapsed Time	00:00:00.00	


Group Statistics	
	Groups	N	Mean	Std. Deviation	Std. Error Mean	
Age	1.00	32	31.5000	5.27930	.93326	
	2.00	32	31.6875	5.17711	.91519	


Independent Samples Test	
	Levene's Test for Equality of Variances	t-test for Equality of Means	
	F	Sig.	t	df	
					
Age	Equal variances assumed	.012	.911	-.143	62	
	Equal variances not assumed			-.143	61.976	

Independent Samples Test	
	t-test for Equality of Means	
	Significance	Mean Difference	Std. Error Difference	
	One-Sided p	Two-Sided p			
Age	Equal variances assumed	.443	.884	-.18750	1.30711	
	Equal variances not assumed	.443	.884	-.18750	1.30711	

Independent Samples Test	
	t-test for Equality of Means	
	95% Confidence Interval of the Difference	
	Lower	Upper	
Age	Equal variances assumed	-2.80038	2.42538	
	Equal variances not assumed	-2.80040	2.42540	


Independent Samples Effect Sizes	
	Standardizer^a^	Point Estimate	95% Confidence Interval	
			Lower	Upper	
Age	Cohen's d	5.22845	-.036	-.526	.454	
	Hedges' correction	5.29278	-.035	-.519	.449	
	Glass's delta	5.17711	-.036	-.526	.454	

a. The denominator used in estimating the effect sizes. 
Cohen's d uses the pooled standard deviation. 
Hedges' correction uses the pooled standard deviation, plus a correction factor. 
Glass's delta uses the sample standard deviation of the control (i.e., the second) group.	


Crosstabs


Notes	
Output Created	14-JUL-2026 09:57:57	
Comments		
Input	Active Dataset	DataSet0	
	Filter	<none>	
	Weight	<none>	
	Split File	<none>	
	N of Rows in Working Data File	64	
Missing Value Handling	Definition of Missing	User-defined missing values are treated as missing.	
	Cases Used	Statistics for each table are based on all the cases with valid data in the specified range(s) for all variables in each table.	
Syntax	CROSSTABS
  /TABLES=Gender BY Groups
  /FORMAT=AVALUE TABLES
  /STATISTICS=CHISQ PHI
  /CELLS=COUNT ROW COLUMN TOTAL
  /COUNT ROUND CELL.	
Resources	Processor Time	00:00:00.00	
	Elapsed Time	00:00:00.00	
	Dimensions Requested	2	
	Cells Available	524245	


Case Processing Summary	
	Cases	
	Valid	Missing	Total	
	N	Percent	N	Percent	N	Percent	
Gender * Groups	64	100.0%	0	0.0%	64	100.0%	


Gender * Groups Crosstabulation	
	Groups	Total	
	1.00	2.00		
Gender	Female	Count	8	7	15	
		% within Gender	53.3%	46.7%	100.0%	
		% within Groups	25.0%	21.9%	23.4%	
		% of Total	12.5%	10.9%	23.4%	
	Male	Count	24	25	49	
		% within Gender	49.0%	51.0%	100.0%	
		% within Groups	75.0%	78.1%	76.6%	
		% of Total	37.5%	39.1%	76.6%	
Total	Count	32	32	64	
	% within Gender	50.0%	50.0%	100.0%	
	% within Groups	100.0%	100.0%	100.0%	
	% of Total	50.0%	50.0%	100.0%	


Chi-Square Tests	
	Value	df	Asymptotic Significance (2-sided)	Exact Sig. (2-sided)	Exact Sig. (1-sided)	
Pearson Chi-Square	.087^a^	1	.767			
Continuity Correction^b^	.000	1	1.000			
Likelihood Ratio	.087	1	.767			
Fisher's Exact Test				1.000	.500	
N of Valid Cases	64					

a. 0 cells (0.0%) have expected count less than 5. The minimum expected count is 7.50.	
b. Computed only for a 2x2 table	


Symmetric Measures	
	Value	Approximate Significance	
Nominal by Nominal	Phi	.037	.767	
	Cramer's V	.037	.767	
N of Valid Cases	64		
