## Supplementary material for "Effects of a standardised *Amaranthus tricolor*–*Spinacia oleracea* extract on exercise performance, muscle strength, endurance and recovery in physically active adults: a randomised, double-blind, placebo-controlled trial": Raw statistical outputs: Anthro_Non para. tests.rtf

NPar Tests


Notes	
Output Created	15-MAY-2026 15:33:12	
Comments		
Input	Active Dataset	DataSet0	
	Filter	<none>	
	Weight	<none>	
	Split File	<none>	
	N of Rows in Working Data File	64	
Missing Value Handling	Definition of Missing	User-defined missing values are treated as missing.	
	Cases Used	Statistics for each test are based on all cases with valid data for the variable(s) used in that test.	
Syntax	NPAR TESTS
  /M-W= Weight_B Weight_Diff_W6 Weight_Diff_W12 BMI_B BMI_Diff_W6 BMI_Diff_W12 Hip_B Hip_Diff_W6
    Hip_Diff_W12 Waist_B Waist_Diff_W6 Waist_Diff_W12 Body_Fat_B Body_Fat_Diff_W6 Body_Fat_Diff_W12 BY
    Groups(1 2)
  /STATISTICS=DESCRIPTIVES QUARTILES
  /MISSING ANALYSIS.	
Resources	Processor Time	00:00:00.00	
	Elapsed Time	00:00:00.00	
	Number of Cases Allowed^a^	149796	

a. Based on availability of workspace memory.	


[DataSet0] 


Descriptive Statistics	
	N	Mean	Std. Deviation	Minimum	Maximum	Percentiles	
						25th	
Weight_B	64	74.8656	5.75437	63.60	90.20	70.4500	
Weight_Diff_W6	64	-.4094	1.45545	-5.90	3.00	-1.1750	
Weight_Diff_W12	64	-.7344	1.86558	-4.40	5.60	-1.8750	
BMI_B	64	26.0559	.94050	23.31	29.76	25.4325	
BMI_Diff_W6	64	-.1506	.50884	-1.93	1.11	-.4175	
BMI_Diff_W12	64	-.2700	.65968	-1.69	2.06	-.7025	
Hip_B	64	92.5781	5.57433	83.00	108.00	89.0000	
Hip_Diff_W6	64	.9969	.89122	.00	5.00	1.0000	
Hip_Diff_W12	64	1.3719	1.12115	.00	5.00	1.0000	
Waist_B	64	86.5469	5.99717	75.00	102.00	82.0000	
Waist_Diff_W6	64	1.2500	1.75481	-7.00	6.00	1.0000	
Waist_Diff_W12	64	1.5156	1.84298	-7.00	7.00	1.0000	
Body_Fat_B	64	18.3969	5.60782	8.20	31.30	14.3000	
Body_Fat_Diff_W6	64	.9094	1.81193	-3.20	7.00	.0000	
Body_Fat_Diff_W12	64	1.1969	1.78201	-4.40	6.50	.0250	
Groups	64	1.5000	.50395	1.00	2.00	1.0000	

Descriptive Statistics	
	Percentiles	
	50th (Median)	75th	
Weight_B	75.1500	78.5500	
Weight_Diff_W6	-.5000	.6750	
Weight_Diff_W12	-.7000	.2000	
BMI_B	26.0400	26.4600	
BMI_Diff_W6	-.1750	.2350	
BMI_Diff_W12	-.2500	.0750	
Hip_B	92.0000	94.0000	
Hip_Diff_W6	1.0000	1.0000	
Hip_Diff_W12	1.0000	2.0000	
Waist_B	86.0000	90.0000	
Waist_Diff_W6	1.0000	2.0000	
Waist_Diff_W12	2.0000	2.0000	
Body_Fat_B	16.8000	21.6000	
Body_Fat_Diff_W6	.7000	1.5000	
Body_Fat_Diff_W12	.8500	2.0000	
Groups	1.5000	2.0000	


Mann-Whitney Test


Ranks	
	Groups	N	Mean Rank	Sum of Ranks	
Weight_B	1.00	32	31.67	1013.50	
	2.00	32	33.33	1066.50	
	Total	64			
Weight_Diff_W6	1.00	32	27.33	874.50	
	2.00	32	37.67	1205.50	
	Total	64			
Weight_Diff_W12	1.00	32	28.58	914.50	
	2.00	32	36.42	1165.50	
	Total	64			
BMI_B	1.00	32	29.84	955.00	
	2.00	32	35.16	1125.00	
	Total	64			
BMI_Diff_W6	1.00	32	27.30	873.50	
	2.00	32	37.70	1206.50	
	Total	64			
BMI_Diff_W12	1.00	32	28.66	917.00	
	2.00	32	36.34	1163.00	
	Total	64			
Hip_B	1.00	32	35.64	1140.50	
	2.00	32	29.36	939.50	
	Total	64			
Hip_Diff_W6	1.00	32	34.19	1094.00	
	2.00	32	30.81	986.00	
	Total	64			
Hip_Diff_W12	1.00	32	36.58	1170.50	
	2.00	32	28.42	909.50	
	Total	64			
Waist_B	1.00	32	28.44	910.00	
	2.00	32	36.56	1170.00	
	Total	64			
Waist_Diff_W6	1.00	32	34.41	1101.00	
	2.00	32	30.59	979.00	
	Total	64			
Waist_Diff_W12	1.00	32	36.42	1165.50	
	2.00	32	28.58	914.50	
	Total	64			
Body_Fat_B	1.00	32	32.41	1037.00	
	2.00	32	32.59	1043.00	
	Total	64			
Body_Fat_Diff_W6	1.00	32	39.92	1277.50	
	2.00	32	25.08	802.50	
	Total	64			
Body_Fat_Diff_W12	1.00	32	37.95	1214.50	
	2.00	32	27.05	865.50	
	Total	64			


Test Statistics^a^	
	Weight_B	Weight_Diff_W6	Weight_Diff_W12	BMI_B	BMI_Diff_W6	
Mann-Whitney U	485.500	346.500	386.500	427.000	345.500	
Wilcoxon W	1013.500	874.500	914.500	955.000	873.500	
Z	-.356	-2.224	-1.687	-1.141	-2.236	
Asymp. Sig. (2-tailed)	.722	.026	.092	.254	.025	

Test Statistics^a^	
	BMI_Diff_W12	Hip_B	Hip_Diff_W6	Hip_Diff_W12	Waist_B	
Mann-Whitney U	389.000	411.500	458.000	381.500	382.000	
Wilcoxon W	917.000	939.500	986.000	909.500	910.000	
Z	-1.652	-1.355	-.888	-1.851	-1.749	
Asymp. Sig. (2-tailed)	.099	.175	.375	.064	.080	

Test Statistics^a^	
	Waist_Diff_W6	Waist_Diff_W12	Body_Fat_B	Body_Fat_Diff_W6	
Mann-Whitney U	451.000	386.500	509.000	274.500	
Wilcoxon W	979.000	914.500	1037.000	802.500	
Z	-.847	-1.741	-.040	-3.200	
Asymp. Sig. (2-tailed)	.397	.082	.968	.001	

Test Statistics^a^	
	Body_Fat_Diff_W12	
Mann-Whitney U	337.500	
Wilcoxon W	865.500	
Z	-2.346	
Asymp. Sig. (2-tailed)	.019	

a. Grouping Variable: Groups	


NPar Tests


Notes	
Output Created	15-MAY-2026 15:34:43	
Comments		
Input	Active Dataset	DataSet0	
	Filter	<none>	
	Weight	<none>	
	Split File	<none>	
	N of Rows in Working Data File	64	
Missing Value Handling	Definition of Missing	User-defined missing values are treated as missing.	
	Cases Used	Statistics for each test are based on all cases with valid data for the variable(s) used in that test.	
Syntax	NPAR TESTS
  /WILCOXON=Weight_B_1 Weight_B_1 BMI_B_1 BMI_B_1 Hip_B_1 Hip_B_1 Waist_B_1 Waist_B_1 Body_Fat_B_1
    Body_Fat_B_1 Weight_B_2 Weight_B_2 BMI_B_2 BMI_B_2 Hip_B_2 Hip_B_2 Waist_B_2 Waist_B_2 Body_Fat_B_2
    Body_Fat_B_2 WITH Weight_W6_1 Weight_W12_1 BMI_W6_1 BMI_W12_1 Hip_W6_1 Hip_W12_1 Waist_W6_1
    Waist_W12_1 Body_Fat_W6_1 Body_Fat_W12_1 Weight_W6_2 Weight_W12_2 BMI_W6_2 BMI_W12_2 Hip_W6_2
    Hip_W12_2 Waist_W6_2 Waist_W12_2 Body_Fat_W6_2 Body_Fat_W12_2 (PAIRED)
  /STATISTICS DESCRIPTIVES QUARTILES
  /MISSING ANALYSIS.	
Resources	Processor Time	00:00:00.00	
	Elapsed Time	00:00:00.00	
	Number of Cases Allowed^a^	89877	

a. Based on availability of workspace memory.	


Descriptive Statistics	
	N	Mean	Std. Deviation	Minimum	Maximum	Percentiles	
						25th	
Weight_B_1	32	74.6531	6.01825	65.20	90.20	70.6000	
BMI_B_1	32	25.8794	.82144	23.31	27.83	25.4325	
Hip_B_1	32	93.5313	6.45573	83.00	108.00	89.2500	
Waist_B_1	32	85.3125	6.58658	75.00	102.00	80.0000	
Body_Fat_B_1	32	18.5438	5.85419	9.30	31.30	14.4000	
Weight_B_2	32	75.0781	5.56607	63.60	89.00	70.1750	
BMI_B_2	32	26.2325	1.02884	25.02	29.76	25.4175	
Hip_B_2	32	91.6250	4.42682	83.00	102.00	89.0000	
Waist_B_2	32	87.7812	5.15398	80.00	100.00	84.0000	
Body_Fat_B_2	32	18.2500	5.44000	8.20	30.20	14.2250	
Weight_W6_1	32	75.4500	5.84416	66.10	90.70	70.9250	
Weight_W12_1	32	75.8031	5.72896	67.00	90.10	71.4500	
BMI_W6_1	32	26.1628	.78985	24.76	27.69	25.5625	
BMI_W12_1	32	26.2912	.89394	23.51	28.16	25.9350	
Hip_W6_1	32	92.3750	6.10526	83.00	107.00	88.2500	
Hip_W12_1	32	91.8750	5.74035	82.00	105.00	89.0000	
Waist_W6_1	32	83.7500	6.47576	74.00	101.00	79.2500	
Waist_W12_1	32	83.3750	6.35382	74.00	100.00	79.2500	
Body_Fat_W6_1	32	17.1219	5.31181	8.20	27.10	13.3000	
Body_Fat_W12_1	32	16.8844	4.90326	8.00	27.90	13.2500	
Weight_W6_2	32	75.1000	5.27930	64.10	88.20	70.5250	
Weight_W12_2	32	75.3969	5.19773	65.40	89.10	71.5250	
BMI_W6_2	32	26.2503	1.08806	24.81	29.69	25.3000	
BMI_W12_2	32	26.3606	1.18367	24.07	29.55	25.4550	
Hip_W6_2	32	90.7875	4.26537	82.00	101.00	88.2500	
Hip_W12_2	32	90.5375	4.29777	81.00	101.00	88.0000	
Waist_W6_2	32	86.8437	5.10603	80.00	98.00	83.0000	
Waist_W12_2	32	86.6875	4.90186	80.00	97.00	83.0000	
Body_Fat_W6_2	32	17.8531	5.30569	11.20	32.20	13.9750	
Body_Fat_W12_2	32	17.5156	5.02517	9.60	26.90	13.0500	

Descriptive Statistics	
	Percentiles	
	50th (Median)	75th	
Weight_B_1	72.6000	78.8750	
BMI_B_1	25.8700	26.4025	
Hip_B_1	92.5000	97.0000	
Waist_B_1	85.5000	89.5000	
Body_Fat_B_1	16.3500	22.0750	
Weight_B_2	75.6500	78.1000	
BMI_B_2	26.1950	26.6025	
Hip_B_2	91.0000	93.7500	
Waist_B_2	87.5000	90.0000	
Body_Fat_B_2	17.5000	21.1500	
Weight_W6_1	74.4500	79.1750	
Weight_W12_1	74.8500	79.9500	
BMI_W6_1	26.1300	26.8750	
BMI_W12_1	26.2800	26.8600	
Hip_W6_1	91.5000	96.0000	
Hip_W12_1	91.0000	95.0000	
Waist_W6_1	83.5000	87.7500	
Waist_W12_1	83.0000	87.5000	
Body_Fat_W6_1	14.9000	20.2000	
Body_Fat_W12_1	15.2000	20.7750	
Weight_W6_2	75.7500	78.1250	
Weight_W12_2	75.5000	78.0750	
BMI_W6_2	26.3200	26.7025	
BMI_W12_2	26.3150	27.0150	
Hip_W6_2	90.0000	92.8000	
Hip_W12_2	90.0000	92.8000	
Waist_W6_2	86.5000	88.7500	
Waist_W12_2	86.5000	88.7500	
Body_Fat_W6_2	16.5500	21.3250	
Body_Fat_W12_2	17.0000	20.5000	


Wilcoxon Signed Ranks Test


Ranks	
	N	Mean Rank	Sum of Ranks	
Weight_W6_1 - Weight_B_1	Negative Ranks	9^a^	14.56	131.00	
	Positive Ranks	23^b^	17.26	397.00	
	Ties	0^c^			
	Total	32			
Weight_W12_1 - Weight_B_1	Negative Ranks	9^d^	9.56	86.00	
	Positive Ranks	23^e^	19.22	442.00	
	Ties	0^f^			
	Total	32			
BMI_W6_1 - BMI_B_1	Negative Ranks	9^g^	14.17	127.50	
	Positive Ranks	23^h^	17.41	400.50	
	Ties	0^i^			
	Total	32			
BMI_W12_1 - BMI_B_1	Negative Ranks	9^j^	9.33	84.00	
	Positive Ranks	23^k^	19.30	444.00	
	Ties	0^l^			
	Total	32			
Hip_W6_1 - Hip_B_1	Negative Ranks	26^m^	13.50	351.00	
	Positive Ranks	0^n^	.00	.00	
	Ties	6^o^			
	Total	32			
Hip_W12_1 - Hip_B_1	Negative Ranks	26^p^	13.50	351.00	
	Positive Ranks	0^q^	.00	.00	
	Ties	6^r^			
	Total	32			
Waist_W6_1 - Waist_B_1	Negative Ranks	26^s^	13.50	351.00	
	Positive Ranks	0^t^	.00	.00	
	Ties	6^u^			
	Total	32			
Waist_W12_1 - Waist_B_1	Negative Ranks	28^v^	14.50	406.00	
	Positive Ranks	0^w^	.00	.00	
	Ties	4^x^			
	Total	32			
Body_Fat_W6_1 - Body_Fat_B_1	Negative Ranks	28^y^	15.80	442.50	
	Positive Ranks	3^z^	17.83	53.50	
	Ties	1^aa^			
	Total	32			
Body_Fat_W12_1 - Body_Fat_B_1	Negative Ranks	27^ab^	15.52	419.00	
	Positive Ranks	2^ac^	8.00	16.00	
	Ties	3^ad^			
	Total	32			
Weight_W6_2 - Weight_B_2	Negative Ranks	14^ae^	18.36	257.00	
	Positive Ranks	18^af^	15.06	271.00	
	Ties	0^ag^			
	Total	32			
Weight_W12_2 - Weight_B_2	Negative Ranks	11^ah^	15.41	169.50	
	Positive Ranks	20^ai^	16.33	326.50	
	Ties	1^aj^			
	Total	32			
BMI_W6_2 - BMI_B_2	Negative Ranks	14^ak^	17.50	245.00	
	Positive Ranks	18^al^	15.72	283.00	
	Ties	0^am^			
	Total	32			
BMI_W12_2 - BMI_B_2	Negative Ranks	11^an^	15.09	166.00	
	Positive Ranks	20^ao^	16.50	330.00	
	Ties	1^ap^			
	Total	32			
Hip_W6_2 - Hip_B_2	Negative Ranks	25^aq^	13.00	325.00	
	Positive Ranks	0^ar^	.00	.00	
	Ties	7^as^			
	Total	32			
Hip_W12_2 - Hip_B_2	Negative Ranks	26^at^	13.50	351.00	
	Positive Ranks	0^au^	.00	.00	
	Ties	6^av^			
	Total	32			
Waist_W6_2 - Waist_B_2	Negative Ranks	23^aw^	13.17	303.00	
	Positive Ranks	4^ax^	18.75	75.00	
	Ties	5^ay^			
	Total	32			
Waist_W12_2 - Waist_B_2	Negative Ranks	23^az^	13.98	321.50	
	Positive Ranks	4^ba^	14.13	56.50	
	Ties	5^bb^			
	Total	32			
Body_Fat_W6_2 - Body_Fat_B_2	Negative Ranks	15^bc^	13.97	209.50	
	Positive Ranks	10^bd^	11.55	115.50	
	Ties	7^be^			
	Total	32			
Body_Fat_W12_2 - Body_Fat_B_2	Negative Ranks	21^bf^	15.14	318.00	
	Positive Ranks	7^bg^	12.57	88.00	
	Ties	4^bh^			
	Total	32			

a. Weight_W6_1 < Weight_B_1	
b. Weight_W6_1 > Weight_B_1	
c. Weight_W6_1 = Weight_B_1	
d. Weight_W12_1 < Weight_B_1	
e. Weight_W12_1 > Weight_B_1	
f. Weight_W12_1 = Weight_B_1	
g. BMI_W6_1 < BMI_B_1	
h. BMI_W6_1 > BMI_B_1	
i. BMI_W6_1 = BMI_B_1	
j. BMI_W12_1 < BMI_B_1	
k. BMI_W12_1 > BMI_B_1	
l. BMI_W12_1 = BMI_B_1	
m. Hip_W6_1 < Hip_B_1	
n. Hip_W6_1 > Hip_B_1	
o. Hip_W6_1 = Hip_B_1	
p. Hip_W12_1 < Hip_B_1	
q. Hip_W12_1 > Hip_B_1	
r. Hip_W12_1 = Hip_B_1	
s. Waist_W6_1 < Waist_B_1	
t. Waist_W6_1 > Waist_B_1	
u. Waist_W6_1 = Waist_B_1	
v. Waist_W12_1 < Waist_B_1	
w. Waist_W12_1 > Waist_B_1	
x. Waist_W12_1 = Waist_B_1	
y. Body_Fat_W6_1 < Body_Fat_B_1	
z. Body_Fat_W6_1 > Body_Fat_B_1	
aa. Body_Fat_W6_1 = Body_Fat_B_1	
ab. Body_Fat_W12_1 < Body_Fat_B_1	
ac. Body_Fat_W12_1 > Body_Fat_B_1	
ad. Body_Fat_W12_1 = Body_Fat_B_1	
ae. Weight_W6_2 < Weight_B_2	
af. Weight_W6_2 > Weight_B_2	
ag. Weight_W6_2 = Weight_B_2	
ah. Weight_W12_2 < Weight_B_2	
ai. Weight_W12_2 > Weight_B_2	
aj. Weight_W12_2 = Weight_B_2	
ak. BMI_W6_2 < BMI_B_2	
al. BMI_W6_2 > BMI_B_2	
am. BMI_W6_2 = BMI_B_2	
an. BMI_W12_2 < BMI_B_2	
ao. BMI_W12_2 > BMI_B_2	
ap. BMI_W12_2 = BMI_B_2	
aq. Hip_W6_2 < Hip_B_2	
ar. Hip_W6_2 > Hip_B_2	
as. Hip_W6_2 = Hip_B_2	
at. Hip_W12_2 < Hip_B_2	
au. Hip_W12_2 > Hip_B_2	
av. Hip_W12_2 = Hip_B_2	
aw. Waist_W6_2 < Waist_B_2	
ax. Waist_W6_2 > Waist_B_2	
ay. Waist_W6_2 = Waist_B_2	
az. Waist_W12_2 < Waist_B_2	
ba. Waist_W12_2 > Waist_B_2	
bb. Waist_W12_2 = Waist_B_2	
bc. Body_Fat_W6_2 < Body_Fat_B_2	
bd. Body_Fat_W6_2 > Body_Fat_B_2	
be. Body_Fat_W6_2 = Body_Fat_B_2	
bf. Body_Fat_W12_2 < Body_Fat_B_2	
bg. Body_Fat_W12_2 > Body_Fat_B_2	
bh. Body_Fat_W12_2 = Body_Fat_B_2	


Test Statistics^a^	
	Weight_W6_1 - Weight_B_1	Weight_W12_1 - Weight_B_1	BMI_W6_1 - BMI_B_1	BMI_W12_1 - BMI_B_1	
Z	-2.488^b^	-3.329^b^	-2.553^b^	-3.366^b^	
Asymp. Sig. (2-tailed)	.013	<.001	.011	<.001	

Test Statistics^a^	
	Hip_W6_1 - Hip_B_1	Hip_W12_1 - Hip_B_1	Waist_W6_1 - Waist_B_1	Waist_W12_1 - Waist_B_1	
Z	-4.815^c^	-4.548^c^	-4.529^c^	-4.718^c^	
Asymp. Sig. (2-tailed)	<.001	<.001	<.001	<.001	

Test Statistics^a^	
	Body_Fat_W6_1 - Body_Fat_B_1	Body_Fat_W12_1 - Body_Fat_B_1	Weight_W6_2 - Weight_B_2	Weight_W12_2 - Weight_B_2	
Z	-3.818^c^	-4.358^c^	-.131^b^	-1.539^b^	
Asymp. Sig. (2-tailed)	<.001	<.001	.896	.124	

Test Statistics^a^	
	BMI_W6_2 - BMI_B_2	BMI_W12_2 - BMI_B_2	Hip_W6_2 - Hip_B_2	Hip_W12_2 - Hip_B_2	
Z	-.355^b^	-1.607^b^	-4.772^c^	-4.623^c^	
Asymp. Sig. (2-tailed)	.722	.108	<.001	<.001	

Test Statistics^a^	
	Waist_W6_2 - Waist_B_2	Waist_W12_2 - Waist_B_2	Body_Fat_W6_2 - Body_Fat_B_2	Body_Fat_W12_2 - Body_Fat_B_2	
Z	-2.779^c^	-3.227^c^	-1.268^c^	-2.620^c^	
Asymp. Sig. (2-tailed)	.005	.001	.205	.009	

a. Wilcoxon Signed Ranks Test	
b. Based on negative ranks.	
c. Based on positive ranks.	
