## Supplementary material for "Effects of a standardised *Amaranthus tricolor*–*Spinacia oleracea* extract on exercise performance, muscle strength, endurance and recovery in physically active adults: a randomised, double-blind, placebo-controlled trial": Raw statistical outputs: biomarker(Blood lact, Plas nitrite, S.creatinine)By ANCOVA.doc

Univariate Analysis of Variance


Notes	
Output Created	07-MAY-2026 11:20:45	
Comments		
Input	Active Dataset	DataSet0	
	Filter	<none>	
	Weight	<none>	
	Split File	<none>	
	N of Rows in Working Data File	64	
Missing Value Handling	Definition of Missing	User-defined missing values are treated as missing.	
	Cases Used	Statistics are based on all cases with valid data for all variables in the model.	
Syntax	UNIANOVA D90_Blood_lactate BY Groups WITH Bas_Blood_lactate
  /METHOD=SSTYPE(3)
  /INTERCEPT=INCLUDE
  /EMMEANS=TABLES(Groups) WITH(Bas_Blood_lactate=MEAN) COMPARE ADJ(BONFERRONI)
  /PRINT ETASQ DESCRIPTIVE HOMOGENEITY
  /CRITERIA=ALPHA(.05)
  /DESIGN=Bas_Blood_lactate Groups.	
Resources	Processor Time	00:00:00.00	
	Elapsed Time	00:00:00.02	


[DataSet0] 


Between-Subjects Factors	
	N	
Groups	Placebo	32	
	Test	32	


Descriptive Statistics	
Dependent Variable:   D90_Blood_lactate  	
Groups	Mean	Std. Deviation	N	
Placebo	71.3169	16.94312	32	
Test	40.0591	8.68717	32	
Total	55.6880	20.65262	64	


Levene's Test of Equality of Error Variances^a^	
Dependent Variable:   D90_Blood_lactate  	
F	df1	df2	Sig.	
1.169	1	62	.284	

Tests the null hypothesis that the error variance of the dependent variable is equal across groups.^a^	
a. Design: Intercept + Bas_Blood_lactate + Groups	


Tests of Between-Subjects Effects	
Dependent Variable:   D90_Blood_lactate  	
Source	Type III Sum of Squares	df	Mean Square	F	Sig.	
Corrected Model	19599.056^a^	2	9799.528	82.197	<.001	
Intercept	654.452	1	654.452	5.489	.022	
Bas_Blood_lactate	3966.242	1	3966.242	33.268	<.001	
Groups	13158.334	1	13158.334	110.371	<.001	
Error	7272.381	61	119.219			
Total	225345.028	64				
Corrected Total	26871.437	63				

Tests of Between-Subjects Effects	
Dependent Variable:   D90_Blood_lactate  	
Source	Partial Eta Squared	
Corrected Model	.729	
Intercept	.083	
Bas_Blood_lactate	.353	
Groups	.644	
Error		
Total		
Corrected Total		

a. R Squared = .729 (Adjusted R Squared = .720)	


Estimated Marginal Means


Groups


Estimates	
Dependent Variable:   D90_Blood_lactate  	
Groups	Mean	Std. Error	95% Confidence Interval	
			Lower Bound	Upper Bound	
Placebo	70.176^a^	1.940	66.297	74.056	
Test	41.200^a^	1.940	37.320	45.079	

a. Covariates appearing in the model are evaluated at the following values: Bas_Blood_lactate = 70.8158.	


Pairwise Comparisons	
Dependent Variable:   D90_Blood_lactate  	
(I) Groups	(J) Groups	Mean Difference (I-J)	Std. Error	Sig.^b^	95% Confidence Interval for Difference^b^	
					Lower Bound	Upper Bound	
Placebo	Test	28.977^*^	2.758	<.001	23.461	34.492	
Test	Placebo	-28.977^*^	2.758	<.001	-34.492	-23.461	

Based on estimated marginal means	
*. The mean difference is significant at the .05 level.	
b. Adjustment for multiple comparisons: Bonferroni.	


Univariate Tests	
Dependent Variable:   D90_Blood_lactate  	
	Sum of Squares	df	Mean Square	F	Sig.	Partial Eta Squared	
Contrast	13158.334	1	13158.334	110.371	<.001	.644	
Error	7272.381	61	119.219				

The F tests the effect of Groups. This test is based on the linearly independent pairwise comparisons among the estimated marginal means.	


Univariate Analysis of Variance


Notes	
Output Created	07-MAY-2026 11:26:47	
Comments		
Input	Active Dataset	DataSet0	
	Filter	<none>	
	Weight	<none>	
	Split File	<none>	
	N of Rows in Working Data File	64	
Missing Value Handling	Definition of Missing	User-defined missing values are treated as missing.	
	Cases Used	Statistics are based on all cases with valid data for all variables in the model.	
Syntax	UNIANOVA D90_Plasma_Nitrite BY Groups WITH Bas_Plasma_Nitrite
  /METHOD=SSTYPE(3)
  /INTERCEPT=INCLUDE
  /EMMEANS=TABLES(Groups) WITH(Bas_Plasma_Nitrite=MEAN) COMPARE ADJ(BONFERRONI)
  /PRINT ETASQ DESCRIPTIVE HOMOGENEITY
  /CRITERIA=ALPHA(.05)
  /DESIGN=Bas_Plasma_Nitrite Groups.	
Resources	Processor Time	00:00:00.00	
	Elapsed Time	00:00:00.00	


Between-Subjects Factors	
	N	
Groups	Placebo	32	
	Test	32	


Descriptive Statistics	
Dependent Variable:   D90_Plasma_Nitrite  	
Groups	Mean	Std. Deviation	N	
Placebo	14.6881	2.51397	32	
Test	22.3619	3.68426	32	
Total	18.5250	4.97437	64	


Levene's Test of Equality of Error Variances^a^	
Dependent Variable:   D90_Plasma_Nitrite  	
F	df1	df2	Sig.	
19.672	1	62	<.001	

Tests the null hypothesis that the error variance of the dependent variable is equal across groups.^a^	
a. Design: Intercept + Bas_Plasma_Nitrite + Groups	


Tests of Between-Subjects Effects	
Dependent Variable:   D90_Plasma_Nitrite  	
Source	Type III Sum of Squares	df	Mean Square	F	Sig.	
Corrected Model	1160.000^a^	2	580.000	88.696	<.001	
Intercept	52.599	1	52.599	8.044	.006	
Bas_Plasma_Nitrite	217.817	1	217.817	33.309	<.001	
Groups	514.766	1	514.766	78.720	<.001	
Error	398.891	61	6.539			
Total	23522.131	64				
Corrected Total	1558.891	63				

Tests of Between-Subjects Effects	
Dependent Variable:   D90_Plasma_Nitrite  	
Source	Partial Eta Squared	
Corrected Model	.744	
Intercept	.117	
Bas_Plasma_Nitrite	.353	
Groups	.563	
Error		
Total		
Corrected Total		

a. R Squared = .744 (Adjusted R Squared = .736)	


Estimated Marginal Means


Groups


Estimates	
Dependent Variable:   D90_Plasma_Nitrite  	
Groups	Mean	Std. Error	95% Confidence Interval	
			Lower Bound	Upper Bound	
Placebo	15.454^a^	.471	14.512	16.396	
Test	21.596^a^	.471	20.654	22.538	

a. Covariates appearing in the model are evaluated at the following values: Bas_Plasma_Nitrite = 14.7625.	


Pairwise Comparisons	
Dependent Variable:   D90_Plasma_Nitrite  	
(I) Groups	(J) Groups	Mean Difference (I-J)	Std. Error	Sig.^b^	95% Confidence Interval for Difference^b^	
					Lower Bound	Upper Bound	
Placebo	Test	-6.142^*^	.692	<.001	-7.526	-4.757	
Test	Placebo	6.142^*^	.692	<.001	4.757	7.526	

Based on estimated marginal means	
*. The mean difference is significant at the .05 level.	
b. Adjustment for multiple comparisons: Bonferroni.	


Univariate Tests	
Dependent Variable:   D90_Plasma_Nitrite  	
	Sum of Squares	df	Mean Square	F	Sig.	Partial Eta Squared	
Contrast	514.766	1	514.766	78.720	<.001	.563	
Error	398.891	61	6.539				

The F tests the effect of Groups. This test is based on the linearly independent pairwise comparisons among the estimated marginal means.	


Univariate Analysis of Variance


Notes	
Output Created	07-MAY-2026 11:27:57	
Comments		
Input	Active Dataset	DataSet0	
	Filter	<none>	
	Weight	<none>	
	Split File	<none>	
	N of Rows in Working Data File	64	
Missing Value Handling	Definition of Missing	User-defined missing values are treated as missing.	
	Cases Used	Statistics are based on all cases with valid data for all variables in the model.	
Syntax	UNIANOVA D90_Serum_Creat BY Groups WITH Bas_Serum_Creat
  /METHOD=SSTYPE(3)
  /INTERCEPT=INCLUDE
  /EMMEANS=TABLES(Groups) WITH(Bas_Serum_Creat=MEAN) COMPARE ADJ(BONFERRONI)
  /PRINT ETASQ DESCRIPTIVE HOMOGENEITY
  /CRITERIA=ALPHA(.05)
  /DESIGN=Bas_Serum_Creat Groups.	
Resources	Processor Time	00:00:00.02	
	Elapsed Time	00:00:00.00	


Between-Subjects Factors	
	N	
Groups	Placebo	32	
	Test	32	


Descriptive Statistics	
Dependent Variable:   D90_Serum_Creat  	
Groups	Mean	Std. Deviation	N	
Placebo	4.1562	.77239	32	
Test	5.8353	.94858	32	
Total	4.9958	1.20512	64	


Levene's Test of Equality of Error Variances^a^	
Dependent Variable:   D90_Serum_Creat  	
F	df1	df2	Sig.	
7.465	1	62	.008	

Tests the null hypothesis that the error variance of the dependent variable is equal across groups.^a^	
a. Design: Intercept + Bas_Serum_Creat + Groups	


Tests of Between-Subjects Effects	
Dependent Variable:   D90_Serum_Creat  	
Source	Type III Sum of Squares	df	Mean Square	F	Sig.	
Corrected Model	60.558^a^	2	30.279	59.699	<.001	
Intercept	2.561	1	2.561	5.050	.028	
Bas_Serum_Creat	15.450	1	15.450	30.462	<.001	
Groups	33.069	1	33.069	65.200	<.001	
Error	30.939	61	.507			
Total	1688.797	64				
Corrected Total	91.496	63				

Tests of Between-Subjects Effects	
Dependent Variable:   D90_Serum_Creat  	
Source	Partial Eta Squared	
Corrected Model	.662	
Intercept	.076	
Bas_Serum_Creat	.333	
Groups	.517	
Error		
Total		
Corrected Total		

a. R Squared = .662 (Adjusted R Squared = .651)	


Estimated Marginal Means


Groups


Estimates	
Dependent Variable:   D90_Serum_Creat  	
Groups	Mean	Std. Error	95% Confidence Interval	
			Lower Bound	Upper Bound	
Placebo	4.261^a^	.127	4.006	4.515	
Test	5.731^a^	.127	5.476	5.985	

a. Covariates appearing in the model are evaluated at the following values: Bas_Serum_Creat = 3.9981.	


Pairwise Comparisons	
Dependent Variable:   D90_Serum_Creat  	
(I) Groups	(J) Groups	Mean Difference (I-J)	Std. Error	Sig.^b^	95% Confidence Interval for Difference^b^	
					Lower Bound	Upper Bound	
Placebo	Test	-1.470^*^	.182	<.001	-1.834	-1.106	
Test	Placebo	1.470^*^	.182	<.001	1.106	1.834	

Based on estimated marginal means	
*. The mean difference is significant at the .05 level.	
b. Adjustment for multiple comparisons: Bonferroni.	


Univariate Tests	
Dependent Variable:   D90_Serum_Creat  	
	Sum of Squares	df	Mean Square	F	Sig.	Partial Eta Squared	
Contrast	33.069	1	33.069	65.200	<.001	.517	
Error	30.939	61	.507				

The F tests the effect of Groups. This test is based on the linearly independent pairwise comparisons among the estimated marginal means.	
