## Supplementary material for "Effects of a standardised *Amaranthus tricolor*–*Spinacia oleracea* extract on exercise performance, muscle strength, endurance and recovery in physically active adults: a randomised, double-blind, placebo-controlled trial": Raw statistical outputs: Biomarkers ( CK, LDH, cortisol, S. ferrittin, S. iron)- non para tests.rtf

NPar Tests


Notes	
Output Created	11-MAY-2026 14:31:37	
Comments		
Input	Active Dataset	DataSet0	
	Filter	<none>	
	Weight	<none>	
	Split File	<none>	
	N of Rows in Working Data File	64	
Missing Value Handling	Definition of Missing	User-defined missing values are treated as missing.	
	Cases Used	Statistics for each test are based on all cases with valid data for the variable(s) used in that test.	
Syntax	NPAR TESTS
  /M-W= B_Creat_Kinase Diff_Creat_Kinase B_lactate_dehydr Diff_lactate_dehydr B_cortiscol
    Diff_cortiscol B_Ser_Ferritine Diff_Ser_Ferritine B_Ser_Iron Diff_Ser_Iron BY Groups(1 2)
  /STATISTICS=DESCRIPTIVES QUARTILES
  /MISSING ANALYSIS.	
Resources	Processor Time	00:00:00.02	
	Elapsed Time	00:00:00.01	
	Number of Cases Allowed^a^	196608	

a. Based on availability of workspace memory.	


[DataSet0] 


Descriptive Statistics	
	N	Mean	Std. Deviation	Minimum	Maximum	Percentiles	
						25th	
B_Creat_Kinase	64	120.2848	38.80060	49.20	210.59	94.3725	
Diff_Creat_Kinase	64	12.3350	11.03816	-15.39	48.90	5.4125	
B_lactate_dehydr	64	236.4061	39.14569	115.80	409.00	223.2750	
Diff_lactate_dehydr	64	25.3906	22.84295	-19.60	96.30	10.9000	
B_cortiscol	64	144.9905	50.65417	87.00	289.63	106.8550	
Diff_cortiscol	64	15.1078	23.70969	-30.51	112.26	1.6875	
B_Ser_Ferritine	64	41.4411	52.09669	9.32	244.00	16.1550	
Diff_Ser_Ferritine	64	-14.4102	84.82086	-235.09	226.00	-33.0500	
B_Ser_Iron	64	75.2872	36.76899	20.90	192.09	46.6825	
Diff_Ser_Iron	64	-26.6092	48.46818	-120.65	95.21	-70.2950	
Groups	64	1.5000	.50395	1.00	2.00	1.0000	

Descriptive Statistics	
	Percentiles	
	50th (Median)	75th	
B_Creat_Kinase	107.2250	148.5400	
Diff_Creat_Kinase	10.7350	16.4450	
B_lactate_dehydr	236.0000	244.1750	
Diff_lactate_dehydr	21.5500	37.3750	
B_cortiscol	120.5400	186.8300	
Diff_cortiscol	13.0500	22.8425	
B_Ser_Ferritine	19.5850	42.3775	
Diff_Ser_Ferritine	-3.4050	13.1625	
B_Ser_Iron	63.8850	100.0550	
Diff_Ser_Iron	-28.0600	12.3325	
Groups	1.5000	2.0000	


Mann-Whitney Test


Ranks	
	Groups	N	Mean Rank	Sum of Ranks	
B_Creat_Kinase	1.00	32	32.75	1048.00	
	2.00	32	32.25	1032.00	
	Total	64			
Diff_Creat_Kinase	1.00	32	41.08	1314.50	
	2.00	32	23.92	765.50	
	Total	64			
B_lactate_dehydr	1.00	32	30.22	967.00	
	2.00	32	34.78	1113.00	
	Total	64			
Diff_lactate_dehydr	1.00	32	40.16	1285.00	
	2.00	32	24.84	795.00	
	Total	64			
B_cortiscol	1.00	32	32.52	1040.50	
	2.00	32	32.48	1039.50	
	Total	64			
Diff_cortiscol	1.00	32	38.45	1230.50	
	2.00	32	26.55	849.50	
	Total	64			
B_Ser_Ferritine	1.00	32	33.73	1079.50	
	2.00	32	31.27	1000.50	
	Total	64			
Diff_Ser_Ferritine	1.00	32	34.44	1102.00	
	2.00	32	30.56	978.00	
	Total	64			
B_Ser_Iron	1.00	32	35.05	1121.50	
	2.00	32	29.95	958.50	
	Total	64			
Diff_Ser_Iron	1.00	32	36.33	1162.50	
	2.00	32	28.67	917.50	
	Total	64			


Test Statistics^a^	
	B_Creat_Kinase	Diff_Creat_Kinase	B_lactate_dehydr	Diff_lactate_dehydr	
Mann-Whitney U	504.000	237.500	439.000	267.000	
Wilcoxon W	1032.000	765.500	967.000	795.000	
Z	-.107	-3.686	-.980	-3.290	
Asymp. Sig. (2-tailed)	.914	<.001	.327	.001	

Test Statistics^a^	
	B_cortiscol	Diff_cortiscol	B_Ser_Ferritine	Diff_Ser_Ferritine	B_Ser_Iron	
Mann-Whitney U	511.500	321.500	472.500	450.000	430.500	
Wilcoxon W	1039.500	849.500	1000.500	978.000	958.500	
Z	-.007	-2.558	-.530	-.832	-1.094	
Asymp. Sig. (2-tailed)	.995	.011	.596	.405	.274	

Test Statistics^a^	
	Diff_Ser_Iron	
Mann-Whitney U	389.500	
Wilcoxon W	917.500	
Z	-1.645	
Asymp. Sig. (2-tailed)	.100	

a. Grouping Variable: Groups	


NPar Tests


Notes	
Output Created	11-MAY-2026 14:34:03	
Comments		
Input	Active Dataset	DataSet0	
	Filter	<none>	
	Weight	<none>	
	Split File	<none>	
	N of Rows in Working Data File	64	
Missing Value Handling	Definition of Missing	User-defined missing values are treated as missing.	
	Cases Used	Statistics for each test are based on all cases with valid data for the variable(s) used in that test.	
Syntax	NPAR TESTS
  /WILCOXON=B_Creat_Kinase_1 B_lactate_dehydr_1 B_cortiscol_1 B_Ser_Ferritine_1 B_Ser_Iron_1
    B_Creat_Kinase_2 B_lactate_dehydr_2 B_cortiscol_2 B_Ser_Ferritine_2 B_Ser_Iron_2 WITH
    W12_Creat_Kinase_1 W12_lactate_dehydr_1 W12_cortiscol_1 W12_Ser_Ferritine_1 W12_Ser_Iron_1
    W12_Creat_Kinase_2 W12_lactate_dehydr_2 W12_cortiscol_2 W12_Ser_Ferritine_2 W12_Ser_Iron_2 (PAIRED)
  /STATISTICS DESCRIPTIVES QUARTILES
  /MISSING ANALYSIS.	
Resources	Processor Time	00:00:00.00	
	Elapsed Time	00:00:00.00	
	Number of Cases Allowed^a^	125829	

a. Based on availability of workspace memory.	


Descriptive Statistics	
	N	Mean	Std. Deviation	Minimum	Maximum	Percentiles	
						25th	
B_Creat_Kinase_1	32	122.5181	44.58493	49.20	210.59	94.6400	
B_lactate_dehydr_1	32	234.8819	22.70212	181.70	298.70	221.1000	
B_cortiscol_1	32	145.9662	53.29647	87.00	289.63	108.6300	
B_Ser_Ferritine_1	32	41.2441	46.59499	9.32	187.41	16.7150	
B_Ser_Iron_1	32	80.0272	36.75208	30.41	192.09	53.1800	
B_Creat_Kinase_2	32	118.0516	32.57951	56.32	198.00	93.2100	
B_lactate_dehydr_2	32	237.9303	50.93152	115.80	409.00	231.2075	
B_cortiscol_2	32	144.0147	48.70298	88.02	247.00	102.8800	
B_Ser_Ferritine_2	32	41.6381	57.83172	9.61	244.00	15.4575	
B_Ser_Iron_2	32	70.5472	36.74818	20.90	157.80	45.0675	
W12_Creat_Kinase_1	32	104.8706	42.66702	39.23	199.89	71.7850	
W12_lactate_dehydr_1	32	203.6878	21.67636	135.10	243.10	190.6000	
W12_cortiscol_1	32	127.4913	51.63015	84.70	274.21	89.6300	
W12_Ser_Ferritine_1	32	51.4975	63.07316	12.33	241.21	19.5250	
W12_Ser_Iron_1	32	97.9107	32.51386	58.00	160.14	65.1700	
W12_Creat_Kinase_2	32	111.0275	30.84211	51.93	183.09	91.1000	
W12_lactate_dehydr_2	32	218.3431	53.44814	102.30	394.10	198.2000	
W12_cortiscol_2	32	132.2741	50.20841	86.70	256.63	90.9725	
W12_Ser_Ferritine_2	32	60.2050	62.12594	12.10	254.10	18.8775	
W12_Ser_Iron_2	32	105.8822	37.69460	53.65	170.25	65.5575	

Descriptive Statistics	
	Percentiles	
	50th (Median)	75th	
B_Creat_Kinase_1	105.3100	162.7375	
B_lactate_dehydr_1	235.2500	240.6750	
B_cortiscol_1	120.9950	186.8300	
B_Ser_Ferritine_1	20.6500	42.3775	
B_Ser_Iron_1	77.7150	100.0550	
B_Creat_Kinase_2	110.9250	128.1850	
B_lactate_dehydr_2	238.2600	248.1250	
B_cortiscol_2	120.2550	187.0775	
B_Ser_Ferritine_2	19.3050	42.2250	
B_Ser_Iron_2	60.0200	100.0850	
W12_Creat_Kinase_1	91.0950	137.7525	
W12_lactate_dehydr_1	200.2700	214.5500	
W12_cortiscol_1	102.4100	165.0850	
W12_Ser_Ferritine_1	20.4900	50.4650	
W12_Ser_Iron_1	89.3150	121.3575	
W12_Creat_Kinase_2	101.7050	118.7850	
W12_lactate_dehydr_2	226.6000	237.7250	
W12_cortiscol_2	109.5650	187.0400	
W12_Ser_Ferritine_2	26.9250	102.5000	
W12_Ser_Iron_2	110.7750	140.0100	


Wilcoxon Signed Ranks Test


Ranks	
	N	Mean Rank	Sum of Ranks	
W12_Creat_Kinase_1 - B_Creat_Kinase_1	Negative Ranks	32^a^	16.50	528.00	
	Positive Ranks	0^b^	.00	.00	
	Ties	0^c^			
	Total	32			
W12_lactate_dehydr_1 - B_lactate_dehydr_1	Negative Ranks	30^d^	17.50	525.00	
	Positive Ranks	2^e^	1.50	3.00	
	Ties	0^f^			
	Total	32			
W12_cortiscol_1 - B_cortiscol_1	Negative Ranks	30^g^	17.03	511.00	
	Positive Ranks	2^h^	8.50	17.00	
	Ties	0^i^			
	Total	32			
W12_Ser_Ferritine_1 - B_Ser_Ferritine_1	Negative Ranks	16^j^	15.19	243.00	
	Positive Ranks	16^k^	17.81	285.00	
	Ties	0^l^			
	Total	32			
W12_Ser_Iron_1 - B_Ser_Iron_1	Negative Ranks	14^m^	12.43	174.00	
	Positive Ranks	18^n^	19.67	354.00	
	Ties	0^o^			
	Total	32			
W12_Creat_Kinase_2 - B_Creat_Kinase_2	Negative Ranks	28^p^	16.82	471.00	
	Positive Ranks	4^q^	14.25	57.00	
	Ties	0^r^			
	Total	32			
W12_lactate_dehydr_2 - B_lactate_dehydr_2	Negative Ranks	26^s^	18.65	485.00	
	Positive Ranks	6^t^	7.17	43.00	
	Ties	0^u^			
	Total	32			
W12_cortiscol_2 - B_cortiscol_2	Negative Ranks	23^v^	16.87	388.00	
	Positive Ranks	9^w^	15.56	140.00	
	Ties	0^x^			
	Total	32			
W12_Ser_Ferritine_2 - B_Ser_Ferritine_2	Negative Ranks	12^y^	15.67	188.00	
	Positive Ranks	20^z^	17.00	340.00	
	Ties	0^aa^			
	Total	32			
W12_Ser_Iron_2 - B_Ser_Iron_2	Negative Ranks	6^ab^	9.67	58.00	
	Positive Ranks	26^ac^	18.08	470.00	
	Ties	0^ad^			
	Total	32			

a. W12_Creat_Kinase_1 < B_Creat_Kinase_1	
b. W12_Creat_Kinase_1 > B_Creat_Kinase_1	
c. W12_Creat_Kinase_1 = B_Creat_Kinase_1	
d. W12_lactate_dehydr_1 < B_lactate_dehydr_1	
e. W12_lactate_dehydr_1 > B_lactate_dehydr_1	
f. W12_lactate_dehydr_1 = B_lactate_dehydr_1	
g. W12_cortiscol_1 < B_cortiscol_1	
h. W12_cortiscol_1 > B_cortiscol_1	
i. W12_cortiscol_1 = B_cortiscol_1	
j. W12_Ser_Ferritine_1 < B_Ser_Ferritine_1	
k. W12_Ser_Ferritine_1 > B_Ser_Ferritine_1	
l. W12_Ser_Ferritine_1 = B_Ser_Ferritine_1	
m. W12_Ser_Iron_1 < B_Ser_Iron_1	
n. W12_Ser_Iron_1 > B_Ser_Iron_1	
o. W12_Ser_Iron_1 = B_Ser_Iron_1	
p. W12_Creat_Kinase_2 < B_Creat_Kinase_2	
q. W12_Creat_Kinase_2 > B_Creat_Kinase_2	
r. W12_Creat_Kinase_2 = B_Creat_Kinase_2	
s. W12_lactate_dehydr_2 < B_lactate_dehydr_2	
t. W12_lactate_dehydr_2 > B_lactate_dehydr_2	
u. W12_lactate_dehydr_2 = B_lactate_dehydr_2	
v. W12_cortiscol_2 < B_cortiscol_2	
w. W12_cortiscol_2 > B_cortiscol_2	
x. W12_cortiscol_2 = B_cortiscol_2	
y. W12_Ser_Ferritine_2 < B_Ser_Ferritine_2	
z. W12_Ser_Ferritine_2 > B_Ser_Ferritine_2	
aa. W12_Ser_Ferritine_2 = B_Ser_Ferritine_2	
ab. W12_Ser_Iron_2 < B_Ser_Iron_2	
ac. W12_Ser_Iron_2 > B_Ser_Iron_2	
ad. W12_Ser_Iron_2 = B_Ser_Iron_2	


Test Statistics^a^	
	W12_Creat_Kinase_1 - B_Creat_Kinase_1	W12_lactate_dehydr_1 - B_lactate_dehydr_1	W12_cortiscol_1 - B_cortiscol_1	W12_Ser_Ferritine_1 - B_Ser_Ferritine_1	
Z	-4.937^b^	-4.881^b^	-4.619^b^	-.393^c^	
Asymp. Sig. (2-tailed)	<.001	<.001	<.001	.695	

Test Statistics^a^	
	W12_Ser_Iron_1 - B_Ser_Iron_1	W12_Creat_Kinase_2 - B_Creat_Kinase_2	W12_lactate_dehydr_2 - B_lactate_dehydr_2	W12_cortiscol_2 - B_cortiscol_2	
Z	-1.683^c^	-3.871^b^	-4.133^b^	-2.319^b^	
Asymp. Sig. (2-tailed)	.092	<.001	<.001	.020	

Test Statistics^a^	
	W12_Ser_Ferritine_2 - B_Ser_Ferritine_2	W12_Ser_Iron_2 - B_Ser_Iron_2	
Z	-1.421^c^	-3.852^c^	
Asymp. Sig. (2-tailed)	.155	<.001	

a. Wilcoxon Signed Ranks Test	
b. Based on positive ranks.	
c. Based on negative ranks.	
