## Supplementary material for "Effects of a standardised *Amaranthus tricolor*–*Spinacia oleracea* extract on exercise performance, muscle strength, endurance and recovery in physically active adults: a randomised, double-blind, placebo-controlled trial": Raw statistical outputs: CBC_normal data_out- By ANCOVA.rtf

Univariate Analysis of Variance


Notes	
Output Created	11-MAY-2026 12:13:04	
Comments		
Input	Active Dataset	DataSet0	
	Filter	<none>	
	Weight	<none>	
	Split File	<none>	
	N of Rows in Working Data File	64	
Missing Value Handling	Definition of Missing	User-defined missing values are treated as missing.	
	Cases Used	Statistics are based on all cases with valid data for all variables in the model.	
Syntax	UNIANOVA W12_neutro BY Groups WITH B_Neutro
  /METHOD=SSTYPE(3)
  /INTERCEPT=INCLUDE
  /EMMEANS=TABLES(Groups) WITH(B_Neutro=MEAN) COMPARE ADJ(BONFERRONI)
  /PRINT ETASQ DESCRIPTIVE
  /CRITERIA=ALPHA(.05)
  /DESIGN=B_Neutro Groups.	
Resources	Processor Time	00:00:00.00	
	Elapsed Time	00:00:00.00	


Between-Subjects Factors	
	N	
Groups	1	32	
	2	32	


Descriptive Statistics	
Dependent Variable:   W12_neutro  	
Groups	Mean	Std. Deviation	N	
1	69.5937	6.37464	32	
2	66.7813	8.16655	32	
Total	68.1875	7.40415	64	


Tests of Between-Subjects Effects	
Dependent Variable:   W12_neutro  	
Source	Type III Sum of Squares	df	Mean Square	F	Sig.	Partial Eta Squared	
Corrected Model	303.159^a^	2	151.580	2.935	.061	.088	
Intercept	3543.238	1	3543.238	68.602	<.001	.529	
B_Neutro	176.597	1	176.597	3.419	.069	.053	
Groups	67.357	1	67.357	1.304	.258	.021	
Error	3150.591	61	51.649				
Total	301024.000	64					
Corrected Total	3453.750	63					

a. R Squared = .088 (Adjusted R Squared = .058)	


Estimated Marginal Means


Groups


Estimates	
Dependent Variable:   W12_neutro  	
Groups	Mean	Std. Error	95% Confidence Interval	
			Lower Bound	Upper Bound	
1	69.237^a^	1.285	66.667	71.806	
2	67.138^a^	1.285	64.569	69.708	

a. Covariates appearing in the model are evaluated at the following values: B_Neutro = 66.5156.	


Pairwise Comparisons	
Dependent Variable:   W12_neutro  	
(I) Groups	(J) Groups	Mean Difference (I-J)	Std. Error	Sig.^a^	95% Confidence Interval for Difference^a^	
					Lower Bound	Upper Bound	
1	2	2.099	1.838	.258	-1.576	5.773	
2	1	-2.099	1.838	.258	-5.773	1.576	

Based on estimated marginal means	
a. Adjustment for multiple comparisons: Bonferroni.	


Univariate Tests	
Dependent Variable:   W12_neutro  	
	Sum of Squares	df	Mean Square	F	Sig.	Partial Eta Squared	
Contrast	67.357	1	67.357	1.304	.258	.021	
Error	3150.591	61	51.649				

The F tests the effect of Groups. This test is based on the linearly independent pairwise comparisons among the estimated marginal means.	


Univariate Analysis of Variance


Notes	
Output Created	11-MAY-2026 12:13:16	
Comments		
Input	Active Dataset	DataSet0	
	Filter	<none>	
	Weight	<none>	
	Split File	<none>	
	N of Rows in Working Data File	64	
Missing Value Handling	Definition of Missing	User-defined missing values are treated as missing.	
	Cases Used	Statistics are based on all cases with valid data for all variables in the model.	
Syntax	UNIANOVA W12_Total_RBC BY Groups WITH B_Total_RBC
  /METHOD=SSTYPE(3)
  /INTERCEPT=INCLUDE
  /EMMEANS=TABLES(Groups) WITH(B_Total_RBC=MEAN) COMPARE ADJ(BONFERRONI)
  /PRINT ETASQ DESCRIPTIVE
  /CRITERIA=ALPHA(.05)
  /DESIGN=B_Total_RBC Groups.	
Resources	Processor Time	00:00:00.02	
	Elapsed Time	00:00:00.01	


Between-Subjects Factors	
	N	
Groups	1	32	
	2	32	


Descriptive Statistics	
Dependent Variable:   W12_Total_RBC  	
Groups	Mean	Std. Deviation	N	
1	5.2991	.55490	32	
2	5.2063	.58350	32	
Total	5.2527	.56678	64	


Tests of Between-Subjects Effects	
Dependent Variable:   W12_Total_RBC  	
Source	Type III Sum of Squares	df	Mean Square	F	Sig.	Partial Eta Squared	
Corrected Model	1.348^a^	2	.674	2.176	.122	.067	
Intercept	14.649	1	14.649	47.307	<.001	.437	
B_Total_RBC	1.210	1	1.210	3.908	.053	.060	
Groups	.171	1	.171	.552	.460	.009	
Error	18.890	61	.310				
Total	1786.023	64					
Corrected Total	20.238	63					

a. R Squared = .067 (Adjusted R Squared = .036)	


Estimated Marginal Means


Groups


Estimates	
Dependent Variable:   W12_Total_RBC  	
Groups	Mean	Std. Error	95% Confidence Interval	
			Lower Bound	Upper Bound	
1	5.304^a^	.098	5.108	5.501	
2	5.201^a^	.098	5.004	5.398	

a. Covariates appearing in the model are evaluated at the following values: B_Total_RBC = 5.2311.	


Pairwise Comparisons	
Dependent Variable:   W12_Total_RBC  	
(I) Groups	(J) Groups	Mean Difference (I-J)	Std. Error	Sig.^a^	95% Confidence Interval for Difference^a^	
					Lower Bound	Upper Bound	
1	2	.103	.139	.460	-.175	.382	
2	1	-.103	.139	.460	-.382	.175	

Based on estimated marginal means	
a. Adjustment for multiple comparisons: Bonferroni.	


Univariate Tests	
Dependent Variable:   W12_Total_RBC  	
	Sum of Squares	df	Mean Square	F	Sig.	Partial Eta Squared	
Contrast	.171	1	.171	.552	.460	.009	
Error	18.890	61	.310				

The F tests the effect of Groups. This test is based on the linearly independent pairwise comparisons among the estimated marginal means.	


Univariate Analysis of Variance


Notes	
Output Created	11-MAY-2026 12:13:30	
Comments		
Input	Active Dataset	DataSet0	
	Filter	<none>	
	Weight	<none>	
	Split File	<none>	
	N of Rows in Working Data File	64	
Missing Value Handling	Definition of Missing	User-defined missing values are treated as missing.	
	Cases Used	Statistics are based on all cases with valid data for all variables in the model.	
Syntax	UNIANOVA W12_Heamato BY Groups WITH B_Heamato
  /METHOD=SSTYPE(3)
  /INTERCEPT=INCLUDE
  /EMMEANS=TABLES(Groups) WITH(B_Heamato=MEAN) COMPARE ADJ(BONFERRONI)
  /PRINT ETASQ DESCRIPTIVE
  /CRITERIA=ALPHA(.05)
  /DESIGN=B_Heamato Groups.	
Resources	Processor Time	00:00:00.00	
	Elapsed Time	00:00:00.01	


Between-Subjects Factors	
	N	
Groups	1	32	
	2	32	


Descriptive Statistics	
Dependent Variable:   W12_Heamato  	
Groups	Mean	Std. Deviation	N	
1	42.1719	3.69535	32	
2	41.8719	3.83593	32	
Total	42.0219	3.73934	64	


Tests of Between-Subjects Effects	
Dependent Variable:   W12_Heamato  	
Source	Type III Sum of Squares	df	Mean Square	F	Sig.	Partial Eta Squared	
Corrected Model	68.239^a^	2	34.120	2.561	.086	.077	
Intercept	888.608	1	888.608	66.700	<.001	.522	
B_Heamato	66.799	1	66.799	5.014	.029	.076	
Groups	1.211	1	1.211	.091	.764	.001	
Error	812.670	61	13.322				
Total	113894.540	64					
Corrected Total	880.909	63					

a. R Squared = .077 (Adjusted R Squared = .047)	


Estimated Marginal Means


Groups


Estimates	
Dependent Variable:   W12_Heamato  	
Groups	Mean	Std. Error	95% Confidence Interval	
			Lower Bound	Upper Bound	
1	42.159^a^	.645	40.869	43.450	
2	41.884^a^	.645	40.594	43.175	

a. Covariates appearing in the model are evaluated at the following values: B_Heamato = 42.8875.	


Pairwise Comparisons	
Dependent Variable:   W12_Heamato  	
(I) Groups	(J) Groups	Mean Difference (I-J)	Std. Error	Sig.^a^	95% Confidence Interval for Difference^a^	
					Lower Bound	Upper Bound	
1	2	.275	.913	.764	-1.550	2.100	
2	1	-.275	.913	.764	-2.100	1.550	

Based on estimated marginal means	
a. Adjustment for multiple comparisons: Bonferroni.	


Univariate Tests	
Dependent Variable:   W12_Heamato  	
	Sum of Squares	df	Mean Square	F	Sig.	Partial Eta Squared	
Contrast	1.211	1	1.211	.091	.764	.001	
Error	812.670	61	13.322				

The F tests the effect of Groups. This test is based on the linearly independent pairwise comparisons among the estimated marginal means.	


Univariate Analysis of Variance


Notes	
Output Created	11-MAY-2026 12:13:45	
Comments		
Input	Active Dataset	DataSet0	
	Filter	<none>	
	Weight	<none>	
	Split File	<none>	
	N of Rows in Working Data File	64	
Missing Value Handling	Definition of Missing	User-defined missing values are treated as missing.	
	Cases Used	Statistics are based on all cases with valid data for all variables in the model.	
Syntax	UNIANOVA W12_platelets BY Groups WITH B_platelets
  /METHOD=SSTYPE(3)
  /INTERCEPT=INCLUDE
  /EMMEANS=TABLES(Groups) WITH(B_platelets=MEAN) COMPARE ADJ(BONFERRONI)
  /PRINT ETASQ DESCRIPTIVE
  /CRITERIA=ALPHA(.05)
  /DESIGN=B_platelets Groups.	
Resources	Processor Time	00:00:00.00	
	Elapsed Time	00:00:00.00	


Between-Subjects Factors	
	N	
Groups	1	32	
	2	32	


Descriptive Statistics	
Dependent Variable:   W12_platelets  	
Groups	Mean	Std. Deviation	N	
1	289.3750	82.83592	32	
2	302.5313	74.65317	32	
Total	295.9531	78.50294	64	


Tests of Between-Subjects Effects	
Dependent Variable:   W12_platelets  	
Source	Type III Sum of Squares	df	Mean Square	F	Sig.	Partial Eta Squared	
Corrected Model	134781.678^a^	2	67390.839	16.218	<.001	.347	
Intercept	49368.603	1	49368.603	11.881	.001	.163	
B_platelets	132012.288	1	132012.288	31.770	<.001	.342	
Groups	8095.979	1	8095.979	1.948	.168	.031	
Error	253469.181	61	4155.232				
Total	5993899.000	64					
Corrected Total	388250.859	63					

a. R Squared = .347 (Adjusted R Squared = .326)	


Estimated Marginal Means


Groups


Estimates	
Dependent Variable:   W12_platelets  	
Groups	Mean	Std. Error	95% Confidence Interval	
			Lower Bound	Upper Bound	
1	284.645^a^	11.426	261.797	307.493	
2	307.261^a^	11.426	284.413	330.109	

a. Covariates appearing in the model are evaluated at the following values: B_platelets = 287.2344.	


Pairwise Comparisons	
Dependent Variable:   W12_platelets  	
(I) Groups	(J) Groups	Mean Difference (I-J)	Std. Error	Sig.^a^	95% Confidence Interval for Difference^a^	
					Lower Bound	Upper Bound	
1	2	-22.616	16.202	.168	-55.015	9.783	
2	1	22.616	16.202	.168	-9.783	55.015	

Based on estimated marginal means	
a. Adjustment for multiple comparisons: Bonferroni.	


Univariate Tests	
Dependent Variable:   W12_platelets  	
	Sum of Squares	df	Mean Square	F	Sig.	Partial Eta Squared	
Contrast	8095.979	1	8095.979	1.948	.168	.031	
Error	253469.181	61	4155.232				

The F tests the effect of Groups. This test is based on the linearly independent pairwise comparisons among the estimated marginal means.	


T-Test


Notes	
Output Created	11-MAY-2026 12:15:21	
Comments		
Input	Active Dataset	DataSet0	
	Filter	<none>	
	Weight	<none>	
	Split File	<none>	
	N of Rows in Working Data File	64	
Missing Value Handling	Definition of Missing	User defined missing values are treated as missing.	
	Cases Used	Statistics for each analysis are based on the cases with no missing or out-of-range data for any variable in the analysis.	
Syntax	T-TEST PAIRS=B_Neutro_1 B_Total_RBC_1 B_Heamato_1 B_platelets_1 B_Neutro_2 B_Total_RBC_2
    B_Heamato_2 B_platelets_2 WITH W12_Neutro_1 W12_Total_RBC_1 W12_Heamato_1 W12_platelets_1
    W12_Neutro_2 W12_Total_RBC_2 W12_Heamato_2 W12_platelets_2 (PAIRED)
  /ES DISPLAY(TRUE) STANDARDIZER(SD)
  /CRITERIA=CI(.9500)
  /MISSING=ANALYSIS.	
Resources	Processor Time	00:00:00.02	
	Elapsed Time	00:00:00.01	


Paired Samples Statistics	
	Mean	N	Std. Deviation	Std. Error Mean	
Pair 1	B_Neutro_1	68.4375	32	7.78310	1.37587	
	W12_Neutro_1	69.5938	32	6.37464	1.12689	
Pair 2	B_Total_RBC_1	5.2072	32	.46133	.08155	
	W12_Total_RBC_1	5.2991	32	.55490	.09809	
Pair 3	B_Heamato_1	42.9469	32	4.56211	.80648	
	W12_Heamato_1	42.1719	32	3.69535	.65325	
Pair 4	B_platelets_1	294.7188	32	81.01304	14.32122	
	W12_platelets_1	289.3750	32	82.83592	14.64346	
Pair 5	B_Neutro_2	64.5938	32	10.22602	1.80772	
	W12_Neutro_2	66.7813	32	8.16655	1.44366	
Pair 6	B_Total_RBC_2	5.2550	32	.75656	.13374	
	W12_Total_RBC_2	5.2063	32	.58350	.10315	
Pair 7	B_Heamato_2	42.8281	32	5.30588	.93796	
	W12_Heamato_2	41.8719	32	3.83593	.67810	
Pair 8	B_platelets_2	279.7500	32	64.02620	11.31834	
	W12_platelets_2	302.5313	32	74.65317	13.19694	


Paired Samples Correlations	
	N	Correlation	Significance	
			One-Sided p	Two-Sided p	
Pair 1	B_Neutro_1 & W12_Neutro_1	32	.393	.013	.026	
Pair 2	B_Total_RBC_1 & W12_Total_RBC_1	32	.299	.048	.096	
Pair 3	B_Heamato_1 & W12_Heamato_1	32	.479	.003	.006	
Pair 4	B_platelets_1 & W12_platelets_1	32	.728	<.001	<.001	
Pair 5	B_Neutro_2 & W12_Neutro_2	32	.134	.232	.464	
Pair 6	B_Total_RBC_2 & W12_Total_RBC_2	32	.223	.110	.220	
Pair 7	B_Heamato_2 & W12_Heamato_2	32	.108	.278	.557	
Pair 8	B_platelets_2 & W12_platelets_2	32	.387	.014	.029	


Paired Samples Test	
	Paired Differences	
	Mean	Std. Deviation	Std. Error Mean	95% Confidence Interval of the Difference	
				Lower	
Pair 1	B_Neutro_1 - W12_Neutro_1	-1.15625	7.89083	1.39491	-4.00120	
Pair 2	B_Total_RBC_1 - W12_Total_RBC_1	-.09188	.60628	.10718	-.31046	
Pair 3	B_Heamato_1 - W12_Heamato_1	.77500	4.27943	.75650	-.76790	
Pair 4	B_platelets_1 - W12_platelets_1	5.34375	60.40060	10.67742	-16.43299	
Pair 5	B_Neutro_2 - W12_Neutro_2	-2.18750	12.20110	2.15687	-6.58647	
Pair 6	B_Total_RBC_2 - W12_Total_RBC_2	.04875	.84609	.14957	-.25630	
Pair 7	B_Heamato_2 - W12_Heamato_2	.95625	6.20307	1.09656	-1.28019	
Pair 8	B_platelets_2 - W12_platelets_2	-22.78125	77.28111	13.66150	-50.64406	

Paired Samples Test	
	Paired Differences	t	df	Significance	
	95% Confidence Interval of the Difference			One-Sided p	
	Upper				
Pair 1	B_Neutro_1 - W12_Neutro_1	1.68870	-.829	31	.207	
Pair 2	B_Total_RBC_1 - W12_Total_RBC_1	.12671	-.857	31	.199	
Pair 3	B_Heamato_1 - W12_Heamato_1	2.31790	1.024	31	.157	
Pair 4	B_platelets_1 - W12_platelets_1	27.12049	.500	31	.310	
Pair 5	B_Neutro_2 - W12_Neutro_2	2.21147	-1.014	31	.159	
Pair 6	B_Total_RBC_2 - W12_Total_RBC_2	.35380	.326	31	.373	
Pair 7	B_Heamato_2 - W12_Heamato_2	3.19269	.872	31	.195	
Pair 8	B_platelets_2 - W12_platelets_2	5.08156	-1.668	31	.053	

Paired Samples Test	
	Significance	
	Two-Sided p	
		
Pair 1	B_Neutro_1 - W12_Neutro_1	.413	
Pair 2	B_Total_RBC_1 - W12_Total_RBC_1	.398	
Pair 3	B_Heamato_1 - W12_Heamato_1	.314	
Pair 4	B_platelets_1 - W12_platelets_1	.620	
Pair 5	B_Neutro_2 - W12_Neutro_2	.318	
Pair 6	B_Total_RBC_2 - W12_Total_RBC_2	.747	
Pair 7	B_Heamato_2 - W12_Heamato_2	.390	
Pair 8	B_platelets_2 - W12_platelets_2	.105	


Paired Samples Effect Sizes	
	Standardizer^a^	Point Estimate	95% Confidence Interval	
			Lower	
Pair 1	B_Neutro_1 - W12_Neutro_1	Cohen's d	7.89083	-.147	-.494	
		Hedges' correction	8.08837	-.143	-.482	
Pair 2	B_Total_RBC_1 - W12_Total_RBC_1	Cohen's d	.60628	-.152	-.499	
		Hedges' correction	.62145	-.148	-.487	
Pair 3	B_Heamato_1 - W12_Heamato_1	Cohen's d	4.27943	.181	-.170	
		Hedges' correction	4.38657	.177	-.166	
Pair 4	B_platelets_1 - W12_platelets_1	Cohen's d	60.40060	.088	-.259	
		Hedges' correction	61.91273	.086	-.253	
Pair 5	B_Neutro_2 - W12_Neutro_2	Cohen's d	12.20110	-.179	-.527	
		Hedges' correction	12.50656	-.175	-.514	
Pair 6	B_Total_RBC_2 - W12_Total_RBC_2	Cohen's d	.84609	.058	-.290	
		Hedges' correction	.86727	.056	-.283	
Pair 7	B_Heamato_2 - W12_Heamato_2	Cohen's d	6.20307	.154	-.196	
		Hedges' correction	6.35836	.150	-.191	
Pair 8	B_platelets_2 - W12_platelets_2	Cohen's d	77.28111	-.295	-.647	
		Hedges' correction	79.21584	-.288	-.631	

Paired Samples Effect Sizes	
	95% Confidence Interval^a^	
	Upper	
Pair 1	B_Neutro_1 - W12_Neutro_1	Cohen's d	.203	
		Hedges' correction	.198	
Pair 2	B_Total_RBC_1 - W12_Total_RBC_1	Cohen's d	.198	
		Hedges' correction	.193	
Pair 3	B_Heamato_1 - W12_Heamato_1	Cohen's d	.529	
		Hedges' correction	.516	
Pair 4	B_platelets_1 - W12_platelets_1	Cohen's d	.435	
		Hedges' correction	.424	
Pair 5	B_Neutro_2 - W12_Neutro_2	Cohen's d	.171	
		Hedges' correction	.167	
Pair 6	B_Total_RBC_2 - W12_Total_RBC_2	Cohen's d	.404	
		Hedges' correction	.394	
Pair 7	B_Heamato_2 - W12_Heamato_2	Cohen's d	.502	
		Hedges' correction	.489	
Pair 8	B_platelets_2 - W12_platelets_2	Cohen's d	.062	
		Hedges' correction	.060	

a. The denominator used in estimating the effect sizes. 
Cohen's d uses the sample standard deviation of the mean difference. 
Hedges' correction uses the sample standard deviation of the mean difference, plus a correction factor.	
