## Supplementary material for "Effects of a standardised *Amaranthus tricolor*–*Spinacia oleracea* extract on exercise performance, muscle strength, endurance and recovery in physically active adults: a randomised, double-blind, placebo-controlled trial": Raw statistical outputs: CBC_Output- Non parametric tests.rtf

NPar Tests


Notes	
Output Created	11-MAY-2026 14:45:03	
Comments		
Input	Active Dataset	DataSet0	
	Filter	<none>	
	Weight	<none>	
	Split File	<none>	
	N of Rows in Working Data File	64	
Missing Value Handling	Definition of Missing	User-defined missing values are treated as missing.	
	Cases Used	Statistics for each test are based on all cases with valid data for the variable(s) used in that test.	
Syntax	NPAR TESTS
  /M-W= B_Total_Leuco Diff_Total_Leuco B_Lympho Diff_Lympho B_Mono Diff_Mono B_Eosino Diff_Eosino
    B_Baso Diff_Baso B_Heamoglobin Diff_Heamoglobin BY Groups(1 2)
  /STATISTICS=DESCRIPTIVES QUARTILES
  /MISSING ANALYSIS.	
Resources	Processor Time	00:00:00.00	
	Elapsed Time	00:00:00.00	
	Number of Cases Allowed^a^	174762	

a. Based on availability of workspace memory.	


Descriptive Statistics	
	N	Mean	Std. Deviation	Minimum	Maximum	Percentiles	
						25th	
B_Total_Leuco	64	7992.1563	1611.83325	3510.00	10810.00	6875.0000	
Diff_Total_Leuco	64	385.4375	2006.44315	-4770.00	5800.00	-722.5000	
B_Lympho	64	30.3594	8.02450	14.00	50.00	25.0000	
Diff_Lympho	64	2.7813	7.46094	-11.00	22.00	-2.5000	
B_Mono	64	2.3281	.50567	2.00	4.00	2.0000	
Diff_Mono	64	-.3750	1.40859	-8.00	2.00	-1.0000	
B_Eosino	64	1.2969	.49376	1.00	3.00	1.0000	
Diff_Eosino	64	-.3437	.82074	-3.00	2.00	-1.0000	
B_Baso	64	.0000	.00000	.00	.00	.0000	
Diff_Baso	64	.0000	.00000	.00	.00	.0000	
B_Heamoglobin	64	14.3875	.93546	12.80	16.80	13.6250	
Diff_Heamoglobin	64	.1062	.57235	-1.60	1.90	-.3000	
Groups	64	1.5000	.50395	1.00	2.00	1.0000	

Descriptive Statistics	
	Percentiles	
	50th (Median)	75th	
B_Total_Leuco	7979.5000	9435.0000	
Diff_Total_Leuco	225.0000	1087.5000	
B_Lympho	30.0000	36.7500	
Diff_Lympho	2.5000	7.0000	
B_Mono	2.0000	3.0000	
Diff_Mono	.0000	.0000	
B_Eosino	1.0000	2.0000	
Diff_Eosino	.0000	.0000	
B_Baso	.0000	.0000	
Diff_Baso	.0000	.0000	
B_Heamoglobin	14.3000	14.8750	
Diff_Heamoglobin	.0000	.2750	
Groups	1.5000	2.0000	


Mann-Whitney Test


Ranks	
	Groups	N	Mean Rank	Sum of Ranks	
B_Total_Leuco	1.00	32	36.80	1177.50	
	2.00	32	28.20	902.50	
	Total	64			
Diff_Total_Leuco	1.00	32	36.34	1163.00	
	2.00	32	28.66	917.00	
	Total	64			
B_Lympho	1.00	32	29.27	936.50	
	2.00	32	35.73	1143.50	
	Total	64			
Diff_Lympho	1.00	32	32.48	1039.50	
	2.00	32	32.52	1040.50	
	Total	64			
B_Mono	1.00	32	30.38	972.00	
	2.00	32	34.63	1108.00	
	Total	64			
Diff_Mono	1.00	32	30.70	982.50	
	2.00	32	34.30	1097.50	
	Total	64			
B_Eosino	1.00	32	30.39	972.50	
	2.00	32	34.61	1107.50	
	Total	64			
Diff_Eosino	1.00	32	32.38	1036.00	
	2.00	32	32.63	1044.00	
	Total	64			
B_Baso	1.00	32	32.50	1040.00	
	2.00	32	32.50	1040.00	
	Total	64			
Diff_Baso	1.00	32	32.50	1040.00	
	2.00	32	32.50	1040.00	
	Total	64			
B_Heamoglobin	1.00	32	33.59	1075.00	
	2.00	32	31.41	1005.00	
	Total	64			
Diff_Heamoglobin	1.00	32	32.53	1041.00	
	2.00	32	32.47	1039.00	
	Total	64			


Test Statistics^a^	
	B_Total_Leuco	Diff_Total_Leuco	B_Lympho	Diff_Lympho	B_Mono	
Mann-Whitney U	374.500	389.000	408.500	511.500	444.000	
Wilcoxon W	902.500	917.000	936.500	1039.500	972.000	
Z	-1.846	-1.652	-1.393	-.007	-1.133	
Asymp. Sig. (2-tailed)	.065	.099	.164	.995	.257	

Test Statistics^a^	
	Diff_Mono	B_Eosino	Diff_Eosino	B_Baso	Diff_Baso	B_Heamoglobin	
Mann-Whitney U	454.500	444.500	508.000	512.000	512.000	477.000	
Wilcoxon W	982.500	972.500	1036.000	1040.000	1040.000	1005.000	
Z	-.818	-1.160	-.059	.000	.000	-.471	
Asymp. Sig. (2-tailed)	.413	.246	.953	1.000	1.000	.638	

Test Statistics^a^	
	Diff_Heamoglobin	
Mann-Whitney U	511.000	
Wilcoxon W	1039.000	
Z	-.014	
Asymp. Sig. (2-tailed)	.989	

a. Grouping Variable: Groups	


NPar Tests


Notes	
Output Created	11-MAY-2026 14:46:31	
Comments		
Input	Active Dataset	DataSet0	
	Filter	<none>	
	Weight	<none>	
	Split File	<none>	
	N of Rows in Working Data File	64	
Missing Value Handling	Definition of Missing	User-defined missing values are treated as missing.	
	Cases Used	Statistics for each test are based on all cases with valid data for the variable(s) used in that test.	
Syntax	NPAR TESTS
  /WILCOXON=B_Total_Leuco_1 B_Lympho_1 B_Mono_1 B_Eosino_1 B_Baso_1 B_Heamoglobin_1 B_Total_Leuco_2
    B_Lympho_2 B_Mono_2 B_Eosino_2 B_Baso_2 B_Heamoglobin_2 WITH W12_Total_Leuco_1 W12_Lympho_1
    W12_Mono_1 W12_Eosino_1 W12_Baso_1 W12_Heamoglobin_1 W12_Total_Leuco_2 W12_Lympho_2 W12_Mono_2
    W12_Eosino_2 W12_Baso_2 W12_Heamoglobin_2 (PAIRED)
  /STATISTICS DESCRIPTIVES QUARTILES
  /MISSING ANALYSIS.	
Resources	Processor Time	00:00:00.00	
	Elapsed Time	00:00:00.00	
	Number of Cases Allowed^a^	108473	

a. Based on availability of workspace memory.	


Descriptive Statistics	
	N	Mean	Std. Deviation	Minimum	Maximum	Percentiles	
						25th	
B_Total_Leuco_1	32	8419.3438	1524.44223	5830.00	10810.00	6860.0000	
B_Lympho_1	32	29.0938	6.10320	21.00	42.00	24.2500	
B_Mono_1	32	2.2500	.43994	2.00	3.00	2.0000	
B_Eosino_1	32	1.2188	.42001	1.00	2.00	1.0000	
B_Baso_1	32	.0000	.00000	.00	.00	.0000	
B_Heamoglobin_1	32	14.4563	.91895	13.30	16.30	13.7000	
B_Total_Leuco_2	32	7564.9687	1605.97123	3510.00	10510.00	6875.0000	
B_Lympho_2	32	31.6250	9.50297	14.00	50.00	25.2500	
B_Mono_2	32	2.4063	.55992	2.00	4.00	2.0000	
B_Eosino_2	32	1.3750	.55358	1.00	3.00	1.0000	
B_Baso_2	32	.0000	.00000	.00	.00	.0000	
B_Heamoglobin_2	32	14.3188	.96133	12.80	16.80	13.6000	
W12_Total_Leuco_1	32	7607.8125	1985.71143	4440.00	11390.00	5757.5000	
W12_Lympho_1	32	26.6562	6.28675	18.00	40.00	21.0000	
W12_Mono_1	32	2.5625	.66901	2.00	4.00	2.0000	
W12_Eosino_1	32	1.5625	.61892	1.00	3.00	1.0000	
W12_Baso_1	32	.0000	.00000	.00	.00	.0000	
W12_Heamoglobin_1	32	14.3375	.84653	12.30	16.30	13.8000	
W12_Total_Leuco_2	32	7605.6250	1994.58127	2800.00	10940.00	6610.0000	
W12_Lympho_2	32	28.5000	6.12767	17.00	44.00	25.0000	
W12_Mono_2	32	2.8438	1.62856	2.00	10.00	2.0000	
W12_Eosino_2	32	1.7187	.68318	1.00	4.00	1.0000	
W12_Baso_2	32	.0000	.00000	.00	.00	.0000	
W12_Heamoglobin_2	32	14.2250	.89587	12.80	15.90	13.5250	

Descriptive Statistics	
	Percentiles	
	50th (Median)	75th	
B_Total_Leuco_1	8545.0000	9712.5000	
B_Lympho_1	27.5000	31.7500	
B_Mono_1	2.0000	2.7500	
B_Eosino_1	1.0000	1.0000	
B_Baso_1	.0000	.0000	
B_Heamoglobin_1	14.5000	15.1250	
B_Total_Leuco_2	7570.0000	8642.5000	
B_Lympho_2	33.0000	40.0000	
B_Mono_2	2.0000	3.0000	
B_Eosino_2	1.0000	2.0000	
B_Baso_2	.0000	.0000	
B_Heamoglobin_2	14.2000	14.8000	
W12_Total_Leuco_1	7495.0000	8860.0000	
W12_Lympho_1	25.5000	30.7500	
W12_Mono_1	2.0000	3.0000	
W12_Eosino_1	1.5000	2.0000	
W12_Baso_1	.0000	.0000	
W12_Heamoglobin_1	14.2000	14.8500	
W12_Total_Leuco_2	7420.0000	8945.0000	
W12_Lympho_2	28.0000	32.5000	
W12_Mono_2	2.0000	3.0000	
W12_Eosino_2	2.0000	2.0000	
W12_Baso_2	.0000	.0000	
W12_Heamoglobin_2	14.1000	14.8750	


Wilcoxon Signed Ranks Test


Ranks	
	N	Mean Rank	Sum of Ranks	
W12_Total_Leuco_1 - B_Total_Leuco_1	Negative Ranks	21^a^	18.24	383.00	
	Positive Ranks	11^b^	13.18	145.00	
	Ties	0^c^			
	Total	32			
W12_Lympho_1 - B_Lympho_1	Negative Ranks	21^d^	15.62	328.00	
	Positive Ranks	9^e^	15.22	137.00	
	Ties	2^f^			
	Total	32			
W12_Mono_1 - B_Mono_1	Negative Ranks	7^g^	10.50	73.50	
	Positive Ranks	15^h^	11.97	179.50	
	Ties	10^i^			
	Total	32			
W12_Eosino_1 - B_Eosino_1	Negative Ranks	3^j^	8.00	24.00	
	Positive Ranks	13^k^	8.62	112.00	
	Ties	16^l^			
	Total	32			
W12_Baso_1 - B_Baso_1	Negative Ranks	0^m^	.00	.00	
	Positive Ranks	0^n^	.00	.00	
	Ties	32^o^			
	Total	32			
W12_Heamoglobin_1 - B_Heamoglobin_1	Negative Ranks	17^p^	14.21	241.50	
	Positive Ranks	12^q^	16.13	193.50	
	Ties	3^r^			
	Total	32			
W12_Total_Leuco_2 - B_Total_Leuco_2	Negative Ranks	15^s^	17.00	255.00	
	Positive Ranks	17^t^	16.06	273.00	
	Ties	0^u^			
	Total	32			
W12_Lympho_2 - B_Lympho_2	Negative Ranks	18^v^	17.33	312.00	
	Positive Ranks	12^w^	12.75	153.00	
	Ties	2^x^			
	Total	32			
W12_Mono_2 - B_Mono_2	Negative Ranks	6^y^	5.92	35.50	
	Positive Ranks	8^z^	8.69	69.50	
	Ties	18^aa^			
	Total	32			
W12_Eosino_2 - B_Eosino_2	Negative Ranks	4^ab^	9.50	38.00	
	Positive Ranks	13^ac^	8.85	115.00	
	Ties	15^ad^			
	Total	32			
W12_Baso_2 - B_Baso_2	Negative Ranks	0^ae^	.00	.00	
	Positive Ranks	0^af^	.00	.00	
	Ties	32^ag^			
	Total	32			
W12_Heamoglobin_2 - B_Heamoglobin_2	Negative Ranks	13^ah^	11.54	150.00	
	Positive Ranks	8^ai^	10.13	81.00	
	Ties	11^aj^			
	Total	32			

a. W12_Total_Leuco_1 < B_Total_Leuco_1	
b. W12_Total_Leuco_1 > B_Total_Leuco_1	
c. W12_Total_Leuco_1 = B_Total_Leuco_1	
d. W12_Lympho_1 < B_Lympho_1	
e. W12_Lympho_1 > B_Lympho_1	
f. W12_Lympho_1 = B_Lympho_1	
g. W12_Mono_1 < B_Mono_1	
h. W12_Mono_1 > B_Mono_1	
i. W12_Mono_1 = B_Mono_1	
j. W12_Eosino_1 < B_Eosino_1	
k. W12_Eosino_1 > B_Eosino_1	
l. W12_Eosino_1 = B_Eosino_1	
m. W12_Baso_1 < B_Baso_1	
n. W12_Baso_1 > B_Baso_1	
o. W12_Baso_1 = B_Baso_1	
p. W12_Heamoglobin_1 < B_Heamoglobin_1	
q. W12_Heamoglobin_1 > B_Heamoglobin_1	
r. W12_Heamoglobin_1 = B_Heamoglobin_1	
s. W12_Total_Leuco_2 < B_Total_Leuco_2	
t. W12_Total_Leuco_2 > B_Total_Leuco_2	
u. W12_Total_Leuco_2 = B_Total_Leuco_2	
v. W12_Lympho_2 < B_Lympho_2	
w. W12_Lympho_2 > B_Lympho_2	
x. W12_Lympho_2 = B_Lympho_2	
y. W12_Mono_2 < B_Mono_2	
z. W12_Mono_2 > B_Mono_2	
aa. W12_Mono_2 = B_Mono_2	
ab. W12_Eosino_2 < B_Eosino_2	
ac. W12_Eosino_2 > B_Eosino_2	
ad. W12_Eosino_2 = B_Eosino_2	
ae. W12_Baso_2 < B_Baso_2	
af. W12_Baso_2 > B_Baso_2	
ag. W12_Baso_2 = B_Baso_2	
ah. W12_Heamoglobin_2 < B_Heamoglobin_2	
ai. W12_Heamoglobin_2 > B_Heamoglobin_2	
aj. W12_Heamoglobin_2 = B_Heamoglobin_2	


Test Statistics^a^	
	W12_Total_Leuco_1 - B_Total_Leuco_1	W12_Lympho_1 - B_Lympho_1	W12_Mono_1 - B_Mono_1	W12_Eosino_1 - B_Eosino_1	
Z	-2.225^b^	-1.968^b^	-1.895^c^	-2.524^c^	
Asymp. Sig. (2-tailed)	.026	.049	.058	.012	

Test Statistics^a^	
	W12_Baso_1 - B_Baso_1	W12_Heamoglobin_1 - B_Heamoglobin_1	W12_Total_Leuco_2 - B_Total_Leuco_2	W12_Lympho_2 - B_Lympho_2	
Z	.000^d^	-.520^b^	-.168^c^	-1.637^b^	
Asymp. Sig. (2-tailed)	1.000	.603	.866	.102	

Test Statistics^a^	
	W12_Mono_2 - B_Mono_2	W12_Eosino_2 - B_Eosino_2	W12_Baso_2 - B_Baso_2	W12_Heamoglobin_2 - B_Heamoglobin_2	
Z	-1.101^c^	-1.951^c^	.000^d^	-1.203^b^	
Asymp. Sig. (2-tailed)	.271	.051	1.000	.229	

a. Wilcoxon Signed Ranks Test	
b. Based on positive ranks.	
c. Based on negative ranks.	
d. The sum of negative ranks equals the sum of positive ranks.	
