## Supplementary material for "Effects of a standardised *Amaranthus tricolor*–*Spinacia oleracea* extract on exercise performance, muscle strength, endurance and recovery in physically active adults: a randomised, double-blind, placebo-controlled trial": Raw statistical outputs: Chest circumfe. & skeletal musc.- By LMM model analysis.rtf

Mixed Model Analysis


Notes	
Output Created	18-MAY-2026 15:45:27	
Comments		
Input	Active Dataset	DataSet0	
	Filter	<none>	
	Weight	<none>	
	Split File	<none>	
	N of Rows in Working Data File	348	
Missing Value Handling	Definition of Missing	User-defined missing values are treated as valid data.	
	Cases Used	Statistics are based on all cases with valid data for all variables in the model.	
Weight Handling	not applicable	
Syntax	MIXED Chest_circum BY Treatment Time
  /CRITERIA=DFMETHOD(SATTERTHWAITE) CIN(95) MXITER(100) MXSTEP(10) SCORING(1)
    SINGULAR(0.000000000001) HCONVERGE(0.00000001, RELATIVE) LCONVERGE(0, ABSOLUTE) PCONVERGE(0,
    ABSOLUTE)
  /FIXED=Treatment Time Treatment*Time | SSTYPE(3)
  /METHOD=REML
  /PRINT=COVB DESCRIPTIVES G  SOLUTION
  /RANDOM=Treatment Time Treatment*Time | COVTYPE(VC)
  /EMMEANS=TABLES(Treatment) COMPARE ADJ(BONFERRONI)
  /EMMEANS=TABLES(Time) COMPARE ADJ(BONFERRONI)
  /EMMEANS=TABLES(Treatment*Time) .	
Resources	Processor Time	00:00:00.03	
	Elapsed Time	00:00:00.04	


Descriptive Statistics	
Chest_circum  	
Treatment	Time	Count	Mean	Standard Deviation	Coefficient of Variation	
1.00	1.00	32	91.0938	4.88816	5.4%	
	2.00	32	93.0938	3.71014	4.0%	
	3.00	32	93.4375	3.50058	3.7%	
	Total	96	92.5417	4.16733	4.5%	
2.00	1.00	32	89.0625	3.00470	3.4%	
	2.00	32	89.9062	2.64404	2.9%	
	3.00	32	90.3125	2.63277	2.9%	
	Total	96	89.7604	2.78622	3.1%	
Total	1.00	64	90.0781	4.15304	4.6%	
	2.00	64	91.5000	3.57682	3.9%	
	3.00	64	91.8750	3.45263	3.8%	
	Total	192	91.1510	3.80040	4.2%	


Model Dimension^a^	
	Number of Levels	Covariance Structure	Number of Parameters	
Fixed Effects	Intercept	1		1	
	Treatment	2		1	
	Time	3		2	
	Treatment * Time	6		2	
Random Effects	Treatment + Time + Treatment * Time	11	Variance Components	3	
Residual			1	
Total	23		10	

a. Dependent Variable: Chest_circum.	


Information Criteria^a^	
-2 Restricted Log Likelihood	1013.05067436	
Akaike's Information Criterion (AIC)	1021.05067436	
Hurvich and Tsai's Criterion (AICC)	1021.27166884	
Bozdogan's Criterion (CAIC)	1037.95366106	
Schwarz's Bayesian Criterion (BIC)	1033.95366106	

The information criteria are displayed in smaller-is-better form.^a^	
a. Dependent Variable: Chest_circum.	


Coefficients of Determination	
Pseudo-R Square Measures	Marginal	.110	
	Conditional	.492	


Intraclass Correlation Coefficients	
Overall ICCs	Adjusted	.429	
	Conditional	.382	


Fixed Effects


Type III Tests of Fixed Effects^a^	
Source	Numerator df	Denominator df	F	Sig.	
Intercept	1	5.403	2634.666	<.001	
Treatment	1	2.783	.917	.414	
Time	2	4.346	.189	.834	
Treatment * Time	2	4.037	.065	.938	

a. Dependent Variable: Chest_circum.	


Estimates of Fixed Effects^a^	
Parameter	Estimate	Std. Error	df	t	Sig.	95% Confidence Interval	
						Lower Bound	
Intercept	90.313	3.084	9.366	29.286	<.001	83.378	
[Treatment=1.00]	3.125	3.570	5.163	.875	.420	-5.965	
[Treatment=2.00]	0^b^	0	.	.	.	.	
[Time=1.00]	-1.250	3.570	6.820	-.350	.737	-9.737	
[Time=2.00]	-.406	3.570	6.820	-.114	.913	-8.893	
[Time=3.00]	0^b^	0	.	.	.	.	
[Treatment=1.00] * [Time=1.00]	-1.094	3.597	4.037	-.304	.776	-11.045	
[Treatment=1.00] * [Time=2.00]	.063	3.597	4.037	.017	.987	-9.889	
[Treatment=1.00] * [Time=3.00]	0^b^	0	.	.	.	.	
[Treatment=2.00] * [Time=1.00]	0^b^	0	.	.	.	.	
[Treatment=2.00] * [Time=2.00]	0^b^	0	.	.	.	.	
[Treatment=2.00] * [Time=3.00]	0^b^	0	.	.	.	.	

Estimates of Fixed Effects^a^	
Parameter	95% Confidence Interval	
	Upper Bound	
Intercept	97.247	
[Treatment=1.00]	12.215	
[Treatment=2.00]	.	
[Time=1.00]	7.237	
[Time=2.00]	8.081	
[Time=3.00]	.	
[Treatment=1.00] * [Time=1.00]	8.858	
[Treatment=1.00] * [Time=2.00]	10.014	
[Treatment=1.00] * [Time=3.00]	.	
[Treatment=2.00] * [Time=1.00]	.	
[Treatment=2.00] * [Time=2.00]	.	
[Treatment=2.00] * [Time=3.00]	.	

a. Dependent Variable: Chest_circum.	
b. This parameter is set to zero because it is redundant.	


Covariance Matrix for Estimates of Fixed Effects^a^	
Parameter	Intercept	[Treatment=1.00]	[Treatment=2.00]	[Time=1.00]	[Time=2.00]	
Intercept	9.510	-6.372	0^b^	-6.372	-6.372	
[Treatment=1.00]	-6.372	12.745	0^b^	3.235	3.235	
[Treatment=2.00]	0^b^	0^b^	0^b^	0^b^	0^b^	
[Time=1.00]	-6.372	3.235	0^b^	12.745	6.372	
[Time=2.00]	-6.372	3.235	0^b^	6.372	12.745	
[Time=3.00]	0^b^	0^b^	0^b^	0^b^	0^b^	
[Treatment=1.00] * [Time=1.00]	3.235	-6.470	0^b^	-6.470	-3.235	
[Treatment=1.00] * [Time=2.00]	3.235	-6.470	0^b^	-3.235	-6.470	
[Treatment=1.00] * [Time=3.00]	0^b^	0^b^	0^b^	0^b^	0^b^	
[Treatment=2.00] * [Time=1.00]	0^b^	0^b^	0^b^	0^b^	0^b^	
[Treatment=2.00] * [Time=2.00]	0^b^	0^b^	0^b^	0^b^	0^b^	
[Treatment=2.00] * [Time=3.00]	0^b^	0^b^	0^b^	0^b^	0^b^	

Covariance Matrix for Estimates of Fixed Effects^a^	
Parameter	[Time=3.00]	[Treatment=1.00] * [Time=1.00]	[Treatment=1.00] * [Time=2.00]	[Treatment=1.00] * [Time=3.00]	
Intercept	0^b^	3.235	3.235	0^b^	
[Treatment=1.00]	0^b^	-6.470	-6.470	0^b^	
[Treatment=2.00]	0^b^	0^b^	0^b^	0^b^	
[Time=1.00]	0^b^	-6.470	-3.235	0^b^	
[Time=2.00]	0^b^	-3.235	-6.470	0^b^	
[Time=3.00]	0^b^	0^b^	0^b^	0^b^	
[Treatment=1.00] * [Time=1.00]	0^b^	12.941	6.470	0^b^	
[Treatment=1.00] * [Time=2.00]	0^b^	6.470	12.941	0^b^	
[Treatment=1.00] * [Time=3.00]	0^b^	0^b^	0^b^	0^b^	
[Treatment=2.00] * [Time=1.00]	0^b^	0^b^	0^b^	0^b^	
[Treatment=2.00] * [Time=2.00]	0^b^	0^b^	0^b^	0^b^	
[Treatment=2.00] * [Time=3.00]	0^b^	0^b^	0^b^	0^b^	

Covariance Matrix for Estimates of Fixed Effects^a^	
Parameter	[Treatment=2.00] * [Time=1.00]	[Treatment=2.00] * [Time=2.00]	[Treatment=2.00] * [Time=3.00]	
Intercept	0^b^	0^b^	0^b^	
[Treatment=1.00]	0^b^	0^b^	0^b^	
[Treatment=2.00]	0^b^	0^b^	0^b^	
[Time=1.00]	0^b^	0^b^	0^b^	
[Time=2.00]	0^b^	0^b^	0^b^	
[Time=3.00]	0^b^	0^b^	0^b^	
[Treatment=1.00] * [Time=1.00]	0^b^	0^b^	0^b^	
[Treatment=1.00] * [Time=2.00]	0^b^	0^b^	0^b^	
[Treatment=1.00] * [Time=3.00]	0^b^	0^b^	0^b^	
[Treatment=2.00] * [Time=1.00]	0^b^	0^b^	0^b^	
[Treatment=2.00] * [Time=2.00]	0^b^	0^b^	0^b^	
[Treatment=2.00] * [Time=3.00]	0^b^	0^b^	0^b^	

a. Dependent Variable: Chest_circum.	
b. The covariance is set to zero because it is associated with a redundant parameter.	


Covariance Parameters


Estimates of Covariance Parameters^a^	
Parameter	Estimate	Std. Error	
Residual	12.144	1.259	
Treatment	Variance	3.137	3.608	
Time	Variance	3.137	3.390	
Treatment * Time	Variance	2.856	2.277	

a. Dependent Variable: Chest_circum.	


Covariance Matrix for Estimates of Covariance Parameters^a^	
Parameter	Residual	Treatment	Time	Treatment * Time	
		Variance	Variance	Variance	
Residual	1.586	-3.936E-14	-3.411E-14	-.050	
Treatment	Variance	-3.936E-14	13.020	-1.571	-1.238	
Time	Variance	-3.411E-14	-1.571	11.493	-2.384	
Treatment * Time	Variance	-.050	-1.238	-2.384	5.187	

a. Dependent Variable: Chest_circum.	


Random Effects Covariance Structures (G)


Treatment^a^	
	[Treatment=1.00]	[Treatment=2.00]	
[Treatment=1.00]	3.137	0	
[Treatment=2.00]	0	3.137	

Variance Components^a^	
a. Dependent Variable: Chest_circum.	


Time^a^	
	[Time=1.00]	[Time=2.00]	[Time=3.00]	
[Time=1.00]	3.137	0	0	
[Time=2.00]	0	3.137	0	
[Time=3.00]	0	0	3.137	

Variance Components^a^	
a. Dependent Variable: Chest_circum.	


Treatment * Time^a^	
	[Treatment=1.00] * [Time=1.00]	[Treatment=1.00] * [Time=2.00]	[Treatment=1.00] * [Time=3.00]	[Treatment=2.00] * [Time=1.00]	
[Treatment=1.00] * [Time=1.00]	2.856	0	0	0	
[Treatment=1.00] * [Time=2.00]	0	2.856	0	0	
[Treatment=1.00] * [Time=3.00]	0	0	2.856	0	
[Treatment=2.00] * [Time=1.00]	0	0	0	2.856	
[Treatment=2.00] * [Time=2.00]	0	0	0	0	
[Treatment=2.00] * [Time=3.00]	0	0	0	0	

Treatment * Time^a^	
	[Treatment=2.00] * [Time=2.00]	[Treatment=2.00] * [Time=3.00]	
[Treatment=1.00] * [Time=1.00]	0	0	
[Treatment=1.00] * [Time=2.00]	0	0	
[Treatment=1.00] * [Time=3.00]	0	0	
[Treatment=2.00] * [Time=1.00]	0	0	
[Treatment=2.00] * [Time=2.00]	2.856	0	
[Treatment=2.00] * [Time=3.00]	0	2.856	

Variance Components^a^	
a. Dependent Variable: Chest_circum.	


Estimated Marginal Means


1. Treatment


Estimates^a^	
Treatment	Mean	Std. Error	df	95% Confidence Interval	
				Lower Bound	Upper Bound	
1.00	92.542	2.294	4.440	86.414	98.669	
2.00	89.760	2.294	4.440	83.633	95.888	

a. Dependent Variable: Chest_circum.	


Pairwise Comparisons^a^	
(I) Treatment	(J) Treatment	Mean Difference (I-J)	Std. Error	df	Sig.^b^	95% Confidence Interval for Difference^b^	
						Lower Bound	
1.00	2.00	2.781	2.904	2.783	.414	-6.880	
2.00	1.00	-2.781	2.904	2.783	.414	-12.443	

Pairwise Comparisons^a^	
(I) Treatment	(J) Treatment	95% Confidence Interval for Difference	
		Upper Bound	
1.00	2.00	12.443	
2.00	1.00	6.880	

Based on estimated marginal means^a^	
a. Dependent Variable: Chest_circum.	
b. Adjustment for multiple comparisons: Bonferroni.	


Univariate Tests^a^	
Numerator df	Denominator df	F	Sig.	
1	2.783	.917	.414	

The F tests the effect of Treatment. This test is based on the linearly independent pairwise comparisons among the estimated marginal means.^a^	
a. Dependent Variable: Chest_circum.	


2. Time


Estimates^a^	
Time	Mean	Std. Error	df	95% Confidence Interval	
				Lower Bound	Upper Bound	
1.00	90.078	2.515	6.972	84.127	96.029	
2.00	91.500	2.515	6.972	85.549	97.451	
3.00	91.875	2.515	6.972	85.924	97.826	

a. Dependent Variable: Chest_circum.	


Pairwise Comparisons^a^	
(I) Time	(J) Time	Mean Difference (I-J)	Std. Error	df	Sig.^b^	95% Confidence Interval for Difference^b^	
						Lower Bound	
1.00	2.00	-1.422	3.084	4.346	1.000	-13.086	
	3.00	-1.797	3.084	4.346	1.000	-13.461	
2.00	1.00	1.422	3.084	4.346	1.000	-10.242	
	3.00	-.375	3.084	4.346	1.000	-12.039	
3.00	1.00	1.797	3.084	4.346	1.000	-9.867	
	2.00	.375	3.084	4.346	1.000	-11.289	

Pairwise Comparisons^a^	
(I) Time	(J) Time	95% Confidence Interval for Difference	
		Upper Bound	
1.00	2.00	10.242	
	3.00	9.867	
2.00	1.00	13.086	
	3.00	11.289	
3.00	1.00	13.461	
	2.00	12.039	

Based on estimated marginal means^a^	
a. Dependent Variable: Chest_circum.	
b. Adjustment for multiple comparisons: Bonferroni.	


Univariate Tests^a^	
Numerator df	Denominator df	F	Sig.	
2	4.346	.189	.834	

The F tests the effect of Time. This test is based on the linearly independent pairwise comparisons among the estimated marginal means.^a^	
a. Dependent Variable: Chest_circum.	


3. Treatment * Time^a^	
Treatment	Time	Mean	Std. Error	df	95% Confidence Interval	
					Lower Bound	Upper Bound	
1.00	1.00	91.094	3.084	9.366	84.159	98.028	
	2.00	93.094	3.084	9.366	86.159	100.028	
	3.00	93.438	3.084	9.366	86.503	100.372	
2.00	1.00	89.063	3.084	9.366	82.128	95.997	
	2.00	89.906	3.084	9.366	82.972	96.841	
	3.00	90.313	3.084	9.366	83.378	97.247	

a. Dependent Variable: Chest_circum.	


Mixed Model Analysis


Notes	
Output Created	18-MAY-2026 15:45:36	
Comments		
Input	Active Dataset	DataSet0	
	Filter	<none>	
	Weight	<none>	
	Split File	<none>	
	N of Rows in Working Data File	348	
Missing Value Handling	Definition of Missing	User-defined missing values are treated as valid data.	
	Cases Used	Statistics are based on all cases with valid data for all variables in the model.	
Weight Handling	not applicable	
Syntax	MIXED Skeletal_muscle BY Treatment Time
  /CRITERIA=DFMETHOD(SATTERTHWAITE) CIN(95) MXITER(100) MXSTEP(10) SCORING(1)
    SINGULAR(0.000000000001) HCONVERGE(0.00000001, RELATIVE) LCONVERGE(0, ABSOLUTE) PCONVERGE(0,
    ABSOLUTE)
  /FIXED=Treatment Time Treatment*Time | SSTYPE(3)
  /METHOD=REML
  /PRINT=COVB DESCRIPTIVES G  SOLUTION
  /RANDOM=Treatment Time Treatment*Time | COVTYPE(VC)
  /EMMEANS=TABLES(Treatment) COMPARE ADJ(BONFERRONI)
  /EMMEANS=TABLES(Time) COMPARE ADJ(BONFERRONI)
  /EMMEANS=TABLES(Treatment*Time) .	
Resources	Processor Time	00:00:00.05	
	Elapsed Time	00:00:00.03	


Descriptive Statistics	
Skeletal_muscle  	
Treatment	Time	Count	Mean	Standard Deviation	Coefficient of Variation	
1.00	1.00	32	38.4125	4.93733	12.9%	
	2.00	32	39.3469	5.20180	13.2%	
	3.00	32	39.7000	5.24232	13.2%	
	Total	96	39.1531	5.10395	13.0%	
2.00	1.00	32	37.4156	4.62107	12.4%	
	2.00	32	37.7438	4.65957	12.3%	
	3.00	32	38.0375	4.56224	12.0%	
	Total	96	37.7323	4.57277	12.1%	
Total	1.00	64	37.9141	4.77024	12.6%	
	2.00	64	38.5453	4.96495	12.9%	
	3.00	64	38.8687	4.94638	12.7%	
	Total	192	38.4427	4.88515	12.7%	


Model Dimension^a^	
	Number of Levels	Covariance Structure	Number of Parameters	
Fixed Effects	Intercept	1		1	
	Treatment	2		1	
	Time	3		2	
	Treatment * Time	6		2	
Random Effects	Treatment + Time + Treatment * Time	11	Variance Components	3	
Residual			1	
Total	23		10	

a. Dependent Variable: Skeletal_muscle.	


Information Criteria^a^	
-2 Restricted Log Likelihood	1138.19815284	
Akaike's Information Criterion (AIC)	1146.19815284	
Hurvich and Tsai's Criterion (AICC)	1146.41914731	
Bozdogan's Criterion (CAIC)	1163.10113953	
Schwarz's Bayesian Criterion (BIC)	1159.10113953	

The information criteria are displayed in smaller-is-better form.^a^	
a. Dependent Variable: Skeletal_muscle.	


Coefficients of Determination	
Pseudo-R Square Measures	Marginal	.016	
	Conditional	.438	


Intraclass Correlation Coefficients	
Overall ICCs	Adjusted	.429	
	Conditional	.422	


Fixed Effects


Type III Tests of Fixed Effects^a^	
Source	Numerator df	Denominator df	F	Sig.	
Intercept	1	5.403	239.123	<.001	
Treatment	1	2.783	.122	.751	
Time	2	4.346	.025	.975	
Treatment * Time	2	4.037	.011	.989	

a. Dependent Variable: Skeletal_muscle.	


Estimates of Fixed Effects^a^	
Parameter	Estimate	Std. Error	df	t	Sig.	95% Confidence Interval	
						Lower Bound	
Intercept	38.038	4.317	9.366	8.811	<.001	28.329	
[Treatment=1.00]	1.662	4.998	5.163	.333	.752	-11.063	
[Treatment=2.00]	0^b^	0	.	.	.	.	
[Time=1.00]	-.622	4.998	6.820	-.124	.905	-12.503	
[Time=2.00]	-.294	4.998	6.820	-.059	.955	-12.175	
[Time=3.00]	0^b^	0	.	.	.	.	
[Treatment=1.00] * [Time=1.00]	-.666	5.036	4.037	-.132	.901	-14.597	
[Treatment=1.00] * [Time=2.00]	-.059	5.036	4.037	-.012	.991	-13.991	
[Treatment=1.00] * [Time=3.00]	0^b^	0	.	.	.	.	
[Treatment=2.00] * [Time=1.00]	0^b^	0	.	.	.	.	
[Treatment=2.00] * [Time=2.00]	0^b^	0	.	.	.	.	
[Treatment=2.00] * [Time=3.00]	0^b^	0	.	.	.	.	

Estimates of Fixed Effects^a^	
Parameter	95% Confidence Interval	
	Upper Bound	
Intercept	47.746	
[Treatment=1.00]	14.388	
[Treatment=2.00]	.	
[Time=1.00]	11.260	
[Time=2.00]	11.588	
[Time=3.00]	.	
[Treatment=1.00] * [Time=1.00]	13.266	
[Treatment=1.00] * [Time=2.00]	13.872	
[Treatment=1.00] * [Time=3.00]	.	
[Treatment=2.00] * [Time=1.00]	.	
[Treatment=2.00] * [Time=2.00]	.	
[Treatment=2.00] * [Time=3.00]	.	

a. Dependent Variable: Skeletal_muscle.	
b. This parameter is set to zero because it is redundant.	


Covariance Matrix for Estimates of Fixed Effects^a^	
Parameter	Intercept	[Treatment=1.00]	[Treatment=2.00]	[Time=1.00]	[Time=2.00]	
Intercept	18.637	-12.489	0^b^	-12.489	-12.489	
[Treatment=1.00]	-12.489	24.977	0^b^	6.340	6.340	
[Treatment=2.00]	0^b^	0^b^	0^b^	0^b^	0^b^	
[Time=1.00]	-12.489	6.340	0^b^	24.977	12.489	
[Time=2.00]	-12.489	6.340	0^b^	12.489	24.977	
[Time=3.00]	0^b^	0^b^	0^b^	0^b^	0^b^	
[Treatment=1.00] * [Time=1.00]	6.340	-12.681	0^b^	-12.681	-6.340	
[Treatment=1.00] * [Time=2.00]	6.340	-12.681	0^b^	-6.340	-12.681	
[Treatment=1.00] * [Time=3.00]	0^b^	0^b^	0^b^	0^b^	0^b^	
[Treatment=2.00] * [Time=1.00]	0^b^	0^b^	0^b^	0^b^	0^b^	
[Treatment=2.00] * [Time=2.00]	0^b^	0^b^	0^b^	0^b^	0^b^	
[Treatment=2.00] * [Time=3.00]	0^b^	0^b^	0^b^	0^b^	0^b^	

Covariance Matrix for Estimates of Fixed Effects^a^	
Parameter	[Time=3.00]	[Treatment=1.00] * [Time=1.00]	[Treatment=1.00] * [Time=2.00]	[Treatment=1.00] * [Time=3.00]	
Intercept	0^b^	6.340	6.340	0^b^	
[Treatment=1.00]	0^b^	-12.681	-12.681	0^b^	
[Treatment=2.00]	0^b^	0^b^	0^b^	0^b^	
[Time=1.00]	0^b^	-12.681	-6.340	0^b^	
[Time=2.00]	0^b^	-6.340	-12.681	0^b^	
[Time=3.00]	0^b^	0^b^	0^b^	0^b^	
[Treatment=1.00] * [Time=1.00]	0^b^	25.361	12.681	0^b^	
[Treatment=1.00] * [Time=2.00]	0^b^	12.681	25.361	0^b^	
[Treatment=1.00] * [Time=3.00]	0^b^	0^b^	0^b^	0^b^	
[Treatment=2.00] * [Time=1.00]	0^b^	0^b^	0^b^	0^b^	
[Treatment=2.00] * [Time=2.00]	0^b^	0^b^	0^b^	0^b^	
[Treatment=2.00] * [Time=3.00]	0^b^	0^b^	0^b^	0^b^	

Covariance Matrix for Estimates of Fixed Effects^a^	
Parameter	[Treatment=2.00] * [Time=1.00]	[Treatment=2.00] * [Time=2.00]	[Treatment=2.00] * [Time=3.00]	
Intercept	0^b^	0^b^	0^b^	
[Treatment=1.00]	0^b^	0^b^	0^b^	
[Treatment=2.00]	0^b^	0^b^	0^b^	
[Time=1.00]	0^b^	0^b^	0^b^	
[Time=2.00]	0^b^	0^b^	0^b^	
[Time=3.00]	0^b^	0^b^	0^b^	
[Treatment=1.00] * [Time=1.00]	0^b^	0^b^	0^b^	
[Treatment=1.00] * [Time=2.00]	0^b^	0^b^	0^b^	
[Treatment=1.00] * [Time=3.00]	0^b^	0^b^	0^b^	
[Treatment=2.00] * [Time=1.00]	0^b^	0^b^	0^b^	
[Treatment=2.00] * [Time=2.00]	0^b^	0^b^	0^b^	
[Treatment=2.00] * [Time=3.00]	0^b^	0^b^	0^b^	

a. Dependent Variable: Skeletal_muscle.	
b. The covariance is set to zero because it is associated with a redundant parameter.	


Covariance Parameters


Estimates of Covariance Parameters^a^	
Parameter	Estimate	Std. Error	
Residual	23.800	2.468	
Treatment	Variance	6.148	7.071	
Time	Variance	6.148	6.644	
Treatment * Time	Variance	5.597	4.463	

a. Dependent Variable: Skeletal_muscle.	


Covariance Matrix for Estimates of Covariance Parameters^a^	
Parameter	Residual	Treatment	Time	Treatment * Time	
		Variance	Variance	Variance	
Residual	6.091	-1.078E-13	-7.034E-16	-.190	
Treatment	Variance	-1.078E-13	50.005	-6.035	-4.753	
Time	Variance	-7.034E-16	-6.035	44.142	-9.158	
Treatment * Time	Variance	-.190	-4.753	-9.158	19.921	

a. Dependent Variable: Skeletal_muscle.	


Random Effects Covariance Structures (G)


Treatment^a^	
	[Treatment=1.00]	[Treatment=2.00]	
[Treatment=1.00]	6.148	0	
[Treatment=2.00]	0	6.148	

Variance Components^a^	
a. Dependent Variable: Skeletal_muscle.	


Time^a^	
	[Time=1.00]	[Time=2.00]	[Time=3.00]	
[Time=1.00]	6.148	0	0	
[Time=2.00]	0	6.148	0	
[Time=3.00]	0	0	6.148	

Variance Components^a^	
a. Dependent Variable: Skeletal_muscle.	


Treatment * Time^a^	
	[Treatment=1.00] * [Time=1.00]	[Treatment=1.00] * [Time=2.00]	[Treatment=1.00] * [Time=3.00]	[Treatment=2.00] * [Time=1.00]	
[Treatment=1.00] * [Time=1.00]	5.597	0	0	0	
[Treatment=1.00] * [Time=2.00]	0	5.597	0	0	
[Treatment=1.00] * [Time=3.00]	0	0	5.597	0	
[Treatment=2.00] * [Time=1.00]	0	0	0	5.597	
[Treatment=2.00] * [Time=2.00]	0	0	0	0	
[Treatment=2.00] * [Time=3.00]	0	0	0	0	

Treatment * Time^a^	
	[Treatment=2.00] * [Time=2.00]	[Treatment=2.00] * [Time=3.00]	
[Treatment=1.00] * [Time=1.00]	0	0	
[Treatment=1.00] * [Time=2.00]	0	0	
[Treatment=1.00] * [Time=3.00]	0	0	
[Treatment=2.00] * [Time=1.00]	0	0	
[Treatment=2.00] * [Time=2.00]	5.597	0	
[Treatment=2.00] * [Time=3.00]	0	5.597	

Variance Components^a^	
a. Dependent Variable: Skeletal_muscle.	


Estimated Marginal Means


1. Treatment


Estimates^a^	
Treatment	Mean	Std. Error	df	95% Confidence Interval	
				Lower Bound	Upper Bound	
1.00	39.153	3.211	4.440	30.575	47.731	
2.00	37.732	3.211	4.440	29.154	46.310	

a. Dependent Variable: Skeletal_muscle.	


Pairwise Comparisons^a^	
(I) Treatment	(J) Treatment	Mean Difference (I-J)	Std. Error	df	Sig.^b^	95% Confidence Interval for Difference^b^	
						Lower Bound	
1.00	2.00	1.421	4.065	2.783	.751	-12.105	
2.00	1.00	-1.421	4.065	2.783	.751	-14.947	

Pairwise Comparisons^a^	
(I) Treatment	(J) Treatment	95% Confidence Interval for Difference	
		Upper Bound	
1.00	2.00	14.947	
2.00	1.00	12.105	

Based on estimated marginal means^a^	
a. Dependent Variable: Skeletal_muscle.	
b. Adjustment for multiple comparisons: Bonferroni.	


Univariate Tests^a^	
Numerator df	Denominator df	F	Sig.	
1	2.783	.122	.751	

The F tests the effect of Treatment. This test is based on the linearly independent pairwise comparisons among the estimated marginal means.^a^	
a. Dependent Variable: Skeletal_muscle.	


2. Time


Estimates^a^	
Time	Mean	Std. Error	df	95% Confidence Interval	
				Lower Bound	Upper Bound	
1.00	37.914	3.520	6.972	29.583	46.245	
2.00	38.545	3.520	6.972	30.214	46.876	
3.00	38.869	3.520	6.972	30.538	47.200	

a. Dependent Variable: Skeletal_muscle.	


Pairwise Comparisons^a^	
(I) Time	(J) Time	Mean Difference (I-J)	Std. Error	df	Sig.^b^	95% Confidence Interval for Difference^b^	
						Lower Bound	
1.00	2.00	-.631	4.317	4.346	1.000	-16.960	
	3.00	-.955	4.317	4.346	1.000	-17.283	
2.00	1.00	.631	4.317	4.346	1.000	-15.697	
	3.00	-.323	4.317	4.346	1.000	-16.652	
3.00	1.00	.955	4.317	4.346	1.000	-15.374	
	2.00	.323	4.317	4.346	1.000	-16.005	

Pairwise Comparisons^a^	
(I) Time	(J) Time	95% Confidence Interval for Difference	
		Upper Bound	
1.00	2.00	15.697	
	3.00	15.374	
2.00	1.00	16.960	
	3.00	16.005	
3.00	1.00	17.283	
	2.00	16.652	

Based on estimated marginal means^a^	
a. Dependent Variable: Skeletal_muscle.	
b. Adjustment for multiple comparisons: Bonferroni.	


Univariate Tests^a^	
Numerator df	Denominator df	F	Sig.	
2	4.346	.025	.975	

The F tests the effect of Time. This test is based on the linearly independent pairwise comparisons among the estimated marginal means.^a^	
a. Dependent Variable: Skeletal_muscle.	


3. Treatment * Time^a^	
Treatment	Time	Mean	Std. Error	df	95% Confidence Interval	
					Lower Bound	Upper Bound	
1.00	1.00	38.413	4.317	9.366	28.704	48.121	
	2.00	39.347	4.317	9.366	29.639	49.055	
	3.00	39.700	4.317	9.366	29.992	49.408	
2.00	1.00	37.416	4.317	9.366	27.708	47.124	
	2.00	37.744	4.317	9.366	28.036	47.452	
	3.00	38.038	4.317	9.366	28.329	47.746	

a. Dependent Variable: Skeletal_muscle.	
