## Supplementary material for "Effects of a standardised *Amaranthus tricolor*–*Spinacia oleracea* extract on exercise performance, muscle strength, endurance and recovery in physically active adults: a randomised, double-blind, placebo-controlled trial": Raw statistical outputs: CPET_Mean Exhaushion time_Non para-Mann whiteney & Wilcoxan.rtf

NPar Tests


Notes	
Output Created	18-MAY-2026 15:08:24	
Comments		
Input	Active Dataset	DataSet0	
	Filter	<none>	
	Weight	<none>	
	Split File	<none>	
	N of Rows in Working Data File	64	
Missing Value Handling	Definition of Missing	User-defined missing values are treated as missing.	
	Cases Used	Statistics for each test are based on all cases with valid data for the variable(s) used in that test.	
Syntax	NPAR TESTS
  /M-W= Exhaustion_B Exhaustion_Diff_W6 Exhaustion_Diff_W12
    BY Groups(1 2)
  /STATISTICS=DESCRIPTIVES QUARTILES
  /MISSING ANALYSIS.	
Resources	Processor Time	00:00:00.00	
	Elapsed Time	00:00:00.02	
	Number of Cases Allowed^a^	262144	

a. Based on availability of workspace memory.	


[DataSet0] 


Descriptive Statistics	
	N	Mean	Std. Deviation	Minimum	Maximum	Percentiles	
						25th	
Exhaustion_B	60	7.8573	1.18173	6.00	11.00	7.0325	
Exhaustion_Diff_W6	54	-.5815	1.06800	-3.13	2.96	-1.0600	
Exhaustion_Diff_W12	60	-1.6703	1.39217	-4.93	2.95	-2.6225	
Groups	60	1.5000	.50422	1.00	2.00	1.0000	

Descriptive Statistics	
	Percentiles	
	50th (Median)	75th	
Exhaustion_B	8.0000	8.6150	
Exhaustion_Diff_W6	-.5300	-.0500	
Exhaustion_Diff_W12	-1.2150	-.9625	
Groups	1.5000	2.0000	


Mann-Whitney Test


Ranks	
	Groups	N	Mean Rank	Sum of Ranks	
Exhaustion_B	1.00	30	32.88	986.50	
	2.00	30	28.12	843.50	
	Total	60			
Exhaustion_Diff_W6	1.00	28	21.59	604.50	
	2.00	26	33.87	880.50	
	Total	54			
Exhaustion_Diff_W12	1.00	30	20.47	614.00	
	2.00	30	40.53	1216.00	
	Total	60			


	Exhaustion_B	Exhaustion_Diff_W6	Exhaustion_Diff_W12	
Mann-Whitney U	378.500	198.500	149.000	
Wilcoxon W	843.500	604.500	614.000	
Z	-1.058	-2.867	-4.451	
Asymp. Sig. (2-tailed)	.290	.004	<.001	


a. Grouping Variable: Groups	


NPar Tests


Notes	
Output Created	19-MAY-2026 11:16:23	
Comments		
Input	Active Dataset	DataSet0	
	Filter	<none>	
	Weight	<none>	
	Split File	<none>	
	N of Rows in Working Data File	35	
Missing Value Handling	Definition of Missing	User-defined missing values are treated as missing.	
	Cases Used	Statistics for each test are based on all cases with valid data for the variable(s) used in that test.	
Syntax	NPAR TESTS
  /WILCOXON= Exhau_B_1 Exhau_Bas_1 Exha_B_2
    Exha_Bas_2 Exhau_W6_1 Exhau_W12_1
    Exha_W6_2 Exha_W12_2 (PAIRED)
  /STATISTICS DESCRIPTIVES QUARTILES
  /MISSING ANALYSIS.	
Resources	Processor Time	00:00:00.03	
	Elapsed Time	00:00:00.03	
	Number of Cases Allowed^a^	149796	

a. Based on availability of workspace memory.	


[DataSet0] 


Descriptive Statistics	
	N	Mean	Std. Deviation	Minimum	Maximum	Percentiles	
						25th	
Exhau_B_1	28	8.0679	1.19110	6.00	10.28	7.0350	
Exhau_Bas_1	30	8.0650	1.26095	6.00	10.28	7.0275	
Exha_B_2	26	7.7608	1.09653	6.02	11.00	7.0350	
Exha_Bas_2	30	7.6497	1.07791	6.00	11.00	7.0350	
xhau_W6_1	28	8.8511	1.53461	5.50	12.00	8.0125	
Exhau_W12_1	30	10.4500	1.51632	7.00	13.00	9.7950	
Exha_W6_2	26	8.1250	1.15800	6.05	11.20	7.1125	
Exha_W12_2	30	8.6053	1.17739	6.12	11.80	8.0350	

Descriptive Statistics	
	Percentiles	
	50th (Median)	75th	
Exhau_B_1	8.0000	9.0000	
Exhau_Bas_1	8.0000	9.0425	
Exha_B_2	8.0000	8.1850	
Exha_Bas_2	7.4600	8.1275	
xhau_W6_1	9.0200	10.0525	
Exhau_W12_1	10.7000	11.1200	
Exha_W6_2	8.0500	8.6575	
Exha_W12_2	8.3250	9.0525	


Wilcoxon Signed Ranks Test


Ranks	
	N	Mean Rank	Sum of Ranks	
xhau_W6_1 - Exhau_B_1	Negative Ranks	5^m^	16.00	80.00	
	Positive Ranks	23^n^	14.17	326.00	
	Ties	0^o^			
	Total	28			
Exhau_W12_1 - Exhau_Bas_1	Negative Ranks	1^p^	16.00	16.00	
	Positive Ranks	29^q^	15.48	449.00	
	Ties	0^r^			
	Total	30			
Exha_W6_2 - Exha_B_2	Negative Ranks	0^s^	.00	.00	
	Positive Ranks	26^t^	13.50	351.00	
	Ties	0^u^			
	Total	26			
Exha_W12_2 - Exha_Bas_2	Negative Ranks	0^v^	.00	.00	
	Positive Ranks	30^w^	15.50	465.00	
	Ties	0^x^			
	Total	30			

a. xhau_W6_1 < Exhau_B_1	
b. xhau_W6_1 > Exhau_B_1	
c. xhau_W6_1 = Exhau_B_1	
d. Exhau_W12_1 < Exhau_Bas_1	
e. Exhau_W12_1 > Exhau_Bas_1	
f. Exhau_W12_1 = Exhau_Bas_1	
g. Exha_W6_2 < Exha_B_2	
h. Exha_W6_2 > Exha_B_2	
i. Exha_W6_2 = Exha_B_2	
j. Exha_W12_2 < Exha_Bas_2	
k. Exha_W12_2 > Exha_Bas_2	
l. Exha_W12_2 = Exha_Bas_2	


Test Statistics^a^	
	xhau_W6_1 - Exhau_B_1	Exhau_W12_1 - Exhau_Bas_1	Exha_W6_2 - Exha_B_2	Exha_W12_2 - Exha_Bas_2	
Z	-2.802^c^	-4.453^c^	-4.461^c^	-4.783^c^	
Asymp. Sig. (2-tailed)	.005	<.001	<.001	<.001	

a. Wilcoxon Signed Ranks Test	
b. Based on positive ranks.	
c. Based on negative ranks.	
