## Supplementary material for "Effects of a standardised *Amaranthus tricolor*–*Spinacia oleracea* extract on exercise performance, muscle strength, endurance and recovery in physically active adults: a randomised, double-blind, placebo-controlled trial": Raw statistical outputs: DOMS_Non para Test.rtf

NPar Tests


Notes	
Output Created	15-MAY-2026 15:02:15	
Comments		
Input	Active Dataset	DataSet0	
	Filter	<none>	
	Weight	<none>	
	Split File	<none>	
	N of Rows in Working Data File	64	
Missing Value Handling	Definition of Missing	User-defined missing values are treated as missing.	
	Cases Used	Statistics for each test are based on all cases with valid data for the variable(s) used in that test.	
Syntax	NPAR TESTS
  /M-W= Upper_Soreness_B Upper_Soreness_Diff_W6 Upper_Soreness_Diff_W12 Upper_Pain_B
    Upper_Pain_Diff_W6 Upper_Pain_Diff_W12 Lower_Soreness_B Lower_Soreness_Diff_W6
    Lower_Soreness_Diff_W12 Lower_Pain_B Lower_Pain_Diff_W6 Lower_Pain_Diff_W12 BY Groups(1 2)
  /STATISTICS=DESCRIPTIVES QUARTILES
  /MISSING ANALYSIS.	
Resources	Processor Time	00:00:00.02	
	Elapsed Time	00:00:00.02	
	Number of Cases Allowed^a^	174762	

a. Based on availability of workspace memory.	


[DataSet0] 


Descriptive Statistics	
	N	Mean	Std. Deviation	Minimum	Maximum	Percentiles	
						25th	
Upper_Soreness_B	64	5.6875	.79433	4.00	7.00	5.0000	
Upper_Soreness_Diff_W6	64	.8437	.82074	-1.00	3.00	.0000	
Upper_Soreness_Diff_W12	64	1.8750	1.17514	-1.00	5.00	1.0000	
Upper_Pain_B	64	2.6563	.59678	2.00	4.00	2.0000	
Upper_Pain_Diff_W6	64	.5625	.61399	-1.00	2.00	.0000	
Upper_Pain_Diff_W12	64	1.0156	.70130	.00	3.00	1.0000	
Lower_Soreness_B	64	5.7188	.78617	4.00	7.00	5.0000	
Lower_Soreness_Diff_W6	64	.8437	.91233	-1.00	3.00	.0000	
Lower_Soreness_Diff_W12	64	1.5937	1.25633	-1.00	5.00	1.0000	
Lower_Pain_B	64	2.7344	.64837	2.00	4.00	2.0000	
Lower_Pain_Diff_W6	64	.5781	.52869	.00	2.00	.0000	
Lower_Pain_Diff_W12	64	1.0781	.80287	.00	3.00	.2500	
Groups	64	1.5000	.50395	1.00	2.00	1.0000	

Descriptive Statistics	
	Percentiles	
	50th (Median)	75th	
Upper_Soreness_B	6.0000	6.0000	
Upper_Soreness_Diff_W6	1.0000	1.0000	
Upper_Soreness_Diff_W12	2.0000	3.0000	
Upper_Pain_B	3.0000	3.0000	
Upper_Pain_Diff_W6	1.0000	1.0000	
Upper_Pain_Diff_W12	1.0000	1.0000	
Lower_Soreness_B	6.0000	6.0000	
Lower_Soreness_Diff_W6	1.0000	1.0000	
Lower_Soreness_Diff_W12	1.0000	3.0000	
Lower_Pain_B	3.0000	3.0000	
Lower_Pain_Diff_W6	1.0000	1.0000	
Lower_Pain_Diff_W12	1.0000	2.0000	
Groups	1.5000	2.0000	


Mann-Whitney Test


Ranks	
	Groups	N	Mean Rank	Sum of Ranks	
Upper_Soreness_B	1.00	32	33.27	1064.50	
	2.00	32	31.73	1015.50	
	Total	64			
Upper_Soreness_Diff_W6	1.00	32	36.58	1170.50	
	2.00	32	28.42	909.50	
	Total	64			
Upper_Soreness_Diff_W12	1.00	32	41.73	1335.50	
	2.00	32	23.27	744.50	
	Total	64			
Upper_Pain_B	1.00	32	33.44	1070.00	
	2.00	32	31.56	1010.00	
	Total	64			
Upper_Pain_Diff_W6	1.00	32	36.75	1176.00	
	2.00	32	28.25	904.00	
	Total	64			
Upper_Pain_Diff_W12	1.00	32	38.78	1241.00	
	2.00	32	26.22	839.00	
	Total	64			
Lower_Soreness_B	1.00	32	34.34	1099.00	
	2.00	32	30.66	981.00	
	Total	64			
Lower_Soreness_Diff_W6	1.00	32	37.97	1215.00	
	2.00	32	27.03	865.00	
	Total	64			
Lower_Soreness_Diff_W12	1.00	32	44.34	1419.00	
	2.00	32	20.66	661.00	
	Total	64			
Lower_Pain_B	1.00	32	36.11	1155.50	
	2.00	32	28.89	924.50	
	Total	64			
Lower_Pain_Diff_W6	1.00	32	37.70	1206.50	
	2.00	32	27.30	873.50	
	Total	64			
Lower_Pain_Diff_W12	1.00	32	42.41	1357.00	
	2.00	32	22.59	723.00	
	Total	64			


Test Statistics^a^	
	Upper_Soreness_B	Upper_Soreness_Diff_W6	Upper_Soreness_Diff_W12	Upper_Pain_B	
Mann-Whitney U	487.500	381.500	216.500	482.000	
Wilcoxon W	1015.500	909.500	744.500	1010.000	
Z	-.353	-1.905	-4.120	-.455	
Asymp. Sig. (2-tailed)	.724	.057	<.001	.649	

Test Statistics^a^	
	Upper_Pain_Diff_W6	Upper_Pain_Diff_W12	Lower_Soreness_B	Lower_Soreness_Diff_W6	
Mann-Whitney U	376.000	311.000	453.000	337.000	
Wilcoxon W	904.000	839.000	981.000	865.000	
Z	-2.094	-3.011	-.850	-2.574	
Asymp. Sig. (2-tailed)	.036	.003	.395	.010	

Test Statistics^a^	
	Lower_Soreness_Diff_W12	Lower_Pain_B	Lower_Pain_Diff_W6	Lower_Pain_Diff_W12	
Mann-Whitney U	133.000	396.500	345.500	195.000	
Wilcoxon W	661.000	924.500	873.500	723.000	
Z	-5.249	-1.724	-2.577	-4.556	
Asymp. Sig. (2-tailed)	<.001	.085	.010	<.001	

a. Grouping Variable: Groups	


NPar Tests


Notes	
Output Created	15-MAY-2026 15:04:18	
Comments		
Input	Active Dataset	DataSet0	
	Filter	<none>	
	Weight	<none>	
	Split File	<none>	
	N of Rows in Working Data File	64	
Missing Value Handling	Definition of Missing	User-defined missing values are treated as missing.	
	Cases Used	Statistics for each test are based on all cases with valid data for the variable(s) used in that test.	
Syntax	NPAR TESTS
  /WILCOXON=Upper_Soreness_B_1 Upper_Soreness_B_1 Upper_Pain_B_1 Upper_Pain_B_1 Lower_Soreness_B_1
    Lower_Soreness_B_1 Lower_Pain_B_1 Lower_Pain_B_1 Upper_Soreness_B_2 Upper_Soreness_B_2
    Upper_Pain_B_2 Upper_Pain_B_2 Lower_Soreness_B_2 Lower_Soreness_B_2 Lower_Pain_B_2 Lower_Pain_B_2
    WITH Upper_Soreness_W6_1 Upper_Soreness_W12_1 Upper_Pain_W6_1 Upper_Pain_W12_1 Lower_Soreness_W6_1
    Lower_Soreness_W12_1 Lower_Pain_W6_1 Lower_Pain_W12_1 Upper_Soreness_W6_2 Upper_Soreness_W12_2
    Upper_Pain_W6_2 Upper_Pain_W12_2 Lower_Soreness_W6_2 Lower_Soreness_W12_2 Lower_Pain_W6_2
    Lower_Pain_W12_2 (PAIRED)
  /STATISTICS DESCRIPTIVES QUARTILES
  /MISSING ANALYSIS.	
Resources	Processor Time	00:00:00.02	
	Elapsed Time	00:00:00.00	
	Number of Cases Allowed^a^	108473	

a. Based on availability of workspace memory.	


Descriptive Statistics	
	N	Mean	Std. Deviation	Minimum	Maximum	Percentiles	
						25th	
Upper_Soreness_B_1	32	5.7188	.85135	4.00	7.00	5.0000	
Upper_Pain_B_1	32	2.6875	.59229	2.00	4.00	2.0000	
Lower_Soreness_B_1	32	5.8125	.85901	4.00	7.00	5.0000	
Lower_Pain_B_1	32	2.8750	.65991	2.00	4.00	2.0000	
Upper_Soreness_B_2	32	5.6563	.74528	4.00	7.00	5.0000	
Upper_Pain_B_2	32	2.6250	.60907	2.00	4.00	2.0000	
Lower_Soreness_B_2	32	5.6250	.70711	4.00	7.00	5.0000	
Lower_Pain_B_2	32	2.5937	.61484	2.00	4.00	2.0000	
Upper_Soreness_W6_1	32	4.6875	.69270	4.00	6.00	4.0000	
Upper_Soreness_W12_1	32	3.2500	.87988	2.00	5.00	3.0000	
Upper_Pain_W6_1	32	1.9687	.40035	1.00	3.00	2.0000	
Upper_Pain_W12_1	32	1.4063	.49899	1.00	2.00	1.0000	
Lower_Soreness_W6_1	32	4.6875	.82060	4.00	7.00	4.0000	
Lower_Soreness_W12_1	32	3.4375	.87759	2.00	5.00	3.0000	
Lower_Pain_W6_1	32	2.1250	.49187	1.00	3.00	2.0000	
Lower_Pain_W12_1	32	1.3438	.48256	1.00	2.00	1.0000	
Upper_Soreness_W6_2	32	5.0000	.71842	4.00	6.00	4.2500	
Upper_Soreness_W12_2	32	4.3750	.60907	3.00	5.00	4.0000	
Upper_Pain_W6_2	32	2.2188	.60824	1.00	4.00	2.0000	
Upper_Pain_W12_2	32	1.8750	.60907	1.00	3.00	1.2500	
Lower_Soreness_W6_2	32	5.0625	.87759	4.00	7.00	4.2500	
Lower_Soreness_W12_2	32	4.8125	.73780	4.00	6.00	4.0000	
Lower_Pain_W6_2	32	2.1875	.59229	1.00	3.00	2.0000	
Lower_Pain_W12_2	32	1.9688	.53788	1.00	3.00	2.0000	

Descriptive Statistics	
	Percentiles	
	50th (Median)	75th	
Upper_Soreness_B_1	6.0000	6.0000	
Upper_Pain_B_1	3.0000	3.0000	
Lower_Soreness_B_1	6.0000	6.7500	
Lower_Pain_B_1	3.0000	3.0000	
Upper_Soreness_B_2	6.0000	6.0000	
Upper_Pain_B_2	3.0000	3.0000	
Lower_Soreness_B_2	6.0000	6.0000	
Lower_Pain_B_2	3.0000	3.0000	
Upper_Soreness_W6_1	5.0000	5.0000	
Upper_Soreness_W12_1	3.0000	4.0000	
Upper_Pain_W6_1	2.0000	2.0000	
Upper_Pain_W12_1	1.0000	2.0000	
Lower_Soreness_W6_1	4.5000	5.0000	
Lower_Soreness_W12_1	3.0000	4.0000	
Lower_Pain_W6_1	2.0000	2.0000	
Lower_Pain_W12_1	1.0000	2.0000	
Upper_Soreness_W6_2	5.0000	5.7500	
Upper_Soreness_W12_2	4.0000	5.0000	
Upper_Pain_W6_2	2.0000	2.7500	
Upper_Pain_W12_2	2.0000	2.0000	
Lower_Soreness_W6_2	5.0000	5.0000	
Lower_Soreness_W12_2	5.0000	5.0000	
Lower_Pain_W6_2	2.0000	3.0000	
Lower_Pain_W12_2	2.0000	2.0000	


Wilcoxon Signed Ranks Test


Ranks	
	N	Mean Rank	Sum of Ranks	
Upper_Soreness_W6_1 - Upper_Soreness_B_1	Negative Ranks	25^a^	13.00	325.00	
	Positive Ranks	0^b^	.00	.00	
	Ties	7^c^			
	Total	32			
Upper_Soreness_W12_1 - Upper_Soreness_B_1	Negative Ranks	32^d^	16.50	528.00	
	Positive Ranks	0^e^	.00	.00	
	Ties	0^f^			
	Total	32			
Upper_Pain_W6_1 - Upper_Pain_B_1	Negative Ranks	23^g^	12.52	288.00	
	Positive Ranks	1^h^	12.00	12.00	
	Ties	8^i^			
	Total	32			
Upper_Pain_W12_1 - Upper_Pain_B_1	Negative Ranks	28^j^	14.50	406.00	
	Positive Ranks	0^k^	.00	.00	
	Ties	4^l^			
	Total	32			
Lower_Soreness_W6_1 - Lower_Soreness_B_1	Negative Ranks	26^m^	14.15	368.00	
	Positive Ranks	1^n^	10.00	10.00	
	Ties	5^o^			
	Total	32			
Lower_Soreness_W12_1 - Lower_Soreness_B_1	Negative Ranks	31^p^	16.92	524.50	
	Positive Ranks	1^q^	3.50	3.50	
	Ties	0^r^			
	Total	32			
Lower_Pain_W6_1 - Lower_Pain_B_1	Negative Ranks	23^s^	12.00	276.00	
	Positive Ranks	0^t^	.00	.00	
	Ties	9^u^			
	Total	32			
Lower_Pain_W12_1 - Lower_Pain_B_1	Negative Ranks	30^v^	15.50	465.00	
	Positive Ranks	0^w^	.00	.00	
	Ties	2^x^			
	Total	32			
Upper_Soreness_W6_2 - Upper_Soreness_B_2	Negative Ranks	18^y^	10.72	193.00	
	Positive Ranks	2^z^	8.50	17.00	
	Ties	12^aa^			
	Total	32			
Upper_Soreness_W12_2 - Upper_Soreness_B_2	Negative Ranks	26^ab^	14.25	370.50	
	Positive Ranks	1^ac^	7.50	7.50	
	Ties	5^ad^			
	Total	32			
Upper_Pain_W6_2 - Upper_Pain_B_2	Negative Ranks	15^ae^	9.00	135.00	
	Positive Ranks	2^af^	9.00	18.00	
	Ties	15^ag^			
	Total	32			
Upper_Pain_W12_2 - Upper_Pain_B_2	Negative Ranks	22^ah^	11.50	253.00	
	Positive Ranks	0^ai^	.00	.00	
	Ties	10^aj^			
	Total	32			
Lower_Soreness_W6_2 - Lower_Soreness_B_2	Negative Ranks	18^ak^	11.17	201.00	
	Positive Ranks	3^al^	10.00	30.00	
	Ties	11^am^			
	Total	32			
Lower_Soreness_W12_2 - Lower_Soreness_B_2	Negative Ranks	21^an^	11.62	244.00	
	Positive Ranks	1^ao^	9.00	9.00	
	Ties	10^ap^			
	Total	32			
Lower_Pain_W6_2 - Lower_Pain_B_2	Negative Ranks	13^aq^	7.00	91.00	
	Positive Ranks	0^ar^	.00	.00	
	Ties	19^as^			
	Total	32			
Lower_Pain_W12_2 - Lower_Pain_B_2	Negative Ranks	18^at^	9.50	171.00	
	Positive Ranks	0^au^	.00	.00	
	Ties	14^av^			
	Total	32			

a. Upper_Soreness_W6_1 < Upper_Soreness_B_1	
b. Upper_Soreness_W6_1 > Upper_Soreness_B_1	
c. Upper_Soreness_W6_1 = Upper_Soreness_B_1	
d. Upper_Soreness_W12_1 < Upper_Soreness_B_1	
e. Upper_Soreness_W12_1 > Upper_Soreness_B_1	
f. Upper_Soreness_W12_1 = Upper_Soreness_B_1	
g. Upper_Pain_W6_1 < Upper_Pain_B_1	
h. Upper_Pain_W6_1 > Upper_Pain_B_1	
i. Upper_Pain_W6_1 = Upper_Pain_B_1	
j. Upper_Pain_W12_1 < Upper_Pain_B_1	
k. Upper_Pain_W12_1 > Upper_Pain_B_1	
l. Upper_Pain_W12_1 = Upper_Pain_B_1	
m. Lower_Soreness_W6_1 < Lower_Soreness_B_1	
n. Lower_Soreness_W6_1 > Lower_Soreness_B_1	
o. Lower_Soreness_W6_1 = Lower_Soreness_B_1	
p. Lower_Soreness_W12_1 < Lower_Soreness_B_1	
q. Lower_Soreness_W12_1 > Lower_Soreness_B_1	
r. Lower_Soreness_W12_1 = Lower_Soreness_B_1	
s. Lower_Pain_W6_1 < Lower_Pain_B_1	
t. Lower_Pain_W6_1 > Lower_Pain_B_1	
u. Lower_Pain_W6_1 = Lower_Pain_B_1	
v. Lower_Pain_W12_1 < Lower_Pain_B_1	
w. Lower_Pain_W12_1 > Lower_Pain_B_1	
x. Lower_Pain_W12_1 = Lower_Pain_B_1	
y. Upper_Soreness_W6_2 < Upper_Soreness_B_2	
z. Upper_Soreness_W6_2 > Upper_Soreness_B_2	
aa. Upper_Soreness_W6_2 = Upper_Soreness_B_2	
ab. Upper_Soreness_W12_2 < Upper_Soreness_B_2	
ac. Upper_Soreness_W12_2 > Upper_Soreness_B_2	
ad. Upper_Soreness_W12_2 = Upper_Soreness_B_2	
ae. Upper_Pain_W6_2 < Upper_Pain_B_2	
af. Upper_Pain_W6_2 > Upper_Pain_B_2	
ag. Upper_Pain_W6_2 = Upper_Pain_B_2	
ah. Upper_Pain_W12_2 < Upper_Pain_B_2	
ai. Upper_Pain_W12_2 > Upper_Pain_B_2	
aj. Upper_Pain_W12_2 = Upper_Pain_B_2	
ak. Lower_Soreness_W6_2 < Lower_Soreness_B_2	
al. Lower_Soreness_W6_2 > Lower_Soreness_B_2	
am. Lower_Soreness_W6_2 = Lower_Soreness_B_2	
an. Lower_Soreness_W12_2 < Lower_Soreness_B_2	
ao. Lower_Soreness_W12_2 > Lower_Soreness_B_2	
ap. Lower_Soreness_W12_2 = Lower_Soreness_B_2	
aq. Lower_Pain_W6_2 < Lower_Pain_B_2	
ar. Lower_Pain_W6_2 > Lower_Pain_B_2	
as. Lower_Pain_W6_2 = Lower_Pain_B_2	
at. Lower_Pain_W12_2 < Lower_Pain_B_2	
au. Lower_Pain_W12_2 > Lower_Pain_B_2	
av. Lower_Pain_W12_2 = Lower_Pain_B_2	


Test Statistics^a^	
	Upper_Soreness_W6_1 - Upper_Soreness_B_1	Upper_Soreness_W12_1 - Upper_Soreness_B_1	Upper_Pain_W6_1 - Upper_Pain_B_1	Upper_Pain_W12_1 - Upper_Pain_B_1	
Z	-4.586^b^	-4.984^b^	-4.426^b^	-4.764^b^	
Asymp. Sig. (2-tailed)	<.001	<.001	<.001	<.001	

Test Statistics^a^	
	Lower_Soreness_W6_1 - Lower_Soreness_B_1	Lower_Soreness_W12_1 - Lower_Soreness_B_1	Lower_Pain_W6_1 - Lower_Pain_B_1	Lower_Pain_W12_1 - Lower_Pain_B_1	
Z	-4.493^b^	-4.932^b^	-4.707^b^	-4.904^b^	
Asymp. Sig. (2-tailed)	<.001	<.001	<.001	<.001	

Test Statistics^a^	
	Upper_Soreness_W6_2 - Upper_Soreness_B_2	Upper_Soreness_W12_2 - Upper_Soreness_B_2	Upper_Pain_W6_2 - Upper_Pain_B_2	Upper_Pain_W12_2 - Upper_Pain_B_2	
Z	-3.500^b^	-4.471^b^	-3.153^b^	-4.523^b^	
Asymp. Sig. (2-tailed)	<.001	<.001	.002	<.001	

Test Statistics^a^	
	Lower_Soreness_W6_2 - Lower_Soreness_B_2	Lower_Soreness_W12_2 - Lower_Soreness_B_2	Lower_Pain_W6_2 - Lower_Pain_B_2	Lower_Pain_W12_2 - Lower_Pain_B_2	
Z	-3.266^b^	-4.041^b^	-3.606^b^	-4.066^b^	
Asymp. Sig. (2-tailed)	.001	<.001	<.001	<.001	

a. Wilcoxon Signed Ranks Test	
b. Based on positive ranks.	
