## Supplementary material for "Effects of a standardised *Amaranthus tricolor*–*Spinacia oleracea* extract on exercise performance, muscle strength, endurance and recovery in physically active adults: a randomised, double-blind, placebo-controlled trial": Raw statistical outputs: Kidney function test_Serum creatinine normal ANCOVA.rtf

Univariate Analysis of Variance


Notes	
Output Created	11-MAY-2026 12:43:49	
Comments		
Input	Active Dataset	DataSet0	
	Filter	<none>	
	Weight	<none>	
	Split File	<none>	
	N of Rows in Working Data File	64	
Missing Value Handling	Definition of Missing	User-defined missing values are treated as missing.	
	Cases Used	Statistics are based on all cases with valid data for all variables in the model.	
Syntax	UNIANOVA W12_Ser_creatinine BY Groups WITH B_Ser_creatinine
  /METHOD=SSTYPE(3)
  /INTERCEPT=INCLUDE
  /EMMEANS=TABLES(Groups) WITH(B_Ser_creatinine=MEAN) COMPARE ADJ(BONFERRONI)
  /PRINT ETASQ DESCRIPTIVE
  /CRITERIA=ALPHA(.05)
  /DESIGN=B_Ser_creatinine Groups.	
Resources	Processor Time	00:00:00.00	
	Elapsed Time	00:00:00.00	


Between-Subjects Factors	
	N	
Groups	1	32	
	2	32	


Descriptive Statistics	
Dependent Variable:   W12_Ser_creatinine  	
Groups	Mean	Std. Deviation	N	
1	.8125	.13714	32	
2	.8103	.13823	32	
Total	.8114	.13659	64	


Tests of Between-Subjects Effects	
Dependent Variable:   W12_Ser_creatinine  	
Source	Type III Sum of Squares	df	Mean Square	F	Sig.	Partial Eta Squared	
Corrected Model	.349^a^	2	.175	12.881	<.001	.297	
Intercept	.229	1	.229	16.883	<.001	.217	
B_Ser_creatinine	.349	1	.349	25.757	<.001	.297	
Groups	.003	1	.003	.254	.616	.004	
Error	.826	61	.014				
Total	43.312	64					
Corrected Total	1.175	63					

a. R Squared = .297 (Adjusted R Squared = .274)	


Estimated Marginal Means


Groups


Estimates	
Dependent Variable:   W12_Ser_creatinine  	
Groups	Mean	Std. Error	95% Confidence Interval	
			Lower Bound	Upper Bound	
1	.819^a^	.021	.778	.860	
2	.804^a^	.021	.763	.845	

a. Covariates appearing in the model are evaluated at the following values: B_Ser_creatinine = .8342.	


Pairwise Comparisons	
Dependent Variable:   W12_Ser_creatinine  	
(I) Groups	(J) Groups	Mean Difference (I-J)	Std. Error	Sig.^a^	95% Confidence Interval for Difference^a^	
					Lower Bound	Upper Bound	
1	2	.015	.029	.616	-.044	.073	
2	1	-.015	.029	.616	-.073	.044	

Based on estimated marginal means	
a. Adjustment for multiple comparisons: Bonferroni.	


Univariate Tests	
Dependent Variable:   W12_Ser_creatinine  	
	Sum of Squares	df	Mean Square	F	Sig.	Partial Eta Squared	
Contrast	.003	1	.003	.254	.616	.004	
Error	.826	61	.014				

The F tests the effect of Groups. This test is based on the linearly independent pairwise comparisons among the estimated marginal means.	


T-Test


Notes	
Output Created	11-MAY-2026 12:44:15	
Comments		
Input	Active Dataset	DataSet0	
	Filter	<none>	
	Weight	<none>	
	Split File	<none>	
	N of Rows in Working Data File	64	
Missing Value Handling	Definition of Missing	User defined missing values are treated as missing.	
	Cases Used	Statistics for each analysis are based on the cases with no missing or out-of-range data for any variable in the analysis.	
Syntax	T-TEST PAIRS=B_Ser_creatinine_1 B_Ser_creatinine_2 WITH W12_Ser_creatinine_1 W12_Ser_creatinine_2
    (PAIRED)
  /ES DISPLAY(TRUE) STANDARDIZER(SD)
  /CRITERIA=CI(.9500)
  /MISSING=ANALYSIS.	
Resources	Processor Time	00:00:00.00	
	Elapsed Time	00:00:00.00	


Paired Samples Statistics	
	Mean	N	Std. Deviation	Std. Error Mean	
Pair 1	B_Ser_creatinine_1	.8225	32	.11019	.01948	
	W12_Ser_creatinine_1	.8125	32	.13714	.02424	
Pair 2	B_Ser_creatinine_2	.8459	32	.16521	.02921	
	W12_Ser_creatinine_2	.8103	32	.13823	.02444	


Paired Samples Correlations	
	N	Correlation	Significance	
			One-Sided p	Two-Sided p	
Pair 1	B_Ser_creatinine_1 & W12_Ser_creatinine_1	32	.558	<.001	<.001	
Pair 2	B_Ser_creatinine_2 & W12_Ser_creatinine_2	32	.553	<.001	.001	


Paired Samples Test	
	Paired Differences	
	Mean	Std. Deviation	Std. Error Mean	95% Confidence Interval of the Difference	
				Lower	
Pair 1	B_Ser_creatinine_1 - W12_Ser_creatinine_1	.01000	.11865	.02097	-.03278	
Pair 2	B_Ser_creatinine_2 - W12_Ser_creatinine_2	.03563	.14538	.02570	-.01679	

Paired Samples Test	
	Paired Differences	t	df	Significance	
	95% Confidence Interval of the Difference			One-Sided p	
	Upper				
Pair 1	B_Ser_creatinine_1 - W12_Ser_creatinine_1	.05278	.477	31	.318	
Pair 2	B_Ser_creatinine_2 - W12_Ser_creatinine_2	.08804	1.386	31	.088	


Paired Samples Test	
	Significance	
	Two-Sided p	
		
Pair 1	B_Ser_creatinine_1 - W12_Ser_creatinine_1	.637	
Pair 2	B_Ser_creatinine_2 - W12_Ser_creatinine_2	.176	


Paired Samples Effect Sizes	
	Standardizer^a^	Point Estimate	95% Confidence Interval	
			Lower	
Pair 1	B_Ser_creatinine_1 - W12_Ser_creatinine_1	Cohen's d	.11865	.084	-.263	
		Hedges' correction	.12162	.082	-.257	
Pair 2	B_Ser_creatinine_2 - W12_Ser_creatinine_2	Cohen's d	.14538	.245	-.109	
		Hedges' correction	.14902	.239	-.106	

Paired Samples Effect Sizes	
	95% Confidence Interval^a^	
	Upper	
Pair 1	B_Ser_creatinine_1 - W12_Ser_creatinine_1	Cohen's d	.431	
		Hedges' correction	.420	
Pair 2	B_Ser_creatinine_2 - W12_Ser_creatinine_2	Cohen's d	.595	
		Hedges' correction	.580	

a. The denominator used in estimating the effect sizes. 
Cohen's d uses the sample standard deviation of the mean difference. 
Hedges' correction uses the sample standard deviation of the mean difference, plus a correction factor.	
