## Supplementary material for "Effects of a standardised *Amaranthus tricolor*–*Spinacia oleracea* extract on exercise performance, muscle strength, endurance and recovery in physically active adults: a randomised, double-blind, placebo-controlled trial": Raw statistical outputs: Kidney function_By Non parametric Test.rtf

NPar Tests


Notes	
Output Created	11-MAY-2026 15:05:28	
Comments		
Input	Active Dataset	DataSet0	
	Filter	<none>	
	Weight	<none>	
	Split File	<none>	
	N of Rows in Working Data File	64	
Missing Value Handling	Definition of Missing	User-defined missing values are treated as missing.	
	Cases Used	Statistics for each test are based on all cases with valid data for the variable(s) used in that test.	
Syntax	NPAR TESTS
  /M-W= B_ser_Uric_Acid Diff_ser_Uric_Acid B_Ser_Blood_Urea Diff_Ser_Blood_Urea BY Groups(1 2)
  /STATISTICS=DESCRIPTIVES QUARTILES
  /MISSING ANALYSIS.	
Resources	Processor Time	00:00:00.00	
	Elapsed Time	00:00:00.00	
	Number of Cases Allowed^a^	314572	

a. Based on availability of workspace memory.	


Descriptive Statistics	
	N	Mean	Std. Deviation	Minimum	Maximum	Percentiles	
						25th	
B_ser_Uric_Acid	64	4.6711	1.25134	2.25	9.47	3.8125	
Diff_ser_Uric_Acid	64	.3153	1.33874	-2.69	5.81	-.3975	
B_Ser_Blood_Urea	64	20.2217	4.39708	12.39	32.63	16.9250	
Diff_Ser_Blood_Urea	64	1.8956	4.83452	-7.73	15.84	-1.7575	
Groups	64	1.5000	.50395	1.00	2.00	1.0000	

Descriptive Statistics	
	Percentiles	
	50th (Median)	75th	
B_ser_Uric_Acid	4.6150	5.3575	
Diff_ser_Uric_Acid	.2850	1.1275	
B_Ser_Blood_Urea	19.2200	22.6400	
Diff_Ser_Blood_Urea	2.2750	4.7975	
Groups	1.5000	2.0000	


Mann-Whitney Test


Ranks	
	Groups	N	Mean Rank	Sum of Ranks	
B_ser_Uric_Acid	1.00	32	29.50	944.00	
	2.00	32	35.50	1136.00	
	Total	64			
Diff_ser_Uric_Acid	1.00	32	31.52	1008.50	
	2.00	32	33.48	1071.50	
	Total	64			
B_Ser_Blood_Urea	1.00	32	35.80	1145.50	
	2.00	32	29.20	934.50	
	Total	64			
Diff_Ser_Blood_Urea	1.00	32	34.11	1091.50	
	2.00	32	30.89	988.50	
	Total	64			


Test Statistics^a^	
	B_ser_Uric_Acid	Diff_ser_Uric_Acid	B_Ser_Blood_Urea	Diff_Ser_Blood_Urea	
Mann-Whitney U	416.000	480.500	406.500	460.500	
Wilcoxon W	944.000	1008.500	934.500	988.500	
Z	-1.289	-.423	-1.417	-.692	
Asymp. Sig. (2-tailed)	.197	.672	.157	.489	

a. Grouping Variable: Groups	


NPar Tests


Notes	
Output Created	11-MAY-2026 15:05:53	
Comments		
Input	Active Dataset	DataSet0	
	Filter	<none>	
	Weight	<none>	
	Split File	<none>	
	N of Rows in Working Data File	64	
Missing Value Handling	Definition of Missing	User-defined missing values are treated as missing.	
	Cases Used	Statistics for each test are based on all cases with valid data for the variable(s) used in that test.	
Syntax	NPAR TESTS
  /WILCOXON=B_ser_Uric_Acid_1 B_Ser_Blood_Urea_1 B_ser_Uric_Acid_2 B_Ser_Blood_Urea_2 WITH
    W12_ser_Uric_Acid_1 W12_Ser_Blood_Urea_1 W12_ser_Uric_Acid_2 W12_Ser_Blood_Urea_2 (PAIRED)
  /STATISTICS DESCRIPTIVES QUARTILES
  /MISSING ANALYSIS.	
Resources	Processor Time	00:00:00.00	
	Elapsed Time	00:00:00.00	
	Number of Cases Allowed^a^	241979	

a. Based on availability of workspace memory.	


Descriptive Statistics	
	N	Mean	Std. Deviation	Minimum	Maximum	Percentiles	
						25th	
B_ser_Uric_Acid_1	32	4.5316	1.28448	2.60	9.47	3.6450	
B_Ser_Blood_Urea_1	32	20.9497	4.51395	13.51	31.87	17.4300	
B_ser_Uric_Acid_2	32	4.8106	1.22154	2.25	7.05	3.8350	
B_Ser_Blood_Urea_2	32	19.4937	4.22168	12.39	32.63	16.7900	
W12_ser_Uric_Acid_1	32	4.2344	1.03987	2.65	6.26	3.5375	
W12_Ser_Blood_Urea_1	32	18.7328	4.61196	11.38	32.98	15.4625	
W12_ser_Uric_Acid_2	32	4.4772	1.10633	2.65	7.04	3.8225	
W12_Ser_Blood_Urea_2	32	17.9194	3.80296	9.08	25.85	15.4300	

Descriptive Statistics	
	Percentiles	
	50th (Median)	75th	
B_ser_Uric_Acid_1	4.4600	4.9500	
B_Ser_Blood_Urea_1	20.0050	24.6875	
B_ser_Uric_Acid_2	4.7500	5.6025	
B_Ser_Blood_Urea_2	18.3100	21.6175	
W12_ser_Uric_Acid_1	4.1600	4.8700	
W12_Ser_Blood_Urea_1	19.4000	21.6250	
W12_ser_Uric_Acid_2	4.3550	4.8450	
W12_Ser_Blood_Urea_2	16.3900	21.1275	


Wilcoxon Signed Ranks Test


Ranks	
	N	Mean Rank	Sum of Ranks	
W12_ser_Uric_Acid_1 - B_ser_Uric_Acid_1	Negative Ranks	19^a^	17.61	334.50	
	Positive Ranks	13^b^	14.88	193.50	
	Ties	0^c^			
	Total	32			
W12_Ser_Blood_Urea_1 - B_Ser_Blood_Urea_1	Negative Ranks	22^d^	17.43	383.50	
	Positive Ranks	9^e^	12.50	112.50	
	Ties	1^f^			
	Total	32			
W12_ser_Uric_Acid_2 - B_ser_Uric_Acid_2	Negative Ranks	19^g^	17.79	338.00	
	Positive Ranks	13^h^	14.62	190.00	
	Ties	0^i^			
	Total	32			
W12_Ser_Blood_Urea_2 - B_Ser_Blood_Urea_2	Negative Ranks	19^j^	18.53	352.00	
	Positive Ranks	13^k^	13.54	176.00	
	Ties	0^l^			
	Total	32			

a. W12_ser_Uric_Acid_1 < B_ser_Uric_Acid_1	
b. W12_ser_Uric_Acid_1 > B_ser_Uric_Acid_1	
c. W12_ser_Uric_Acid_1 = B_ser_Uric_Acid_1	
d. W12_Ser_Blood_Urea_1 < B_Ser_Blood_Urea_1	
e. W12_Ser_Blood_Urea_1 > B_Ser_Blood_Urea_1	
f. W12_Ser_Blood_Urea_1 = B_Ser_Blood_Urea_1	
g. W12_ser_Uric_Acid_2 < B_ser_Uric_Acid_2	
h. W12_ser_Uric_Acid_2 > B_ser_Uric_Acid_2	
i. W12_ser_Uric_Acid_2 = B_ser_Uric_Acid_2	
j. W12_Ser_Blood_Urea_2 < B_Ser_Blood_Urea_2	
k. W12_Ser_Blood_Urea_2 > B_Ser_Blood_Urea_2	
l. W12_Ser_Blood_Urea_2 = B_Ser_Blood_Urea_2	


Test Statistics^a^	
	W12_ser_Uric_Acid_1 - B_ser_Uric_Acid_1	W12_Ser_Blood_Urea_1 - B_Ser_Blood_Urea_1	W12_ser_Uric_Acid_2 - B_ser_Uric_Acid_2	W12_Ser_Blood_Urea_2 - B_Ser_Blood_Urea_2	
Z	-1.318^b^	-2.655^b^	-1.384^b^	-1.646^b^	
Asymp. Sig. (2-tailed)	.187	.008	.166	.100	

a. Wilcoxon Signed Ranks Test	
b. Based on positive ranks.	
