## Supplementary material for "Effects of a standardised *Amaranthus tricolor*–*Spinacia oleracea* extract on exercise performance, muscle strength, endurance and recovery in physically active adults: a randomised, double-blind, placebo-controlled trial": Raw statistical outputs: lipid profile_HDL- By Non parametric test.rtf

NPar Tests


Notes	
Output Created	11-MAY-2026 15:14:56	
Comments		
Input	Active Dataset	DataSet0	
	Filter	<none>	
	Weight	<none>	
	Split File	<none>	
	N of Rows in Working Data File	64	
Missing Value Handling	Definition of Missing	User-defined missing values are treated as missing.	
	Cases Used	Statistics for each test are based on all cases with valid data for the variable(s) used in that test.	
Syntax	NPAR TESTS
  /M-W= B_HDL_choles Diff_HDL_choles BY Groups(1 2)
  /STATISTICS=DESCRIPTIVES QUARTILES
  /MISSING ANALYSIS.	
Resources	Processor Time	00:00:00.00	
	Elapsed Time	00:00:00.00	
	Number of Cases Allowed^a^	393216	

a. Based on availability of workspace memory.	


Descriptive Statistics	
	N	Mean	Std. Deviation	Minimum	Maximum	Percentiles	
						25th	
B_HDL_choles	64	55.5653	11.64329	34.80	77.87	46.0275	
Diff_HDL_choles	64	4.1903	14.01854	-31.74	34.67	-4.7400	
Groups	64	1.5000	.50395	1.00	2.00	1.0000	

Descriptive Statistics	
	Percentiles	
	50th (Median)	75th	
B_HDL_choles	53.5050	66.3175	
Diff_HDL_choles	4.3150	13.3825	
Groups	1.5000	2.0000	


Mann-Whitney Test


Ranks	
	Groups	N	Mean Rank	Sum of Ranks	
B_HDL_choles	1.00	32	30.94	990.00	
	2.00	32	34.06	1090.00	
	Total	64			
Diff_HDL_choles	1.00	32	31.61	1011.50	
	2.00	32	33.39	1068.50	
	Total	64			


Test Statistics^a^	
	B_HDL_choles	Diff_HDL_choles	
Mann-Whitney U	462.000	483.500	
Wilcoxon W	990.000	1011.500	
Z	-.671	-.383	
Asymp. Sig. (2-tailed)	.502	.702	

a. Grouping Variable: Groups	


NPar Tests


Notes	
Output Created	11-MAY-2026 15:15:11	
Comments		
Input	Active Dataset	DataSet0	
	Filter	<none>	
	Weight	<none>	
	Split File	<none>	
	N of Rows in Working Data File	64	
Missing Value Handling	Definition of Missing	User-defined missing values are treated as missing.	
	Cases Used	Statistics for each test are based on all cases with valid data for the variable(s) used in that test.	
Syntax	NPAR TESTS
  /WILCOXON=B_HDL_choles_1 B_HDL_choles_2 WITH W12_HDL_choles_1 W12_HDL_choles_2 (PAIRED)
  /STATISTICS DESCRIPTIVES QUARTILES
  /MISSING ANALYSIS.	
Resources	Processor Time	00:00:00.00	
	Elapsed Time	00:00:00.00	
	Number of Cases Allowed^a^	349525	

a. Based on availability of workspace memory.	


Descriptive Statistics	
	N	Mean	Std. Deviation	Minimum	Maximum	Percentiles	
						25th	
B_HDL_choles_1	32	54.8219	11.48234	37.32	75.42	45.7050	
B_HDL_choles_2	32	56.3088	11.93818	34.80	77.87	47.4450	
W12_HDL_choles_1	32	51.5647	11.97256	30.17	80.89	41.0900	
W12_HDL_choles_2	32	51.1853	11.93318	33.11	76.40	43.2425	

Descriptive Statistics	
	Percentiles	
	50th (Median)	75th	
B_HDL_choles_1	52.7600	66.3175	
B_HDL_choles_2	54.2350	67.7350	
W12_HDL_choles_1	51.1600	59.1475	
W12_HDL_choles_2	46.7200	59.0675	


Wilcoxon Signed Ranks Test


Ranks	
	N	Mean Rank	Sum of Ranks	
W12_HDL_choles_1 - B_HDL_choles_1	Negative Ranks	20^a^	17.23	344.50	
	Positive Ranks	12^b^	15.29	183.50	
	Ties	0^c^			
	Total	32			
W12_HDL_choles_2 - B_HDL_choles_2	Negative Ranks	23^d^	16.52	380.00	
	Positive Ranks	9^e^	16.44	148.00	
	Ties	0^f^			
	Total	32			

a. W12_HDL_choles_1 < B_HDL_choles_1	
b. W12_HDL_choles_1 > B_HDL_choles_1	
c. W12_HDL_choles_1 = B_HDL_choles_1	
d. W12_HDL_choles_2 < B_HDL_choles_2	
e. W12_HDL_choles_2 > B_HDL_choles_2	
f. W12_HDL_choles_2 = B_HDL_choles_2	


Test Statistics^a^	
	W12_HDL_choles_1 - B_HDL_choles_1	W12_HDL_choles_2 - B_HDL_choles_2	
Z	-1.505^b^	-2.169^b^	
Asymp. Sig. (2-tailed)	.132	.030	

a. Wilcoxon Signed Ranks Test	
b. Based on positive ranks.	
