## Supplementary material for "Effects of a standardised *Amaranthus tricolor*–*Spinacia oleracea* extract on exercise performance, muscle strength, endurance and recovery in physically active adults: a randomised, double-blind, placebo-controlled trial": Raw statistical outputs: Lipid profile_normal by ANCOVA test.rtf

Univariate Analysis of Variance


Notes	
Output Created	11-MAY-2026 12:33:39	
Comments		
Input	Active Dataset	DataSet0	
	Filter	<none>	
	Weight	<none>	
	Split File	<none>	
	N of Rows in Working Data File	64	
Missing Value Handling	Definition of Missing	User-defined missing values are treated as missing.	
	Cases Used	Statistics are based on all cases with valid data for all variables in the model.	
Syntax	UNIANOVA W12_Total_Choles BY Groups WITH B_Total_Choles
  /METHOD=SSTYPE(3)
  /INTERCEPT=INCLUDE
  /EMMEANS=TABLES(Groups) WITH(B_Total_Choles=MEAN) COMPARE ADJ(BONFERRONI)
  /PRINT ETASQ DESCRIPTIVE
  /CRITERIA=ALPHA(.05)
  /DESIGN=B_Total_Choles Groups.	
Resources	Processor Time	00:00:00.00	
	Elapsed Time	00:00:00.00	


Between-Subjects Factors	
	N	
Groups	1	32	
	2	32	


Descriptive Statistics	
Dependent Variable:   W12_Total_Choles  	
Groups	Mean	Std. Deviation	N	
1	169.1431	28.68579	32	
2	159.9769	37.25327	32	
Total	164.5600	33.30366	64	


Tests of Between-Subjects Effects	
Dependent Variable:   W12_Total_Choles  	
Source	Type III Sum of Squares	df	Mean Square	F	Sig.	Partial Eta Squared	
Corrected Model	7940.159^a^	2	3970.079	3.910	.025	.114	
Intercept	23626.387	1	23626.387	23.270	<.001	.276	
B_Total_Choles	6595.837	1	6595.837	6.496	.013	.096	
Groups	1192.817	1	1192.817	1.175	.283	.019	
Error	61935.254	61	1015.332				
Total	1802995.003	64					
Corrected Total	69875.412	63					

a. R Squared = .114 (Adjusted R Squared = .085)	


Estimated Marginal Means


Groups


Estimates	
Dependent Variable:   W12_Total_Choles  	
Groups	Mean	Std. Error	95% Confidence Interval	
			Lower Bound	Upper Bound	
1	168.879^a^	5.634	157.613	180.144	
2	160.241^a^	5.634	148.976	171.507	

a. Covariates appearing in the model are evaluated at the following values: B_Total_Choles = 169.9297.	


Pairwise Comparisons	
Dependent Variable:   W12_Total_Choles  	
(I) Groups	(J) Groups	Mean Difference (I-J)	Std. Error	Sig.^a^	95% Confidence Interval for Difference^a^	
					Lower Bound	Upper Bound	
1	2	8.637	7.969	.283	-7.297	24.572	
2	1	-8.637	7.969	.283	-24.572	7.297	

Based on estimated marginal means	
a. Adjustment for multiple comparisons: Bonferroni.	


Univariate Tests	
Dependent Variable:   W12_Total_Choles  	
	Sum of Squares	df	Mean Square	F	Sig.	Partial Eta Squared	
Contrast	1192.817	1	1192.817	1.175	.283	.019	
Error	61935.254	61	1015.332				

The F tests the effect of Groups. This test is based on the linearly independent pairwise comparisons among the estimated marginal means.	


Univariate Analysis of Variance


Notes	
Output Created	11-MAY-2026 12:33:51	
Comments		
Input	Active Dataset	DataSet0	
	Filter	<none>	
	Weight	<none>	
	Split File	<none>	
	N of Rows in Working Data File	64	
Missing Value Handling	Definition of Missing	User-defined missing values are treated as missing.	
	Cases Used	Statistics are based on all cases with valid data for all variables in the model.	
Syntax	UNIANOVA W12_Triglyceride BY Groups WITH B_Triglyceride
  /METHOD=SSTYPE(3)
  /INTERCEPT=INCLUDE
  /EMMEANS=TABLES(Groups) WITH(B_Triglyceride=MEAN) COMPARE ADJ(BONFERRONI)
  /PRINT ETASQ DESCRIPTIVE
  /CRITERIA=ALPHA(.05)
  /DESIGN=B_Triglyceride Groups.	
Resources	Processor Time	00:00:00.00	
	Elapsed Time	00:00:00.02	


Between-Subjects Factors	
	N	
Groups	1	32	
	2	32	


Descriptive Statistics	
Dependent Variable:   W12_Triglyceride  	
Groups	Mean	Std. Deviation	N	
1	125.6797	33.48296	32	
2	127.2406	31.33827	32	
Total	126.4602	32.17957	64	


Tests of Between-Subjects Effects	
Dependent Variable:   W12_Triglyceride  	
Source	Type III Sum of Squares	df	Mean Square	F	Sig.	Partial Eta Squared	
Corrected Model	2499.028^a^	2	1249.514	1.215	.304	.038	
Intercept	42642.854	1	42642.854	41.461	<.001	.405	
B_Triglyceride	2460.044	1	2460.044	2.392	.127	.038	
Groups	28.149	1	28.149	.027	.869	.000	
Error	62739.028	61	1028.509				
Total	1088737.008	64					
Corrected Total	65238.056	63					

a. R Squared = .038 (Adjusted R Squared = .007)	


Estimated Marginal Means


Groups


Estimates	
Dependent Variable:   W12_Triglyceride  	
Groups	Mean	Std. Error	95% Confidence Interval	
			Lower Bound	Upper Bound	
1	125.797^a^	5.670	114.459	137.134	
2	127.123^a^	5.670	115.786	138.461	

a. Covariates appearing in the model are evaluated at the following values: B_Triglyceride = 130.2648.	


Pairwise Comparisons	
Dependent Variable:   W12_Triglyceride  	
(I) Groups	(J) Groups	Mean Difference (I-J)	Std. Error	Sig.^a^	95% Confidence Interval for Difference^a^	
					Lower Bound	Upper Bound	
1	2	-1.327	8.019	.869	-17.362	14.708	
2	1	1.327	8.019	.869	-14.708	17.362	

Based on estimated marginal means	
a. Adjustment for multiple comparisons: Bonferroni.	


Univariate Tests	
Dependent Variable:   W12_Triglyceride  	
	Sum of Squares	df	Mean Square	F	Sig.	Partial Eta Squared	
Contrast	28.149	1	28.149	.027	.869	.000	
Error	62739.028	61	1028.509				

The F tests the effect of Groups. This test is based on the linearly independent pairwise comparisons among the estimated marginal means.	


Univariate Analysis of Variance


Notes	
Output Created	11-MAY-2026 12:34:08	
Comments		
Input	Active Dataset	DataSet0	
	Filter	<none>	
	Weight	<none>	
	Split File	<none>	
	N of Rows in Working Data File	64	
Missing Value Handling	Definition of Missing	User-defined missing values are treated as missing.	
	Cases Used	Statistics are based on all cases with valid data for all variables in the model.	
Syntax	UNIANOVA W12_LDL_Choles BY Groups WITH B_LDL_Choles
  /METHOD=SSTYPE(3)
  /INTERCEPT=INCLUDE
  /EMMEANS=TABLES(Groups) WITH(B_LDL_Choles=MEAN) COMPARE ADJ(BONFERRONI)
  /PRINT ETASQ DESCRIPTIVE
  /CRITERIA=ALPHA(.05)
  /DESIGN=B_LDL_Choles Groups.	
Resources	Processor Time	00:00:00.02	
	Elapsed Time	00:00:00.00	


Between-Subjects Factors	
	N	
Groups	1	32	
	2	32	


Descriptive Statistics	
Dependent Variable:   W12_LDL_Choles  	
Groups	Mean	Std. Deviation	N	
1	92.4425	25.77866	32	
2	83.3434	34.29541	32	
Total	87.8930	30.44298	64	


Tests of Between-Subjects Effects	
Dependent Variable:   W12_LDL_Choles  	
Source	Type III Sum of Squares	df	Mean Square	F	Sig.	Partial Eta Squared	
Corrected Model	3344.760^a^	2	1672.380	1.853	.165	.057	
Intercept	32889.290	1	32889.290	36.449	<.001	.374	
B_LDL_Choles	2020.073	1	2020.073	2.239	.140	.035	
Groups	1143.353	1	1143.353	1.267	.265	.020	
Error	55042.068	61	902.329				
Total	552797.962	64					
Corrected Total	58386.829	63					

a. R Squared = .057 (Adjusted R Squared = .026)	


Estimated Marginal Means


Groups


Estimates	
Dependent Variable:   W12_LDL_Choles  	
Groups	Mean	Std. Error	95% Confidence Interval	
			Lower Bound	Upper Bound	
1	92.126^a^	5.314	81.500	102.753	
2	83.660^a^	5.314	73.033	94.286	

a. Covariates appearing in the model are evaluated at the following values: B_LDL_Choles = 88.3114.	


Pairwise Comparisons	
Dependent Variable:   W12_LDL_Choles  	
(I) Groups	(J) Groups	Mean Difference (I-J)	Std. Error	Sig.^a^	95% Confidence Interval for Difference^a^	
					Lower Bound	Upper Bound	
1	2	8.467	7.522	.265	-6.574	23.507	
2	1	-8.467	7.522	.265	-23.507	6.574	

Based on estimated marginal means	
a. Adjustment for multiple comparisons: Bonferroni.	


Univariate Tests	
Dependent Variable:   W12_LDL_Choles  	
	Sum of Squares	df	Mean Square	F	Sig.	Partial Eta Squared	
Contrast	1143.353	1	1143.353	1.267	.265	.020	
Error	55042.068	61	902.329				

The F tests the effect of Groups. This test is based on the linearly independent pairwise comparisons among the estimated marginal means.	


Univariate Analysis of Variance


Notes	
Output Created	11-MAY-2026 12:34:21	
Comments		
Input	Active Dataset	DataSet0	
	Filter	<none>	
	Weight	<none>	
	Split File	<none>	
	N of Rows in Working Data File	64	
Missing Value Handling	Definition of Missing	User-defined missing values are treated as missing.	
	Cases Used	Statistics are based on all cases with valid data for all variables in the model.	
Syntax	UNIANOVA W12_VLDL_Choles BY Groups WITH B_VLDL_Choles
  /METHOD=SSTYPE(3)
  /INTERCEPT=INCLUDE
  /EMMEANS=TABLES(Groups) WITH(B_VLDL_Choles=MEAN) COMPARE ADJ(BONFERRONI)
  /PRINT ETASQ DESCRIPTIVE
  /CRITERIA=ALPHA(.05)
  /DESIGN=B_VLDL_Choles Groups.	
Resources	Processor Time	00:00:00.02	
	Elapsed Time	00:00:00.00	


Between-Subjects Factors	
	N	
Groups	1	32	
	2	32	


Descriptive Statistics	
Dependent Variable:   W12_VLDL_Choles  	
Groups	Mean	Std. Deviation	N	
1	25.1359	6.69659	32	
2	25.4481	6.26765	32	
Total	25.2920	6.43591	64	


Tests of Between-Subjects Effects	
Dependent Variable:   W12_VLDL_Choles  	
Source	Type III Sum of Squares	df	Mean Square	F	Sig.	Partial Eta Squared	
Corrected Model	99.961^a^	2	49.981	1.215	.304	.038	
Intercept	1705.714	1	1705.714	41.461	<.001	.405	
B_VLDL_Choles	98.402	1	98.402	2.392	.127	.038	
Groups	1.126	1	1.126	.027	.869	.000	
Error	2509.561	61	41.140				
Total	43549.480	64					
Corrected Total	2609.522	63					

a. R Squared = .038 (Adjusted R Squared = .007)	


Estimated Marginal Means


Groups


Estimates	
Dependent Variable:   W12_VLDL_Choles  	
Groups	Mean	Std. Error	95% Confidence Interval	
			Lower Bound	Upper Bound	
1	25.159^a^	1.134	22.892	27.427	
2	25.425^a^	1.134	23.157	27.692	

a. Covariates appearing in the model are evaluated at the following values: B_VLDL_Choles = 26.0530.	


Pairwise Comparisons	
Dependent Variable:   W12_VLDL_Choles  	
(I) Groups	(J) Groups	Mean Difference (I-J)	Std. Error	Sig.^a^	95% Confidence Interval for Difference^a^	
					Lower Bound	Upper Bound	
1	2	-.265	1.604	.869	-3.472	2.942	
2	1	.265	1.604	.869	-2.942	3.472	

Based on estimated marginal means	
a. Adjustment for multiple comparisons: Bonferroni.	


Univariate Tests	
Dependent Variable:   W12_VLDL_Choles  	
	Sum of Squares	df	Mean Square	F	Sig.	Partial Eta Squared	
Contrast	1.126	1	1.126	.027	.869	.000	
Error	2509.561	61	41.140				

The F tests the effect of Groups. This test is based on the linearly independent pairwise comparisons among the estimated marginal means.	


Univariate Analysis of Variance


Notes	
Output Created	11-MAY-2026 12:34:36	
Comments		
Input	Active Dataset	DataSet0	
	Filter	<none>	
	Weight	<none>	
	Split File	<none>	
	N of Rows in Working Data File	64	
Missing Value Handling	Definition of Missing	User-defined missing values are treated as missing.	
	Cases Used	Statistics are based on all cases with valid data for all variables in the model.	
Syntax	UNIANOVA W12_TC_HDL_Ratio BY Groups WITH B_TC_HDL_Ratio
  /METHOD=SSTYPE(3)
  /INTERCEPT=INCLUDE
  /EMMEANS=TABLES(Groups) WITH(B_TC_HDL_Ratio=MEAN) COMPARE ADJ(BONFERRONI)
  /PRINT ETASQ DESCRIPTIVE
  /CRITERIA=ALPHA(.05)
  /DESIGN=B_TC_HDL_Ratio Groups.	
Resources	Processor Time	00:00:00.00	
	Elapsed Time	00:00:00.01	


Between-Subjects Factors	
	N	
Groups	1	32	
	2	32	


Descriptive Statistics	
Dependent Variable:   W12_TC_HDL_Ratio  	
Groups	Mean	Std. Deviation	N	
1	3.3813	.68343	32	
2	3.2225	.80002	32	
Total	3.3019	.74240	64	


Tests of Between-Subjects Effects	
Dependent Variable:   W12_TC_HDL_Ratio  	
Source	Type III Sum of Squares	df	Mean Square	F	Sig.	
Corrected Model	.590^a^	2	.295	.527	.593	
Intercept	31.915	1	31.915	57.035	<.001	
B_TC_HDL_Ratio	.186	1	.186	.333	.566	
Groups	.353	1	.353	.631	.430	
Error	34.134	61	.560			
Total	732.476	64				
Corrected Total	34.723	63				

Tests of Between-Subjects Effects	
Dependent Variable:   W12_TC_HDL_Ratio  	
Source	Partial Eta Squared	
Corrected Model	.017	
Intercept	.483	
B_TC_HDL_Ratio	.005	
Groups	.010	
Error		
Total		
Corrected Total		

a. R Squared = .017 (Adjusted R Squared = -.015)	


Estimated Marginal Means


Groups


Estimates	
Dependent Variable:   W12_TC_HDL_Ratio  	
Groups	Mean	Std. Error	95% Confidence Interval	
			Lower Bound	Upper Bound	
1	3.376^a^	.132	3.111	3.641	
2	3.227^a^	.132	2.962	3.492	

a. Covariates appearing in the model are evaluated at the following values: B_TC_HDL_Ratio = 3.1644.	


Pairwise Comparisons	
Dependent Variable:   W12_TC_HDL_Ratio  	
(I) Groups	(J) Groups	Mean Difference (I-J)	Std. Error	Sig.^a^	95% Confidence Interval for Difference^a^	
					Lower Bound	Upper Bound	
1	2	.149	.188	.430	-.226	.525	
2	1	-.149	.188	.430	-.525	.226	

Based on estimated marginal means	
a. Adjustment for multiple comparisons: Bonferroni.	


Univariate Tests	
Dependent Variable:   W12_TC_HDL_Ratio  	
	Sum of Squares	df	Mean Square	F	Sig.	Partial Eta Squared	
Contrast	.353	1	.353	.631	.430	.010	
Error	34.134	61	.560				

The F tests the effect of Groups. This test is based on the linearly independent pairwise comparisons among the estimated marginal means.	


Univariate Analysis of Variance


Notes	
Output Created	11-MAY-2026 12:34:49	
Comments		
Input	Active Dataset	DataSet0	
	Filter	<none>	
	Weight	<none>	
	Split File	<none>	
	N of Rows in Working Data File	64	
Missing Value Handling	Definition of Missing	User-defined missing values are treated as missing.	
	Cases Used	Statistics are based on all cases with valid data for all variables in the model.	
Syntax	UNIANOVA W12_LDL_HDL_Ratio BY Groups WITH B_LDL_HDL_Ratio
  /METHOD=SSTYPE(3)
  /INTERCEPT=INCLUDE
  /EMMEANS=TABLES(Groups) WITH(B_LDL_HDL_Ratio=MEAN) COMPARE ADJ(BONFERRONI)
  /PRINT ETASQ DESCRIPTIVE
  /CRITERIA=ALPHA(.05)
  /DESIGN=B_LDL_HDL_Ratio Groups.	
Resources	Processor Time	00:00:00.02	
	Elapsed Time	00:00:00.00	


Between-Subjects Factors	
	N	
Groups	1	32	
	2	32	


Descriptive Statistics	
Dependent Variable:   W12_LDL_HDL_Ratio  	
Groups	Mean	Std. Deviation	N	
1	1.8531	.60083	32	
2	1.6941	.70922	32	
Total	1.7736	.65693	64	


Tests of Between-Subjects Effects	
Dependent Variable:   W12_LDL_HDL_Ratio  	
Source	Type III Sum of Squares	df	Mean Square	F	Sig.	
Corrected Model	.450^a^	2	.225	.514	.601	
Intercept	24.619	1	24.619	56.166	<.001	
B_LDL_HDL_Ratio	.046	1	.046	.104	.748	
Groups	.375	1	.375	.854	.359	
Error	26.738	61	.438			
Total	228.509	64				
Corrected Total	27.188	63				

Tests of Between-Subjects Effects	
Dependent Variable:   W12_LDL_HDL_Ratio  	
Source	Partial Eta Squared	
Corrected Model	.017	
Intercept	.479	
B_LDL_HDL_Ratio	.002	
Groups	.014	
Error		
Total		
Corrected Total		

a. R Squared = .017 (Adjusted R Squared = -.016)	


Estimated Marginal Means


Groups


Estimates	
Dependent Variable:   W12_LDL_HDL_Ratio  	
Groups	Mean	Std. Error	95% Confidence Interval	
			Lower Bound	Upper Bound	
1	1.850^a^	.117	1.616	2.085	
2	1.697^a^	.117	1.462	1.931	

a. Covariates appearing in the model are evaluated at the following values: B_LDL_HDL_Ratio = 1.6663.	


Pairwise Comparisons	
Dependent Variable:   W12_LDL_HDL_Ratio  	
(I) Groups	(J) Groups	Mean Difference (I-J)	Std. Error	Sig.^a^	95% Confidence Interval for Difference^a^	
					Lower Bound	Upper Bound	
1	2	.154	.166	.359	-.179	.486	
2	1	-.154	.166	.359	-.486	.179	

Based on estimated marginal means	
a. Adjustment for multiple comparisons: Bonferroni.	


Univariate Tests	
Dependent Variable:   W12_LDL_HDL_Ratio  	
	Sum of Squares	df	Mean Square	F	Sig.	Partial Eta Squared	
Contrast	.375	1	.375	.854	.359	.014	
Error	26.738	61	.438				

The F tests the effect of Groups. This test is based on the linearly independent pairwise comparisons among the estimated marginal means.	


T-Test


Notes	
Output Created	11-MAY-2026 12:37:06	
Comments		
Input	Active Dataset	DataSet0	
	Filter	<none>	
	Weight	<none>	
	Split File	<none>	
	N of Rows in Working Data File	64	
Missing Value Handling	Definition of Missing	User defined missing values are treated as missing.	
	Cases Used	Statistics for each analysis are based on the cases with no missing or out-of-range data for any variable in the analysis.	
Syntax	T-TEST PAIRS=B_Total_Choles_1 B_Triglyceride_1 B_LDL_Choles_1 B_VLDL_Choles_1 B_TC_HDL_Ratio_1
    B_LDL_HDL_Ratio_1 B_Total_Choles_2 B_Triglyceride_2 B_LDL_Choles_2 B_VLDL_Choles_2 B_TC_HDL_Ratio_2
    B_LDL_HDL_Ratio_2 WITH W12_Total_Choles_1 W12_Triglyceride_1 W12_LDL_Choles_1 W12_VLDL_Choles_1
    W12_TC_HDL_Ratio_1 W12_LDL_HDL_Ratio_1 W12_Total_Choles_2 W12_Triglyceride_2 W12_LDL_Choles_2
    W12_VLDL_Choles_2 W12_TC_HDL_Ratio_2 W12_LDL_HDL_Ratio_2 (PAIRED)
  /ES DISPLAY(TRUE) STANDARDIZER(SD)
  /CRITERIA=CI(.9500)
  /MISSING=ANALYSIS.	
Resources	Processor Time	00:00:00.00	
	Elapsed Time	00:00:00.01	


Paired Samples Statistics	
	Mean	N	Std. Deviation	Std. Error Mean	
Pair 1	B_Total_Choles_1	170.7281	32	32.35899	5.72031	
	W12_Total_Choles_1	169.1431	32	28.68579	5.07098	
Pair 2	B_Triglyceride_1	129.6250	32	33.82241	5.97901	
	W12_Triglyceride_1	125.6797	32	33.48296	5.91901	
Pair 3	B_LDL_Choles_1	89.9812	32	33.42503	5.90877	
	W12_LDL_Choles_1	92.4425	32	25.77866	4.55707	
Pair 4	B_VLDL_Choles_1	25.9250	32	6.76448	1.19580	
	W12_VLDL_Choles_1	25.1359	32	6.69659	1.18380	
Pair 5	B_TC_HDL_Ratio_1	3.2309	32	.83342	.14733	
	W12_TC_HDL_Ratio_1	3.3812	32	.68343	.12081	
Pair 6	B_LDL_HDL_Ratio_1	1.7309	32	.74656	.13197	
	W12_LDL_HDL_Ratio_1	1.8531	32	.60083	.10621	
Pair 7	B_Total_Choles_2	169.1312	32	29.85881	5.27834	
	W12_Total_Choles_2	159.9769	32	37.25327	6.58551	
Pair 8	B_Triglyceride_2	130.9047	32	34.96997	6.18188	
	W12_Triglyceride_2	127.2406	32	31.33827	5.53988	
Pair 9	B_LDL_Choles_2	86.6416	32	26.46901	4.67910	
	W12_LDL_Choles_2	83.3434	32	34.29541	6.06263	
Pair 10	B_VLDL_Choles_2	26.1809	32	6.99399	1.23638	
	W12_VLDL_Choles_2	25.4481	32	6.26765	1.10798	
Pair 11	B_TC_HDL_Ratio_2	3.0978	32	.67651	.11959	
	W12_TC_HDL_Ratio_2	3.2225	32	.80002	.14142	
Pair 12	B_LDL_HDL_Ratio_2	1.6016	32	.56265	.09946	
	W12_LDL_HDL_Ratio_2	1.6941	32	.70922	.12537	


Paired Samples Correlations	
	N	Correlation	Significance	
			One-Sided p	Two-Sided p	
Pair 1	B_Total_Choles_1 & W12_Total_Choles_1	32	.342	.028	.055	
Pair 2	B_Triglyceride_1 & W12_Triglyceride_1	32	.347	.026	.052	
Pair 3	B_LDL_Choles_1 & W12_LDL_Choles_1	32	.182	.160	.320	
Pair 4	B_VLDL_Choles_1 & W12_VLDL_Choles_1	32	.347	.026	.052	
Pair 5	B_TC_HDL_Ratio_1 & W12_TC_HDL_Ratio_1	32	-.042	.409	.818	
Pair 6	B_LDL_HDL_Ratio_1 & W12_LDL_HDL_Ratio_1	32	-.068	.356	.712	
Pair 7	B_Total_Choles_2 & W12_Total_Choles_2	32	.292	.053	.105	
Pair 8	B_Triglyceride_2 & W12_Triglyceride_2	32	.037	.420	.841	
Pair 9	B_LDL_Choles_2 & W12_LDL_Choles_2	32	.207	.128	.256	
Pair 10	B_VLDL_Choles_2 & W12_VLDL_Choles_2	32	.037	.420	.841	
Pair 11	B_TC_HDL_Ratio_2 & W12_TC_HDL_Ratio_2	32	.198	.138	.276	
Pair 12	B_LDL_HDL_Ratio_2 & W12_LDL_HDL_Ratio_2	32	.166	.182	.363	


Paired Samples Test	
	Paired Differences	
	Mean	Std. Deviation	Std. Error Mean	95% Confidence Interval of the Difference	
				Lower	
Pair 1	B_Total_Choles_1 - W12_Total_Choles_1	1.58500	35.13492	6.21104	-11.08249	
Pair 2	B_Triglyceride_1 - W12_Triglyceride_1	3.94531	38.46300	6.79936	-9.92208	
Pair 3	B_LDL_Choles_1 - W12_LDL_Choles_1	-2.46125	38.32521	6.77500	-16.27896	
Pair 4	B_VLDL_Choles_1 - W12_VLDL_Choles_1	.78906	7.69260	1.35987	-1.98442	
Pair 5	B_TC_HDL_Ratio_1 - W12_TC_HDL_Ratio_1	-.15031	1.09999	.19445	-.54690	
Pair 6	B_LDL_HDL_Ratio_1 - W12_LDL_HDL_Ratio_1	-.12219	.98957	.17493	-.47896	
Pair 7	B_Total_Choles_2 - W12_Total_Choles_2	9.15437	40.37784	7.13786	-5.40339	
Pair 8	B_Triglyceride_2 - W12_Triglyceride_2	3.66406	46.08549	8.14684	-12.95153	
Pair 9	B_LDL_Choles_2 - W12_LDL_Choles_2	3.29812	38.74766	6.84968	-10.67190	
Pair 10	B_VLDL_Choles_2 - W12_VLDL_Choles_2	.73281	9.21710	1.62937	-2.59031	
Pair 11	B_TC_HDL_Ratio_2 - W12_TC_HDL_Ratio_2	-.12469	.93964	.16611	-.46346	
Pair 12	B_LDL_HDL_Ratio_2 - W12_LDL_HDL_Ratio_2	-.09250	.82881	.14651	-.39132	

Paired Samples Test	
	Paired Differences	t	df	Significance	
	95% Confidence Interval of the Difference			One-Sided p	
	Upper				
Pair 1	B_Total_Choles_1 - W12_Total_Choles_1	14.25249	.255	31	.400	
Pair 2	B_Triglyceride_1 - W12_Triglyceride_1	17.81270	.580	31	.283	
Pair 3	B_LDL_Choles_1 - W12_LDL_Choles_1	11.35646	-.363	31	.359	
Pair 4	B_VLDL_Choles_1 - W12_VLDL_Choles_1	3.56254	.580	31	.283	
Pair 5	B_TC_HDL_Ratio_1 - W12_TC_HDL_Ratio_1	.24628	-.773	31	.223	
Pair 6	B_LDL_HDL_Ratio_1 - W12_LDL_HDL_Ratio_1	.23459	-.698	31	.245	
Pair 7	B_Total_Choles_2 - W12_Total_Choles_2	23.71214	1.283	31	.105	
Pair 8	B_Triglyceride_2 - W12_Triglyceride_2	20.27965	.450	31	.328	
Pair 9	B_LDL_Choles_2 - W12_LDL_Choles_2	17.26815	.482	31	.317	
Pair 10	B_VLDL_Choles_2 - W12_VLDL_Choles_2	4.05593	.450	31	.328	
Pair 11	B_TC_HDL_Ratio_2 - W12_TC_HDL_Ratio_2	.21409	-.751	31	.229	
Pair 12	B_LDL_HDL_Ratio_2 - W12_LDL_HDL_Ratio_2	.20632	-.631	31	.266	


Paired Samples Test	
	Significance	
	Two-Sided p	
		
Pair 1	B_Total_Choles_1 - W12_Total_Choles_1	.800	
Pair 2	B_Triglyceride_1 - W12_Triglyceride_1	.566	
Pair 3	B_LDL_Choles_1 - W12_LDL_Choles_1	.719	
Pair 4	B_VLDL_Choles_1 - W12_VLDL_Choles_1	.566	
Pair 5	B_TC_HDL_Ratio_1 - W12_TC_HDL_Ratio_1	.445	
Pair 6	B_LDL_HDL_Ratio_1 - W12_LDL_HDL_Ratio_1	.490	
Pair 7	B_Total_Choles_2 - W12_Total_Choles_2	.209	
Pair 8	B_Triglyceride_2 - W12_Triglyceride_2	.656	
Pair 9	B_LDL_Choles_2 - W12_LDL_Choles_2	.634	
Pair 10	B_VLDL_Choles_2 - W12_VLDL_Choles_2	.656	
Pair 11	B_TC_HDL_Ratio_2 - W12_TC_HDL_Ratio_2	.459	
Pair 12	B_LDL_HDL_Ratio_2 - W12_LDL_HDL_Ratio_2	.532	


Paired Samples Effect Sizes	
	Standardizer^a^	Point Estimate	95% Confidence Interval	
			Lower	
Pair 1	B_Total_Choles_1 - W12_Total_Choles_1	Cohen's d	35.13492	.045	-.302	
		Hedges' correction	36.01452	.044	-.295	
Pair 2	B_Triglyceride_1 - W12_Triglyceride_1	Cohen's d	38.46300	.103	-.246	
		Hedges' correction	39.42592	.100	-.240	
Pair 3	B_LDL_Choles_1 - W12_LDL_Choles_1	Cohen's d	38.32521	-.064	-.411	
		Hedges' correction	39.28468	-.063	-.401	
Pair 4	B_VLDL_Choles_1 - W12_VLDL_Choles_1	Cohen's d	7.69260	.103	-.246	
		Hedges' correction	7.88518	.100	-.240	
Pair 5	B_TC_HDL_Ratio_1 - W12_TC_HDL_Ratio_1	Cohen's d	1.09999	-.137	-.484	
		Hedges' correction	1.12753	-.133	-.472	
Pair 6	B_LDL_HDL_Ratio_1 - W12_LDL_HDL_Ratio_1	Cohen's d	.98957	-.123	-.470	
		Hedges' correction	1.01434	-.120	-.459	
Pair 7	B_Total_Choles_2 - W12_Total_Choles_2	Cohen's d	40.37784	.227	-.126	
		Hedges' correction	41.38870	.221	-.123	
Pair 8	B_Triglyceride_2 - W12_Triglyceride_2	Cohen's d	46.08549	.080	-.268	
		Hedges' correction	47.23924	.078	-.262	
Pair 9	B_LDL_Choles_2 - W12_LDL_Choles_2	Cohen's d	38.74766	.085	-.263	
		Hedges' correction	39.71771	.083	-.256	
Pair 10	B_VLDL_Choles_2 - W12_VLDL_Choles_2	Cohen's d	9.21710	.080	-.268	
		Hedges' correction	9.44785	.078	-.262	
Pair 11	B_TC_HDL_Ratio_2 - W12_TC_HDL_Ratio_2	Cohen's d	.93964	-.133	-.480	
		Hedges' correction	.96317	-.129	-.468	
Pair 12	B_LDL_HDL_Ratio_2 - W12_LDL_HDL_Ratio_2	Cohen's d	.82881	-.112	-.458	
		Hedges' correction	.84956	-.109	-.447	

Paired Samples Effect Sizes	
	95% Confidence Interval^a^	
	Upper	
Pair 1	B_Total_Choles_1 - W12_Total_Choles_1	Cohen's d	.391	
		Hedges' correction	.382	
Pair 2	B_Triglyceride_1 - W12_Triglyceride_1	Cohen's d	.449	
		Hedges' correction	.438	
Pair 3	B_LDL_Choles_1 - W12_LDL_Choles_1	Cohen's d	.283	
		Hedges' correction	.276	
Pair 4	B_VLDL_Choles_1 - W12_VLDL_Choles_1	Cohen's d	.449	
		Hedges' correction	.438	
Pair 5	B_TC_HDL_Ratio_1 - W12_TC_HDL_Ratio_1	Cohen's d	.213	
		Hedges' correction	.207	
Pair 6	B_LDL_HDL_Ratio_1 - W12_LDL_HDL_Ratio_1	Cohen's d	.225	
		Hedges' correction	.220	
Pair 7	B_Total_Choles_2 - W12_Total_Choles_2	Cohen's d	.576	
		Hedges' correction	.562	
Pair 8	B_Triglyceride_2 - W12_Triglyceride_2	Cohen's d	.426	
		Hedges' correction	.416	
Pair 9	B_LDL_Choles_2 - W12_LDL_Choles_2	Cohen's d	.432	
		Hedges' correction	.421	
Pair 10	B_VLDL_Choles_2 - W12_VLDL_Choles_2	Cohen's d	.426	
		Hedges' correction	.416	
Pair 11	B_TC_HDL_Ratio_2 - W12_TC_HDL_Ratio_2	Cohen's d	.216	
		Hedges' correction	.211	
Pair 12	B_LDL_HDL_Ratio_2 - W12_LDL_HDL_Ratio_2	Cohen's d	.237	
		Hedges' correction	.231	

a. The denominator used in estimating the effect sizes. 
Cohen's d uses the sample standard deviation of the mean difference. 
Hedges' correction uses the sample standard deviation of the mean difference, plus a correction factor.	
