## Supplementary material for "Effects of a standardised *Amaranthus tricolor*–*Spinacia oleracea* extract on exercise performance, muscle strength, endurance and recovery in physically active adults: a randomised, double-blind, placebo-controlled trial": Raw statistical outputs: Liver function test- Non parametric test.rtf

NPar Tests


Notes	
Output Created	11-MAY-2026 15:00:04	
Comments		
Input	Active Dataset	DataSet0	
	Filter	<none>	
	Weight	<none>	
	Split File	<none>	
	N of Rows in Working Data File	64	
Missing Value Handling	Definition of Missing	User-defined missing values are treated as missing.	
	Cases Used	Statistics for each test are based on all cases with valid data for the variable(s) used in that test.	
Syntax	NPAR TESTS
  /M-W= B_Bilrub_Total Diff_Bilrub_Total B_Bilrub_Indirect Diff_Bilrub_Indirect B_SGPT Diff_SGPT
    B_Total_Protein Diff_Total_Protein B_Ser_Album Diff_Ser_Album BY Groups(1 2)
  /STATISTICS=DESCRIPTIVES QUARTILES
  /MISSING ANALYSIS.	
Resources	Processor Time	00:00:00.00	
	Elapsed Time	00:00:00.00	
	Number of Cases Allowed^a^	196608	

a. Based on availability of workspace memory.	


Descriptive Statistics	
	N	Mean	Std. Deviation	Minimum	Maximum	Percentiles	
						25th	
B_Bilrub_Total	64	.6286	.21594	.23	1.15	.4925	
Diff_Bilrub_Total	64	-.1452	.30690	-.89	.38	-.3575	
B_Bilrub_Indirect	64	.4105	.21648	.11	1.15	.2625	
Diff_Bilrub_Indirect	64	-.1173	.28725	-.89	.48	-.2975	
B_SGPT	64	30.3603	10.91419	10.25	58.44	22.7425	
Diff_SGPT	64	2.9348	13.92977	-35.08	38.73	-3.2750	
B_Total_Protein	64	6.9283	.37440	6.21	7.58	6.7000	
Diff_Total_Protein	64	.0241	.64218	-3.31	1.50	-.2800	
B_Ser_Album	64	4.0452	.43079	3.17	4.79	3.7050	
Diff_Ser_Album	64	.2109	.61474	-1.08	1.23	-.2525	
Groups	64	1.5000	.50395	1.00	2.00	1.0000	

Descriptive Statistics	
	Percentiles	
	50th (Median)	75th	
B_Bilrub_Total	.5850	.7675	
Diff_Bilrub_Total	-.1050	.1000	
B_Bilrub_Indirect	.3450	.5275	
Diff_Bilrub_Indirect	-.1000	.0975	
B_SGPT	29.7950	38.4200	
Diff_SGPT	1.4000	14.1350	
B_Total_Protein	6.9200	7.1750	
Diff_Total_Protein	.0950	.3700	
B_Ser_Album	3.9850	4.4175	
Diff_Ser_Album	.2000	.8250	
Groups	1.5000	2.0000	


Mann-Whitney Test


Ranks	
	Groups	N	Mean Rank	Sum of Ranks	
B_Bilrub_Total	1.00	32	31.23	999.50	
	2.00	32	33.77	1080.50	
	Total	64			
Diff_Bilrub_Total	1.00	32	30.44	974.00	
	2.00	32	34.56	1106.00	
	Total	64			
B_Bilrub_Indirect	1.00	32	32.41	1037.00	
	2.00	32	32.59	1043.00	
	Total	64			
Diff_Bilrub_Indirect	1.00	32	32.05	1025.50	
	2.00	32	32.95	1054.50	
	Total	64			
B_SGPT	1.00	32	30.22	967.00	
	2.00	32	34.78	1113.00	
	Total	64			
Diff_SGPT	1.00	32	27.59	883.00	
	2.00	32	37.41	1197.00	
	Total	64			
B_Total_Protein	1.00	32	30.64	980.50	
	2.00	32	34.36	1099.50	
	Total	64			
Diff_Total_Protein	1.00	32	28.66	917.00	
	2.00	32	36.34	1163.00	
	Total	64			
B_Ser_Album	1.00	32	31.34	1003.00	
	2.00	32	33.66	1077.00	
	Total	64			
Diff_Ser_Album	1.00	32	29.23	935.50	
	2.00	32	35.77	1144.50	
	Total	64			


Test Statistics^a^	
	B_Bilrub_Total	Diff_Bilrub_Total	B_Bilrub_Indirect	Diff_Bilrub_Indirect	B_SGPT	
Mann-Whitney U	471.500	446.000	509.000	497.500	439.000	
Wilcoxon W	999.500	974.000	1037.000	1025.500	967.000	
Z	-.544	-.886	-.040	-.195	-.980	
Asymp. Sig. (2-tailed)	.586	.375	.968	.846	.327	

Test Statistics^a^	
	Diff_SGPT	B_Total_Protein	Diff_Total_Protein	B_Ser_Album	Diff_Ser_Album	
Mann-Whitney U	355.000	452.500	389.000	475.000	407.500	
Wilcoxon W	883.000	980.500	917.000	1003.000	935.500	
Z	-2.108	-.799	-1.652	-.497	-1.403	
Asymp. Sig. (2-tailed)	.035	.424	.099	.619	.161	

a. Grouping Variable: Groups	


NPar Tests


Notes	
Output Created	11-MAY-2026 15:01:07	
Comments		
Input	Active Dataset	DataSet0	
	Filter	<none>	
	Weight	<none>	
	Split File	<none>	
	N of Rows in Working Data File	64	
Missing Value Handling	Definition of Missing	User-defined missing values are treated as missing.	
	Cases Used	Statistics for each test are based on all cases with valid data for the variable(s) used in that test.	
Syntax	NPAR TESTS
  /WILCOXON=B_Bilrub_Total_1 B_Bilrub_Indirect_1 B_SGPT_1 B_Total_Protein_1 B_Ser_Album_1
    B_Bilrub_Total_2 B_Bilrub_Indirect_2 B_SGPT_2 B_Total_Protein_2 B_Ser_Album_2 WITH
    W12_Bilrub_Total_1 W12_Bilrub_Indirect_1 W12_SGPT_1 W12_Total_Protein_1 W12_Ser_Album_1
    W12_Bilrub_Total_2 W12_Bilrub_Indirect_2 W12_SGPT_2 W12_Total_Protein_2 W12_Ser_Album_2 (PAIRED)
  /STATISTICS DESCRIPTIVES QUARTILES
  /MISSING ANALYSIS.	
Resources	Processor Time	00:00:00.00	
	Elapsed Time	00:00:00.00	
	Number of Cases Allowed^a^	125829	

a. Based on availability of workspace memory.	


Descriptive Statistics	
	N	Mean	Std. Deviation	Minimum	Maximum	Percentiles	
						25th	
B_Bilrub_Total_1	32	.6109	.20915	.23	1.15	.4925	
B_Bilrub_Indirect_1	32	.4131	.21818	.11	1.15	.2650	
B_SGPT_1	32	29.5353	12.42024	11.33	58.44	22.7425	
B_Total_Protein_1	32	6.8966	.35425	6.21	7.58	6.6725	
B_Ser_Album_1	32	4.0175	.39633	3.28	4.62	3.7250	
B_Bilrub_Total_2	32	.6463	.22444	.31	1.15	.4850	
B_Bilrub_Indirect_2	32	.4078	.21823	.11	1.00	.2550	
B_SGPT_2	32	31.1853	9.29592	10.25	43.48	22.1075	
B_Total_Protein_2	32	6.9600	.39661	6.22	7.54	6.7900	
B_Ser_Album_2	32	4.0728	.46742	3.17	4.79	3.5850	
W12_Bilrub_Total_1	32	.7863	.24943	.32	1.25	.6325	
W12_Bilrub_Indirect_1	32	.5366	.20553	.15	1.15	.4125	
W12_SGPT_1	32	30.2191	14.61171	4.75	69.06	18.8275	
W12_Total_Protein_1	32	6.9822	.77402	5.82	10.42	6.5275	
W12_Ser_Album_1	32	3.9184	.42968	3.26	4.67	3.5425	
W12_Bilrub_Total_2	32	.7613	.26870	.33	1.36	.5450	
W12_Bilrub_Indirect_2	32	.5191	.23503	.11	1.06	.3825	
W12_SGPT_2	32	24.6319	10.13548	8.77	39.10	14.4325	
W12_Total_Protein_2	32	6.8262	.45381	6.18	8.27	6.4875	
W12_Ser_Album_2	32	3.7500	.29483	3.34	4.57	3.5325	

Descriptive Statistics	
	Percentiles	
	50th (Median)	75th	
B_Bilrub_Total_1	.5700	.7175	
B_Bilrub_Indirect_1	.3400	.5250	
B_SGPT_1	25.3950	37.3675	
B_Total_Protein_1	6.8900	7.1600	
B_Ser_Album_1	3.8750	4.4050	
B_Bilrub_Total_2	.6150	.8250	
B_Bilrub_Indirect_2	.3500	.5275	
B_SGPT_2	34.5300	39.1025	
B_Total_Protein_2	7.0200	7.3000	
B_Ser_Album_2	4.1050	4.4750	
W12_Bilrub_Total_1	.7300	.9975	
W12_Bilrub_Indirect_1	.5150	.6450	
W12_SGPT_1	29.4600	39.5700	
W12_Total_Protein_1	6.9850	7.2250	
W12_Ser_Album_1	3.8050	4.4200	
W12_Bilrub_Total_2	.7150	.9600	
W12_Bilrub_Indirect_2	.4750	.6600	
W12_SGPT_2	24.1150	34.5575	
W12_Total_Protein_2	6.7100	7.1400	
W12_Ser_Album_2	3.7150	3.8550	


Wilcoxon Signed Ranks Test


Ranks	
	N	Mean Rank	Sum of Ranks	
W12_Bilrub_Total_1 - B_Bilrub_Total_1	Negative Ranks	8^a^	11.81	94.50	
	Positive Ranks	23^b^	17.46	401.50	
	Ties	1^c^			
	Total	32			
W12_Bilrub_Indirect_1 - B_Bilrub_Indirect_1	Negative Ranks	11^d^	12.45	137.00	
	Positive Ranks	21^e^	18.62	391.00	
	Ties	0^f^			
	Total	32			
W12_SGPT_1 - B_SGPT_1	Negative Ranks	15^g^	16.60	249.00	
	Positive Ranks	17^h^	16.41	279.00	
	Ties	0^i^			
	Total	32			
W12_Total_Protein_1 - B_Total_Protein_1	Negative Ranks	15^j^	16.07	241.00	
	Positive Ranks	17^k^	16.88	287.00	
	Ties	0^l^			
	Total	32			
W12_Ser_Album_1 - B_Ser_Album_1	Negative Ranks	16^m^	18.22	291.50	
	Positive Ranks	15^n^	13.63	204.50	
	Ties	1^o^			
	Total	32			
W12_Bilrub_Total_2 - B_Bilrub_Total_2	Negative Ranks	15^p^	11.47	172.00	
	Positive Ranks	17^q^	20.94	356.00	
	Ties	0^r^			
	Total	32			
W12_Bilrub_Indirect_2 - B_Bilrub_Indirect_2	Negative Ranks	13^s^	12.65	164.50	
	Positive Ranks	19^t^	19.13	363.50	
	Ties	0^u^			
	Total	32			
W12_SGPT_2 - B_SGPT_2	Negative Ranks	21^v^	18.98	398.50	
	Positive Ranks	11^w^	11.77	129.50	
	Ties	0^x^			
	Total	32			
W12_Total_Protein_2 - B_Total_Protein_2	Negative Ranks	22^y^	16.59	365.00	
	Positive Ranks	10^z^	16.30	163.00	
	Ties	0^aa^			
	Total	32			
W12_Ser_Album_2 - B_Ser_Album_2	Negative Ranks	22^ab^	18.27	402.00	
	Positive Ranks	10^ac^	12.60	126.00	
	Ties	0^ad^			
	Total	32			

a. W12_Bilrub_Total_1 < B_Bilrub_Total_1	
b. W12_Bilrub_Total_1 > B_Bilrub_Total_1	
c. W12_Bilrub_Total_1 = B_Bilrub_Total_1	
d. W12_Bilrub_Indirect_1 < B_Bilrub_Indirect_1	
e. W12_Bilrub_Indirect_1 > B_Bilrub_Indirect_1	
f. W12_Bilrub_Indirect_1 = B_Bilrub_Indirect_1	
g. W12_SGPT_1 < B_SGPT_1	
h. W12_SGPT_1 > B_SGPT_1	
i. W12_SGPT_1 = B_SGPT_1	
j. W12_Total_Protein_1 < B_Total_Protein_1	
k. W12_Total_Protein_1 > B_Total_Protein_1	
l. W12_Total_Protein_1 = B_Total_Protein_1	
m. W12_Ser_Album_1 < B_Ser_Album_1	
n. W12_Ser_Album_1 > B_Ser_Album_1	
o. W12_Ser_Album_1 = B_Ser_Album_1	
p. W12_Bilrub_Total_2 < B_Bilrub_Total_2	
q. W12_Bilrub_Total_2 > B_Bilrub_Total_2	
r. W12_Bilrub_Total_2 = B_Bilrub_Total_2	
s. W12_Bilrub_Indirect_2 < B_Bilrub_Indirect_2	
t. W12_Bilrub_Indirect_2 > B_Bilrub_Indirect_2	
u. W12_Bilrub_Indirect_2 = B_Bilrub_Indirect_2	
v. W12_SGPT_2 < B_SGPT_2	
w. W12_SGPT_2 > B_SGPT_2	
x. W12_SGPT_2 = B_SGPT_2	
y. W12_Total_Protein_2 < B_Total_Protein_2	
z. W12_Total_Protein_2 > B_Total_Protein_2	
aa. W12_Total_Protein_2 = B_Total_Protein_2	
ab. W12_Ser_Album_2 < B_Ser_Album_2	
ac. W12_Ser_Album_2 > B_Ser_Album_2	
ad. W12_Ser_Album_2 = B_Ser_Album_2	


Test Statistics^a^	
	W12_Bilrub_Total_1 - B_Bilrub_Total_1	W12_Bilrub_Indirect_1 - B_Bilrub_Indirect_1	W12_SGPT_1 - B_SGPT_1	W12_Total_Protein_1 - B_Total_Protein_1	
Z	-3.009^b^	-2.375^b^	-.280^b^	-.430^b^	
Asymp. Sig. (2-tailed)	.003	.018	.779	.667	

Test Statistics^a^	
	W12_Ser_Album_1 - B_Ser_Album_1	W12_Bilrub_Total_2 - B_Bilrub_Total_2	W12_Bilrub_Indirect_2 - B_Bilrub_Indirect_2	W12_SGPT_2 - B_SGPT_2	
Z	-.853^c^	-1.721^b^	-1.861^b^	-2.515^c^	
Asymp. Sig. (2-tailed)	.394	.085	.063	.012	

Test Statistics^a^	
	W12_Total_Protein_2 - B_Total_Protein_2	W12_Ser_Album_2 - B_Ser_Album_2	
Z	-1.889^c^	-2.581^c^	
Asymp. Sig. (2-tailed)	.059	.010	

a. Wilcoxon Signed Ranks Test	
b. Based on negative ranks.	
c. Based on positive ranks.	
