## Supplementary material for "Effects of a standardised *Amaranthus tricolor*–*Spinacia oleracea* extract on exercise performance, muscle strength, endurance and recovery in physically active adults: a randomised, double-blind, placebo-controlled trial": Raw statistical outputs: Liver function test_normal data ANCOVA test.rtf

Univariate Analysis of Variance


Notes	
Output Created	11-MAY-2026 12:23:16	
Comments		
Input	Active Dataset	DataSet0	
	Filter	<none>	
	Weight	<none>	
	Split File	<none>	
	N of Rows in Working Data File	64	
Missing Value Handling	Definition of Missing	User-defined missing values are treated as missing.	
	Cases Used	Statistics are based on all cases with valid data for all variables in the model.	
Syntax	UNIANOVA W12_Bilrub_Dire BY Groups WITH B_Bilrub_Dire
  /METHOD=SSTYPE(3)
  /INTERCEPT=INCLUDE
  /EMMEANS=TABLES(Groups) WITH(B_Bilrub_Dire=MEAN) COMPARE ADJ(BONFERRONI)
  /PRINT ETASQ DESCRIPTIVE
  /CRITERIA=ALPHA(.05)
  /DESIGN=B_Bilrub_Dire Groups.	
Resources	Processor Time	00:00:00.00	
	Elapsed Time	00:00:00.01	


Between-Subjects Factors	
	N	
Groups	1	32	
	2	32	


Descriptive Statistics	
Dependent Variable:   W12_Bilrub_Dire  	
Groups	Mean	Std. Deviation	N	
1	.2591	.09223	32	
2	.2422	.07508	32	
Total	.2506	.08385	64	


Tests of Between-Subjects Effects	
Dependent Variable:   W12_Bilrub_Dire  	
Source	Type III Sum of Squares	df	Mean Square	F	Sig.	Partial Eta Squared	
Corrected Model	.039^a^	2	.020	2.971	.059	.089	
Intercept	.272	1	.272	41.093	<.001	.403	
B_Bilrub_Dire	.035	1	.035	5.253	.025	.079	
Groups	.010	1	.010	1.562	.216	.025	
Error	.404	61	.007				
Total	4.463	64					
Corrected Total	.443	63					

a. R Squared = .089 (Adjusted R Squared = .059)	


Estimated Marginal Means


Groups


Estimates	
Dependent Variable:   W12_Bilrub_Dire  	
Groups	Mean	Std. Error	95% Confidence Interval	
			Lower Bound	Upper Bound	
1	.264^a^	.015	.235	.293	
2	.238^a^	.015	.209	.267	

a. Covariates appearing in the model are evaluated at the following values: B_Bilrub_Dire = .2225.	


Pairwise Comparisons	
Dependent Variable:   W12_Bilrub_Dire  	
(I) Groups	(J) Groups	Mean Difference (I-J)	Std. Error	Sig.^a^	95% Confidence Interval for Difference^a^	
					Lower Bound	Upper Bound	
1	2	.026	.021	.216	-.016	.067	
2	1	-.026	.021	.216	-.067	.016	

Based on estimated marginal means	
a. Adjustment for multiple comparisons: Bonferroni.	


Univariate Tests	
Dependent Variable:   W12_Bilrub_Dire  	
	Sum of Squares	df	Mean Square	F	Sig.	Partial Eta Squared	
Contrast	.010	1	.010	1.562	.216	.025	
Error	.404	61	.007				

The F tests the effect of Groups. This test is based on the linearly independent pairwise comparisons among the estimated marginal means.	


Univariate Analysis of Variance


Notes	
Output Created	11-MAY-2026 12:23:33	
Comments		
Input	Active Dataset	DataSet0	
	Filter	<none>	
	Weight	<none>	
	Split File	<none>	
	N of Rows in Working Data File	64	
Missing Value Handling	Definition of Missing	User-defined missing values are treated as missing.	
	Cases Used	Statistics are based on all cases with valid data for all variables in the model.	
Syntax	UNIANOVA W12_SGOT BY Groups WITH B_SGOT
  /METHOD=SSTYPE(3)
  /INTERCEPT=INCLUDE
  /EMMEANS=TABLES(Groups) WITH(B_SGOT=MEAN) COMPARE ADJ(BONFERRONI)
  /PRINT ETASQ DESCRIPTIVE
  /CRITERIA=ALPHA(.05)
  /DESIGN=B_SGOT Groups.	
Resources	Processor Time	00:00:00.02	
	Elapsed Time	00:00:00.02	


Between-Subjects Factors	
	N	
Groups	1	32	
	2	32	


Descriptive Statistics	
Dependent Variable:   W12_SGOT  	
Groups	Mean	Std. Deviation	N	
1	28.2934	9.43844	32	
2	26.0831	10.41010	32	
Total	27.1883	9.91972	64	


Tests of Between-Subjects Effects	
Dependent Variable:   W12_SGOT  	
Source	Type III Sum of Squares	df	Mean Square	F	Sig.	Partial Eta Squared	
Corrected Model	253.959^a^	2	126.979	1.303	.279	.041	
Intercept	3436.810	1	3436.810	35.262	<.001	.366	
B_SGOT	175.791	1	175.791	1.804	.184	.029	
Groups	79.536	1	79.536	.816	.370	.013	
Error	5945.289	61	97.464				
Total	53508.216	64					
Corrected Total	6199.248	63					

a. R Squared = .041 (Adjusted R Squared = .010)	


Estimated Marginal Means


Groups


Estimates	
Dependent Variable:   W12_SGOT  	
Groups	Mean	Std. Error	95% Confidence Interval	
			Lower Bound	Upper Bound	
1	28.303^a^	1.745	24.813	31.793	
2	26.073^a^	1.745	22.584	29.563	

a. Covariates appearing in the model are evaluated at the following values: B_SGOT = 28.6878.	


Pairwise Comparisons	
Dependent Variable:   W12_SGOT  	
(I) Groups	(J) Groups	Mean Difference (I-J)	Std. Error	Sig.^a^	95% Confidence Interval for Difference^a^	
					Lower Bound	Upper Bound	
1	2	2.230	2.468	.370	-2.706	7.165	
2	1	-2.230	2.468	.370	-7.165	2.706	

Based on estimated marginal means	
a. Adjustment for multiple comparisons: Bonferroni.	


Univariate Tests	
Dependent Variable:   W12_SGOT  	
	Sum of Squares	df	Mean Square	F	Sig.	Partial Eta Squared	
Contrast	79.536	1	79.536	.816	.370	.013	
Error	5945.289	61	97.464				

The F tests the effect of Groups. This test is based on the linearly independent pairwise comparisons among the estimated marginal means.	


Univariate Analysis of Variance


Notes	
Output Created	11-MAY-2026 12:23:45	
Comments		
Input	Active Dataset	DataSet0	
	Filter	<none>	
	Weight	<none>	
	Split File	<none>	
	N of Rows in Working Data File	64	
Missing Value Handling	Definition of Missing	User-defined missing values are treated as missing.	
	Cases Used	Statistics are based on all cases with valid data for all variables in the model.	
Syntax	UNIANOVA W12_Alkalin_Phosp BY Groups WITH B_Alkalin_Phosp
  /METHOD=SSTYPE(3)
  /INTERCEPT=INCLUDE
  /EMMEANS=TABLES(Groups) WITH(B_Alkalin_Phosp=MEAN) COMPARE ADJ(BONFERRONI)
  /PRINT ETASQ DESCRIPTIVE
  /CRITERIA=ALPHA(.05)
  /DESIGN=B_Alkalin_Phosp Groups.	
Resources	Processor Time	00:00:00.02	
	Elapsed Time	00:00:00.01	


Between-Subjects Factors	
	N	
Groups	1	32	
	2	32	


Descriptive Statistics	
Dependent Variable:   W12_Alkalin_Phosp  	
Groups	Mean	Std. Deviation	N	
1	105.7585	23.95415	32	
2	102.0153	21.54462	32	
Total	103.8869	22.67833	64	


Tests of Between-Subjects Effects	
Dependent Variable:   W12_Alkalin_Phosp  	
Source	Type III Sum of Squares	df	Mean Square	F	Sig.	Partial Eta Squared	
Corrected Model	703.805^a^	2	351.903	.677	.512	.022	
Intercept	19677.248	1	19677.248	37.868	<.001	.383	
B_Alkalin_Phosp	479.626	1	479.626	.923	.340	.015	
Groups	221.859	1	221.859	.427	.516	.007	
Error	31697.505	61	519.631				
Total	723120.417	64					
Corrected Total	32401.310	63					

a. R Squared = .022 (Adjusted R Squared = -.010)	


Estimated Marginal Means


Groups


Estimates	
Dependent Variable:   W12_Alkalin_Phosp  	
Groups	Mean	Std. Error	95% Confidence Interval	
			Lower Bound	Upper Bound	
1	105.749^a^	4.030	97.691	113.807	
2	102.025^a^	4.030	93.967	110.083	

a. Covariates appearing in the model are evaluated at the following values: B_Alkalin_Phosp = 107.3480.	


Pairwise Comparisons	
Dependent Variable:   W12_Alkalin_Phosp  	
(I) Groups	(J) Groups	Mean Difference (I-J)	Std. Error	Sig.^a^	95% Confidence Interval for Difference^a^	
					Lower Bound	Upper Bound	
1	2	3.724	5.699	.516	-7.672	15.119	
2	1	-3.724	5.699	.516	-15.119	7.672	

Based on estimated marginal means	
a. Adjustment for multiple comparisons: Bonferroni.	


Univariate Tests	
Dependent Variable:   W12_Alkalin_Phosp  	
	Sum of Squares	df	Mean Square	F	Sig.	Partial Eta Squared	
Contrast	221.859	1	221.859	.427	.516	.007	
Error	31697.505	61	519.631				

The F tests the effect of Groups. This test is based on the linearly independent pairwise comparisons among the estimated marginal means.	


Univariate Analysis of Variance


Notes	
Output Created	11-MAY-2026 12:23:59	
Comments		
Input	Active Dataset	DataSet0	
	Filter	<none>	
	Weight	<none>	
	Split File	<none>	
	N of Rows in Working Data File	64	
Missing Value Handling	Definition of Missing	User-defined missing values are treated as missing.	
	Cases Used	Statistics are based on all cases with valid data for all variables in the model.	
Syntax	UNIANOVA W12_Ser_Globu BY Groups WITH B_Ser_Globu
  /METHOD=SSTYPE(3)
  /INTERCEPT=INCLUDE
  /EMMEANS=TABLES(Groups) WITH(B_Ser_Globu=MEAN) COMPARE ADJ(BONFERRONI)
  /PRINT ETASQ DESCRIPTIVE
  /CRITERIA=ALPHA(.05)
  /DESIGN=B_Ser_Globu Groups.	
Resources	Processor Time	00:00:00.00	
	Elapsed Time	00:00:00.00	


Between-Subjects Factors	
	N	
Groups	1	32	
	2	32	


Descriptive Statistics	
Dependent Variable:   W12_Ser_Globu  	
Groups	Mean	Std. Deviation	N	
1	2.9638	.55240	32	
2	3.0763	.52493	32	
Total	3.0200	.53754	64	


Tests of Between-Subjects Effects	
Dependent Variable:   W12_Ser_Globu  	
Source	Type III Sum of Squares	df	Mean Square	F	Sig.	Partial Eta Squared	
Corrected Model	.247^a^	2	.123	.419	.660	.014	
Intercept	10.493	1	10.493	35.645	<.001	.369	
B_Ser_Globu	.044	1	.044	.150	.700	.002	
Groups	.205	1	.205	.695	.408	.011	
Error	17.957	61	.294				
Total	601.910	64					
Corrected Total	18.204	63					

a. R Squared = .014 (Adjusted R Squared = -.019)	


Estimated Marginal Means


Groups


Estimates	
Dependent Variable:   W12_Ser_Globu  	
Groups	Mean	Std. Error	95% Confidence Interval	
			Lower Bound	Upper Bound	
1	2.963^a^	.096	2.772	3.155	
2	3.077^a^	.096	2.885	3.268	

a. Covariates appearing in the model are evaluated at the following values: B_Ser_Globu = 2.8831.	


Pairwise Comparisons	
Dependent Variable:   W12_Ser_Globu  	
(I) Groups	(J) Groups	Mean Difference (I-J)	Std. Error	Sig.^a^	95% Confidence Interval for Difference^a^	
					Lower Bound	Upper Bound	
1	2	-.113	.136	.408	-.384	.158	
2	1	.113	.136	.408	-.158	.384	

Based on estimated marginal means	
a. Adjustment for multiple comparisons: Bonferroni.	


Univariate Tests	
Dependent Variable:   W12_Ser_Globu  	
	Sum of Squares	df	Mean Square	F	Sig.	Partial Eta Squared	
Contrast	.205	1	.205	.695	.408	.011	
Error	17.957	61	.294				

The F tests the effect of Groups. This test is based on the linearly independent pairwise comparisons among the estimated marginal means.	


Univariate Analysis of Variance


Notes	
Output Created	11-MAY-2026 12:24:19	
Comments		
Input	Active Dataset	DataSet0	
	Filter	<none>	
	Weight	<none>	
	Split File	<none>	
	N of Rows in Working Data File	64	
Missing Value Handling	Definition of Missing	User-defined missing values are treated as missing.	
	Cases Used	Statistics are based on all cases with valid data for all variables in the model.	
Syntax	UNIANOVA W12_A_G_Ratio BY Groups WITH Bas_A_G_Ratio
  /METHOD=SSTYPE(3)
  /INTERCEPT=INCLUDE
  /EMMEANS=TABLES(Groups) WITH(Bas_A_G_Ratio=MEAN) COMPARE ADJ(BONFERRONI)
  /PRINT ETASQ DESCRIPTIVE
  /CRITERIA=ALPHA(.05)
  /DESIGN=Bas_A_G_Ratio Groups.	
Resources	Processor Time	00:00:00.02	
	Elapsed Time	00:00:00.00	


Between-Subjects Factors	
	N	
Groups	1	32	
	2	32	


Descriptive Statistics	
Dependent Variable:   W12_A_G_Ratio  	
Groups	Mean	Std. Deviation	N	
1	1.3672	.36414	32	
2	1.3259	.50623	32	
Total	1.3466	.43793	64	


Tests of Between-Subjects Effects	
Dependent Variable:   W12_A_G_Ratio  	
Source	Type III Sum of Squares	df	Mean Square	F	Sig.	Partial Eta Squared	
Corrected Model	.428^a^	2	.214	1.121	.333	.035	
Intercept	7.289	1	7.289	38.154	<.001	.385	
Bas_A_G_Ratio	.401	1	.401	2.099	.153	.033	
Groups	.016	1	.016	.086	.770	.001	
Error	11.654	61	.191				
Total	128.129	64					
Corrected Total	12.082	63					

a. R Squared = .035 (Adjusted R Squared = .004)	


Estimated Marginal Means


Groups


Estimates	
Dependent Variable:   W12_A_G_Ratio  	
Groups	Mean	Std. Error	95% Confidence Interval	
			Lower Bound	Upper Bound	
1	1.363^a^	.077	1.208	1.517	
2	1.330^a^	.077	1.176	1.485	

a. Covariates appearing in the model are evaluated at the following values: Bas_A_G_Ratio = 1.4350.	


Pairwise Comparisons	
Dependent Variable:   W12_A_G_Ratio  	
(I) Groups	(J) Groups	Mean Difference (I-J)	Std. Error	Sig.^a^	95% Confidence Interval for Difference^a^	
					Lower Bound	Upper Bound	
1	2	.032	.109	.770	-.187	.251	
2	1	-.032	.109	.770	-.251	.187	

Based on estimated marginal means	
a. Adjustment for multiple comparisons: Bonferroni.	


Univariate Tests	
Dependent Variable:   W12_A_G_Ratio  	
	Sum of Squares	df	Mean Square	F	Sig.	Partial Eta Squared	
Contrast	.016	1	.016	.086	.770	.001	
Error	11.654	61	.191				

The F tests the effect of Groups. This test is based on the linearly independent pairwise comparisons among the estimated marginal means.	


T-Test


Notes	
Output Created	11-MAY-2026 12:25:38	
Comments		
Input	Active Dataset	DataSet0	
	Filter	<none>	
	Weight	<none>	
	Split File	<none>	
	N of Rows in Working Data File	64	
Missing Value Handling	Definition of Missing	User defined missing values are treated as missing.	
	Cases Used	Statistics for each analysis are based on the cases with no missing or out-of-range data for any variable in the analysis.	
Syntax	T-TEST PAIRS=B_Bilrub_Dire_1 B_SGOT_1 B_Alkalin_Phosp_1 B_Ser_Globu_1 Bas_A_G_Ratio_1
    B_Bilrub_Dire_2 B_SGOT_2 B_Alkalin_Phosp_2 B_Ser_Globu_2 Bas_A_G_Ratio_2 WITH W12_Bilrub_Dire_1
    W12_SGOT_1 W12_Alkalin_Phosp_1 W12_Ser_Globu_1 W12_A_G_Ratio_1 W12_Bilrub_Dire_2 W12_SGOT_2
    W12_Alkalin_Phosp_2 W12_Ser_Globu_2 W12_A_G_Ratio_2 (PAIRED)
  /ES DISPLAY(TRUE) STANDARDIZER(SD)
  /CRITERIA=CI(.9500)
  /MISSING=ANALYSIS.	
Resources	Processor Time	00:00:00.02	
	Elapsed Time	00:00:00.00	


Paired Samples Statistics	
	Mean	N	Std. Deviation	Std. Error Mean	
Pair 1	B_Bilrub_Dire_1	.2066	32	.08827	.01560	
	W12_Bilrub_Dire_1	.2591	32	.09223	.01630	
Pair 2	B_SGOT_1	28.6300	32	11.25236	1.98916	
	W12_SGOT_1	28.2934	32	9.43844	1.66850	
Pair 3	B_Alkalin_Phosp_1	107.4234	32	22.29486	3.94121	
	W12_Alkalin_Phosp_1	105.7585	32	23.95415	4.23454	
Pair 4	B_Ser_Globu_1	2.8791	32	.31490	.05567	
	W12_Ser_Globu_1	2.9637	32	.55240	.09765	
Pair 5	Bas_A_G_Ratio_1	1.4188	32	.25378	.04486	
	W12_A_G_Ratio_1	1.3672	32	.36414	.06437	
Pair 6	B_Bilrub_Dire_2	.2384	32	.07891	.01395	
	W12_Bilrub_Dire_2	.2422	32	.07508	.01327	
Pair 7	B_SGOT_2	28.7456	32	8.77514	1.55124	
	W12_SGOT_2	26.0831	32	10.41010	1.84026	
Pair 8	B_Alkalin_Phosp_2	107.2725	32	20.96769	3.70660	
	W12_Alkalin_Phosp_2	102.0153	32	21.54462	3.80859	
Pair 9	B_Ser_Globu_2	2.8872	32	.41858	.07399	
	W12_Ser_Globu_2	3.0763	32	.52493	.09280	
Pair 10	Bas_A_G_Ratio_2	1.4513	32	.31622	.05590	
	W12_A_G_Ratio_2	1.3259	32	.50623	.08949	


Paired Samples Correlations	
	N	Correlation	Significance	
			One-Sided p	Two-Sided p	
Pair 1	B_Bilrub_Dire_1 & W12_Bilrub_Dire_1	32	.416	.009	.018	
Pair 2	B_SGOT_1 & W12_SGOT_1	32	.303	.046	.092	
Pair 3	B_Alkalin_Phosp_1 & W12_Alkalin_Phosp_1	32	.108	.278	.555	
Pair 4	B_Ser_Globu_1 & W12_Ser_Globu_1	32	.047	.398	.797	
Pair 5	Bas_A_G_Ratio_1 & W12_A_G_Ratio_1	32	-.089	.315	.629	
Pair 6	B_Bilrub_Dire_2 & W12_Bilrub_Dire_2	32	.098	.297	.593	
Pair 7	B_SGOT_2 & W12_SGOT_2	32	.020	.458	.915	
Pair 8	B_Alkalin_Phosp_2 & W12_Alkalin_Phosp_2	32	.138	.225	.450	
Pair 9	B_Ser_Globu_2 & W12_Ser_Globu_2	32	-.128	.243	.487	
Pair 10	Bas_A_G_Ratio_2 & W12_A_G_Ratio_2	32	-.237	.096	.192	


Paired Samples Test	
	Paired Differences	
	Mean	Std. Deviation	Std. Error Mean	95% Confidence Interval of the Difference	
				Lower	
Pair 1	B_Bilrub_Dire_1 - W12_Bilrub_Dire_1	-.05250	.09762	.01726	-.08769	
Pair 2	B_SGOT_1 - W12_SGOT_1	.33656	12.30118	2.17456	-4.09848	
Pair 3	B_Alkalin_Phosp_1 - W12_Alkalin_Phosp_1	1.66497	30.90546	5.46337	-9.47764	
Pair 4	B_Ser_Globu_1 - W12_Ser_Globu_1	-.08469	.62275	.11009	-.30921	
Pair 5	Bas_A_G_Ratio_1 - W12_A_G_Ratio_1	.05156	.46196	.08166	-.11499	
Pair 6	B_Bilrub_Dire_2 - W12_Bilrub_Dire_2	-.00375	.10345	.01829	-.04105	
Pair 7	B_SGOT_2 - W12_SGOT_2	2.66250	13.48308	2.38349	-2.19867	
Pair 8	B_Alkalin_Phosp_2 - W12_Alkalin_Phosp_2	5.25719	27.90626	4.93318	-4.80409	
Pair 9	B_Ser_Globu_2 - W12_Ser_Globu_2	-.18906	.71190	.12585	-.44573	
Pair 10	Bas_A_G_Ratio_2 - W12_A_G_Ratio_2	.12531	.65733	.11620	-.11168	

Paired Samples Test	
	Paired Differences	t	df	Significance	
	95% Confidence Interval of the Difference			One-Sided p	
	Upper				
Pair 1	B_Bilrub_Dire_1 - W12_Bilrub_Dire_1	-.01731	-3.042	31	.002	
Pair 2	B_SGOT_1 - W12_SGOT_1	4.77161	.155	31	.439	
Pair 3	B_Alkalin_Phosp_1 - W12_Alkalin_Phosp_1	12.80758	.305	31	.381	
Pair 4	B_Ser_Globu_1 - W12_Ser_Globu_1	.13984	-.769	31	.224	
Pair 5	Bas_A_G_Ratio_1 - W12_A_G_Ratio_1	.21812	.631	31	.266	
Pair 6	B_Bilrub_Dire_2 - W12_Bilrub_Dire_2	.03355	-.205	31	.419	
Pair 7	B_SGOT_2 - W12_SGOT_2	7.52367	1.117	31	.136	
Pair 8	B_Alkalin_Phosp_2 - W12_Alkalin_Phosp_2	15.31847	1.066	31	.147	
Pair 9	B_Ser_Globu_2 - W12_Ser_Globu_2	.06760	-1.502	31	.072	
Pair 10	Bas_A_G_Ratio_2 - W12_A_G_Ratio_2	.36231	1.078	31	.145	


Paired Samples Test	
	Significance	
	Two-Sided p	
		
Pair 1	B_Bilrub_Dire_1 - W12_Bilrub_Dire_1	.005	
Pair 2	B_SGOT_1 - W12_SGOT_1	.878	
Pair 3	B_Alkalin_Phosp_1 - W12_Alkalin_Phosp_1	.763	
Pair 4	B_Ser_Globu_1 - W12_Ser_Globu_1	.448	
Pair 5	Bas_A_G_Ratio_1 - W12_A_G_Ratio_1	.532	
Pair 6	B_Bilrub_Dire_2 - W12_Bilrub_Dire_2	.839	
Pair 7	B_SGOT_2 - W12_SGOT_2	.273	
Pair 8	B_Alkalin_Phosp_2 - W12_Alkalin_Phosp_2	.295	
Pair 9	B_Ser_Globu_2 - W12_Ser_Globu_2	.143	
Pair 10	Bas_A_G_Ratio_2 - W12_A_G_Ratio_2	.289	


Paired Samples Effect Sizes	
	Standardizer^a^	Point Estimate	95% Confidence Interval	
			Lower	
Pair 1	B_Bilrub_Dire_1 - W12_Bilrub_Dire_1	Cohen's d	.09762	-.538	-.905	
		Hedges' correction	.10006	-.525	-.883	
Pair 2	B_SGOT_1 - W12_SGOT_1	Cohen's d	12.30118	.027	-.319	
		Hedges' correction	12.60914	.027	-.312	
Pair 3	B_Alkalin_Phosp_1 - W12_Alkalin_Phosp_1	Cohen's d	30.90546	.054	-.293	
		Hedges' correction	31.67918	.053	-.286	
Pair 4	B_Ser_Globu_1 - W12_Ser_Globu_1	Cohen's d	.62275	-.136	-.483	
		Hedges' correction	.63834	-.133	-.471	
Pair 5	Bas_A_G_Ratio_1 - W12_A_G_Ratio_1	Cohen's d	.46196	.112	-.237	
		Hedges' correction	.47353	.109	-.231	
Pair 6	B_Bilrub_Dire_2 - W12_Bilrub_Dire_2	Cohen's d	.10345	-.036	-.383	
		Hedges' correction	.10604	-.035	-.373	
Pair 7	B_SGOT_2 - W12_SGOT_2	Cohen's d	13.48308	.197	-.154	
		Hedges' correction	13.82063	.193	-.150	
Pair 8	B_Alkalin_Phosp_2 - W12_Alkalin_Phosp_2	Cohen's d	27.90626	.188	-.163	
		Hedges' correction	28.60490	.184	-.159	
Pair 9	B_Ser_Globu_2 - W12_Ser_Globu_2	Cohen's d	.71190	-.266	-.616	
		Hedges' correction	.72972	-.259	-.601	
Pair 10	Bas_A_G_Ratio_2 - W12_A_G_Ratio_2	Cohen's d	.65733	.191	-.161	
		Hedges' correction	.67379	.186	-.157	

Paired Samples Effect Sizes	
	95% Confidence Interval^a^	
	Upper	
Pair 1	B_Bilrub_Dire_1 - W12_Bilrub_Dire_1	Cohen's d	-.163	
		Hedges' correction	-.159	
Pair 2	B_SGOT_1 - W12_SGOT_1	Cohen's d	.374	
		Hedges' correction	.365	
Pair 3	B_Alkalin_Phosp_1 - W12_Alkalin_Phosp_1	Cohen's d	.400	
		Hedges' correction	.390	
Pair 4	B_Ser_Globu_1 - W12_Ser_Globu_1	Cohen's d	.213	
		Hedges' correction	.208	
Pair 5	Bas_A_G_Ratio_1 - W12_A_G_Ratio_1	Cohen's d	.458	
		Hedges' correction	.447	
Pair 6	B_Bilrub_Dire_2 - W12_Bilrub_Dire_2	Cohen's d	.311	
		Hedges' correction	.303	
Pair 7	B_SGOT_2 - W12_SGOT_2	Cohen's d	.546	
		Hedges' correction	.532	
Pair 8	B_Alkalin_Phosp_2 - W12_Alkalin_Phosp_2	Cohen's d	.537	
		Hedges' correction	.523	
Pair 9	B_Ser_Globu_2 - W12_Ser_Globu_2	Cohen's d	.089	
		Hedges' correction	.087	
Pair 10	Bas_A_G_Ratio_2 - W12_A_G_Ratio_2	Cohen's d	.539	
		Hedges' correction	.526	

a. The denominator used in estimating the effect sizes. 
Cohen's d uses the sample standard deviation of the mean difference. 
Hedges' correction uses the sample standard deviation of the mean difference, plus a correction factor.	
