## Supplementary material for "Effects of a standardised *Amaranthus tricolor*–*Spinacia oleracea* extract on exercise performance, muscle strength, endurance and recovery in physically active adults: a randomised, double-blind, placebo-controlled trial": Raw statistical outputs: Mus strength 1 RM Bench Press - non parametric test.rtf

NPar Tests


Notes	
Output Created	11-JUN-2026 10:46:59	
Comments		
Input	Active Dataset	DataSet0	
	Filter	<none>	
	Weight	<none>	
	Split File	<none>	
	N of Rows in Working Data File	64	
Missing Value Handling	Definition of Missing	User-defined missing values are treated as missing.	
	Cases Used	Statistics for each test are based on all cases with valid data for the variable(s) used in that test.	
Syntax	NPAR TESTS
  /WILCOXON=Baseline_1 Baseline_1 Baseline_2 Baseline_2 WITH Day_45_1 Day_90_1 Day_45_2 Day_90_2
    (PAIRED)
  /STATISTICS DESCRIPTIVES QUARTILES
  /MISSING ANALYSIS.	
Resources	Processor Time	00:00:00.00	
	Elapsed Time	00:00:00.00	
	Number of Cases Allowed^a^	285975	

a. Based on availability of workspace memory.	


Descriptive Statistics	
	N	Mean	Std. Deviation	Minimum	Maximum	Percentiles	
						25th	50th (Median)	
Baseline_1	32	71.1174	8.26227	57.18	87.00	64.5913	70.4116	
Baseline_2	32	70.6029	7.91117	44.00	84.71	65.6501	70.4151	
Day_45_1	32	74.3603	7.88023	62.47	90.01	66.8230	73.0481	
Day_90_1	32	77.6677	6.68143	66.38	91.00	72.2885	78.7579	
Day_45_2	32	72.5546	6.36198	63.28	86.19	67.6413	70.3688	
Day_90_2	32	72.9010	6.64779	63.28	84.38	66.8230	70.3527	

Descriptive Statistics	
	Percentiles	
	75th	
Baseline_1	77.2978	
Baseline_2	77.0656	
Day_45_1	81.5514	
Day_90_1	82.9770	
Day_45_2	79.3694	
Day_90_2	80.4604	


Wilcoxon Signed Ranks Test


Ranks	
	N	Mean Rank	Sum of Ranks	
Day_45_1 - Baseline_1	Negative Ranks	7^a^	16.71	117.00	
	Positive Ranks	23^b^	15.13	348.00	
	Ties	2^c^			
	Total	32			
Day_90_1 - Baseline_1	Negative Ranks	4^d^	16.50	66.00	
	Positive Ranks	28^e^	16.50	462.00	
	Ties	0^f^			
	Total	32			
Day_45_2 - Baseline_2	Negative Ranks	7^g^	23.93	167.50	
	Positive Ranks	25^h^	14.42	360.50	
	Ties	0^i^			
	Total	32			
Day_90_2 - Baseline_2	Negative Ranks	11^j^	17.82	196.00	
	Positive Ranks	21^k^	15.81	332.00	
	Ties	0^l^			
	Total	32			

a. Day_45_1 < Baseline_1	
b. Day_45_1 > Baseline_1	
c. Day_45_1 = Baseline_1	
d. Day_90_1 < Baseline_1	
e. Day_90_1 > Baseline_1	
f. Day_90_1 = Baseline_1	
g. Day_45_2 < Baseline_2	
h. Day_45_2 > Baseline_2	
i. Day_45_2 = Baseline_2	
j. Day_90_2 < Baseline_2	
k. Day_90_2 > Baseline_2	
l. Day_90_2 = Baseline_2	


Test Statistics^a^	
	Day_45_1 - Baseline_1	Day_90_1 - Baseline_1	Day_45_2 - Baseline_2	Day_90_2 - Baseline_2	
Z	-2.376^b^	-3.702^b^	-1.805^b^	-1.272^b^	
Asymp. Sig. (2-tailed)	.018	<.001	.071	.204	

a. Wilcoxon Signed Ranks Test	
b. Based on negative ranks.	


NPar Tests


Notes	
Output Created	11-JUN-2026 10:47:41	
Comments		
Input	Active Dataset	DataSet0	
	Filter	<none>	
	Weight	<none>	
	Split File	<none>	
	N of Rows in Working Data File	64	
Missing Value Handling	Definition of Missing	User-defined missing values are treated as missing.	
	Cases Used	Statistics for each test are based on all cases with valid data for the variable(s) used in that test.	
Syntax	NPAR TESTS
  /M-W= Diff_45 Diff_90 Baseline BY Groups(1 2)
  /STATISTICS=DESCRIPTIVES QUARTILES
  /MISSING ANALYSIS.	
Resources	Processor Time	00:00:00.00	
	Elapsed Time	00:00:00.00	
	Number of Cases Allowed^a^	349525	

a. Based on availability of workspace memory.	


Descriptive Statistics	
	N	Mean	Std. Deviation	Minimum	Maximum	Percentiles	
						25th	50th (Median)	
Diff_45	64	-2.5973	7.22695	-21.46	12.00	-6.3996	-2.0714	
Diff_90	64	-4.4242	8.49817	-32.00	19.45	-9.0241	-4.0203	
Baseline	64	70.8602	8.02836	44.00	87.00	64.5913	70.4116	
Groups	64	1.5000	.50395	1.00	2.00	1.0000	1.5000	

Descriptive Statistics	
	Percentiles	
	75th	
Diff_45	-.2170	
Diff_90	-.9965	
Baseline	77.2978	
Groups	2.0000	


Mann-Whitney Test


Ranks	
	Groups	N	Mean Rank	Sum of Ranks	
Diff_45	1.00	32	31.13	996.00	
	2.00	32	33.88	1084.00	
	Total	64			
Diff_90	1.00	32	26.20	838.50	
	2.00	32	38.80	1241.50	
	Total	64			
Baseline	1.00	32	32.22	1031.00	
	2.00	32	32.78	1049.00	
	Total	64			


Test Statistics^a^	
	Diff_45	Diff_90	Baseline	
Mann-Whitney U	468.000	310.500	503.000	
Wilcoxon W	996.000	838.500	1031.000	
Z	-.591	-2.706	-.121	
Asymp. Sig. (2-tailed)	.555	.007	.904	

a. Grouping Variable: Groups	
