## Supplementary material for "Effects of a standardised *Amaranthus tricolor*–*Spinacia oleracea* extract on exercise performance, muscle strength, endurance and recovery in physically active adults: a randomised, double-blind, placebo-controlled trial": Raw statistical outputs: Mus strength 1 RM Leg Press- Non Para tests.rtf

NPar Tests


Notes	
Output Created	11-JUN-2026 10:49:08	
Comments		
Input	Active Dataset	DataSet0	
	Filter	<none>	
	Weight	<none>	
	Split File	<none>	
	N of Rows in Working Data File	64	
Missing Value Handling	Definition of Missing	User-defined missing values are treated as missing.	
	Cases Used	Statistics for each test are based on all cases with valid data for the variable(s) used in that test.	
Syntax	NPAR TESTS
  /WILCOXON=Baseline_1 Baseline_1 Baseline_2 Baseline_2 WITH Day_45_1 Day_90_1 Day_45_2 Day_90_2
    (PAIRED)
  /STATISTICS DESCRIPTIVES QUARTILES
  /MISSING ANALYSIS.	
Resources	Processor Time	00:00:00.00	
	Elapsed Time	00:00:00.00	
	Number of Cases Allowed^a^	285975	

a. Based on availability of workspace memory.	


Descriptive Statistics	
	N	Mean	Std. Deviation	Minimum	Maximum	Percentiles	
						25th	50th (Median)	
Baseline_1	32	90.0003	7.29884	75.18	103.77	85.2393	90.0042	
Baseline_2	32	88.7903	8.84251	56.00	102.71	82.8568	90.0042	
Day_45_1	32	93.5241	6.83837	80.73	106.92	88.5139	94.3705	
Day_90_1	32	97.1572	6.23256	81.01	109.14	94.7907	97.8848	
Day_45_2	32	91.4634	8.03062	74.12	105.83	85.0794	92.4280	
Day_90_2	32	92.5878	7.55536	77.46	109.14	86.1881	94.9160	

Descriptive Statistics	
	Percentiles	
	75th	
Baseline_1	95.2986	
Baseline_2	93.9750	
Day_45_1	98.1890	
Day_90_1	101.2601	
Day_45_2	96.0070	
Day_90_2	96.7517	


Wilcoxon Signed Ranks Test


Ranks	
	N	Mean Rank	Sum of Ranks	
Day_45_1 - Baseline_1	Negative Ranks	4^a^	17.50	70.00	
	Positive Ranks	27^b^	15.78	426.00	
	Ties	1^c^			
	Total	32			
Day_90_1 - Baseline_1	Negative Ranks	4^d^	4.25	17.00	
	Positive Ranks	28^e^	18.25	511.00	
	Ties	0^f^			
	Total	32			
Day_45_2 - Baseline_2	Negative Ranks	6^g^	10.83	65.00	
	Positive Ranks	23^h^	16.09	370.00	
	Ties	3^i^			
	Total	32			
Day_90_2 - Baseline_2	Negative Ranks	5^j^	9.40	47.00	
	Positive Ranks	26^k^	17.27	449.00	
	Ties	1^l^			
	Total	32			

a. Day_45_1 < Baseline_1	
b. Day_45_1 > Baseline_1	
c. Day_45_1 = Baseline_1	
d. Day_90_1 < Baseline_1	
e. Day_90_1 > Baseline_1	
f. Day_90_1 = Baseline_1	
g. Day_45_2 < Baseline_2	
h. Day_45_2 > Baseline_2	
i. Day_45_2 = Baseline_2	
j. Day_90_2 < Baseline_2	
k. Day_90_2 > Baseline_2	
l. Day_90_2 = Baseline_2	


Test Statistics^a^	
	Day_45_1 - Baseline_1	Day_90_1 - Baseline_1	Day_45_2 - Baseline_2	Day_90_2 - Baseline_2	
Z	-3.489^b^	-4.619^b^	-3.299^b^	-3.939^b^	
Asymp. Sig. (2-tailed)	<.001	<.001	<.001	<.001	

a. Wilcoxon Signed Ranks Test	
b. Based on negative ranks.	


NPar Tests


Notes	
Output Created	11-JUN-2026 10:49:18	
Comments		
Input	Active Dataset	DataSet0	
	Filter	<none>	
	Weight	<none>	
	Split File	<none>	
	N of Rows in Working Data File	64	
Missing Value Handling	Definition of Missing	User-defined missing values are treated as missing.	
	Cases Used	Statistics for each test are based on all cases with valid data for the variable(s) used in that test.	
Syntax	NPAR TESTS
  /M-W= Diff_45 Diff_90 Baseline BY Groups(1 2)
  /STATISTICS=DESCRIPTIVES QUARTILES
  /MISSING ANALYSIS.	
Resources	Processor Time	00:00:00.02	
	Elapsed Time	00:00:00.01	
	Number of Cases Allowed^a^	349525	

a. Based on availability of workspace memory.	


Descriptive Statistics	
	N	Mean	Std. Deviation	Minimum	Maximum	Percentiles	
						25th	50th (Median)	
Diff_45	64	-3.0985	4.70002	-18.12	9.14	-4.7833	-2.8261	
Diff_90	64	-5.4773	5.39841	-21.46	6.86	-7.9848	-5.9786	
Baseline	64	89.3953	8.06598	56.00	103.77	83.9157	90.0042	
Groups	64	1.5000	.50395	1.00	2.00	1.0000	1.5000	

Descriptive Statistics	
	Percentiles	
	75th	
Diff_45	-1.0589	
Diff_90	-2.8209	
Baseline	95.0339	
Groups	2.0000	


Mann-Whitney Test


Ranks	
	Groups	N	Mean Rank	Sum of Ranks	
Diff_45	1.00	32	28.91	925.00	
	2.00	32	36.09	1155.00	
	Total	64			
Diff_90	1.00	32	23.89	764.50	
	2.00	32	41.11	1315.50	
	Total	64			
Baseline	1.00	32	33.13	1060.00	
	2.00	32	31.88	1020.00	
	Total	64			


Test Statistics^a^	
	Diff_45	Diff_90	Baseline	
Mann-Whitney U	397.000	236.500	492.000	
Wilcoxon W	925.000	764.500	1020.000	
Z	-1.545	-3.700	-.269	
Asymp. Sig. (2-tailed)	.122	<.001	.788	

a. Grouping Variable: Groups	
