## Supplementary material for "Effects of a standardised *Amaranthus tricolor*–*Spinacia oleracea* extract on exercise performance, muscle strength, endurance and recovery in physically active adults: a randomised, double-blind, placebo-controlled trial": Raw statistical outputs: Muscle Endurance & Fatigue index _Not normal tests.rtf

NPar Tests


Notes	
Output Created	18-MAY-2026 15:09:00	
Comments		
Input	Active Dataset	DataSet0	
	Filter	<none>	
	Weight	<none>	
	Split File	<none>	
	N of Rows in Working Data File	64	
Missing Value Handling	Definition of Missing	User-defined missing values are treated as missing.	
	Cases Used	Statistics for each test are based on all cases with valid data for the variable(s) used in that test.	
Syntax	NPAR TESTS
  /M-W= Bench_Press_reps_B Bench_reps_Diff_W6 Bench_reps_Diff_W12 Leg_press_reps_B Leg_reps_Diff_W6
    Leg_reps_Diff_W12 Fatigue_Leg_B Fatigue_Leg_Diff_W6 Fatigue_Leg_Diff_W12 Fatigue_Bench_B
    Fatigue_Bench_Diff_W6 Fatigue_Bench_Diff_W12 BY group(1 2)
  /STATISTICS=DESCRIPTIVES QUARTILES
  /MISSING ANALYSIS.	
Resources	Processor Time	00:00:00.02	
	Elapsed Time	00:00:00.00	
	Number of Cases Allowed^a^	174762	

a. Based on availability of workspace memory.	


Descriptive Statistics	
	N	Mean	Std. Deviation	Minimum	Maximum	Percentiles	
						25th	
Bench_Press_reps_B	64	11.8594	.87045	10.00	14.00	11.0000	
Bench_reps_Diff_W6	64	-1.1719	.67975	-3.00	1.00	-1.0000	
Bench_reps_Diff_W12	64	-2.1094	1.50256	-6.00	1.00	-3.0000	
Leg_press_reps_B	64	13.0625	.95743	11.00	16.00	13.0000	
Leg_reps_Diff_W6	64	-1.7031	1.16401	-6.00	1.00	-2.0000	
Leg_reps_Diff_W12	64	-2.5313	1.85137	-7.00	.00	-4.0000	
Fatigue_Leg_B	64	23.3998	5.12414	9.09	35.71	18.1818	
Fatigue_Leg_Diff_W6	64	1.4687	7.12742	-22.86	26.62	.0000	
Fatigue_Leg_Diff_W12	64	3.8940	7.52729	-15.58	28.02	1.2363	
Fatigue_Bench_B	64	31.6130	5.88161	20.00	45.45	27.2727	
Fatigue_Bench_Diff_W6	64	1.8662	5.25542	-16.36	15.15	.0000	
Fatigue_Bench_Diff_W12	64	5.6408	8.43607	-16.36	23.33	.0000	
group	64	1.5000	.50395	1.00	2.00	1.0000	

Descriptive Statistics	
	Percentiles	
	50th (Median)	75th	
Bench_Press_reps_B	12.0000	12.0000	
Bench_reps_Diff_W6	-1.0000	-1.0000	
Bench_reps_Diff_W12	-2.0000	-1.0000	
Leg_press_reps_B	13.0000	13.0000	
Leg_reps_Diff_W6	-1.5000	-1.0000	
Leg_reps_Diff_W12	-2.0000	-1.0000	
Fatigue_Leg_B	25.0000	25.0000	
Fatigue_Leg_Diff_W6	1.9231	2.5641	
Fatigue_Leg_Diff_W12	3.5714	8.6767	
Fatigue_Bench_B	30.0000	35.6061	
Fatigue_Bench_Diff_W6	2.7273	3.6364	
Fatigue_Bench_Diff_W12	5.5303	12.3470	
group	1.5000	2.0000	


Mann-Whitney Test


Ranks	
	group	N	Mean Rank	Sum of Ranks	
Bench_Press_reps_B	1.00	32	34.97	1119.00	
	2.00	32	30.03	961.00	
	Total	64			
Bench_reps_Diff_W6	1.00	32	24.09	771.00	
	2.00	32	40.91	1309.00	
	Total	64			
Bench_reps_Diff_W12	1.00	32	17.59	563.00	
	2.00	32	47.41	1517.00	
	Total	64			
Leg_press_reps_B	1.00	32	36.77	1176.50	
	2.00	32	28.23	903.50	
	Total	64			
Leg_reps_Diff_W6	1.00	32	17.94	574.00	
	2.00	32	47.06	1506.00	
	Total	64			
Leg_reps_Diff_W12	1.00	32	19.73	631.50	
	2.00	32	45.27	1448.50	
	Total	64			
Fatigue_Leg_B	1.00	32	31.92	1021.50	
	2.00	32	33.08	1058.50	
	Total	64			
Fatigue_Leg_Diff_W6	1.00	32	35.25	1128.00	
	2.00	32	29.75	952.00	
	Total	64			
Fatigue_Leg_Diff_W12	1.00	32	38.31	1226.00	
	2.00	32	26.69	854.00	
	Total	64			
Fatigue_Bench_B	1.00	32	33.02	1056.50	
	2.00	32	31.98	1023.50	
	Total	64			
Fatigue_Bench_Diff_W6	1.00	32	37.34	1195.00	
	2.00	32	27.66	885.00	
	Total	64			
Fatigue_Bench_Diff_W12	1.00	32	40.09	1283.00	
	2.00	32	24.91	797.00	
	Total	64			


Test Statistics^a^	
	Bench_Press_reps_B	Bench_reps_Diff_W6	Bench_reps_Diff_W12	Leg_press_reps_B	
Mann-Whitney U	433.000	243.000	35.000	375.500	
Wilcoxon W	961.000	771.000	563.000	903.500	
Z	-1.152	-4.490	-6.585	-2.004	
Asymp. Sig. (2-tailed)	.249	<.001	<.001	.045	

Test Statistics^a^	
	Leg_reps_Diff_W6	Leg_reps_Diff_W12	Fatigue_Leg_B	Fatigue_Leg_Diff_W6	
Mann-Whitney U	46.000	103.500	493.500	424.000	
Wilcoxon W	574.000	631.500	1021.500	952.000	
Z	-6.614	-5.610	-.254	-1.192	
Asymp. Sig. (2-tailed)	<.001	<.001	.799	.233	

Test Statistics^a^	
	Fatigue_Leg_Diff_W12	Fatigue_Bench_B	Fatigue_Bench_Diff_W6	Fatigue_Bench_Diff_W12	
Mann-Whitney U	326.000	495.500	357.000	269.000	
Wilcoxon W	854.000	1023.500	885.000	797.000	
Z	-2.505	-.226	-2.112	-3.271	
Asymp. Sig. (2-tailed)	.012	.821	.035	.001	

a. Grouping Variable: group	


NPar Tests


Notes	
Output Created	19-MAY-2026 11:18:41	
Comments		
Input	Active Dataset	DataSet0	
	Filter	<none>	
	Weight	<none>	
	Split File	<none>	
	N of Rows in Working Data File	35	
Missing Value Handling	Definition of Missing	User-defined missing values are treated as missing.	
	Cases Used	Statistics for each test are based on all cases with valid data for the variable(s) used in that test.	
Syntax	NPAR TESTS
  /WILCOXON=Bench_reps_B_1 Bench_reps_B_1 Bench_reps_B_2 Bench_reps_B_2 Leg_reps_B_1 Leg_reps_B_1
    Leg_reps_B_2 Leg_reps_B_2 Fatigu_Leg_B_1 Fatigu_Leg_B_1 Fatigu_Leg_B_2 Fatigu_Leg_B_2
    Fatigu_Bench_B_1 Fatigu_Bench_B_1 Fatigu_Bench_B_2 Fatigu_Bench_B_2 Spec_Gravity_B_1
    Spec_Gravity_B_2 WITH Bench_reps_W6_1 Bench_reps_W12_1 Bench_reps_W6_2 Bench_reps_W12_2
    Leg_reps_W6_1 Leg_reps_W12_1 Leg_reps_W6_2 Leg_reps_W12_2 Fatigu_Leg_W6_1 Fatigu_Leg_W12_1
    Fatigu_Leg_W6_2 Fatigu_Leg_W12_2 Fatigu_Bench_W6_1 Fatigu_Bench_W12_1 Fatigu_Bench_W6_2
    Fatigu_Bench_W12_2 Spec_Gravity_W12_1 Spec_Gravity_W12_2 (PAIRED)
  /STATISTICS DESCRIPTIVES QUARTILES
  /MISSING ANALYSIS.	
Resources	Processor Time	00:00:00.02	
	Elapsed Time	00:00:00.00	
	Number of Cases Allowed^a^	95325	

a. Based on availability of workspace memory.	


Descriptive Statistics	
	N	Mean	Std. Deviation	Minimum	Maximum	Percentiles	
						25th	
Bench_reps_B_1	32	11.9687	.89747	10.00	14.00	12.0000	
Bench_reps_B_2	32	11.7500	.84242	10.00	13.00	11.0000	
Leg_reps_B_1	32	13.3438	1.06587	12.00	16.00	13.0000	
Leg_reps_B_2	32	12.7813	.75067	11.00	14.00	12.0000	
Fatigu_Leg_B_1	32	23.4159	5.75970	9.09	35.71	18.1818	
Fatigu_Leg_B_2	32	23.3836	4.49287	16.67	33.33	18.1818	
Fatigu_Bench_B_1	32	31.8355	5.53107	22.22	45.45	27.2727	
Fatigu_Bench_B_2	32	31.3905	6.29346	20.00	45.45	27.2727	
Spec_Gravity_B_1	32	1.0173	.00783	1.01	1.03	1.0100	
Spec_Gravity_B_2	32	1.0203	.00782	1.01	1.03	1.0200	
Bench_reps_W6_1	32	13.5000	1.04727	11.00	16.00	13.0000	
Bench_reps_W12_1	32	15.1875	1.53323	13.00	18.00	14.0000	
Bench_reps_W6_2	32	12.5625	.91361	10.00	14.00	12.0000	
Bench_reps_W12_2	32	12.7500	.95038	11.00	15.00	12.0000	
Leg_reps_W6_1	32	15.8750	1.18458	14.00	18.00	15.0000	
Leg_reps_W12_1	32	17.1250	1.68005	14.00	20.00	16.0000	
Leg_reps_W6_2	32	13.6563	.82733	12.00	15.00	13.0000	
Leg_reps_W12_2	32	14.0625	.91361	12.00	16.00	14.0000	
Fatigu_Leg_W6_1	32	21.0409	5.57378	9.09	35.71	16.6667	
Fatigu_Leg_W12_1	32	16.9345	4.12589	7.69	26.67	14.5604	
Fatigu_Leg_W6_2	32	22.8212	5.52973	15.38	42.86	18.1818	
Fatigu_Leg_W12_2	32	22.0770	6.76971	9.09	42.86	17.0455	
Fatigu_Bench_W6_1	32	29.2561	5.78146	18.18	41.67	25.5682	
Fatigu_Bench_W12_1	32	22.6764	6.72017	8.33	33.33	18.1818	
Fatigu_Bench_W6_2	32	30.2375	6.84169	18.18	41.67	25.0000	
Fatigu_Bench_W12_2	32	29.2680	7.92350	16.67	41.67	22.2222	
Spec_Gravity_W12_1	32	1.0175	.00783	1.00	1.03	1.0100	
Spec_Gravity_W12_2	32	1.0180	.00781	1.01	1.03	1.0100	

Descriptive Statistics	
	Percentiles	
	50th (Median)	75th	
Bench_reps_B_1	12.0000	12.0000	
Bench_reps_B_2	12.0000	12.0000	
Leg_reps_B_1	13.0000	14.0000	
Leg_reps_B_2	13.0000	13.0000	
Fatigu_Leg_B_1	24.0385	25.0000	
Fatigu_Leg_B_2	25.0000	25.0000	
Fatigu_Bench_B_1	30.0000	35.6061	
Fatigu_Bench_B_2	30.0000	36.4583	
Spec_Gravity_B_1	1.0200	1.0250	
Spec_Gravity_B_2	1.0200	1.0250	
Bench_reps_W6_1	13.0000	14.0000	
Bench_reps_W12_1	15.0000	16.0000	
Bench_reps_W6_2	13.0000	13.0000	
Bench_reps_W12_2	13.0000	13.0000	
Leg_reps_W6_1	16.0000	17.0000	
Leg_reps_W12_1	18.0000	18.0000	
Leg_reps_W6_2	14.0000	14.0000	
Leg_reps_W12_2	14.0000	15.0000	
Fatigu_Leg_W6_1	23.0769	23.0769	
Fatigu_Leg_W12_1	15.3846	21.4286	
Fatigu_Leg_W6_2	23.0769	25.0000	
Fatigu_Leg_W12_2	21.4286	26.7045	
Fatigu_Bench_W6_1	30.0000	33.3333	
Fatigu_Bench_W12_1	24.0385	27.2727	
Fatigu_Bench_W6_2	30.0000	36.3636	
Fatigu_Bench_W12_2	30.0000	36.3636	
Spec_Gravity_W12_1	1.0200	1.0200	
Spec_Gravity_W12_2	1.0200	1.0238	


Wilcoxon Signed Ranks Test


Ranks	
	N	Mean Rank	Sum of Ranks	
Bench_reps_W6_1 - Bench_reps_B_1	Negative Ranks	0^a^	.00	.00	
	Positive Ranks	32^b^	16.50	528.00	
	Ties	0^c^			
	Total	32			
Bench_reps_W12_1 - Bench_reps_B_1	Negative Ranks	0^d^	.00	.00	
	Positive Ranks	32^e^	16.50	528.00	
	Ties	0^f^			
	Total	32			
Bench_reps_W6_2 - Bench_reps_B_2	Negative Ranks	1^g^	14.50	14.50	
	Positive Ranks	27^h^	14.50	391.50	
	Ties	4^i^			
	Total	32			
Bench_reps_W12_2 - Bench_reps_B_2	Negative Ranks	1^j^	10.50	10.50	
	Positive Ranks	26^k^	14.13	367.50	
	Ties	5^l^			
	Total	32			
Leg_reps_W6_1 - Leg_reps_B_1	Negative Ranks	0^m^	.00	.00	
	Positive Ranks	32^n^	16.50	528.00	
	Ties	0^o^			
	Total	32			
Leg_reps_W12_1 - Leg_reps_B_1	Negative Ranks	0^p^	.00	.00	
	Positive Ranks	31^q^	16.00	496.00	
	Ties	1^r^			
	Total	32			
Leg_reps_W6_2 - Leg_reps_B_2	Negative Ranks	1^s^	13.50	13.50	
	Positive Ranks	27^t^	14.54	392.50	
	Ties	4^u^			
	Total	32			
Leg_reps_W12_2 - Leg_reps_B_2	Negative Ranks	0^v^	.00	.00	
	Positive Ranks	28^w^	14.50	406.00	
	Ties	4^x^			
	Total	32			
Fatigu_Leg_W6_1 - Fatigu_Leg_B_1	Negative Ranks	26^y^	14.56	378.50	
	Positive Ranks	6^z^	24.92	149.50	
	Ties	0^aa^			
	Total	32			
Fatigu_Leg_W12_1 - Fatigu_Leg_B_1	Negative Ranks	29^ab^	16.05	465.50	
	Positive Ranks	2^ac^	15.25	30.50	
	Ties	1^ad^			
	Total	32			
Fatigu_Leg_W6_2 - Fatigu_Leg_B_2	Negative Ranks	21^ae^	12.14	255.00	
	Positive Ranks	7^af^	21.57	151.00	
	Ties	4^ag^			
	Total	32			
Fatigu_Leg_W12_2 - Fatigu_Leg_B_2	Negative Ranks	20^ah^	13.33	266.50	
	Positive Ranks	9^ai^	18.72	168.50	
	Ties	3^aj^			
	Total	32			
Fatigu_Bench_W6_1 - Fatigu_Bench_B_1	Negative Ranks	26^ak^	14.88	387.00	
	Positive Ranks	6^al^	23.50	141.00	
	Ties	0^am^			
	Total	32			
Fatigu_Bench_W12_1 - Fatigu_Bench_B_1	Negative Ranks	26^an^	18.50	481.00	
	Positive Ranks	5^ao^	3.00	15.00	
	Ties	1^ap^			
	Total	32			
Fatigu_Bench_W6_2 - Fatigu_Bench_B_2	Negative Ranks	10^aq^	6.30	63.00	
	Positive Ranks	3^ar^	9.33	28.00	
	Ties	19^as^			
	Total	32			
Fatigu_Bench_W12_2 - Fatigu_Bench_B_2	Negative Ranks	15^at^	12.43	186.50	
	Positive Ranks	8^au^	11.19	89.50	
	Ties	9^av^			
	Total	32			
Spec_Gravity_W12_1 - Spec_Gravity_B_1	Negative Ranks	13^aw^	14.12	183.50	
	Positive Ranks	14^ax^	13.89	194.50	
	Ties	5^ay^			
	Total	32			
Spec_Gravity_W12_2 - Spec_Gravity_B_2	Negative Ranks	17^az^	12.74	216.50	
	Positive Ranks	9^ba^	14.94	134.50	
	Ties	6^bb^			
	Total	32			

a. Bench_reps_W6_1 < Bench_reps_B_1	
b. Bench_reps_W6_1 > Bench_reps_B_1	
c. Bench_reps_W6_1 = Bench_reps_B_1	
d. Bench_reps_W12_1 < Bench_reps_B_1	
e. Bench_reps_W12_1 > Bench_reps_B_1	
f. Bench_reps_W12_1 = Bench_reps_B_1	
g. Bench_reps_W6_2 < Bench_reps_B_2	
h. Bench_reps_W6_2 > Bench_reps_B_2	
i. Bench_reps_W6_2 = Bench_reps_B_2	
j. Bench_reps_W12_2 < Bench_reps_B_2	
k. Bench_reps_W12_2 > Bench_reps_B_2	
l. Bench_reps_W12_2 = Bench_reps_B_2	
m. Leg_reps_W6_1 < Leg_reps_B_1	
n. Leg_reps_W6_1 > Leg_reps_B_1	
o. Leg_reps_W6_1 = Leg_reps_B_1	
p. Leg_reps_W12_1 < Leg_reps_B_1	
q. Leg_reps_W12_1 > Leg_reps_B_1	
r. Leg_reps_W12_1 = Leg_reps_B_1	
s. Leg_reps_W6_2 < Leg_reps_B_2	
t. Leg_reps_W6_2 > Leg_reps_B_2	
u. Leg_reps_W6_2 = Leg_reps_B_2	
v. Leg_reps_W12_2 < Leg_reps_B_2	
w. Leg_reps_W12_2 > Leg_reps_B_2	
x. Leg_reps_W12_2 = Leg_reps_B_2	
y. Fatigu_Leg_W6_1 < Fatigu_Leg_B_1	
z. Fatigu_Leg_W6_1 > Fatigu_Leg_B_1	
aa. Fatigu_Leg_W6_1 = Fatigu_Leg_B_1	
ab. Fatigu_Leg_W12_1 < Fatigu_Leg_B_1	
ac. Fatigu_Leg_W12_1 > Fatigu_Leg_B_1	
ad. Fatigu_Leg_W12_1 = Fatigu_Leg_B_1	
ae. Fatigu_Leg_W6_2 < Fatigu_Leg_B_2	
af. Fatigu_Leg_W6_2 > Fatigu_Leg_B_2	
ag. Fatigu_Leg_W6_2 = Fatigu_Leg_B_2	
ah. Fatigu_Leg_W12_2 < Fatigu_Leg_B_2	
ai. Fatigu_Leg_W12_2 > Fatigu_Leg_B_2	
aj. Fatigu_Leg_W12_2 = Fatigu_Leg_B_2	
ak. Fatigu_Bench_W6_1 < Fatigu_Bench_B_1	
al. Fatigu_Bench_W6_1 > Fatigu_Bench_B_1	
am. Fatigu_Bench_W6_1 = Fatigu_Bench_B_1	
an. Fatigu_Bench_W12_1 < Fatigu_Bench_B_1	
ao. Fatigu_Bench_W12_1 > Fatigu_Bench_B_1	
ap. Fatigu_Bench_W12_1 = Fatigu_Bench_B_1	
aq. Fatigu_Bench_W6_2 < Fatigu_Bench_B_2	
ar. Fatigu_Bench_W6_2 > Fatigu_Bench_B_2	
as. Fatigu_Bench_W6_2 = Fatigu_Bench_B_2	
at. Fatigu_Bench_W12_2 < Fatigu_Bench_B_2	
au. Fatigu_Bench_W12_2 > Fatigu_Bench_B_2	
av. Fatigu_Bench_W12_2 = Fatigu_Bench_B_2	
aw. Spec_Gravity_W12_1 < Spec_Gravity_B_1	
ax. Spec_Gravity_W12_1 > Spec_Gravity_B_1	
ay. Spec_Gravity_W12_1 = Spec_Gravity_B_1	
az. Spec_Gravity_W12_2 < Spec_Gravity_B_2	
ba. Spec_Gravity_W12_2 > Spec_Gravity_B_2	
bb. Spec_Gravity_W12_2 = Spec_Gravity_B_2	


Test Statistics^a^	
	Bench_reps_W6_1 - Bench_reps_B_1	Bench_reps_W12_1 - Bench_reps_B_1	Bench_reps_W6_2 - Bench_reps_B_2	Bench_reps_W12_2 - Bench_reps_B_2	
Z	-5.070^b^	-4.996^b^	-4.914^b^	-4.520^b^	
Asymp. Sig. (2-tailed)	<.001	<.001	<.001	<.001	

Test Statistics^a^	
	Leg_reps_W6_1 - Leg_reps_B_1	Leg_reps_W12_1 - Leg_reps_B_1	Leg_reps_W6_2 - Leg_reps_B_2	Leg_reps_W12_2 - Leg_reps_B_2	
Z	-5.046^b^	-4.877^b^	-4.794^b^	-4.768^b^	
Asymp. Sig. (2-tailed)	<.001	<.001	<.001	<.001	

Test Statistics^a^	
	Fatigu_Leg_W6_1 - Fatigu_Leg_B_1	Fatigu_Leg_W12_1 - Fatigu_Leg_B_1	Fatigu_Leg_W6_2 - Fatigu_Leg_B_2	Fatigu_Leg_W12_2 - Fatigu_Leg_B_2	
Z	-2.147^c^	-4.268^c^	-1.186^c^	-1.061^c^	
Asymp. Sig. (2-tailed)	.032	<.001	.235	.289	

Test Statistics^a^	
	Fatigu_Bench_W6_1 - Fatigu_Bench_B_1	Fatigu_Bench_W12_1 - Fatigu_Bench_B_1	Fatigu_Bench_W6_2 - Fatigu_Bench_B_2	Fatigu_Bench_W12_2 - Fatigu_Bench_B_2	
Z	-2.303^c^	-4.567^c^	-1.224^c^	-1.475^c^	
Asymp. Sig. (2-tailed)	.021	<.001	.221	.140	

Test Statistics^a^	
	Spec_Gravity_W12_1 - Spec_Gravity_B_1	Spec_Gravity_W12_2 - Spec_Gravity_B_2	
Z	-.134^b^	-1.055^c^	
Asymp. Sig. (2-tailed)	.894	.291	

a. Wilcoxon Signed Ranks Test	
b. Based on negative ranks.	
c. Based on positive ranks.	
