## Supplementary material for "Effects of a standardised *Amaranthus tricolor*–*Spinacia oleracea* extract on exercise performance, muscle strength, endurance and recovery in physically active adults: a randomised, double-blind, placebo-controlled trial": Raw statistical outputs: Serum electro_( Potassium,Bicarbonate, inorganic phosphorus).rtf

NPar Tests


Notes	
Output Created	11-MAY-2026 15:11:10	
Comments		
Input	Active Dataset	DataSet0	
	Filter	<none>	
	Weight	<none>	
	Split File	<none>	
	N of Rows in Working Data File	64	
Missing Value Handling	Definition of Missing	User-defined missing values are treated as missing.	
	Cases Used	Statistics for each test are based on all cases with valid data for the variable(s) used in that test.	
Syntax	NPAR TESTS
  /M-W= B_Ser_Potassium Diff_Ser_Potassium B_Bicarbonate Diff_Bicarbonate B_Inorg_Phosph
    Diff_Inorg_Phosph BY Groups(1 2)
  /STATISTICS=DESCRIPTIVES QUARTILES
  /MISSING ANALYSIS.	
Resources	Processor Time	00:00:00.00	
	Elapsed Time	00:00:00.00	
	Number of Cases Allowed^a^	262144	

a. Based on availability of workspace memory.	


Descriptive Statistics	
	N	Mean	Std. Deviation	Minimum	Maximum	Percentiles	
						25th	
B_Ser_Potassium	64	4.4309	.68928	3.57	7.52	3.9600	
Diff_Ser_Potassium	64	.1048	.84219	-1.70	2.52	-.3975	
B_Bicarbonate	64	21.8953	2.18555	18.70	28.50	20.4000	
Diff_Bicarbonate	64	-5.0656	2.01173	-10.00	.00	-7.0000	
B_Inorg_Phosph	64	3.8850	2.98613	2.46	27.10	3.2225	
Diff_Inorg_Phosph	64	.1863	3.10687	-3.22	23.85	-.6350	
Groups	64	1.5000	.50395	1.00	2.00	1.0000	

Descriptive Statistics	
	Percentiles	
	50th (Median)	75th	
B_Ser_Potassium	4.2000	4.8075	
Diff_Ser_Potassium	.0300	.6100	
B_Bicarbonate	21.4000	23.3000	
Diff_Bicarbonate	-5.0000	-4.0000	
B_Inorg_Phosph	3.5200	3.8850	
Diff_Inorg_Phosph	-.0800	.3000	
Groups	1.5000	2.0000	


Mann-Whitney Test


Ranks	
	Groups	N	Mean Rank	Sum of Ranks	
B_Ser_Potassium	1.00	32	30.91	989.00	
	2.00	32	34.09	1091.00	
	Total	64			
Diff_Ser_Potassium	1.00	32	30.73	983.50	
	2.00	32	34.27	1096.50	
	Total	64			
B_Bicarbonate	1.00	32	31.64	1012.50	
	2.00	32	33.36	1067.50	
	Total	64			
Diff_Bicarbonate	1.00	32	25.13	804.00	
	2.00	32	39.88	1276.00	
	Total	64			
B_Inorg_Phosph	1.00	32	33.27	1064.50	
	2.00	32	31.73	1015.50	
	Total	64			
Diff_Inorg_Phosph	1.00	32	31.27	1000.50	
	2.00	32	33.73	1079.50	
	Total	64			


Test Statistics^a^	
	B_Ser_Potassium	Diff_Ser_Potassium	B_Bicarbonate	Diff_Bicarbonate	
Mann-Whitney U	461.000	455.500	484.500	276.000	
Wilcoxon W	989.000	983.500	1012.500	804.000	
Z	-.685	-.759	-.370	-3.205	
Asymp. Sig. (2-tailed)	.493	.448	.712	.001	

Test Statistics^a^	
	B_Inorg_Phosph	Diff_Inorg_Phosph	
Mann-Whitney U	487.500	472.500	
Wilcoxon W	1015.500	1000.500	
Z	-.329	-.530	
Asymp. Sig. (2-tailed)	.742	.596	

a. Grouping Variable: Groups	


NPar Tests


Notes	
Output Created	13-MAY-2026 14:39:33	
Comments		
Input	Active Dataset	DataSet0	
	Filter	<none>	
	Weight	<none>	
	Split File	<none>	
	N of Rows in Working Data File	32	
Missing Value Handling	Definition of Missing	User-defined missing values are treated as missing.	
	Cases Used	Statistics for each test are based on all cases with valid data for the variable(s) used in that test.	
Syntax	NPAR TESTS
  /WILCOXON=B_Ser_Potassium_1 B_Bicarbonate_1 B_Inorg_Phosph_1 B_Ser_Potassium_2 B_Bicarbonate_2
    B_Inorg_Phosph_2 WITH W12_Ser_Potassium_1 W12_Bicarbonate_1 W12_Inorg_Phosph_1 W12_Ser_Potassium_2
    W12_Bicarbonate_2 W12_Inorg_Phosph_2 (PAIRED)
  /STATISTICS DESCRIPTIVES QUARTILES
  /MISSING ANALYSIS.	
Resources	Processor Time	00:00:00.00	
	Elapsed Time	00:00:00.03	
	Number of Cases Allowed^a^	185042	

a. Based on availability of workspace memory.	


[DataSet0] 


Descriptive Statistics	
	N	Mean	Std. Deviation	Minimum	Maximum	Percentiles	
						25th	
B_Ser_Potassium_1	32	4.4075	.77515	3.57	7.52	3.8925	
B_Bicarbonate_1	32	21.7687	2.12154	18.70	27.40	20.1750	
B_Inorg_Phosph_1	32	4.2672	4.18985	2.46	27.10	3.3000	
B_Ser_Potassium_2	32	4.4544	.60293	3.63	6.23	4.0025	
B_Bicarbonate_2	32	22.0219	2.27451	19.20	28.50	20.5000	
B_Inorg_Phosph_2	32	3.5028	.51495	2.50	4.34	3.1700	
W12_Ser_Potassium_1	32	4.3344	.59914	3.52	5.30	3.7350	
W12_Bicarbonate_1	32	27.5563	2.64318	18.70	32.60	26.2750	
W12_Inorg_Phosph_1	32	3.7469	.57269	2.54	5.66	3.3450	
W12_Ser_Potassium_2	32	4.3178	.54877	3.54	5.40	3.9600	
W12_Bicarbonate_2	32	26.3656	2.11989	23.20	32.50	24.6000	
W12_Inorg_Phosph_2	32	3.6506	.90556	2.17	7.21	3.2400	

Descriptive Statistics	
	Percentiles	
	50th (Median)	75th	
B_Ser_Potassium_1	4.1150	4.8000	
B_Bicarbonate_1	21.4000	22.8750	
B_Inorg_Phosph_1	3.5200	3.8475	
B_Ser_Potassium_2	4.3700	4.8250	
B_Bicarbonate_2	21.3000	23.5250	
B_Inorg_Phosph_2	3.5550	4.0275	
W12_Ser_Potassium_1	4.3000	4.9000	
W12_Bicarbonate_1	28.3000	28.8750	
W12_Inorg_Phosph_1	3.6600	4.1025	
W12_Ser_Potassium_2	4.2000	4.6225	
W12_Bicarbonate_2	26.3000	27.4500	
W12_Inorg_Phosph_2	3.5950	3.8700	


Wilcoxon Signed Ranks Test


Ranks	
	N	Mean Rank	Sum of Ranks	
W12_Ser_Potassium_1 - B_Ser_Potassium_1	Negative Ranks	13^a^	18.77	244.00	
	Positive Ranks	18^b^	14.00	252.00	
	Ties	1^c^			
	Total	32			
W12_Bicarbonate_1 - B_Bicarbonate_1	Negative Ranks	0^d^	.00	.00	
	Positive Ranks	31^e^	16.00	496.00	
	Ties	1^f^			
	Total	32			
W12_Inorg_Phosph_1 - B_Inorg_Phosph_1	Negative Ranks	13^g^	14.88	193.50	
	Positive Ranks	19^h^	17.61	334.50	
	Ties	0^i^			
	Total	32			
W12_Ser_Potassium_2 - B_Ser_Potassium_2	Negative Ranks	19^j^	16.63	316.00	
	Positive Ranks	13^k^	16.31	212.00	
	Ties	0^l^			
	Total	32			
W12_Bicarbonate_2 - B_Bicarbonate_2	Negative Ranks	0^m^	.00	.00	
	Positive Ranks	32^n^	16.50	528.00	
	Ties	0^o^			
	Total	32			
W12_Inorg_Phosph_2 - B_Inorg_Phosph_2	Negative Ranks	15^p^	15.57	233.50	
	Positive Ranks	17^q^	17.32	294.50	
	Ties	0^r^			
	Total	32			

a. W12_Ser_Potassium_1 < B_Ser_Potassium_1	
b. W12_Ser_Potassium_1 > B_Ser_Potassium_1	
c. W12_Ser_Potassium_1 = B_Ser_Potassium_1	
d. W12_Bicarbonate_1 < B_Bicarbonate_1	
e. W12_Bicarbonate_1 > B_Bicarbonate_1	
f. W12_Bicarbonate_1 = B_Bicarbonate_1	
g. W12_Inorg_Phosph_1 < B_Inorg_Phosph_1	
h. W12_Inorg_Phosph_1 > B_Inorg_Phosph_1	
i. W12_Inorg_Phosph_1 = B_Inorg_Phosph_1	
j. W12_Ser_Potassium_2 < B_Ser_Potassium_2	
k. W12_Ser_Potassium_2 > B_Ser_Potassium_2	
l. W12_Ser_Potassium_2 = B_Ser_Potassium_2	
m. W12_Bicarbonate_2 < B_Bicarbonate_2	
n. W12_Bicarbonate_2 > B_Bicarbonate_2	
o. W12_Bicarbonate_2 = B_Bicarbonate_2	
p. W12_Inorg_Phosph_2 < B_Inorg_Phosph_2	
q. W12_Inorg_Phosph_2 > B_Inorg_Phosph_2	
r. W12_Inorg_Phosph_2 = B_Inorg_Phosph_2	


Test Statistics^a^	
	W12_Ser_Potassium_1 - B_Ser_Potassium_1	W12_Bicarbonate_1 - B_Bicarbonate_1	W12_Inorg_Phosph_1 - B_Inorg_Phosph_1	W12_Ser_Potassium_2 - B_Ser_Potassium_2	
Z	-.078^b^	-4.882^b^	-1.318^b^	-.972^c^	
Asymp. Sig. (2-tailed)	.938	<.001	.187	.331	

Test Statistics^a^	
	W12_Bicarbonate_2 - B_Bicarbonate_2	W12_Inorg_Phosph_2 - B_Inorg_Phosph_2	
Z	-4.957^b^	-.570^b^	
Asymp. Sig. (2-tailed)	<.001	.568	

a. Wilcoxon Signed Ranks Test	
b. Based on negative ranks.	
c. Based on positive ranks.	
