## Supplementary material for "Effects of a standardised *Amaranthus tricolor*–*Spinacia oleracea* extract on exercise performance, muscle strength, endurance and recovery in physically active adults: a randomised, double-blind, placebo-controlled trial": Raw statistical outputs: Serum Electro_( Sodium, chloride) By ANCOVA test.rtf

Univariate Analysis of Variance


Notes	
Output Created	11-MAY-2026 12:41:00	
Comments		
Input	Active Dataset	DataSet0	
	Filter	<none>	
	Weight	<none>	
	Split File	<none>	
	N of Rows in Working Data File	64	
Missing Value Handling	Definition of Missing	User-defined missing values are treated as missing.	
	Cases Used	Statistics are based on all cases with valid data for all variables in the model.	
Syntax	UNIANOVA W12_Ser_Sodium BY Groups WITH B_Ser_Sodium
  /METHOD=SSTYPE(3)
  /INTERCEPT=INCLUDE
  /EMMEANS=TABLES(Groups) WITH(B_Ser_Sodium=MEAN) COMPARE ADJ(BONFERRONI)
  /PRINT ETASQ DESCRIPTIVE
  /CRITERIA=ALPHA(.05)
  /DESIGN=B_Ser_Sodium Groups.	
Resources	Processor Time	00:00:00.00	
	Elapsed Time	00:00:00.00	


Between-Subjects Factors	
	N	
Groups	1	32	
	2	32	


Descriptive Statistics	
Dependent Variable:   W12_Ser_Sodium  	
Groups	Mean	Std. Deviation	N	
1	138.8691	2.32319	32	
2	139.6975	2.29807	32	
Total	139.2833	2.32996	64	


Tests of Between-Subjects Effects	
Dependent Variable:   W12_Ser_Sodium  	
Source	Type III Sum of Squares	df	Mean Square	F	Sig.	Partial Eta Squared	
Corrected Model	34.722^a^	2	17.361	3.446	.038	.102	
Intercept	147.461	1	147.461	29.273	<.001	.324	
B_Ser_Sodium	23.741	1	23.741	4.713	.034	.072	
Groups	7.762	1	7.762	1.541	.219	.025	
Error	307.288	61	5.038				
Total	1241931.285	64					
Corrected Total	342.009	63					

a. R Squared = .102 (Adjusted R Squared = .072)	


Estimated Marginal Means


Groups


Estimates	
Dependent Variable:   W12_Ser_Sodium  	
Groups	Mean	Std. Error	95% Confidence Interval	
			Lower Bound	Upper Bound	
1	138.933^a^	.398	138.138	139.729	
2	139.633^a^	.398	138.838	140.429	

a. Covariates appearing in the model are evaluated at the following values: B_Ser_Sodium = 138.3356.	


Pairwise Comparisons	
Dependent Variable:   W12_Ser_Sodium  	
(I) Groups	(J) Groups	Mean Difference (I-J)	Std. Error	Sig.^a^	95% Confidence Interval for Difference^a^	
					Lower Bound	Upper Bound	
1	2	-.700	.564	.219	-1.829	.428	
2	1	.700	.564	.219	-.428	1.829	

Based on estimated marginal means	
a. Adjustment for multiple comparisons: Bonferroni.	


Univariate Tests	
Dependent Variable:   W12_Ser_Sodium  	
	Sum of Squares	df	Mean Square	F	Sig.	Partial Eta Squared	
Contrast	7.762	1	7.762	1.541	.219	.025	
Error	307.288	61	5.038				

The F tests the effect of Groups. This test is based on the linearly independent pairwise comparisons among the estimated marginal means.	


Univariate Analysis of Variance


Notes	
Output Created	11-MAY-2026 12:41:12	
Comments		
Input	Active Dataset	DataSet0	
	Filter	<none>	
	Weight	<none>	
	Split File	<none>	
	N of Rows in Working Data File	64	
Missing Value Handling	Definition of Missing	User-defined missing values are treated as missing.	
	Cases Used	Statistics are based on all cases with valid data for all variables in the model.	
Syntax	UNIANOVA W12_Ser_Chloride BY Groups WITH B_Ser_Chloride
  /METHOD=SSTYPE(3)
  /INTERCEPT=INCLUDE
  /EMMEANS=TABLES(Groups) WITH(B_Ser_Chloride=MEAN) COMPARE ADJ(BONFERRONI)
  /PRINT ETASQ DESCRIPTIVE
  /CRITERIA=ALPHA(.05)
  /DESIGN=B_Ser_Chloride Groups.	
Resources	Processor Time	00:00:00.00	
	Elapsed Time	00:00:00.01	


Between-Subjects Factors	
	N	
Groups	1	32	
	2	32	


Descriptive Statistics	
Dependent Variable:   W12_Ser_Chloride  	
Groups	Mean	Std. Deviation	N	
1	99.5344	1.90089	32	
2	99.2778	2.06531	32	
Total	99.4061	1.97323	64	


Tests of Between-Subjects Effects	
Dependent Variable:   W12_Ser_Chloride  	
Source	Type III Sum of Squares	df	Mean Square	F	Sig.	Partial Eta Squared	
Corrected Model	28.617^a^	2	14.309	4.028	.023	.117	
Intercept	65.515	1	65.515	18.444	<.001	.232	
B_Ser_Chloride	27.564	1	27.564	7.760	.007	.113	
Groups	8.451	1	8.451	2.379	.128	.038	
Error	216.681	61	3.552				
Total	632665.873	64					
Corrected Total	245.298	63					

a. R Squared = .117 (Adjusted R Squared = .088)	


Estimated Marginal Means


Groups


Estimates	
Dependent Variable:   W12_Ser_Chloride  	
Groups	Mean	Std. Error	95% Confidence Interval	
			Lower Bound	Upper Bound	
1	99.798^a^	.346	99.105	100.490	
2	99.015^a^	.346	98.322	99.707	

a. Covariates appearing in the model are evaluated at the following values: B_Ser_Chloride = 99.4416.	


Pairwise Comparisons	
Dependent Variable:   W12_Ser_Chloride  	
(I) Groups	(J) Groups	Mean Difference (I-J)	Std. Error	Sig.^a^	95% Confidence Interval for Difference^a^	
					Lower Bound	Upper Bound	
1	2	.783	.508	.128	-.232	1.798	
2	1	-.783	.508	.128	-1.798	.232	

Based on estimated marginal means	
a. Adjustment for multiple comparisons: Bonferroni.	


Univariate Tests	
Dependent Variable:   W12_Ser_Chloride  	
	Sum of Squares	df	Mean Square	F	Sig.	Partial Eta Squared	
Contrast	8.451	1	8.451	2.379	.128	.038	
Error	216.681	61	3.552				

The F tests the effect of Groups. This test is based on the linearly independent pairwise comparisons among the estimated marginal means.	


T-Test


Notes	
Output Created	11-MAY-2026 12:41:46	
Comments		
Input	Active Dataset	DataSet0	
	Filter	<none>	
	Weight	<none>	
	Split File	<none>	
	N of Rows in Working Data File	64	
Missing Value Handling	Definition of Missing	User defined missing values are treated as missing.	
	Cases Used	Statistics for each analysis are based on the cases with no missing or out-of-range data for any variable in the analysis.	
Syntax	T-TEST PAIRS=B_Ser_Sodium_1 B_Ser_Chloride_1 B_Ser_Sodium_2 B_Ser_Chloride_2 WITH W12_Ser_Sodium_1
    W12_Ser_Chloride_1 W12_Ser_Sodium_2 W12_Ser_Chloride_2 (PAIRED)
  /ES DISPLAY(TRUE) STANDARDIZER(SD)
  /CRITERIA=CI(.9500)
  /MISSING=ANALYSIS.	
Resources	Processor Time	00:00:00.00	
	Elapsed Time	00:00:00.00	


Paired Samples Statistics	
	Mean	N	Std. Deviation	Std. Error Mean	
Pair 1	B_Ser_Sodium_1	138.1134	32	1.99371	.35244	
	W12_Ser_Sodium_1	138.8691	32	2.32319	.41069	
Pair 2	B_Ser_Chloride_1	98.7722	32	1.56657	.27693	
	W12_Ser_Chloride_1	99.5344	32	1.90089	.33603	
Pair 3	B_Ser_Sodium_2	138.5578	32	2.28909	.40466	
	W12_Ser_Sodium_2	139.6975	32	2.29807	.40625	
Pair 4	B_Ser_Chloride_2	100.1109	32	1.81525	.32089	
	W12_Ser_Chloride_2	99.2778	32	2.06531	.36510	


Paired Samples Correlations	
	N	Correlation	Significance	
			One-Sided p	Two-Sided p	
Pair 1	B_Ser_Sodium_1 & W12_Ser_Sodium_1	32	.152	.203	.406	
Pair 2	B_Ser_Chloride_1 & W12_Ser_Chloride_1	32	.349	.025	.050	
Pair 3	B_Ser_Sodium_2 & W12_Ser_Sodium_2	32	.371	.018	.037	
Pair 4	B_Ser_Chloride_2 & W12_Ser_Chloride_2	32	.326	.035	.069	


Paired Samples Test	
	Paired Differences	
	Mean	Std. Deviation	Std. Error Mean	95% Confidence Interval of the Difference	
				Lower	
Pair 1	B_Ser_Sodium_1 - W12_Ser_Sodium_1	-.75562	2.82192	.49885	-1.77304	
Pair 2	B_Ser_Chloride_1 - W12_Ser_Chloride_1	-.76219	1.99657	.35295	-1.48203	
Pair 3	B_Ser_Sodium_2 - W12_Ser_Sodium_2	-1.13969	2.57233	.45473	-2.06711	
Pair 4	B_Ser_Chloride_2 - W12_Ser_Chloride_2	.83312	2.26272	.40000	.01733	

Paired Samples Test	
	Paired Differences	t	df	Significance	
	95% Confidence Interval of the Difference			One-Sided p	
	Upper				
Pair 1	B_Ser_Sodium_1 - W12_Ser_Sodium_1	.26179	-1.515	31	.070	
Pair 2	B_Ser_Chloride_1 - W12_Ser_Chloride_1	-.04235	-2.159	31	.019	
Pair 3	B_Ser_Sodium_2 - W12_Ser_Sodium_2	-.21226	-2.506	31	.009	
Pair 4	B_Ser_Chloride_2 - W12_Ser_Chloride_2	1.64892	2.083	31	.023	

Paired Samples Test	
	Significance	
	Two-Sided p	
		
Pair 1	B_Ser_Sodium_1 - W12_Ser_Sodium_1	.140	
Pair 2	B_Ser_Chloride_1 - W12_Ser_Chloride_1	.039	
Pair 3	B_Ser_Sodium_2 - W12_Ser_Sodium_2	.018	
Pair 4	B_Ser_Chloride_2 - W12_Ser_Chloride_2	.046	


Paired Samples Effect Sizes	
	Standardizer^a^	Point Estimate	95% Confidence Interval	
			Lower	
Pair 1	B_Ser_Sodium_1 - W12_Ser_Sodium_1	Cohen's d	2.82192	-.268	-.618	
		Hedges' correction	2.89257	-.261	-.603	
Pair 2	B_Ser_Chloride_1 - W12_Ser_Chloride_1	Cohen's d	1.99657	-.382	-.738	
		Hedges' correction	2.04655	-.372	-.720	
Pair 3	B_Ser_Sodium_2 - W12_Ser_Sodium_2	Cohen's d	2.57233	-.443	-.803	
		Hedges' correction	2.63673	-.432	-.784	
Pair 4	B_Ser_Chloride_2 - W12_Ser_Chloride_2	Cohen's d	2.26272	.368	.007	
		Hedges' correction	2.31937	.359	.007	

Paired Samples Effect Sizes	
	95% Confidence Interval^a^	
	Upper	
Pair 1	B_Ser_Sodium_1 - W12_Ser_Sodium_1	Cohen's d	.087	
		Hedges' correction	.085	
Pair 2	B_Ser_Chloride_1 - W12_Ser_Chloride_1	Cohen's d	-.020	
		Hedges' correction	-.019	
Pair 3	B_Ser_Sodium_2 - W12_Ser_Sodium_2	Cohen's d	-.076	
		Hedges' correction	-.074	
Pair 4	B_Ser_Chloride_2 - W12_Ser_Chloride_2	Cohen's d	.724	
		Hedges' correction	.706	

a. The denominator used in estimating the effect sizes. 
Cohen's d uses the sample standard deviation of the mean difference. 
Hedges' correction uses the sample standard deviation of the mean difference, plus a correction factor.	
