## Supplementary material for "Effects of a standardised *Amaranthus tricolor*–*Spinacia oleracea* extract on exercise performance, muscle strength, endurance and recovery in physically active adults: a randomised, double-blind, placebo-controlled trial": Raw statistical outputs: Ventilatory Threshold (VT)- Non para tests.rtf

NPar 1s


Notes	
Output Created	12-JUN-2026 10:02:06	
Comments		
Input	Active Dataset	DataSet0	
	Filter	<none>	
	Weight	<none>	
	Split File	<none>	
	N of Rows in Working Data File	60	
Missing Value Handling	Definition of Missing	User-defined missing values are treated as missing.	
	Cases Used	Statistics for each 1 are based on all cases with valid data for the variable(s) used in that 1.	
Syntax	NPAR 1S
  /WILCOXON=Bas_VT_1 B_VT_1 Bas_VT_C B_VT_C WITH D45_T D90_T D45_C D90_C (PAIRED)
  /STATISTICS DESCRIPTIVES QUARTILES
  /MISSING ANALYSIS.	
Resources	Processor Time	00:00:00.02	
	Elapsed Time	00:00:00.02	
	Number of Cases Allowed^a^	241979	

a. Based on availability of workspace memory.	


[DataSet0] 


Descriptive Statistics	
	N	Mean	Std. Deviation	Minimum	Maximum	Percentiles	
						25th	
Bas_VT_1	28	1.8186	.39492	1.01	2.76	1.6200	
B_VT_1	30	1.8090	.38468	1.01	2.76	1.6050	
Bas_VT_2	27	1.8381	.49265	1.05	3.74	1.5200	
B_VT_2	30	1.8100	.49184	1.05	3.74	1.5150	
D45_1	28	1.7750	.34723	.91	2.40	1.4900	
D90_1	30	1.9187	.39162	1.09	2.81	1.7050	
D45_2	27	1.7267	.35335	.92	2.32	1.5000	
D90_2	30	1.7363	.37604	.97	2.68	1.5150	

Descriptive Statistics	
	Percentiles	
	50th (Median)	75th	
Bas_VT_1	1.7600	2.0875	
B_VT_1	1.7600	2.0425	
Bas_VT_2	1.8200	1.9800	
B_VT_2	1.8150	1.9850	
D45_1	1.8500	2.0050	
D90_1	1.8850	2.1775	
D45_2	1.7900	1.9400	
D90_2	1.6950	2.0225	


Wilcoxon Signed Ranks 1


Ranks	
	N	Mean Rank	Sum of Ranks	
D45_1 - Bas_VT_1	Negative Ranks	15^a^	16.37	245.50	
	Positive Ranks	13^b^	12.35	160.50	
	Ties	0^c^			
	Total	28			
D90_1 - B_VT_1	Negative Ranks	9^d^	14.17	127.50	
	Positive Ranks	21^e^	16.07	337.50	
	Ties	0^f^			
	Total	30			
D45_2 - Bas_VT_2	Negative Ranks	17^g^	14.00	238.00	
	Positive Ranks	8^h^	10.88	87.00	
	Ties	2^i^			
	Total	27			
D90_2 - B_VT_2	Negative Ranks	18^j^	16.22	292.00	
	Positive Ranks	12^k^	14.42	173.00	
	Ties	0^l^			
	Total	30			

a. D45_1 < Bas_VT_1	
b. D45_1 > Bas_VT_1	
c. D45_1 = Bas_VT_1	
d. D90_1 < B_VT_1	
e. D90_1 > B_VT_1	
f. D90_1 = B_VT_1	
g. D45_2 < Bas_VT_2	
h. D45_2 > Bas_VT_2	
i. D45_2 = Bas_VT_2	
j. D90_2 < B_VT_2	
k. D90_2 > B_VT_2	
l. D90_2 = B_VT_2	


1 Statistics^a^	
	D45_1 - Bas_VT_1	D90_1 - B_VT_1	D45_2 - Bas_VT_2	D90_2 - B_VT_2	
Z	-.968^b^	-2.160^c^	-2.033^b^	-1.224^b^	
Asymp. Sig. (2-tailed)	.333	.031	.042	.221	

a. Wilcoxon Signed Ranks 1	
b. Based on positive ranks.	
c. Based on negative ranks.	


NPar 1s


Notes	
Output Created	12-JUN-2026 10:02:54	
Comments		
Input	Active Dataset	DataSet0	
	Filter	<none>	
	Weight	<none>	
	Split File	<none>	
	N of Rows in Working Data File	60	
Missing Value Handling	Definition of Missing	User-defined missing values are treated as missing.	
	Cases Used	Statistics for each 1 are based on all cases with valid data for the variable(s) used in that 1.	
Syntax	NPAR 1S
  /M-W= Diff_D45 Diff_D90 Baseline BY Group(1 2)
  /STATISTICS=DESCRIPTIVES QUARTILES
  /MISSING ANALYSIS.	
Resources	Processor Time	00:00:00.00	
	Elapsed Time	00:00:00.01	
	Number of Cases Allowed^a^	349525	

a. Based on availability of workspace memory.	


Descriptive Statistics	
	N	Mean	Std. Deviation	Minimum	Maximum	Percentiles	
						25th	50th (Median)	
Diff_D45	55	.0769	.27964	-.26	1.62	-.0800	.0500	
Diff_D90	60	-.0180	.36740	-1.10	1.86	-.1775	-.0350	
Baseline	60	1.8095	.43777	1.01	3.74	1.5450	1.7850	
Group	60	1.5000	.50422	1.00	2.00	1.0000	1.5000	

Descriptive Statistics	
	Percentiles	
	75th	
Diff_D45	.2000	
Diff_D90	.1275	
Baseline	2.0150	
Group	2.0000	


Mann-Whitney 1


Ranks	
	Group	N	Mean Rank	Sum of Ranks	
Diff_D45	1.00	28	26.96	755.00	
	2.00	27	29.07	785.00	
	Total	55			
Diff_D90	1.00	30	25.08	752.50	
	2.00	30	35.92	1077.50	
	Total	60			
Baseline	1.00	30	31.37	941.00	
	2.00	30	29.63	889.00	
	Total	60			


1 Statistics^a^	
	Diff_D45	Diff_D90	Baseline	
Mann-Whitney U	349.000	287.500	424.000	
Wilcoxon W	755.000	752.500	889.000	
Z	-.489	-2.403	-.384	
Asymp. Sig. (2-tailed)	.625	.016	.701	

a. Grouping Variable: Group	
