## Supplementary material for "Effects of a standardised *Amaranthus tricolor*–*Spinacia oleracea* extract on exercise performance, muscle strength, endurance and recovery in physically active adults: a randomised, double-blind, placebo-controlled trial": Raw statistical outputs: VO2 max and Maximum Ventilation cpet_Output_LMM.rtf

Mixed Model Analysis


Notes	
Output Created	18-MAY-2026 15:37:38	
Comments		
Input	Active Dataset	DataSet0	
	Filter	<none>	
	Weight	<none>	
	Split File	<none>	
	N of Rows in Working Data File	180	
Missing Value Handling	Definition of Missing	User-defined missing values are treated as valid data.	
	Cases Used	Statistics are based on all cases with valid data for all variables in the model.	
Weight Handling	not applicable	
Syntax	MIXED Vo2_max BY Treatment Time
  /CRITERIA=DFMETHOD(SATTERTHWAITE) CIN(95) MXITER(100) MXSTEP(10) SCORING(1)
    SINGULAR(0.000000000001) HCONVERGE(0.00000001, RELATIVE) LCONVERGE(0, ABSOLUTE) PCONVERGE(0,
    ABSOLUTE)
  /FIXED=Treatment Time Treatment*Time | SSTYPE(3)
  /METHOD=REML
  /PRINT=COVB DESCRIPTIVES G  SOLUTION
  /RANDOM=Treatment Time Treatment*Time | COVTYPE(VC)
  /EMMEANS=TABLES(Treatment) COMPARE ADJ(BONFERRONI)
  /EMMEANS=TABLES(Time) COMPARE ADJ(BONFERRONI)
  /EMMEANS=TABLES(Treatment*Time) .	
Resources	Processor Time	00:00:00.05	
	Elapsed Time	00:00:00.03	


Descriptive Statistics	
Vo2_max  	
Treatment	Time	Count	Mean	Standard Deviation	Coefficient of Variation	
1.00	1.00	30	2.0963	.41126	19.6%	
	2.00	28	2.1236	.40009	18.8%	
	3.00	30	2.2303	.42630	19.1%	
	Total	88	2.1507	.41240	19.2%	
2.00	1.00	30	2.0873	.44581	21.4%	
	2.00	27	2.0485	.42540	20.8%	
	3.00	30	2.1203	.44557	21.0%	
	Total	87	2.0867	.43535	20.9%	
Total	1.00	60	2.0918	.42526	20.3%	
	2.00	55	2.0867	.41061	19.7%	
	3.00	60	2.1753	.43587	20.0%	
	Total	175	2.1189	.42396	20.0%	


Model Dimension^a^	
	Number of Levels	Covariance Structure	Number of Parameters	
Fixed Effects	Intercept	1		1	
	Treatment	2		1	
	Time	3		2	
	Treatment * Time	6		2	
Random Effects	Treatment + Time + Treatment * Time	11	Variance Components	3	
Residual			1	
Total	23		10	

a. Dependent Variable: Vo2_max.	


Information Criteria^a^	
-2 Restricted Log Likelihood	211.70955755	
Akaike's Information Criterion (AIC)	219.70955755	
Hurvich and Tsai's Criterion (AICC)	219.95345999	
Bozdogan's Criterion (CAIC)	236.22915241	
Schwarz's Bayesian Criterion (BIC)	232.22915241	

The information criteria are displayed in smaller-is-better form.^a^	
a. Dependent Variable: Vo2_max.	


Coefficients of Determination	
Pseudo-R Square Measures	Marginal	.010	
	Conditional	.435	


Intraclass Correlation Coefficients	
Overall ICCs	Adjusted	.429	
	Conditional	.425	


Fixed Effects


Type III Tests of Fixed Effects^a^	
Source	Numerator df	Denominator df	F	Sig.	
Intercept	1	5.401	94.633	<.001	
Treatment	1	2.783	.033	.868	
Time	2	4.346	.035	.966	
Treatment * Time	2	4.037	.027	.973	

a. Dependent Variable: Vo2_max.	


Estimates of Fixed Effects^a^	
Parameter	Estimate	Std. Error	df	t	Sig.	95% Confidence Interval	
						Lower Bound	
Intercept	2.120	.378	9.345	5.611	<.001	1.270	
[Treatment=1.00]	.110	.437	5.147	.251	.811	-1.005	
[Treatment=2.00]	0^b^	0	.	.	.	.	
[Time=1.00]	-.033	.437	6.797	-.075	.942	-1.074	
[Time=2.00]	-.072	.438	6.845	-.164	.875	-1.113	
[Time=3.00]	0^b^	0	.	.	.	.	
[Treatment=1.00] * [Time=1.00]	-.101	.441	4.007	-.229	.830	-1.324	
[Treatment=1.00] * [Time=2.00]	-.035	.442	4.053	-.079	.941	-1.256	
[Treatment=1.00] * [Time=3.00]	0^b^	0	.	.	.	.	
[Treatment=2.00] * [Time=1.00]	0^b^	0	.	.	.	.	
[Treatment=2.00] * [Time=2.00]	0^b^	0	.	.	.	.	
[Treatment=2.00] * [Time=3.00]	0^b^	0	.	.	.	.	

Estimates of Fixed Effects^a^	
Parameter	95% Confidence Interval	
	Upper Bound	
Intercept	2.970	
[Treatment=1.00]	1.225	
[Treatment=2.00]	.	
[Time=1.00]	1.008	
[Time=2.00]	.969	
[Time=3.00]	.	
[Treatment=1.00] * [Time=1.00]	1.122	
[Treatment=1.00] * [Time=2.00]	1.186	
[Treatment=1.00] * [Time=3.00]	.	
[Treatment=2.00] * [Time=1.00]	.	
[Treatment=2.00] * [Time=2.00]	.	
[Treatment=2.00] * [Time=3.00]	.	

a. Dependent Variable: Vo2_max.	
b. This parameter is set to zero because it is redundant.	


Covariance Matrix for Estimates of Fixed Effects^a^	
Parameter	Intercept	[Treatment=1.00]	[Treatment=2.00]	[Time=1.00]	[Time=2.00]	
Intercept	.143	-.096	0^b^	-.096	-.096	
[Treatment=1.00]	-.096	.191	0^b^	.049	.049	
[Treatment=2.00]	0^b^	0^b^	0^b^	0^b^	0^b^	
[Time=1.00]	-.096	.049	0^b^	.191	.096	
[Time=2.00]	-.096	.049	0^b^	.096	.192	
[Time=3.00]	0^b^	0^b^	0^b^	0^b^	0^b^	
[Treatment=1.00] * [Time=1.00]	.049	-.097	0^b^	-.097	-.049	
[Treatment=1.00] * [Time=2.00]	.049	-.097	0^b^	-.049	-.098	
[Treatment=1.00] * [Time=3.00]	0^b^	0^b^	0^b^	0^b^	0^b^	
[Treatment=2.00] * [Time=1.00]	0^b^	0^b^	0^b^	0^b^	0^b^	
[Treatment=2.00] * [Time=2.00]	0^b^	0^b^	0^b^	0^b^	0^b^	
[Treatment=2.00] * [Time=3.00]	0^b^	0^b^	0^b^	0^b^	0^b^	

Covariance Matrix for Estimates of Fixed Effects^a^	
Parameter	[Time=3.00]	[Treatment=1.00] * [Time=1.00]	[Treatment=1.00] * [Time=2.00]	[Treatment=1.00] * [Time=3.00]	
Intercept	0^b^	.049	.049	0^b^	
[Treatment=1.00]	0^b^	-.097	-.097	0^b^	
[Treatment=2.00]	0^b^	0^b^	0^b^	0^b^	
[Time=1.00]	0^b^	-.097	-.049	0^b^	
[Time=2.00]	0^b^	-.049	-.098	0^b^	
[Time=3.00]	0^b^	0^b^	0^b^	0^b^	
[Treatment=1.00] * [Time=1.00]	0^b^	.194	.097	0^b^	
[Treatment=1.00] * [Time=2.00]	0^b^	.097	.195	0^b^	
[Treatment=1.00] * [Time=3.00]	0^b^	0^b^	0^b^	0^b^	
[Treatment=2.00] * [Time=1.00]	0^b^	0^b^	0^b^	0^b^	
[Treatment=2.00] * [Time=2.00]	0^b^	0^b^	0^b^	0^b^	
[Treatment=2.00] * [Time=3.00]	0^b^	0^b^	0^b^	0^b^	

Covariance Matrix for Estimates of Fixed Effects^a^	
Parameter	[Treatment=2.00] * [Time=1.00]	[Treatment=2.00] * [Time=2.00]	[Treatment=2.00] * [Time=3.00]	
Intercept	0^b^	0^b^	0^b^	
[Treatment=1.00]	0^b^	0^b^	0^b^	
[Treatment=2.00]	0^b^	0^b^	0^b^	
[Time=1.00]	0^b^	0^b^	0^b^	
[Time=2.00]	0^b^	0^b^	0^b^	
[Time=3.00]	0^b^	0^b^	0^b^	
[Treatment=1.00] * [Time=1.00]	0^b^	0^b^	0^b^	
[Treatment=1.00] * [Time=2.00]	0^b^	0^b^	0^b^	
[Treatment=1.00] * [Time=3.00]	0^b^	0^b^	0^b^	
[Treatment=2.00] * [Time=1.00]	0^b^	0^b^	0^b^	
[Treatment=2.00] * [Time=2.00]	0^b^	0^b^	0^b^	
[Treatment=2.00] * [Time=3.00]	0^b^	0^b^	0^b^	

a. Dependent Variable: Vo2_max.	
b. The covariance is set to zero because it is associated with a redundant parameter.	


Covariance Parameters


Estimates of Covariance Parameters^a^	
Parameter	Estimate	Std. Error	
Residual	.182	.020	
Treatment	Variance	.047	.054	
Time	Variance	.047	.051	
Treatment * Time	Variance	.042	.034	

a. Dependent Variable: Vo2_max.	


Covariance Matrix for Estimates of Covariance Parameters^a^	
Parameter	Residual	Treatment	Time	Treatment * Time	
		Variance	Variance	Variance	
Residual	.000	5.704E-10	-1.090E-9	-1.343E-5	
Treatment	Variance	5.704E-10	.003	.000	.000	
Time	Variance	-1.090E-9	.000	.003	-.001	
Treatment * Time	Variance	-1.343E-5	.000	-.001	.001	

a. Dependent Variable: Vo2_max.	


Random Effects Covariance Structures (G)


Treatment^a^	
	[Treatment=1.00]	[Treatment=2.00]	
[Treatment=1.00]	.047	0	
[Treatment=2.00]	0	.047	

Variance Components^a^	
a. Dependent Variable: Vo2_max.	


Time^a^	
	[Time=1.00]	[Time=2.00]	[Time=3.00]	
[Time=1.00]	.047	0	0	
[Time=2.00]	0	.047	0	
[Time=3.00]	0	0	.047	

Variance Components^a^	
a. Dependent Variable: Vo2_max.	


Treatment * Time^a^	
	[Treatment=1.00] * [Time=1.00]	[Treatment=1.00] * [Time=2.00]	[Treatment=1.00] * [Time=3.00]	[Treatment=2.00] * [Time=1.00]	
[Treatment=1.00] * [Time=1.00]	.042	0	0	0	
[Treatment=1.00] * [Time=2.00]	0	.042	0	0	
[Treatment=1.00] * [Time=3.00]	0	0	.042	0	
[Treatment=2.00] * [Time=1.00]	0	0	0	.042	
[Treatment=2.00] * [Time=2.00]	0	0	0	0	
[Treatment=2.00] * [Time=3.00]	0	0	0	0	

Treatment * Time^a^	
	[Treatment=2.00] * [Time=2.00]	[Treatment=2.00] * [Time=3.00]	
[Treatment=1.00] * [Time=1.00]	0	0	
[Treatment=1.00] * [Time=2.00]	0	0	
[Treatment=1.00] * [Time=3.00]	0	0	
[Treatment=2.00] * [Time=1.00]	0	0	
[Treatment=2.00] * [Time=2.00]	.042	0	
[Treatment=2.00] * [Time=3.00]	0	.042	

Variance Components^a^	
a. Dependent Variable: Vo2_max.	


Estimated Marginal Means


1. Treatment


Estimates^a^	
Treatment	Mean	Std. Error	df	95% Confidence Interval	
				Lower Bound	Upper Bound	
1.00	2.150	.281	4.437	1.399	2.901	
2.00	2.085	.281	4.440	1.334	2.837	

a. Dependent Variable: Vo2_max.	


Pairwise Comparisons^a^	
(I) Treatment	(J) Treatment	Mean Difference (I-J)	Std. Error	df	Sig.^b^	95% Confidence Interval for Difference^b^	
						Lower Bound	
1.00	2.00	.065	.356	2.783	.868	-1.120	
2.00	1.00	-.065	.356	2.783	.868	-1.249	

Pairwise Comparisons^a^	
(I) Treatment	(J) Treatment	95% Confidence Interval for Difference	
		Upper Bound	
1.00	2.00	1.249	
2.00	1.00	1.120	

Based on estimated marginal means^a^	
a. Dependent Variable: Vo2_max.	
b. Adjustment for multiple comparisons: Bonferroni.	


Univariate Tests^a^	
Numerator df	Denominator df	F	Sig.	
1	2.783	.033	.868	

The F tests the effect of Treatment. This test is based on the linearly independent pairwise comparisons among the estimated marginal means.^a^	
a. Dependent Variable: Vo2_max.	


2. Time


Estimates^a^	
Time	Mean	Std. Error	df	95% Confidence Interval	
				Lower Bound	Upper Bound	
1.00	2.092	.308	6.957	1.362	2.821	
2.00	2.086	.309	6.998	1.356	2.816	
3.00	2.175	.308	6.957	1.446	2.905	

a. Dependent Variable: Vo2_max.	


Pairwise Comparisons^a^	
(I) Time	(J) Time	Mean Difference (I-J)	Std. Error	df	Sig.^b^	95% Confidence Interval for Difference^b^	
						Lower Bound	
1.00	2.00	.006	.378	4.352	1.000	-1.424	
	3.00	-.083	.378	4.335	1.000	-1.515	
2.00	1.00	-.006	.378	4.352	1.000	-1.435	
	3.00	-.089	.378	4.352	1.000	-1.519	
3.00	1.00	.083	.378	4.335	1.000	-1.348	
	2.00	.089	.378	4.352	1.000	-1.340	

Pairwise Comparisons^a^	
(I) Time	(J) Time	95% Confidence Interval for Difference	
		Upper Bound	
1.00	2.00	1.435	
	3.00	1.348	
2.00	1.00	1.424	
	3.00	1.340	
3.00	1.00	1.515	
	2.00	1.519	

Based on estimated marginal means^a^	
a. Dependent Variable: Vo2_max.	
b. Adjustment for multiple comparisons: Bonferroni.	


Univariate Tests^a^	
Numerator df	Denominator df	F	Sig.	
2	4.346	.035	.966	

The F tests the effect of Time. This test is based on the linearly independent pairwise comparisons among the estimated marginal means.^a^	
a. Dependent Variable: Vo2_max.	


3. Treatment * Time^a^	
Treatment	Time	Mean	Std. Error	df	95% Confidence Interval	
					Lower Bound	Upper Bound	
1.00	1.00	2.096	.378	9.345	1.246	2.946	
	2.00	2.124	.378	9.402	1.273	2.974	
	3.00	2.230	.378	9.345	1.380	3.080	
2.00	1.00	2.087	.378	9.345	1.237	2.937	
	2.00	2.049	.379	9.434	1.198	2.899	
	3.00	2.120	.378	9.345	1.270	2.970	

a. Dependent Variable: Vo2_max.	


Mixed Model Analysis


Notes	
Output Created	18-MAY-2026 15:37:52	
Comments		
Input	Active Dataset	DataSet0	
	Filter	<none>	
	Weight	<none>	
	Split File	<none>	
	N of Rows in Working Data File	180	
Missing Value Handling	Definition of Missing	User-defined missing values are treated as valid data.	
	Cases Used	Statistics are based on all cases with valid data for all variables in the model.	
Weight Handling	not applicable	
Syntax	MIXED Max_venti BY Treatment Time
  /CRITERIA=DFMETHOD(SATTERTHWAITE) CIN(95) MXITER(100) MXSTEP(10) SCORING(1)
    SINGULAR(0.000000000001) HCONVERGE(0.00000001, RELATIVE) LCONVERGE(0, ABSOLUTE) PCONVERGE(0,
    ABSOLUTE)
  /FIXED=Treatment Time Treatment*Time | SSTYPE(3)
  /METHOD=REML
  /PRINT=COVB DESCRIPTIVES G  SOLUTION
  /RANDOM=Treatment Time Treatment*Time | COVTYPE(VC)
  /EMMEANS=TABLES(Treatment) COMPARE ADJ(BONFERRONI)
  /EMMEANS=TABLES(Time) COMPARE ADJ(BONFERRONI)
  /EMMEANS=TABLES(Treatment*Time) .	
Resources	Processor Time	00:00:00.03	
	Elapsed Time	00:00:00.02	


Descriptive Statistics	
Max_venti  	
Treatment	Time	Count	Mean	Standard Deviation	Coefficient of Variation	
1.00	1.00	30	79.2590	14.91460	18.8%	
	2.00	28	75.4979	13.93224	18.5%	
	3.00	30	80.7307	15.62682	19.4%	
	Total	88	78.5640	14.85257	18.9%	
2.00	1.00	30	80.6140	21.40994	26.6%	
	2.00	27	72.3944	18.90029	26.1%	
	3.00	30	74.5870	20.44610	27.4%	
	Total	87	75.9848	20.38973	26.8%	
Total	1.00	60	79.9365	18.30608	22.9%	
	2.00	55	73.9744	16.47727	22.3%	
	3.00	60	77.6588	18.30585	23.6%	
	Total	175	77.2818	17.81721	23.1%	


Model Dimension^a^	
	Number of Levels	Covariance Structure	Number of Parameters	
Fixed Effects	Intercept	1		1	
	Treatment	2		1	
	Time	3		2	
	Treatment * Time	6		2	
Random Effects	Treatment + Time + Treatment * Time	11	Variance Components	3	
Residual			1	
Total	23		10	

a. Dependent Variable: Max_venti.	


Information Criteria^a^	
-2 Restricted Log Likelihood	1472.78760478	
Akaike's Information Criterion (AIC)	1480.78760478	
Hurvich and Tsai's Criterion (AICC)	1481.03150722	
Bozdogan's Criterion (CAIC)	1497.30719964	
Schwarz's Bayesian Criterion (BIC)	1493.30719964	

The information criteria are displayed in smaller-is-better form.^a^	
a. Dependent Variable: Max_venti.	


Coefficients of Determination	
Pseudo-R Square Measures	Marginal	.018	
	Conditional	.439	


Intraclass Correlation Coefficients	
Overall ICCs	Adjusted	.429	
	Conditional	.422	


Fixed Effects


Type III Tests of Fixed Effects^a^	
Source	Numerator df	Denominator df	F	Sig.	
Intercept	1	5.401	72.212	<.001	
Treatment	1	2.783	.031	.872	
Time	2	4.346	.073	.930	
Treatment * Time	2	4.037	.084	.921	

a. Dependent Variable: Max_venti.	


Estimates of Fixed Effects^a^	
Parameter	Estimate	Std. Error	df	t	Sig.	95% Confidence Interval	
						Lower Bound	
Intercept	74.587	15.766	9.345	4.731	<.001	39.122	
[Treatment=1.00]	6.144	18.250	5.147	.337	.750	-40.370	
[Treatment=2.00]	0^b^	0	.	.	.	.	
[Time=1.00]	6.027	18.250	6.797	.330	.751	-37.391	
[Time=2.00]	-2.193	18.282	6.845	-.120	.908	-45.623	
[Time=3.00]	0^b^	0	.	.	.	.	
[Treatment=1.00] * [Time=1.00]	-7.499	18.387	4.007	-.408	.704	-58.513	
[Treatment=1.00] * [Time=2.00]	-3.040	18.439	4.053	-.165	.877	-53.973	
[Treatment=1.00] * [Time=3.00]	0^b^	0	.	.	.	.	
[Treatment=2.00] * [Time=1.00]	0^b^	0	.	.	.	.	
[Treatment=2.00] * [Time=2.00]	0^b^	0	.	.	.	.	
[Treatment=2.00] * [Time=3.00]	0^b^	0	.	.	.	.	

Estimates of Fixed Effects^a^	
Parameter	95% Confidence Interval	
	Upper Bound	
Intercept	110.052	
[Treatment=1.00]	52.657	
[Treatment=2.00]	.	
[Time=1.00]	49.445	
[Time=2.00]	41.238	
[Time=3.00]	.	
[Treatment=1.00] * [Time=1.00]	43.516	
[Treatment=1.00] * [Time=2.00]	47.892	
[Treatment=1.00] * [Time=3.00]	.	
[Treatment=2.00] * [Time=1.00]	.	
[Treatment=2.00] * [Time=2.00]	.	
[Treatment=2.00] * [Time=3.00]	.	

a. Dependent Variable: Max_venti.	
b. This parameter is set to zero because it is redundant.	


Covariance Matrix for Estimates of Fixed Effects^a^	
Parameter	Intercept	[Treatment=1.00]	[Treatment=2.00]	[Time=1.00]	[Time=2.00]	
Intercept	248.560	-166.539	0^b^	-166.539	-166.539	
[Treatment=1.00]	-166.539	333.078	0^b^	84.518	84.518	
[Treatment=2.00]	0^b^	0^b^	0^b^	0^b^	0^b^	
[Time=1.00]	-166.539	84.518	0^b^	333.078	166.539	
[Time=2.00]	-166.539	84.518	0^b^	166.539	334.250	
[Time=3.00]	0^b^	0^b^	0^b^	0^b^	0^b^	
[Treatment=1.00] * [Time=1.00]	84.518	-169.036	0^b^	-169.036	-84.518	
[Treatment=1.00] * [Time=2.00]	84.518	-169.036	0^b^	-84.518	-170.208	
[Treatment=1.00] * [Time=3.00]	0^b^	0^b^	0^b^	0^b^	0^b^	
[Treatment=2.00] * [Time=1.00]	0^b^	0^b^	0^b^	0^b^	0^b^	
[Treatment=2.00] * [Time=2.00]	0^b^	0^b^	0^b^	0^b^	0^b^	
[Treatment=2.00] * [Time=3.00]	0^b^	0^b^	0^b^	0^b^	0^b^	

Covariance Matrix for Estimates of Fixed Effects^a^	
Parameter	[Time=3.00]	[Treatment=1.00] * [Time=1.00]	[Treatment=1.00] * [Time=2.00]	[Treatment=1.00] * [Time=3.00]	
Intercept	0^b^	84.518	84.518	0^b^	
[Treatment=1.00]	0^b^	-169.036	-169.036	0^b^	
[Treatment=2.00]	0^b^	0^b^	0^b^	0^b^	
[Time=1.00]	0^b^	-169.036	-84.518	0^b^	
[Time=2.00]	0^b^	-84.518	-170.208	0^b^	
[Time=3.00]	0^b^	0^b^	0^b^	0^b^	
[Treatment=1.00] * [Time=1.00]	0^b^	338.072	169.036	0^b^	
[Treatment=1.00] * [Time=2.00]	0^b^	169.036	339.997	0^b^	
[Treatment=1.00] * [Time=3.00]	0^b^	0^b^	0^b^	0^b^	
[Treatment=2.00] * [Time=1.00]	0^b^	0^b^	0^b^	0^b^	
[Treatment=2.00] * [Time=2.00]	0^b^	0^b^	0^b^	0^b^	
[Treatment=2.00] * [Time=3.00]	0^b^	0^b^	0^b^	0^b^	

Covariance Matrix for Estimates of Fixed Effects^a^	
Parameter	[Treatment=2.00] * [Time=1.00]	[Treatment=2.00] * [Time=2.00]	[Treatment=2.00] * [Time=3.00]	
Intercept	0^b^	0^b^	0^b^	
[Treatment=1.00]	0^b^	0^b^	0^b^	
[Treatment=2.00]	0^b^	0^b^	0^b^	
[Time=1.00]	0^b^	0^b^	0^b^	
[Time=2.00]	0^b^	0^b^	0^b^	
[Time=3.00]	0^b^	0^b^	0^b^	
[Treatment=1.00] * [Time=1.00]	0^b^	0^b^	0^b^	
[Treatment=1.00] * [Time=2.00]	0^b^	0^b^	0^b^	
[Treatment=1.00] * [Time=3.00]	0^b^	0^b^	0^b^	
[Treatment=2.00] * [Time=1.00]	0^b^	0^b^	0^b^	
[Treatment=2.00] * [Time=2.00]	0^b^	0^b^	0^b^	
[Treatment=2.00] * [Time=3.00]	0^b^	0^b^	0^b^	

a. Dependent Variable: Max_venti.	
b. The covariance is set to zero because it is associated with a redundant parameter.	


Covariance Parameters


Estimates of Covariance Parameters^a^	
Parameter	Estimate	Std. Error	
Residual	316.436	34.424	
Treatment	Variance	82.021	94.409	
Time	Variance	82.021	88.737	
Treatment * Time	Variance	73.970	59.722	

a. Dependent Variable: Max_venti.	


Covariance Matrix for Estimates of Covariance Parameters^a^	
Parameter	Residual	Treatment	Time	Treatment * Time	
		Variance	Variance	Variance	
Residual	1184.992	.002	-.003	-40.690	
Treatment	Variance	.002	8912.970	-1075.122	-850.855	
Time	Variance	-.003	-1075.122	7874.337	-1639.426	
Treatment * Time	Variance	-40.690	-850.855	-1639.426	3566.738	

a. Dependent Variable: Max_venti.	


Random Effects Covariance Structures (G)


Treatment^a^	
	[Treatment=1.00]	[Treatment=2.00]	
[Treatment=1.00]	82.021	0	
[Treatment=2.00]	0	82.021	

Variance Components^a^	
a. Dependent Variable: Max_venti.	


Time^a^	
	[Time=1.00]	[Time=2.00]	[Time=3.00]	
[Time=1.00]	82.021	0	0	
[Time=2.00]	0	82.021	0	
[Time=3.00]	0	0	82.021	

Variance Components^a^	
a. Dependent Variable: Max_venti.	


Treatment * Time^a^	
	[Treatment=1.00] * [Time=1.00]	[Treatment=1.00] * [Time=2.00]	[Treatment=1.00] * [Time=3.00]	[Treatment=2.00] * [Time=1.00]	
[Treatment=1.00] * [Time=1.00]	73.970	0	0	0	
[Treatment=1.00] * [Time=2.00]	0	73.970	0	0	
[Treatment=1.00] * [Time=3.00]	0	0	73.970	0	
[Treatment=2.00] * [Time=1.00]	0	0	0	73.970	
[Treatment=2.00] * [Time=2.00]	0	0	0	0	
[Treatment=2.00] * [Time=3.00]	0	0	0	0	

Treatment * Time^a^	
	[Treatment=2.00] * [Time=2.00]	[Treatment=2.00] * [Time=3.00]	
[Treatment=1.00] * [Time=1.00]	0	0	
[Treatment=1.00] * [Time=2.00]	0	0	
[Treatment=1.00] * [Time=3.00]	0	0	
[Treatment=2.00] * [Time=1.00]	0	0	
[Treatment=2.00] * [Time=2.00]	73.970	0	
[Treatment=2.00] * [Time=3.00]	0	73.970	

Variance Components^a^	
a. Dependent Variable: Max_venti.	


Estimated Marginal Means


1. Treatment


Estimates^a^	
Treatment	Mean	Std. Error	df	95% Confidence Interval	
				Lower Bound	Upper Bound	
1.00	78.496	11.731	4.437	47.153	109.838	
2.00	75.865	11.733	4.440	44.525	107.206	

a. Dependent Variable: Max_venti.	


Pairwise Comparisons^a^	
(I) Treatment	(J) Treatment	Mean Difference (I-J)	Std. Error	df	Sig.^b^	95% Confidence Interval for Difference^b^	
						Lower Bound	
1.00	2.00	2.631	14.853	2.783	.872	-46.787	
2.00	1.00	-2.631	14.853	2.783	.872	-52.049	

Pairwise Comparisons^a^	
(I) Treatment	(J) Treatment	95% Confidence Interval for Difference	
		Upper Bound	
1.00	2.00	52.049	
2.00	1.00	46.787	

Based on estimated marginal means^a^	
a. Dependent Variable: Max_venti.	
b. Adjustment for multiple comparisons: Bonferroni.	


Univariate Tests^a^	
Numerator df	Denominator df	F	Sig.	
1	2.783	.031	.872	

The F tests the effect of Treatment. This test is based on the linearly independent pairwise comparisons among the estimated marginal means.^a^	
a. Dependent Variable: Max_venti.	


2. Time


Estimates^a^	
Time	Mean	Std. Error	df	95% Confidence Interval	
				Lower Bound	Upper Bound	
1.00	79.937	12.857	6.957	49.498	110.375	
2.00	73.946	12.875	6.998	43.499	104.393	
3.00	77.659	12.857	6.957	47.220	108.098	

a. Dependent Variable: Max_venti.	


Pairwise Comparisons^a^	
(I) Time	(J) Time	Mean Difference (I-J)	Std. Error	df	Sig.^b^	95% Confidence Interval for Difference^b^	
						Lower Bound	
1.00	2.00	5.990	15.781	4.352	1.000	-53.655	
	3.00	2.278	15.766	4.335	1.000	-57.432	
2.00	1.00	-5.990	15.781	4.352	1.000	-65.636	
	3.00	-3.713	15.781	4.352	1.000	-63.358	
3.00	1.00	-2.278	15.766	4.335	1.000	-61.988	
	2.00	3.713	15.781	4.352	1.000	-55.933	

Pairwise Comparisons^a^	
(I) Time	(J) Time	95% Confidence Interval for Difference	
		Upper Bound	
1.00	2.00	65.636	
	3.00	61.988	
2.00	1.00	53.655	
	3.00	55.933	
3.00	1.00	57.432	
	2.00	63.358	

Based on estimated marginal means^a^	
a. Dependent Variable: Max_venti.	
b. Adjustment for multiple comparisons: Bonferroni.	


Univariate Tests^a^	
Numerator df	Denominator df	F	Sig.	
2	4.346	.073	.930	

The F tests the effect of Time. This test is based on the linearly independent pairwise comparisons among the estimated marginal means.^a^	
a. Dependent Variable: Max_venti.	


3. Treatment * Time^a^	
Treatment	Time	Mean	Std. Error	df	95% Confidence Interval	
					Lower Bound	Upper Bound	
1.00	1.00	79.259	15.766	9.345	43.794	114.724	
	2.00	75.498	15.790	9.402	40.011	110.985	
	3.00	80.731	15.766	9.345	45.266	116.195	
2.00	1.00	80.614	15.766	9.345	45.149	116.079	
	2.00	72.394	15.803	9.434	36.895	107.894	
	3.00	74.587	15.766	9.345	39.122	110.052	

a. Dependent Variable: Max_venti.	


Mixed Model Analysis


Notes	
Output Created	18-MAY-2026 15:38:10	
Comments		
Input	Active Dataset	DataSet0	
	Filter	<none>	
	Weight	<none>	
	Split File	<none>	
	N of Rows in Working Data File	180	
Missing Value Handling	Definition of Missing	User-defined missing values are treated as valid data.	
	Cases Used	Statistics are based on all cases with valid data for all variables in the model.	
Weight Handling	not applicable	
Syntax	MIXED Vo2_peak BY Treatment Time
  /CRITERIA=DFMETHOD(SATTERTHWAITE) CIN(95) MXITER(100) MXSTEP(10) SCORING(1)
    SINGULAR(0.000000000001) HCONVERGE(0.00000001, RELATIVE) LCONVERGE(0, ABSOLUTE) PCONVERGE(0,
    ABSOLUTE)
  /FIXED=Treatment Time Treatment*Time | SSTYPE(3)
  /METHOD=REML
  /PRINT=COVB DESCRIPTIVES G  SOLUTION
  /RANDOM=Treatment Time Treatment*Time | COVTYPE(VC)
  /EMMEANS=TABLES(Treatment) COMPARE ADJ(BONFERRONI)
  /EMMEANS=TABLES(Time) COMPARE ADJ(BONFERRONI)
  /EMMEANS=TABLES(Treatment*Time) .	
Resources	Processor Time	00:00:00.05	
	Elapsed Time	00:00:00.04	


Descriptive Statistics	
Vo2_peak  	
Treatment	Time	Count	Mean	Standard Deviation	Coefficient of Variation	
1.00	1.00	30	28.0777	5.44558	19.4%	
	2.00	28	28.0729	5.40289	19.2%	
	3.00	30	29.2233	5.17864	17.7%	
	Total	88	28.4667	5.30877	18.6%	
2.00	1.00	30	27.7159	5.36420	19.4%	
	2.00	27	27.2215	5.38954	19.8%	
	3.00	30	28.0877	5.57468	19.8%	
	Total	87	27.6907	5.39338	19.5%	
Total	1.00	60	27.8968	5.36215	19.2%	
	2.00	55	27.6549	5.36337	19.4%	
	3.00	60	28.6555	5.36516	18.7%	
	Total	175	28.0809	5.34977	19.1%	


Model Dimension^a^	
	Number of Levels	Covariance Structure	Number of Parameters	
Fixed Effects	Intercept	1		1	
	Treatment	2		1	
	Time	3		2	
	Treatment * Time	6		2	
Random Effects	Treatment + Time + Treatment * Time	11	Variance Components	3	
Residual			1	
Total	23		10	

a. Dependent Variable: Vo2_peak.	


Information Criteria^a^	
-2 Restricted Log Likelihood	1069.45081799	
Akaike's Information Criterion (AIC)	1077.45081799	
Hurvich and Tsai's Criterion (AICC)	1077.69472043	
Bozdogan's Criterion (CAIC)	1093.97041285	
Schwarz's Bayesian Criterion (BIC)	1089.97041285	

The information criteria are displayed in smaller-is-better form.^a^	
a. Dependent Variable: Vo2_peak.	


Coefficients of Determination	
Pseudo-R Square Measures	Marginal	.007	
	Conditional	.433	


Intraclass Correlation Coefficients	
Overall ICCs	Adjusted	.429	
	Conditional	.426	


Fixed Effects


Type III Tests of Fixed Effects^a^	
Source	Numerator df	Denominator df	F	Sig.	
Intercept	1	5.401	103.863	<.001	
Treatment	1	2.783	.030	.874	
Time	2	4.346	.024	.976	
Treatment * Time	2	4.037	.010	.990	

a. Dependent Variable: Vo2_peak.	


Estimates of Fixed Effects^a^	
Parameter	Estimate	Std. Error	df	t	Sig.	95% Confidence Interval	
						Lower Bound	
Intercept	28.088	4.780	9.345	5.876	<.001	17.334	
[Treatment=1.00]	1.136	5.534	5.147	.205	.845	-12.968	
[Treatment=2.00]	0^b^	0	.	.	.	.	
[Time=1.00]	-.372	5.534	6.797	-.067	.948	-13.537	
[Time=2.00]	-.866	5.544	6.845	-.156	.880	-14.035	
[Time=3.00]	0^b^	0	.	.	.	.	
[Treatment=1.00] * [Time=1.00]	-.774	5.575	4.007	-.139	.896	-16.242	
[Treatment=1.00] * [Time=2.00]	-.284	5.591	4.053	-.051	.962	-15.728	
[Treatment=1.00] * [Time=3.00]	0^b^	0	.	.	.	.	
[Treatment=2.00] * [Time=1.00]	0^b^	0	.	.	.	.	
[Treatment=2.00] * [Time=2.00]	0^b^	0	.	.	.	.	
[Treatment=2.00] * [Time=3.00]	0^b^	0	.	.	.	.	

Estimates of Fixed Effects^a^	
Parameter	95% Confidence Interval	
	Upper Bound	
Intercept	38.841	
[Treatment=1.00]	15.239	
[Treatment=2.00]	.	
[Time=1.00]	12.793	
[Time=2.00]	12.303	
[Time=3.00]	.	
[Treatment=1.00] * [Time=1.00]	14.695	
[Treatment=1.00] * [Time=2.00]	15.159	
[Treatment=1.00] * [Time=3.00]	.	
[Treatment=2.00] * [Time=1.00]	.	
[Treatment=2.00] * [Time=2.00]	.	
[Treatment=2.00] * [Time=3.00]	.	

a. Dependent Variable: Vo2_peak.	
b. This parameter is set to zero because it is redundant.	


Covariance Matrix for Estimates of Fixed Effects^a^	
Parameter	Intercept	[Treatment=1.00]	[Treatment=2.00]	[Time=1.00]	[Time=2.00]	
Intercept	22.853	-15.312	0^b^	-15.312	-15.312	
[Treatment=1.00]	-15.312	30.624	0^b^	7.771	7.771	
[Treatment=2.00]	0^b^	0^b^	0^b^	0^b^	0^b^	
[Time=1.00]	-15.312	7.771	0^b^	30.623	15.312	
[Time=2.00]	-15.312	7.771	0^b^	15.312	30.731	
[Time=3.00]	0^b^	0^b^	0^b^	0^b^	0^b^	
[Treatment=1.00] * [Time=1.00]	7.771	-15.541	0^b^	-15.541	-7.771	
[Treatment=1.00] * [Time=2.00]	7.771	-15.541	0^b^	-7.771	-15.649	
[Treatment=1.00] * [Time=3.00]	0^b^	0^b^	0^b^	0^b^	0^b^	
[Treatment=2.00] * [Time=1.00]	0^b^	0^b^	0^b^	0^b^	0^b^	
[Treatment=2.00] * [Time=2.00]	0^b^	0^b^	0^b^	0^b^	0^b^	
[Treatment=2.00] * [Time=3.00]	0^b^	0^b^	0^b^	0^b^	0^b^	

Covariance Matrix for Estimates of Fixed Effects^a^	
Parameter	[Time=3.00]	[Treatment=1.00] * [Time=1.00]	[Treatment=1.00] * [Time=2.00]	[Treatment=1.00] * [Time=3.00]	
Intercept	0^b^	7.771	7.771	0^b^	
[Treatment=1.00]	0^b^	-15.541	-15.541	0^b^	
[Treatment=2.00]	0^b^	0^b^	0^b^	0^b^	
[Time=1.00]	0^b^	-15.541	-7.771	0^b^	
[Time=2.00]	0^b^	-7.771	-15.649	0^b^	
[Time=3.00]	0^b^	0^b^	0^b^	0^b^	
[Treatment=1.00] * [Time=1.00]	0^b^	31.083	15.541	0^b^	
[Treatment=1.00] * [Time=2.00]	0^b^	15.541	31.260	0^b^	
[Treatment=1.00] * [Time=3.00]	0^b^	0^b^	0^b^	0^b^	
[Treatment=2.00] * [Time=1.00]	0^b^	0^b^	0^b^	0^b^	
[Treatment=2.00] * [Time=2.00]	0^b^	0^b^	0^b^	0^b^	
[Treatment=2.00] * [Time=3.00]	0^b^	0^b^	0^b^	0^b^	

Covariance Matrix for Estimates of Fixed Effects^a^	
Parameter	[Treatment=2.00] * [Time=1.00]	[Treatment=2.00] * [Time=2.00]	[Treatment=2.00] * [Time=3.00]	
Intercept	0^b^	0^b^	0^b^	
[Treatment=1.00]	0^b^	0^b^	0^b^	
[Treatment=2.00]	0^b^	0^b^	0^b^	
[Time=1.00]	0^b^	0^b^	0^b^	
[Time=2.00]	0^b^	0^b^	0^b^	
[Time=3.00]	0^b^	0^b^	0^b^	
[Treatment=1.00] * [Time=1.00]	0^b^	0^b^	0^b^	
[Treatment=1.00] * [Time=2.00]	0^b^	0^b^	0^b^	
[Treatment=1.00] * [Time=3.00]	0^b^	0^b^	0^b^	
[Treatment=2.00] * [Time=1.00]	0^b^	0^b^	0^b^	
[Treatment=2.00] * [Time=2.00]	0^b^	0^b^	0^b^	
[Treatment=2.00] * [Time=3.00]	0^b^	0^b^	0^b^	

a. Dependent Variable: Vo2_peak.	
b. The covariance is set to zero because it is associated with a redundant parameter.	


Covariance Parameters


Estimates of Covariance Parameters^a^	
Parameter	Estimate	Std. Error	
Residual	29.093	3.165	
Treatment	Variance	7.541	8.680	
Time	Variance	7.541	8.159	
Treatment * Time	Variance	6.801	5.491	

a. Dependent Variable: Vo2_peak.	


Covariance Matrix for Estimates of Covariance Parameters^a^	
Parameter	Residual	Treatment	Time	Treatment * Time	
		Variance	Variance	Variance	
Residual	10.017	1.461E-5	-2.792E-5	-.344	
Treatment	Variance	1.461E-5	75.343	-9.088	-7.192	
Time	Variance	-2.792E-5	-9.088	66.563	-13.858	
Treatment * Time	Variance	-.344	-7.192	-13.858	30.150	

a. Dependent Variable: Vo2_peak.	


Random Effects Covariance Structures (G)


Treatment^a^	
	[Treatment=1.00]	[Treatment=2.00]	
[Treatment=1.00]	7.541	0	
[Treatment=2.00]	0	7.541	

Variance Components^a^	
a. Dependent Variable: Vo2_peak.	


Time^a^	
	[Time=1.00]	[Time=2.00]	[Time=3.00]	
[Time=1.00]	7.541	0	0	
[Time=2.00]	0	7.541	0	
[Time=3.00]	0	0	7.541	

Variance Components^a^	
a. Dependent Variable: Vo2_peak.	


Treatment * Time^a^	
	[Treatment=1.00] * [Time=1.00]	[Treatment=1.00] * [Time=2.00]	[Treatment=1.00] * [Time=3.00]	[Treatment=2.00] * [Time=1.00]	
[Treatment=1.00] * [Time=1.00]	6.801	0	0	0	
[Treatment=1.00] * [Time=2.00]	0	6.801	0	0	
[Treatment=1.00] * [Time=3.00]	0	0	6.801	0	
[Treatment=2.00] * [Time=1.00]	0	0	0	6.801	
[Treatment=2.00] * [Time=2.00]	0	0	0	0	
[Treatment=2.00] * [Time=3.00]	0	0	0	0	

Treatment * Time^a^	
	[Treatment=2.00] * [Time=2.00]	[Treatment=2.00] * [Time=3.00]	
[Treatment=1.00] * [Time=1.00]	0	0	
[Treatment=1.00] * [Time=2.00]	0	0	
[Treatment=1.00] * [Time=3.00]	0	0	
[Treatment=2.00] * [Time=1.00]	0	0	
[Treatment=2.00] * [Time=2.00]	6.801	0	
[Treatment=2.00] * [Time=3.00]	0	6.801	

Variance Components^a^	
a. Dependent Variable: Vo2_peak.	


Estimated Marginal Means


1. Treatment


Estimates^a^	
Treatment	Mean	Std. Error	df	95% Confidence Interval	
				Lower Bound	Upper Bound	
1.00	28.458	3.557	4.437	18.954	37.962	
2.00	27.675	3.558	4.440	18.172	37.178	

a. Dependent Variable: Vo2_peak.	


Pairwise Comparisons^a^	
(I) Treatment	(J) Treatment	Mean Difference (I-J)	Std. Error	df	Sig.^b^	95% Confidence Interval for Difference^b^	
						Lower Bound	
1.00	2.00	.783	4.504	2.783	.874	-14.201	
2.00	1.00	-.783	4.504	2.783	.874	-15.767	

Pairwise Comparisons^a^	
(I) Treatment	(J) Treatment	95% Confidence Interval for Difference	
		Upper Bound	
1.00	2.00	15.767	
2.00	1.00	14.201	

Based on estimated marginal means^a^	
a. Dependent Variable: Vo2_peak.	
b. Adjustment for multiple comparisons: Bonferroni.	


Univariate Tests^a^	
Numerator df	Denominator df	F	Sig.	
1	2.783	.030	.874	

The F tests the effect of Treatment. This test is based on the linearly independent pairwise comparisons among the estimated marginal means.^a^	
a. Dependent Variable: Vo2_peak.	


2. Time


Estimates^a^	
Time	Mean	Std. Error	df	95% Confidence Interval	
				Lower Bound	Upper Bound	
1.00	27.897	3.898	6.957	18.667	37.126	
2.00	27.647	3.904	6.998	18.415	36.879	
3.00	28.656	3.898	6.957	19.426	37.885	

a. Dependent Variable: Vo2_peak.	


Pairwise Comparisons^a^	
(I) Time	(J) Time	Mean Difference (I-J)	Std. Error	df	Sig.^b^	95% Confidence Interval for Difference^b^	
						Lower Bound	
1.00	2.00	.250	4.785	4.352	1.000	-17.836	
	3.00	-.759	4.780	4.335	1.000	-18.864	
2.00	1.00	-.250	4.785	4.352	1.000	-18.335	
	3.00	-1.008	4.785	4.352	1.000	-19.094	
3.00	1.00	.759	4.780	4.335	1.000	-17.346	
	2.00	1.008	4.785	4.352	1.000	-17.077	

Pairwise Comparisons^a^	
(I) Time	(J) Time	95% Confidence Interval for Difference	
		Upper Bound	
1.00	2.00	18.335	
	3.00	17.346	
2.00	1.00	17.836	
	3.00	17.077	
3.00	1.00	18.864	
	2.00	19.094	

Based on estimated marginal means^a^	
a. Dependent Variable: Vo2_peak.	
b. Adjustment for multiple comparisons: Bonferroni.	


Univariate Tests^a^	
Numerator df	Denominator df	F	Sig.	
2	4.346	.024	.976	

The F tests the effect of Time. This test is based on the linearly independent pairwise comparisons among the estimated marginal means.^a^	
a. Dependent Variable: Vo2_peak.	


3. Treatment * Time^a^	
Treatment	Time	Mean	Std. Error	df	95% Confidence Interval	
					Lower Bound	Upper Bound	
1.00	1.00	28.078	4.780	9.345	17.324	38.831	
	2.00	28.073	4.788	9.402	17.313	38.833	
	3.00	29.223	4.780	9.345	18.470	39.977	
2.00	1.00	27.716	4.780	9.345	16.962	38.469	
	2.00	27.221	4.792	9.434	16.457	37.986	
	3.00	28.088	4.780	9.345	17.334	38.841	

a. Dependent Variable: Vo2_peak.	


Mixed Model Analysis


Notes	
Output Created	18-MAY-2026 15:38:26	
Comments		
Input	Active Dataset	DataSet0	
	Filter	<none>	
	Weight	<none>	
	Split File	<none>	
	N of Rows in Working Data File	180	
Missing Value Handling	Definition of Missing	User-defined missing values are treated as valid data.	
	Cases Used	Statistics are based on all cases with valid data for all variables in the model.	
Weight Handling	not applicable	
Syntax	MIXED Vent_threshold BY Treatment Time
  /CRITERIA=DFMETHOD(SATTERTHWAITE) CIN(95) MXITER(100) MXSTEP(10) SCORING(1)
    SINGULAR(0.000000000001) HCONVERGE(0.00000001, RELATIVE) LCONVERGE(0, ABSOLUTE) PCONVERGE(0,
    ABSOLUTE)
  /FIXED=Treatment Time Treatment*Time | SSTYPE(3)
  /METHOD=REML
  /PRINT=COVB DESCRIPTIVES G  SOLUTION
  /RANDOM=Treatment Time Treatment*Time | COVTYPE(VC)
  /EMMEANS=TABLES(Treatment) COMPARE ADJ(BONFERRONI)
  /EMMEANS=TABLES(Time) COMPARE ADJ(BONFERRONI)
  /EMMEANS=TABLES(Treatment*Time) .	
Resources	Processor Time	00:00:00.03	
	Elapsed Time	00:00:00.04	


Descriptive Statistics	
Vent_threshold  	
Treatment	Time	Count	Mean	Standard Deviation	Coefficient of Variation	
1.00	1.00	30	1.8090	.38468	21.3%	
	2.00	28	1.7750	.34723	19.6%	
	3.00	30	1.9187	.39162	20.4%	
	Total	88	1.8356	.37639	20.5%	
2.00	1.00	30	1.7767	.37701	21.2%	
	2.00	27	1.7267	.35335	20.5%	
	3.00	30	1.7363	.37604	21.7%	
	Total	87	1.7472	.36583	20.9%	
Total	1.00	60	1.7928	.37797	21.1%	
	2.00	55	1.7513	.34784	19.9%	
	3.00	60	1.8275	.39158	21.4%	
	Total	175	1.7917	.37275	20.8%	


Model Dimension^a^	
	Number of Levels	Covariance Structure	Number of Parameters	
Fixed Effects	Intercept	1		1	
	Treatment	2		1	
	Time	3		2	
	Treatment * Time	6		2	
Random Effects	Treatment + Time + Treatment * Time	11	Variance Components	3	
Residual			1	
Total	23		10	

a. Dependent Variable: Vent_threshold.	


Information Criteria^a^	
-2 Restricted Log Likelihood	166.14852491	
Akaike's Information Criterion (AIC)	174.14852491	
Hurvich and Tsai's Criterion (AICC)	174.39242735	
Bozdogan's Criterion (CAIC)	190.66811977	
Schwarz's Bayesian Criterion (BIC)	186.66811977	

The information criteria are displayed in smaller-is-better form.^a^	
a. Dependent Variable: Vent_threshold.	


Coefficients of Determination	
Pseudo-R Square Measures	Marginal	.017	
	Conditional	.439	


Intraclass Correlation Coefficients	
Overall ICCs	Adjusted	.429	
	Conditional	.422	


Fixed Effects


Type III Tests of Fixed Effects^a^	
Source	Numerator df	Denominator df	F	Sig.	
Intercept	1	5.401	88.568	<.001	
Treatment	1	2.783	.079	.798	
Time	2	4.346	.027	.974	
Treatment * Time	2	4.037	.091	.915	

a. Dependent Variable: Vent_threshold.	


Estimates of Fixed Effects^a^	
Parameter	Estimate	Std. Error	df	t	Sig.	95% Confidence Interval	
						Lower Bound	
Intercept	1.736	.330	9.345	5.258	<.001	.993	
[Treatment=1.00]	.182	.382	5.147	.477	.653	-.792	
[Treatment=2.00]	0^b^	0	.	.	.	.	
[Time=1.00]	.040	.382	6.797	.106	.919	-.869	
[Time=2.00]	-.010	.383	6.845	-.025	.981	-.919	
[Time=3.00]	0^b^	0	.	.	.	.	
[Treatment=1.00] * [Time=1.00]	-.150	.385	4.007	-.389	.717	-1.219	
[Treatment=1.00] * [Time=2.00]	-.134	.386	4.053	-.347	.746	-1.201	
[Treatment=1.00] * [Time=3.00]	0^b^	0	.	.	.	.	
[Treatment=2.00] * [Time=1.00]	0^b^	0	.	.	.	.	
[Treatment=2.00] * [Time=2.00]	0^b^	0	.	.	.	.	
[Treatment=2.00] * [Time=3.00]	0^b^	0	.	.	.	.	

Estimates of Fixed Effects^a^	
Parameter	95% Confidence Interval	
	Upper Bound	
Intercept	2.479	
[Treatment=1.00]	1.157	
[Treatment=2.00]	.	
[Time=1.00]	.950	
[Time=2.00]	.900	
[Time=3.00]	.	
[Treatment=1.00] * [Time=1.00]	.919	
[Treatment=1.00] * [Time=2.00]	.933	
[Treatment=1.00] * [Time=3.00]	.	
[Treatment=2.00] * [Time=1.00]	.	
[Treatment=2.00] * [Time=2.00]	.	
[Treatment=2.00] * [Time=3.00]	.	

a. Dependent Variable: Vent_threshold.	
b. This parameter is set to zero because it is redundant.	


Covariance Matrix for Estimates of Fixed Effects^a^	
Parameter	Intercept	[Treatment=1.00]	[Treatment=2.00]	[Time=1.00]	[Time=2.00]	
Intercept	.109	-.073	0^b^	-.073	-.073	
[Treatment=1.00]	-.073	.146	0^b^	.037	.037	
[Treatment=2.00]	0^b^	0^b^	0^b^	0^b^	0^b^	
[Time=1.00]	-.073	.037	0^b^	.146	.073	
[Time=2.00]	-.073	.037	0^b^	.073	.147	
[Time=3.00]	0^b^	0^b^	0^b^	0^b^	0^b^	
[Treatment=1.00] * [Time=1.00]	.037	-.074	0^b^	-.074	-.037	
[Treatment=1.00] * [Time=2.00]	.037	-.074	0^b^	-.037	-.075	
[Treatment=1.00] * [Time=3.00]	0^b^	0^b^	0^b^	0^b^	0^b^	
[Treatment=2.00] * [Time=1.00]	0^b^	0^b^	0^b^	0^b^	0^b^	
[Treatment=2.00] * [Time=2.00]	0^b^	0^b^	0^b^	0^b^	0^b^	
[Treatment=2.00] * [Time=3.00]	0^b^	0^b^	0^b^	0^b^	0^b^	

Covariance Matrix for Estimates of Fixed Effects^a^	
Parameter	[Time=3.00]	[Treatment=1.00] * [Time=1.00]	[Treatment=1.00] * [Time=2.00]	[Treatment=1.00] * [Time=3.00]	
Intercept	0^b^	.037	.037	0^b^	
[Treatment=1.00]	0^b^	-.074	-.074	0^b^	
[Treatment=2.00]	0^b^	0^b^	0^b^	0^b^	
[Time=1.00]	0^b^	-.074	-.037	0^b^	
[Time=2.00]	0^b^	-.037	-.075	0^b^	
[Time=3.00]	0^b^	0^b^	0^b^	0^b^	
[Treatment=1.00] * [Time=1.00]	0^b^	.148	.074	0^b^	
[Treatment=1.00] * [Time=2.00]	0^b^	.074	.149	0^b^	
[Treatment=1.00] * [Time=3.00]	0^b^	0^b^	0^b^	0^b^	
[Treatment=2.00] * [Time=1.00]	0^b^	0^b^	0^b^	0^b^	
[Treatment=2.00] * [Time=2.00]	0^b^	0^b^	0^b^	0^b^	
[Treatment=2.00] * [Time=3.00]	0^b^	0^b^	0^b^	0^b^	

Covariance Matrix for Estimates of Fixed Effects^a^	
Parameter	[Treatment=2.00] * [Time=1.00]	[Treatment=2.00] * [Time=2.00]	[Treatment=2.00] * [Time=3.00]	
Intercept	0^b^	0^b^	0^b^	
[Treatment=1.00]	0^b^	0^b^	0^b^	
[Treatment=2.00]	0^b^	0^b^	0^b^	
[Time=1.00]	0^b^	0^b^	0^b^	
[Time=2.00]	0^b^	0^b^	0^b^	
[Time=3.00]	0^b^	0^b^	0^b^	
[Treatment=1.00] * [Time=1.00]	0^b^	0^b^	0^b^	
[Treatment=1.00] * [Time=2.00]	0^b^	0^b^	0^b^	
[Treatment=1.00] * [Time=3.00]	0^b^	0^b^	0^b^	
[Treatment=2.00] * [Time=1.00]	0^b^	0^b^	0^b^	
[Treatment=2.00] * [Time=2.00]	0^b^	0^b^	0^b^	
[Treatment=2.00] * [Time=3.00]	0^b^	0^b^	0^b^	

a. Dependent Variable: Vent_threshold.	
b. The covariance is set to zero because it is associated with a redundant parameter.	


Covariance Parameters


Estimates of Covariance Parameters^a^	
Parameter	Estimate	Std. Error	
Residual	.139	.015	
Treatment	Variance	.036	.041	
Time	Variance	.036	.039	
Treatment * Time	Variance	.032	.026	

a. Dependent Variable: Vent_threshold.	


Covariance Matrix for Estimates of Covariance Parameters^a^	
Parameter	Residual	Treatment	Time	Treatment * Time	
		Variance	Variance	Variance	
Residual	.000	3.326E-10	-6.357E-10	-7.833E-6	
Treatment	Variance	3.326E-10	.002	.000	.000	
Time	Variance	-6.357E-10	.000	.002	.000	
Treatment * Time	Variance	-7.833E-6	.000	.000	.001	

a. Dependent Variable: Vent_threshold.	


Random Effects Covariance Structures (G)


Treatment^a^	
	[Treatment=1.00]	[Treatment=2.00]	
[Treatment=1.00]	.036	0	
[Treatment=2.00]	0	.036	

Variance Components^a^	
a. Dependent Variable: Vent_threshold.	


Time^a^	
	[Time=1.00]	[Time=2.00]	[Time=3.00]	
[Time=1.00]	.036	0	0	
[Time=2.00]	0	.036	0	
[Time=3.00]	0	0	.036	

Variance Components^a^	
a. Dependent Variable: Vent_threshold.	


Treatment * Time^a^	
	[Treatment=1.00] * [Time=1.00]	[Treatment=1.00] * [Time=2.00]	[Treatment=1.00] * [Time=3.00]	[Treatment=2.00] * [Time=1.00]	
[Treatment=1.00] * [Time=1.00]	.032	0	0	0	
[Treatment=1.00] * [Time=2.00]	0	.032	0	0	
[Treatment=1.00] * [Time=3.00]	0	0	.032	0	
[Treatment=2.00] * [Time=1.00]	0	0	0	.032	
[Treatment=2.00] * [Time=2.00]	0	0	0	0	
[Treatment=2.00] * [Time=3.00]	0	0	0	0	

Treatment * Time^a^	
	[Treatment=2.00] * [Time=2.00]	[Treatment=2.00] * [Time=3.00]	
[Treatment=1.00] * [Time=1.00]	0	0	
[Treatment=1.00] * [Time=2.00]	0	0	
[Treatment=1.00] * [Time=3.00]	0	0	
[Treatment=2.00] * [Time=1.00]	0	0	
[Treatment=2.00] * [Time=2.00]	.032	0	
[Treatment=2.00] * [Time=3.00]	0	.032	

Variance Components^a^	
a. Dependent Variable: Vent_threshold.	


Estimated Marginal Means


1. Treatment


Estimates^a^	
Treatment	Mean	Std. Error	df	95% Confidence Interval	
				Lower Bound	Upper Bound	
1.00	1.834	.246	4.437	1.178	2.491	
2.00	1.747	.246	4.440	1.090	2.403	

a. Dependent Variable: Vent_threshold.	


Pairwise Comparisons^a^	
(I) Treatment	(J) Treatment	Mean Difference (I-J)	Std. Error	df	Sig.^b^	95% Confidence Interval for Difference^b^	
						Lower Bound	
1.00	2.00	.088	.311	2.783	.798	-.947	
2.00	1.00	-.088	.311	2.783	.798	-1.123	

Pairwise Comparisons^a^	
(I) Treatment	(J) Treatment	95% Confidence Interval for Difference	
		Upper Bound	
1.00	2.00	1.123	
2.00	1.00	.947	

Based on estimated marginal means^a^	
a. Dependent Variable: Vent_threshold.	
b. Adjustment for multiple comparisons: Bonferroni.	


Univariate Tests^a^	
Numerator df	Denominator df	F	Sig.	
1	2.783	.079	.798	

The F tests the effect of Treatment. This test is based on the linearly independent pairwise comparisons among the estimated marginal means.^a^	
a. Dependent Variable: Vent_threshold.	


2. Time


Estimates^a^	
Time	Mean	Std. Error	df	95% Confidence Interval	
				Lower Bound	Upper Bound	
1.00	1.793	.269	6.957	1.155	2.430	
2.00	1.751	.270	6.998	1.113	2.389	
3.00	1.827	.269	6.957	1.190	2.465	

a. Dependent Variable: Vent_threshold.	


Pairwise Comparisons^a^	
(I) Time	(J) Time	Mean Difference (I-J)	Std. Error	df	Sig.^b^	95% Confidence Interval for Difference^b^	
						Lower Bound	
1.00	2.00	.042	.331	4.352	1.000	-1.207	
	3.00	-.035	.330	4.335	1.000	-1.285	
2.00	1.00	-.042	.331	4.352	1.000	-1.291	
	3.00	-.077	.331	4.352	1.000	-1.326	
3.00	1.00	.035	.330	4.335	1.000	-1.216	
	2.00	.077	.331	4.352	1.000	-1.173	

Pairwise Comparisons^a^	
(I) Time	(J) Time	95% Confidence Interval for Difference	
		Upper Bound	
1.00	2.00	1.291	
	3.00	1.216	
2.00	1.00	1.207	
	3.00	1.173	
3.00	1.00	1.285	
	2.00	1.326	

Based on estimated marginal means^a^	
a. Dependent Variable: Vent_threshold.	
b. Adjustment for multiple comparisons: Bonferroni.	


Univariate Tests^a^	
Numerator df	Denominator df	F	Sig.	
2	4.346	.027	.974	

The F tests the effect of Time. This test is based on the linearly independent pairwise comparisons among the estimated marginal means.^a^	
a. Dependent Variable: Vent_threshold.	


3. Treatment * Time^a^	
Treatment	Time	Mean	Std. Error	df	95% Confidence Interval	
					Lower Bound	Upper Bound	
1.00	1.00	1.809	.330	9.345	1.066	2.552	
	2.00	1.775	.331	9.402	1.032	2.518	
	3.00	1.919	.330	9.345	1.176	2.662	
2.00	1.00	1.777	.330	9.345	1.034	2.520	
	2.00	1.727	.331	9.434	.983	2.470	
	3.00	1.736	.330	9.345	.993	2.479	

a. Dependent Variable: Vent_threshold.	


Mixed Model Analysis


Notes	
Output Created	18-MAY-2026 15:38:38	
Comments		
Input	Active Dataset	DataSet0	
	Filter	<none>	
	Weight	<none>	
	Split File	<none>	
	N of Rows in Working Data File	180	
Missing Value Handling	Definition of Missing	User-defined missing values are treated as valid data.	
	Cases Used	Statistics are based on all cases with valid data for all variables in the model.	
Weight Handling	not applicable	
Syntax	MIXED Vo2_85 BY Treatment Time
  /CRITERIA=DFMETHOD(SATTERTHWAITE) CIN(95) MXITER(100) MXSTEP(10) SCORING(1)
    SINGULAR(0.000000000001) HCONVERGE(0.00000001, RELATIVE) LCONVERGE(0, ABSOLUTE) PCONVERGE(0,
    ABSOLUTE)
  /FIXED=Treatment Time Treatment*Time | SSTYPE(3)
  /METHOD=REML
  /PRINT=COVB DESCRIPTIVES G  SOLUTION
  /RANDOM=Treatment Time Treatment*Time | COVTYPE(VC)
  /EMMEANS=TABLES(Treatment) COMPARE ADJ(BONFERRONI)
  /EMMEANS=TABLES(Time) COMPARE ADJ(BONFERRONI)
  /EMMEANS=TABLES(Treatment*Time) .	
Resources	Processor Time	00:00:00.03	
	Elapsed Time	00:00:00.03	


Descriptive Statistics	
Vo2_85  	
Treatment	Time	Count	Mean	Standard Deviation	Coefficient of Variation	
1.00	1.00	30	1.7819	.34957	19.6%	
	2.00	28	1.8050	.34008	18.8%	
	3.00	30	1.8958	.36235	19.1%	
	Total	88	1.8281	.35054	19.2%	
2.00	1.00	30	1.7742	.37894	21.4%	
	2.00	27	1.7412	.36159	20.8%	
	3.00	30	1.8023	.37874	21.0%	
	Total	87	1.7737	.37005	20.9%	
Total	1.00	60	1.7781	.36147	20.3%	
	2.00	55	1.7737	.34902	19.7%	
	3.00	60	1.8490	.37049	20.0%	
	Total	175	1.8010	.36037	20.0%	


Model Dimension^a^	
	Number of Levels	Covariance Structure	Number of Parameters	
Fixed Effects	Intercept	1		1	
	Treatment	2		1	
	Time	3		2	
	Treatment * Time	6		2	
Random Effects	Treatment + Time + Treatment * Time	11	Variance Components	3	
Residual			1	
Total	23		10	

a. Dependent Variable: Vo2_85.	


Information Criteria^a^	
-2 Restricted Log Likelihood	156.77815938	
Akaike's Information Criterion (AIC)	164.77815938	
Hurvich and Tsai's Criterion (AICC)	165.02206182	
Bozdogan's Criterion (CAIC)	181.29775424	
Schwarz's Bayesian Criterion (BIC)	177.29775424	

The information criteria are displayed in smaller-is-better form.^a^	
a. Dependent Variable: Vo2_85.	


Coefficients of Determination	
Pseudo-R Square Measures	Marginal	.010	
	Conditional	.435	


Intraclass Correlation Coefficients	
Overall ICCs	Adjusted	.429	
	Conditional	.425	


Fixed Effects


Type III Tests of Fixed Effects^a^	
Source	Numerator df	Denominator df	F	Sig.	
Intercept	1	5.401	94.633	<.001	
Treatment	1	2.783	.033	.868	
Time	2	4.346	.035	.966	
Treatment * Time	2	4.037	.027	.973	

a. Dependent Variable: Vo2_85.	


Estimates of Fixed Effects^a^	
Parameter	Estimate	Std. Error	df	t	Sig.	95% Confidence Interval	
						Lower Bound	
Intercept	1.802	.321	9.345	5.611	<.001	1.080	
[Treatment=1.00]	.094	.372	5.147	.251	.811	-.854	
[Treatment=2.00]	0^b^	0	.	.	.	.	
[Time=1.00]	-.028	.372	6.797	-.075	.942	-.913	
[Time=2.00]	-.061	.372	6.845	-.164	.875	-.946	
[Time=3.00]	0^b^	0	.	.	.	.	
[Treatment=1.00] * [Time=1.00]	-.086	.375	4.007	-.229	.830	-1.125	
[Treatment=1.00] * [Time=2.00]	-.030	.376	4.053	-.079	.941	-1.067	
[Treatment=1.00] * [Time=3.00]	0^b^	0	.	.	.	.	
[Treatment=2.00] * [Time=1.00]	0^b^	0	.	.	.	.	
[Treatment=2.00] * [Time=2.00]	0^b^	0	.	.	.	.	
[Treatment=2.00] * [Time=3.00]	0^b^	0	.	.	.	.	

Estimates of Fixed Effects^a^	
Parameter	95% Confidence Interval	
	Upper Bound	
Intercept	2.525	
[Treatment=1.00]	1.041	
[Treatment=2.00]	.	
[Time=1.00]	.857	
[Time=2.00]	.824	
[Time=3.00]	.	
[Treatment=1.00] * [Time=1.00]	.953	
[Treatment=1.00] * [Time=2.00]	1.008	
[Treatment=1.00] * [Time=3.00]	.	
[Treatment=2.00] * [Time=1.00]	.	
[Treatment=2.00] * [Time=2.00]	.	
[Treatment=2.00] * [Time=3.00]	.	

a. Dependent Variable: Vo2_85.	
b. This parameter is set to zero because it is redundant.	


Covariance Matrix for Estimates of Fixed Effects^a^	
Parameter	Intercept	[Treatment=1.00]	[Treatment=2.00]	[Time=1.00]	[Time=2.00]	
Intercept	.103	-.069	0^b^	-.069	-.069	
[Treatment=1.00]	-.069	.138	0^b^	.035	.035	
[Treatment=2.00]	0^b^	0^b^	0^b^	0^b^	0^b^	
[Time=1.00]	-.069	.035	0^b^	.138	.069	
[Time=2.00]	-.069	.035	0^b^	.069	.139	
[Time=3.00]	0^b^	0^b^	0^b^	0^b^	0^b^	
[Treatment=1.00] * [Time=1.00]	.035	-.070	0^b^	-.070	-.035	
[Treatment=1.00] * [Time=2.00]	.035	-.070	0^b^	-.035	-.071	
[Treatment=1.00] * [Time=3.00]	0^b^	0^b^	0^b^	0^b^	0^b^	
[Treatment=2.00] * [Time=1.00]	0^b^	0^b^	0^b^	0^b^	0^b^	
[Treatment=2.00] * [Time=2.00]	0^b^	0^b^	0^b^	0^b^	0^b^	
[Treatment=2.00] * [Time=3.00]	0^b^	0^b^	0^b^	0^b^	0^b^	

Covariance Matrix for Estimates of Fixed Effects^a^	
Parameter	[Time=3.00]	[Treatment=1.00] * [Time=1.00]	[Treatment=1.00] * [Time=2.00]	[Treatment=1.00] * [Time=3.00]	
Intercept	0^b^	.035	.035	0^b^	
[Treatment=1.00]	0^b^	-.070	-.070	0^b^	
[Treatment=2.00]	0^b^	0^b^	0^b^	0^b^	
[Time=1.00]	0^b^	-.070	-.035	0^b^	
[Time=2.00]	0^b^	-.035	-.071	0^b^	
[Time=3.00]	0^b^	0^b^	0^b^	0^b^	
[Treatment=1.00] * [Time=1.00]	0^b^	.140	.070	0^b^	
[Treatment=1.00] * [Time=2.00]	0^b^	.070	.141	0^b^	
[Treatment=1.00] * [Time=3.00]	0^b^	0^b^	0^b^	0^b^	
[Treatment=2.00] * [Time=1.00]	0^b^	0^b^	0^b^	0^b^	
[Treatment=2.00] * [Time=2.00]	0^b^	0^b^	0^b^	0^b^	
[Treatment=2.00] * [Time=3.00]	0^b^	0^b^	0^b^	0^b^	

Covariance Matrix for Estimates of Fixed Effects^a^	
Parameter	[Treatment=2.00] * [Time=1.00]	[Treatment=2.00] * [Time=2.00]	[Treatment=2.00] * [Time=3.00]	
Intercept	0^b^	0^b^	0^b^	
[Treatment=1.00]	0^b^	0^b^	0^b^	
[Treatment=2.00]	0^b^	0^b^	0^b^	
[Time=1.00]	0^b^	0^b^	0^b^	
[Time=2.00]	0^b^	0^b^	0^b^	
[Time=3.00]	0^b^	0^b^	0^b^	
[Treatment=1.00] * [Time=1.00]	0^b^	0^b^	0^b^	
[Treatment=1.00] * [Time=2.00]	0^b^	0^b^	0^b^	
[Treatment=1.00] * [Time=3.00]	0^b^	0^b^	0^b^	
[Treatment=2.00] * [Time=1.00]	0^b^	0^b^	0^b^	
[Treatment=2.00] * [Time=2.00]	0^b^	0^b^	0^b^	
[Treatment=2.00] * [Time=3.00]	0^b^	0^b^	0^b^	

a. Dependent Variable: Vo2_85.	
b. The covariance is set to zero because it is associated with a redundant parameter.	


Covariance Parameters


Estimates of Covariance Parameters^a^	
Parameter	Estimate	Std. Error	
Residual	.131	.014	
Treatment	Variance	.034	.039	
Time	Variance	.034	.037	
Treatment * Time	Variance	.031	.025	

a. Dependent Variable: Vo2_85.	


Covariance Matrix for Estimates of Covariance Parameters^a^	
Parameter	Residual	Treatment	Time	Treatment * Time	
		Variance	Variance	Variance	
Residual	.000	2.977E-10	-5.690E-10	-7.010E-6	
Treatment	Variance	2.977E-10	.002	.000	.000	
Time	Variance	-5.690E-10	.000	.001	.000	
Treatment * Time	Variance	-7.010E-6	.000	.000	.001	

a. Dependent Variable: Vo2_85.	


Random Effects Covariance Structures (G)


Treatment^a^	
	[Treatment=1.00]	[Treatment=2.00]	
[Treatment=1.00]	.034	0	
[Treatment=2.00]	0	.034	

Variance Components^a^	
a. Dependent Variable: Vo2_85.	


Time^a^	
	[Time=1.00]	[Time=2.00]	[Time=3.00]	
[Time=1.00]	.034	0	0	
[Time=2.00]	0	.034	0	
[Time=3.00]	0	0	.034	

Variance Components^a^	
a. Dependent Variable: Vo2_85.	


Treatment * Time^a^	
	[Treatment=1.00] * [Time=1.00]	[Treatment=1.00] * [Time=2.00]	[Treatment=1.00] * [Time=3.00]	[Treatment=2.00] * [Time=1.00]	
[Treatment=1.00] * [Time=1.00]	.031	0	0	0	
[Treatment=1.00] * [Time=2.00]	0	.031	0	0	
[Treatment=1.00] * [Time=3.00]	0	0	.031	0	
[Treatment=2.00] * [Time=1.00]	0	0	0	.031	
[Treatment=2.00] * [Time=2.00]	0	0	0	0	
[Treatment=2.00] * [Time=3.00]	0	0	0	0	

Treatment * Time^a^	
	[Treatment=2.00] * [Time=2.00]	[Treatment=2.00] * [Time=3.00]	
[Treatment=1.00] * [Time=1.00]	0	0	
[Treatment=1.00] * [Time=2.00]	0	0	
[Treatment=1.00] * [Time=3.00]	0	0	
[Treatment=2.00] * [Time=1.00]	0	0	
[Treatment=2.00] * [Time=2.00]	.031	0	
[Treatment=2.00] * [Time=3.00]	0	.031	

Variance Components^a^	
a. Dependent Variable: Vo2_85.	


Estimated Marginal Means


1. Treatment


Estimates^a^	
Treatment	Mean	Std. Error	df	95% Confidence Interval	
				Lower Bound	Upper Bound	
1.00	1.828	.239	4.437	1.189	2.466	
2.00	1.773	.239	4.440	1.134	2.411	

a. Dependent Variable: Vo2_85.	


Pairwise Comparisons^a^	
(I) Treatment	(J) Treatment	Mean Difference (I-J)	Std. Error	df	Sig.^b^	95% Confidence Interval for Difference^b^	
						Lower Bound	
1.00	2.00	.055	.303	2.783	.868	-.952	
2.00	1.00	-.055	.303	2.783	.868	-1.062	

Pairwise Comparisons^a^	
(I) Treatment	(J) Treatment	95% Confidence Interval for Difference	
		Upper Bound	
1.00	2.00	1.062	
2.00	1.00	.952	

Based on estimated marginal means^a^	
a. Dependent Variable: Vo2_85.	
b. Adjustment for multiple comparisons: Bonferroni.	


Univariate Tests^a^	
Numerator df	Denominator df	F	Sig.	
1	2.783	.033	.868	

The F tests the effect of Treatment. This test is based on the linearly independent pairwise comparisons among the estimated marginal means.^a^	
a. Dependent Variable: Vo2_85.	


2. Time


Estimates^a^	
Time	Mean	Std. Error	df	95% Confidence Interval	
				Lower Bound	Upper Bound	
1.00	1.778	.262	6.957	1.158	2.398	
2.00	1.773	.262	6.998	1.153	2.393	
3.00	1.849	.262	6.957	1.229	2.469	

a. Dependent Variable: Vo2_85.	


Pairwise Comparisons^a^	
(I) Time	(J) Time	Mean Difference (I-J)	Std. Error	df	Sig.^b^	95% Confidence Interval for Difference^b^	
						Lower Bound	
1.00	2.00	.005	.322	4.352	1.000	-1.210	
	3.00	-.071	.321	4.335	1.000	-1.287	
2.00	1.00	-.005	.322	4.352	1.000	-1.220	
	3.00	-.076	.322	4.352	1.000	-1.291	
3.00	1.00	.071	.321	4.335	1.000	-1.146	
	2.00	.076	.322	4.352	1.000	-1.139	

Pairwise Comparisons^a^	
(I) Time	(J) Time	95% Confidence Interval for Difference	
		Upper Bound	
1.00	2.00	1.220	
	3.00	1.146	
2.00	1.00	1.210	
	3.00	1.139	
3.00	1.00	1.287	
	2.00	1.291	

Based on estimated marginal means^a^	
a. Dependent Variable: Vo2_85.	
b. Adjustment for multiple comparisons: Bonferroni.	


Univariate Tests^a^	
Numerator df	Denominator df	F	Sig.	
2	4.346	.035	.966	

The F tests the effect of Time. This test is based on the linearly independent pairwise comparisons among the estimated marginal means.^a^	
a. Dependent Variable: Vo2_85.	


3. Treatment * Time^a^	
Treatment	Time	Mean	Std. Error	df	95% Confidence Interval	
					Lower Bound	Upper Bound	
1.00	1.00	1.782	.321	9.345	1.059	2.504	
	2.00	1.805	.322	9.402	1.082	2.528	
	3.00	1.896	.321	9.345	1.173	2.618	
2.00	1.00	1.774	.321	9.345	1.052	2.497	
	2.00	1.741	.322	9.434	1.018	2.464	
	3.00	1.802	.321	9.345	1.080	2.525	

a. Dependent Variable: Vo2_85.	
